# Asymmetry in the peak in Covid-19 daily cases and the pandemic R-parameter

**DOI:** 10.1101/2023.07.23.23292960

**Authors:** Sayali Bhatkar, Mingyang Ma, Mary Zsolway, Ayush Tarafder, Sebastian Doniach, Gyan Bhanot

## Abstract

Within the context of the standard SIR model of pandemics, we show that the asymmetry in the peak in recorded daily cases during a pandemic can be used to infer the pandemic R-parameter. Using only daily data for symptomatic, confirmed cases, we derive a universal scaling curve that yields: (i) r_eff_, the pandemic R-parameter; (ii) L_eff,_ the effective latency, the average number of days an infected individual is able to infect others and (iii) α, the probability of infection per contact between infected and susceptible individuals. We validate our method using an example and then apply it to estimate these parameters for the first phase of the SARS-Cov-2/Covid-19 pandemic for several countries where there was a well separated peak in identified infected daily cases. The extension of the SIR model developed in this paper differentiates itself from earlier studies in that it provides a simple method to make an a-posteriori estimate of several useful epidemiological parameters, using only data on confirmed, identified cases. Our results are general and can be applied to any pandemic.

## INTRODUCTION

A pandemic occurs when a new pathogen enters a naïve population. The recent SARS-Cov-2 pandemic was caused by a Coronavirus, one of a family of large, enveloped, single-stranded RNA viruses that are widespread in animals and usually cause only mild respiratory illnesses in humans [1-5]. In 2003, a new coronavirus emerged, and was named SARS-CoV (Severe Acute Respiratory Syndrome – Corona Virus). This virus caused a life-threatening respiratory disease in humans, with a fatality rate of almost 10% [6,7]. In fact, after an initial burst of interest in development of treatment options, interest in this virus waned. The emergence of the novel coronavirus SARS-CoV-2, identified in December 2019 in Wuhan, China, has since caused a worldwide pandemic [8-13]. SARS-CoV-2 is the seventh known coronavirus to cause pathology in humans [1]. The associated respiratory illness, called COVID-19, ranges in severity from a symptomless infection [8], to common-cold like symptoms, to viral pneumonia, organ failure, neurological complications, and death [9-11]. While the mortality in SARS-CoV-2 infections is lower than in SARS-CoV [9-12], it has more favorable transmission characteristics, a higher reproduction number, a long latency period and an asymptomatic infective phase [13].

The governments of several countries took significant measures to slow the infection rate of Covid-19, such as social distancing, quarantine, identification, tracking and isolation. However, there was no uniform policy, some governments reacted later than others, and some (e.g. Sweden) decided to keep the country open, leaving counter-measures up to individuals. A large amount of consistent public data is now available on the number of tests performed, the number of confirmed infected cases, and the number of deaths in different contexts, such as locations and health conditions [14]. These provide important sources of information for the development and testing of models to estimate pandemic characteristics, guide public policy and assess the efficacy of interventions [15]. In this paper, we use data from the WHO website: https://covid19.who.int/WHO-COVID-19-global-data.csv.

It is well known that in most pandemics, confirmed infected cases often seriously underestimate the actual number of infections [16,17]: not everyone who is infected is symptomatic, and not everyone who dies from the disease has been tested [18]. Even the number of reported deaths may be underestimated because of co-morbidities; i.e. COVID-19 increases susceptibility to other diseases and conditions [19]. Moreover, the virus can be transmitted by asymptomatic individuals, who can comprise a substantial portion of the infected population [20], militating against accurate estimates of total infection rates. In this context, as indicated in [21], analytical models can provide useful information.

Dynamical (mechanistic) models have been used for forecasting and for making projections. For example, projections and forecasting models of various types were used as early as February 2020 to determine a reproductive number for SARS-CoV-2 [13]. More generally, multiple research groups have models to estimate Case Fatality Ratios (CFRs) [22], to forecast and project the need for hospital beds [23] and to project and forecast mortality [24]. Among the many applications of models to COVID-19, four variable Susceptible-Exposed-Infective-Recovered (SEIR) models have been used to project the impact of social distancing on mortality [25], three variable Susceptible-Infective-Recovered (SIR) models have been used to estimate case fatality and recovery ratios early in the pandemic [26], and a time delayed SIR has been used to evaluate the effectiveness of suppression strategies [27]. One of the most ambitious dynamical models, which includes 8 state variables, and 16 parameters, was fruitfully applied to evaluate intervention strategies in Italy, in spite of the fact that parameter identifiability could not be assured [28]. There is also some model based evidence that the transmission of the SARS-Cov-2 virus is regulated by temperature and humidity [29]. In this paper, we model the Covid-19 pandemic using an extension of the SIR model [30], which partitions the population into three compartments: Susceptibles (S),

Infectious (I) and Removed R. This and other models (using more variables) have been used in a variety of contexts to study the global spread of diseases (For some recent reviews, see [31-33]).

In this paper, within the context of an extension of the standard epidemiological SIR model [30], using only daily recorded case data of symptomatic individuals, we develop a method to estimate: (i) the pandemic parameter r_eff_, which is the average number of individuals infected by a single infected individual, (ii) the effective latency L_eff_, which is the average time an infected individual is able to infect others, and (iii) the probability *α* of infection from a single encounter between an infected and a susceptible individual.

## METHOD

The spread of a virus depends on several factors, such as patterns of contacts among infectious and susceptible persons, the latency period between being infected and becoming infective and/or developing symptoms, the duration of infection/infectivity, immunity acquired by previous exposures, effects of vaccination, etc. In spite of these complications, some of the features of the spread are well described by a simple epidemiological model, the so-called SIR model [30]. In this model, at any given instant in time, the population is divided into three compartments : susceptible (S), infected (I) and recovered (R) with S+I+R = N. The parameter N is assumed to be constant in time and is the number of individuals exposed to the virus in a given region. It is assumed that individuals in the S compartment are equally susceptible to being infected by individuals in the I compartment and individuals in the R compartment are both immune to further infection and unable to infect others. The R compartment also includes those who are “removed” (dead, quarantined etc.).

At any given moment in time, the reported number of infected individuals is a subset of the actual number of infected individuals [16,17]. Let I_0_(t) and I_1_(t) be the number of individuals in the unidentifiable and identifiable compartments respectively, with I_1_(t) = *ω*I(t) and I_0_(t) = (1-*ω*)I(t). Similarly, let R(t) = R_0_(t) + R_1_(t) where R_0_(t) are derived from I_0_(t), and R_1_(t) are derived from I_1_(t). We assume that individuals in the I_1_ compartment, once identified, are no longer able to infect others because they would be isolated, confined, or quarantined. On the other hand, individuals in the I_0_ compartment, being asymptomatic and unaware of their infected state, would continue to infect others until they become non-infective. Let L_0_ be the average number of days an asymptomatic individual is infective and L_1_ be the average number of days a symptomatic individual is infective. Let 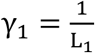 and 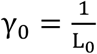 be the rates at which these two types of infected individuals leave the infective pool. Under these assumptions, we propose the following simple extension of the SIR model for the pandemic dynamics:

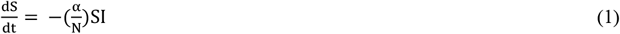

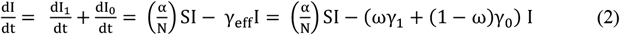

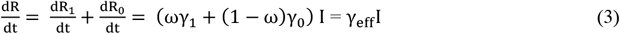

α is the probability of infection in a single encounter between an infected and susceptible individual and

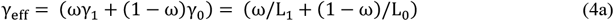

is the rate at which an individual in the I compartment transitions to the R compartment. The reciprocal L_eff_ of γ_eff_ is the average effective latency, the average number of days that an infected individual (symptomatic or not) is infective. Thus,

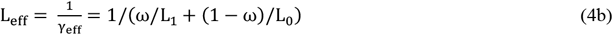

Let X(t) be the observed rate at which daily cases are identified. From Eq. 3,

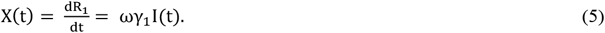

The key relationship we exploit in this paper is Eq. 5, the fact that the number of daily observed cases 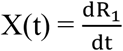, is proportional to I(t). This proportionality implies that the width and location (in units of time t) of the peak in X(t) and I(t) are the same. The location of the peak in I(t) or X(t) is difficult to estimate because it depends on identifying the “start” of the pandemic, i.e., the day when the first person was infected. However, other characteristics of the peak in X(t) are easier to measure. As we show below, scaling laws relating the pandemic parameter R can be inferred using dimensionless quantities from the form of X(t) using Eq. 1-5. In this paper, we will use the left and right half-widths at the peak in X(t) to estimate parameters.

To do this, we rescale the time t to 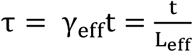 Eq. 1-3 can then be rewritten in terms of the fractions s = S/N, i = I/N, r= R/N, r_1_ = R_1_/N and x = X/N as follows:

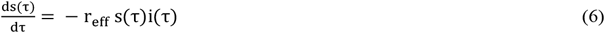

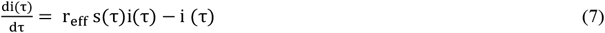

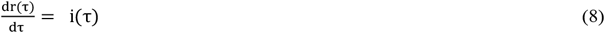

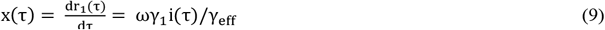

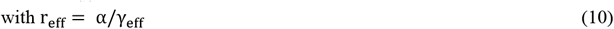

Let W_L_ and W_R_ be the left and right half widths of the peak in X(t). From Eq. 5, we note that the dimensionless quantity W_R_/W_L_ is the same for X(t), I(t). Furthermore, since x(τ) and X(t) are proportional and the relationship between τ and t is just a scale factor (τ = γ_eff_t), W_R_/W_L_ is the same for x(τ) and X(t). Finally, from Eq 6-9, it is obvious that W_R_/W_L_ depends only on r_eff_. The functional relationship between W_R_/W_L_ and r_eff_, can be obtained by numerically solving Eqs 6-9 and is shown in Fig. 1 (detailed values in Supplementary Table 1). Note that the result of Fig. 1 is a universal scaling law in the SIR model which applies to any pandemic. We will now demonstrate with an example how the parameters r_eff_, L_eff_ and *α* can be obtained using only the results in Fig. 1 and Supplementary Table 1 and data for X(t). We note that the fraction f_inf_ = 1-s(∞) of individuals infected at t=∞ can also be computed from r_eff_ using the relationship 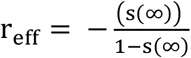 (Appendix A, Eq. A8)

**Figure 1.**
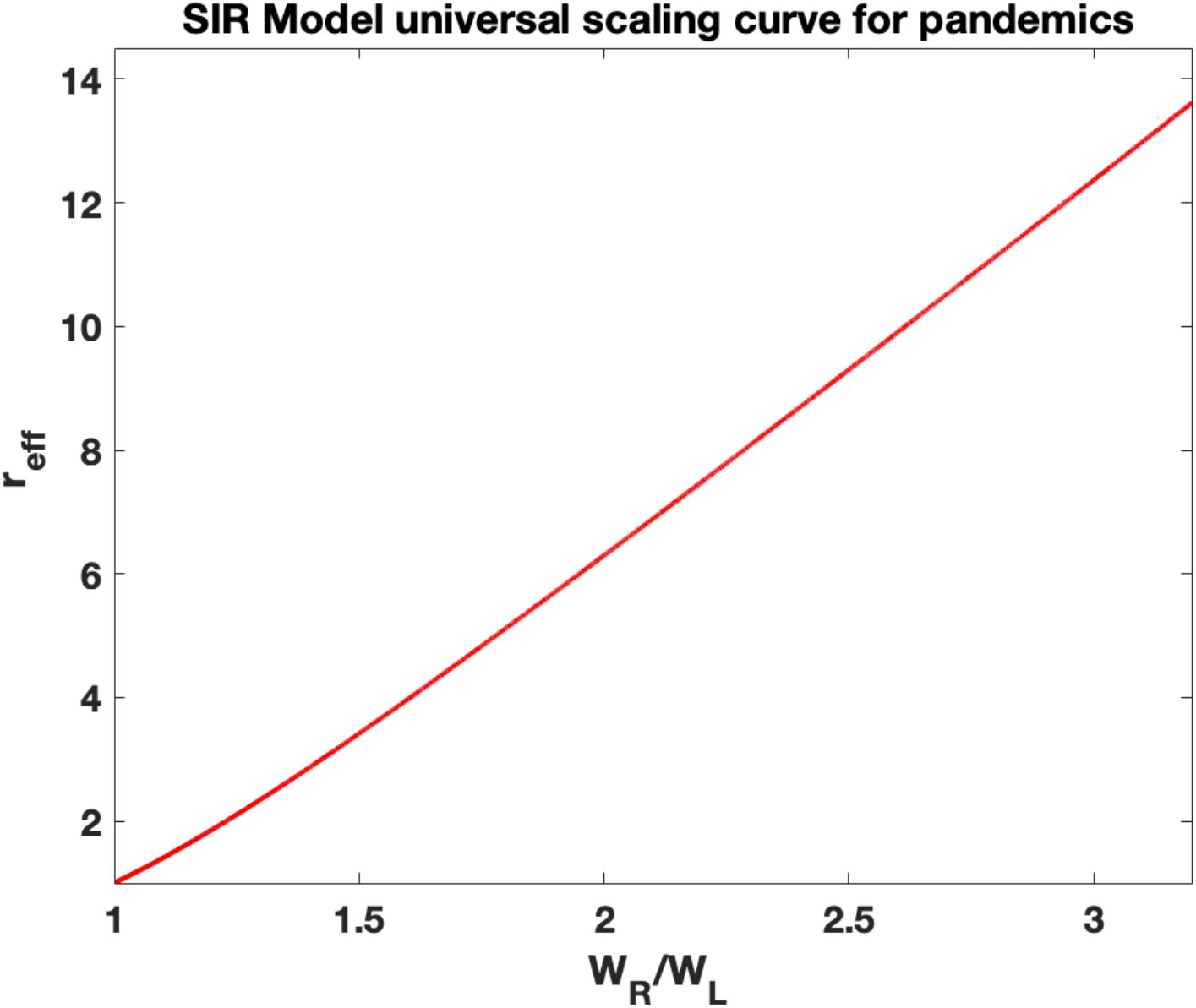
Universal curve in the SIR model for reff as a function of the ratio W_R_/W_L_, where W_L_ and W_R_ are the left and right half widths of the peak in X(t). Note that this scaling function is universal in the SIR model and applies to any pandemic.

Figs 2a, 2b, 2c show an example of the procedure we follow to find parameters from only X(t) data. Fig 2a shows X(t) from a numerical solution of Eqs. 1-4 for parameter values N = 4.6×10^5^, L_0_ = 10 days, L_1_ = 5 days, *ω* = 0.25, *α* = 0.46875. These parameter values correspond to *γ*_eff_ = 0.125 (Eq. 4a), L_eff_ = 8 (Eq. 4b) and r_eff_ = 3.75 (Eq. 10).

**Figure 2 a-b:**
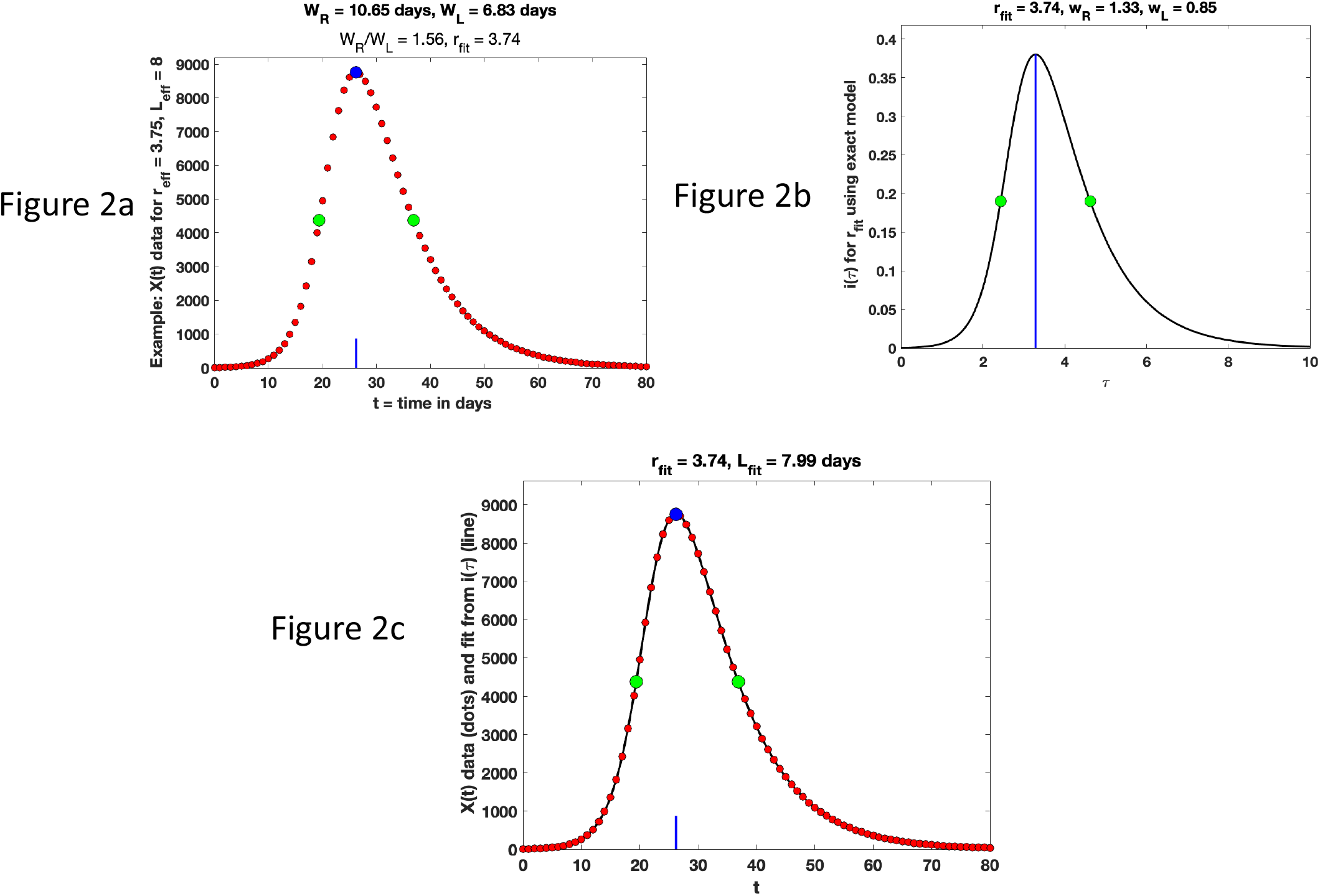
Results obtained by numerically solving the SIR equations (Eqs. 1-5) for parameter values N = 4.6×10^5^, L_0_ = 10 days, L_1_ = 5 days, ω = 0.25, α = 0.46875, which corresponds to r_eff_ = 3.75. The red dots in Fig 2a show daily case data (X(t)) from a numerical solution of Eqs. 1-5 using these parameter values. The blue line shows the location of the maximum and green dots are the locations of the right and left half width points. Using the values of W_R_ and W_L_ inferred from X(t), an estimate (r_fit_) of the pandemic parameter (r_eff_) was obtained by interpolation from Supplementary Table 1. Fig 2b shows i(τ) by solving Eq. 6-9 for the value r_fit_ = 3.74 obtained from Fig 2a. The solid line in Fig 2c shows the result of mapping this solution to the data for X(t) by matching the peak of X(t) to the peak of i(τ) and rescaling the τ axis to match the half width for i(τ). The rescaling of the half width from τ to t provides the estimate L_eff_ = (W_R_+W_L_)/(w_R_+w_L_) = 7.99 days.

The procedure we follow to estimate the parameters from just the data for X(t) (Fig 2a) and the universal scaling results in Figure 1/Supplementary Table 1 is as follows:

- We first estimate the ratio W_R_/W_L_ from the data for X(t) (Fig. 2a). We find W_R_ = 10.65 days, W_L_ = 6.83 days, and W_R_/W_L_ = 1.56. We also obtain the maximum value of X(t) as
- M_X(t)_ = 8768.8 (shown as a blue dot in Fig. 2a).
- The value of W_R_/W_L_ is used to infer r_eff_ by interpolating the data in Supplementary Table 1. This yields the value r_eff_(estimated) = r_fit_ = 3.74.
- Using r_fit_ =3.74, we solve Eqs. 6-9 to generate i(τ), shown in Figure 2b.
- The right and left widths w_R_ and w_L_ at half maximum are estimated from the data for i(τ). This gives w_R_ = 1.33, w_L_ = 0.85. Note that these quantities are dimensionless. Similarly, the maximum in i(τ) is measured to be m_i(*τ*)_ = 0.3859. These quantities are measured very accurately because they result from solving a set of equations.
- An estimate L_eff_ is now obtained as the ratio L_eff_ = (W_R_+W_L_)/(w_R_+w_L_) = 7.99 days.
- To check whether the fit is good, we multiply i(*τ*) in Fig 2b by M_X(t)_/m_i(*τ*)_, scale the *τ* axis by a factor of L_fit_ and shift the location of the maximum in X(t) to coincide with the peak in X(t). This fit is shown in Fig. 2c.
- In this example, we used a small time-step to generate accurate data for X(t). In reality, only daily data is available and there would be an error of approximately 0.5 days in estimates of W_R_ and W_L_. Using this, we can estimate the error in the parameters. The final results for our example are: W_R_/W_L_ = 1.56 +/-0.14, r_eff_ = 3.74 +/-0.76, L_eff_ = 7.99 +/-1.42 days.

Note that in the procedure described above, we used only the observed daily cases X(t) and the universal scaling results in Figure 1 and Table 1.

**Table 1:**
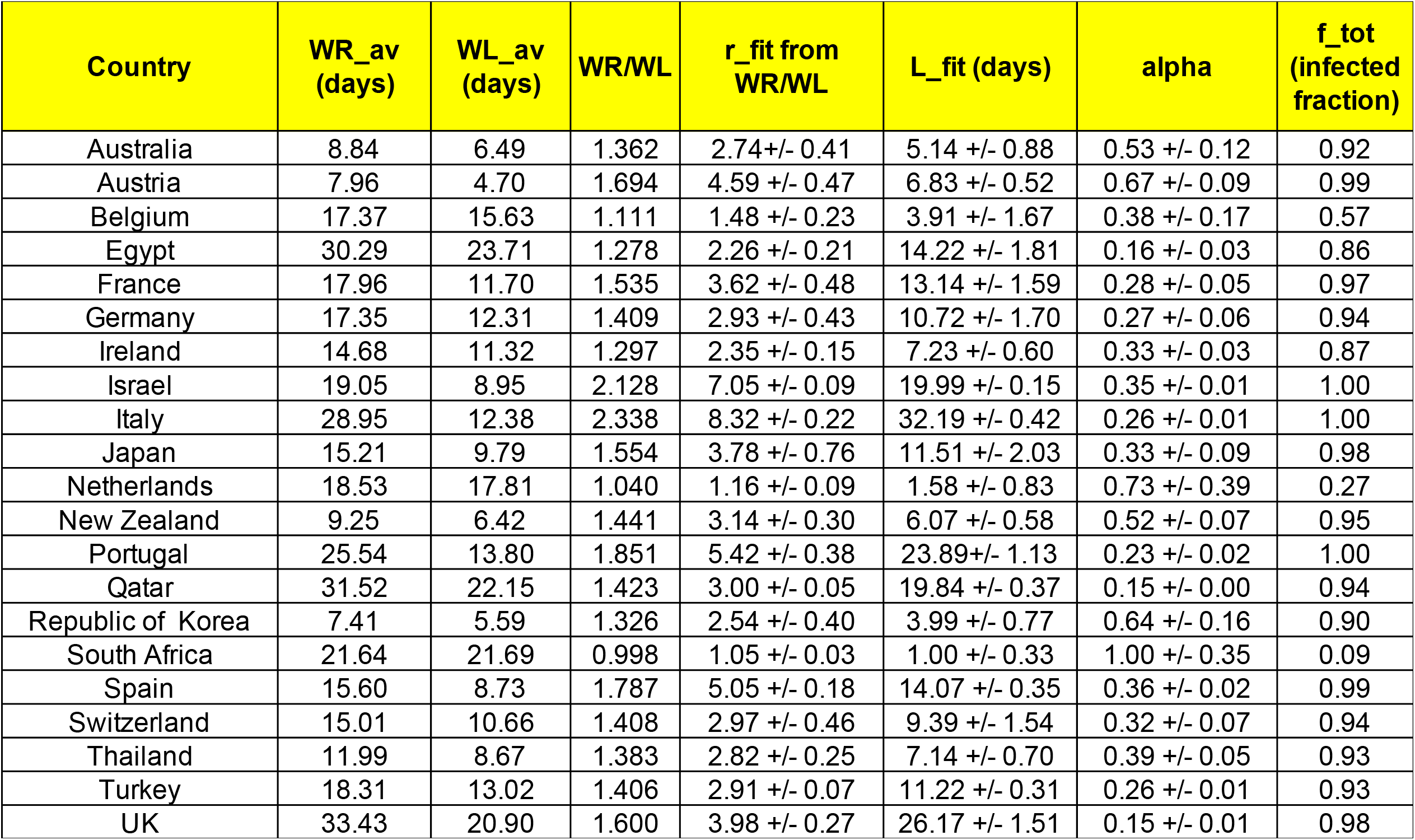
Results for W_R_, W_L_, r_eff_, L_eff_, f_tot_, and α for twenty-one countries using the scaling law in Fig 1 and Supplementary Table 1. Here W_L_ and W_R_ are the left and right half widths of the peak in X(t), the number of recorded daily cases.

When we apply this procedure to pandemic data, we have to first contend with one additional issue, which is that the data shows a three, five, or seven day cycle, depending on the country. This is presumably due to the logistics of data collection and reporting, and requires smoothing before our method can be applied. In this paper, we averaged the raw data for each country over five, seven and nine days; inferred r_eff_ for each averaging scheme and used the average of the values obtained as the final result. Finally, as our method is sensitive to the position of the peak in daily cases, and since we are interested in the asymmetry near the peak, we fitted the data near the peak to a cubic to obtain the location (in time) and height of the peak and used these values and the smoothed data points to find W_R_ and W_L_.

## RESULTS

Worldwide data for confirmed Covid-19 cases and deaths from January 3, 2020 was downloaded from the World Health Organization (WHO) website: https://covid19.who.int/WHO-COVID-19-global-data.csv (Supplementary Table 2). The data for daily cases obtained from this source directly measures the function X(t) in our analysis. Before performing any analysis, the data for daily cases was smoothed/averaged as described above.

Our model assumes a single circulating viral strain infecting a homogeneous set of individuals in a given region who were equally susceptible to infection (uniform immune response). The model also assumes that exposed individuals observed the same rules regarding the use of masks/isolation/quarantine, there was no significant variation in population density among them, little variation in their movements, equal vaccination status, and symptomatic cases once identified were quarantined. These requirements would most likely be valid, at least for some countries, for the first wave of the Covid-19 pandemic when the world population was naïve to the virus (no immunity) and everyone was equally susceptible. Furthermore, at least in some countries, many of the other assumptions of the model did apply, such as homogeneity of response, lack of vaccines resulting in no innate immunity, standard medical protocols, good testing practices, and a single circulating strain of the virus. Such countries would show an initial exponential rise in daily cases at small times followed by a clear peak with no overlap with subsequent peaks. We applied our method to analyze the first peak in daily cases for twenty-one countries that satisfied these conditions.

Supplementary Table 1 gives the scaling function data from solving Eq 6-9 for different values of r_eff_. For each value of r_eff_ the columns show w_L_ = W_L_/L_eff_, w_R_ = W_R_/L_eff_, W_R_/W_L_, m_i(τ)_, f_inf_ = 1-s(∞), r_eff_ calculated from Eq. A8 and the ratio W_R_/W_L_ = w_R_/w_L_. This data is universal and applies to all pandemics. Figs 3-5 show the fits for three countries from very different geographic regions Spain (Europe), Thailand (Asia) and Australia (Oceania). Results and figures for all twenty-one countries are in Table 1 and Supplementary Figures 1.

**Figure 3 a-c:**
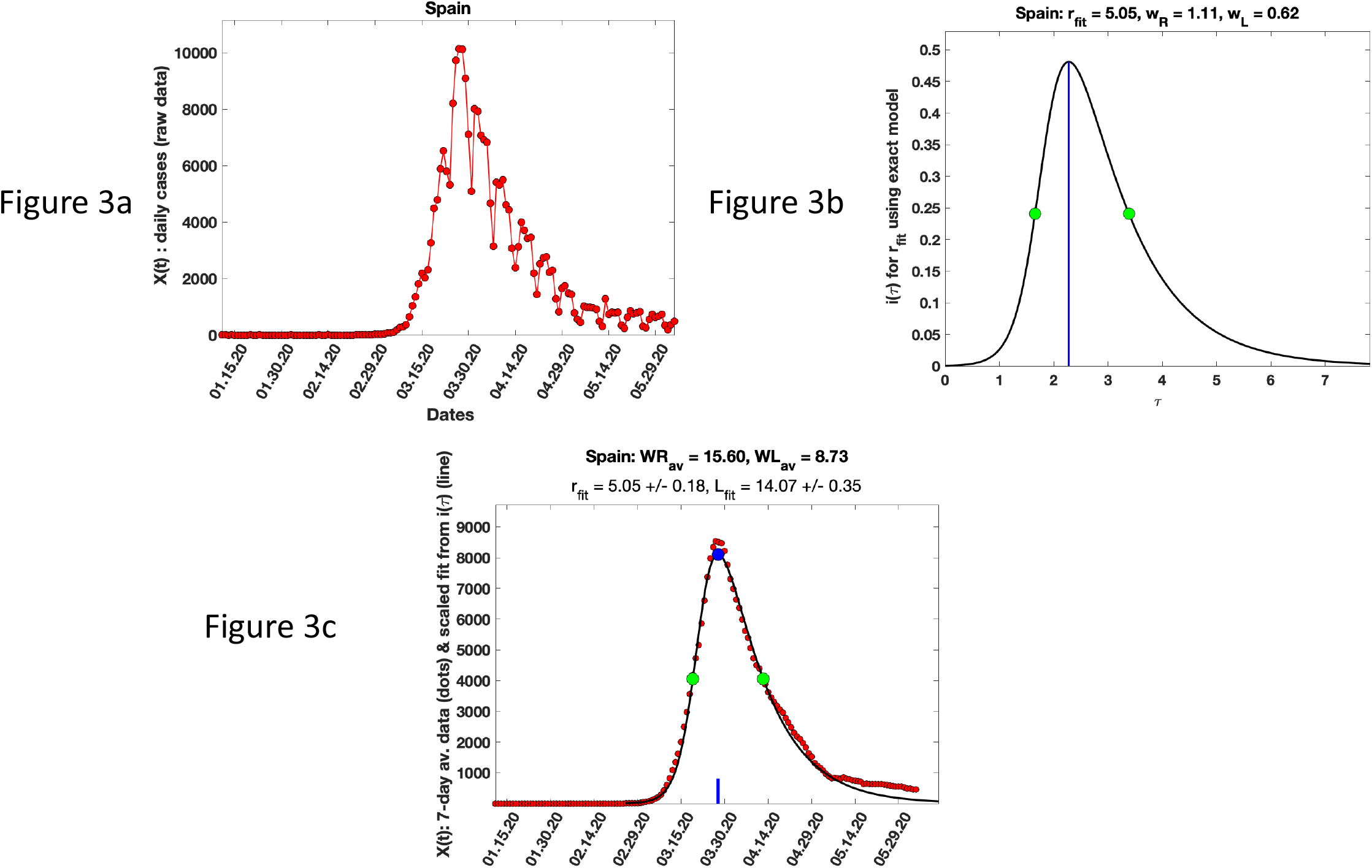
Fit of our model to data for X(t) for Spain (data for X(t) from Supplementary Table 2) from the World Health Organization. Fig 3a is the raw X(t) data for Spain. Fig 3b shows the i(τ) curve for the r_fit_ value obtained from its average value from analysis of X(t) for five, seven and nine day averaging applied to the raw data in Fig 3a. Fig 3c shows the mapping of the i(τ) data to X(t) to match its peak location and height. The points plotted for X(t) are seven day averaged data. The scaling of the τ axis to the t axis allows an estimate of L_eff_ as in the example in Fig 2.

**Figure 4 a-c:**
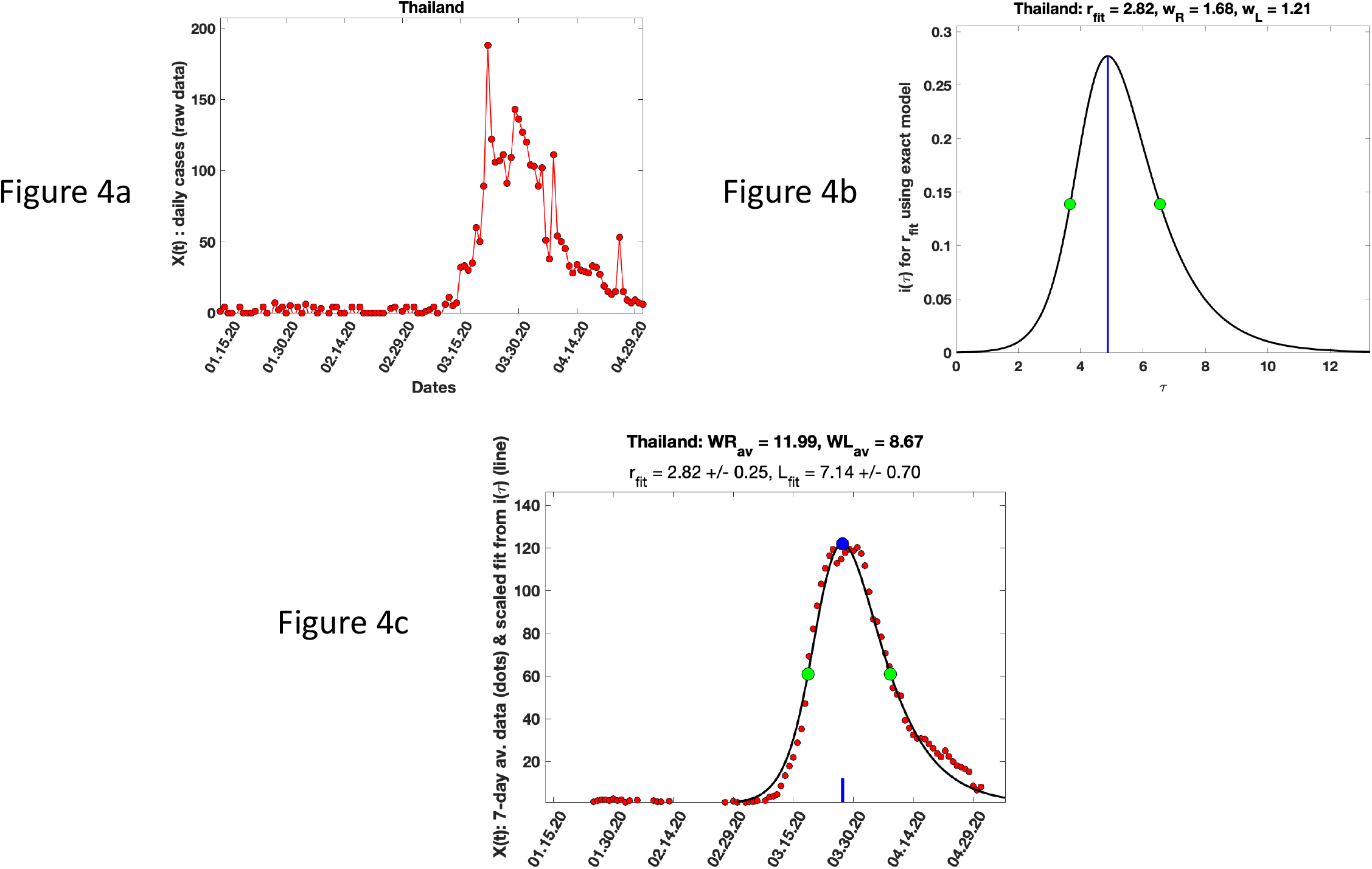
Analysis of X(t) data for Thailand as in Figure 3.

**Figure 5 a-c:**
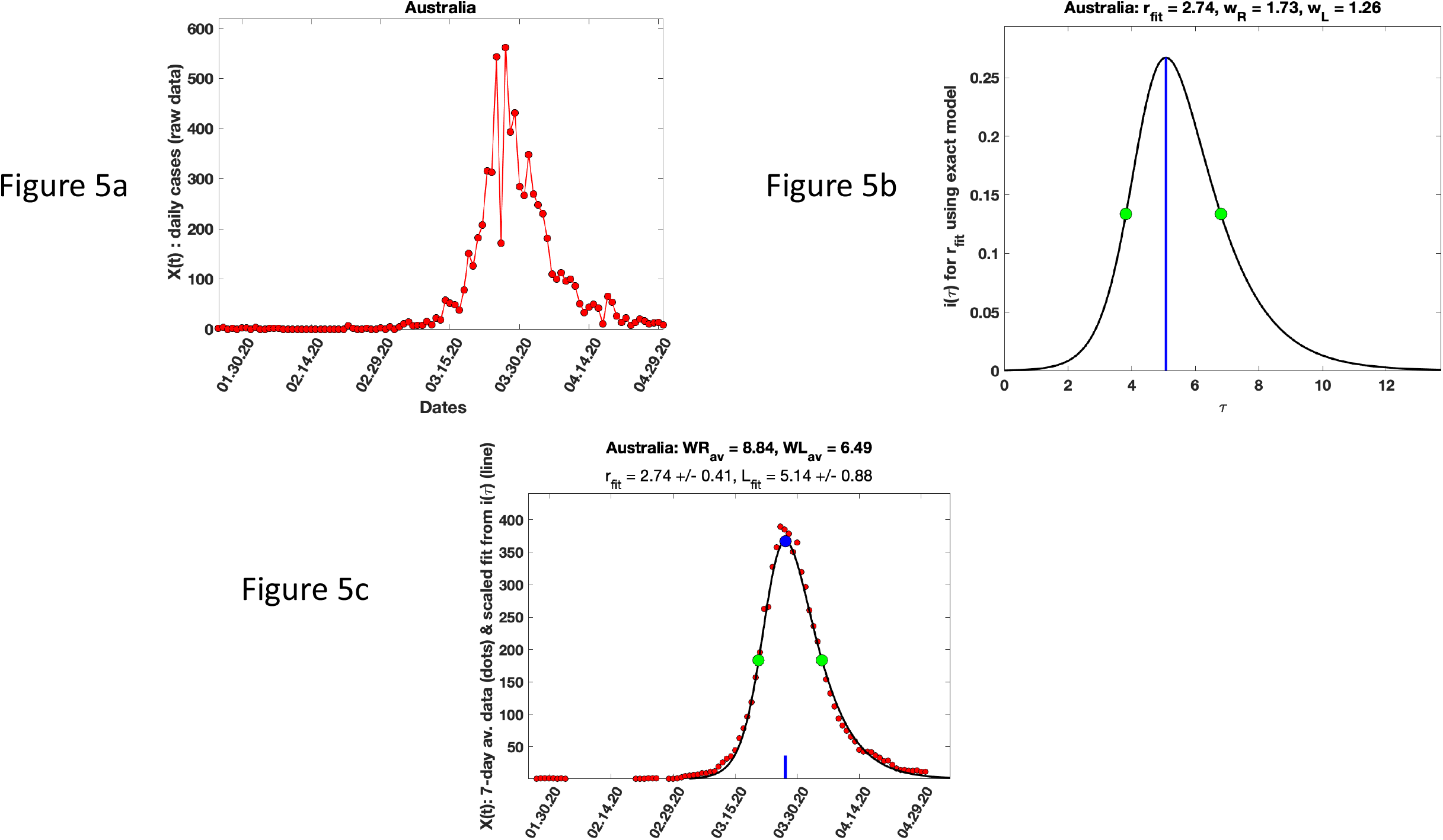
Analysis of X(t) data for Australia as in Figure 3.

Using Eq A in Appendix A, we estimate the total fraction f_tot_ of the exposed subset of the population that was infected (Table 1).Note that this represents both the cases that were counted (symptomatic and/or tested) as well as those that were not identified. By our estimates, 50-100% of exposed individuals (fraction of N in the SIR model) in the countries we analyzed were infected.

Finally, we note that for the daily case data to be reliable, the total number of tests (including positives and negatives) should be sufficiently large. Using test data from (https://www.ecdc.europa.eu/en/publications-data/covid-19-testing) we checked that the number of tests performed on each day were much higher than the number of cases for countries where test data was available for the time range of interest (Supplementary Figures 2).

## DISCUSSION

In this paper we show that the asymmetry of the peak in recorded daily cases in any pandemic contains important information. In particular, for the standard epidemiological SIR model [30], this asymmetry can be used to relate the pandemic R-parameter to the ratio of right and left half widths of the peak, which in turn provides an a-posteriori estimate of several important pandemic parameters, including the effects of both recorded (symptomatic/tested) cases as well as unmeasured/asymptomatic cases, namely: r_eff_, the effective pandemic R-parameter, the effective latency L_eff_ (the average number of days an infected individual is able to infect others), and the infection probability α of transmission from an infected individual to a susceptible individual in a single encounter. These can be inferred using a universal scaling function (Figure 1 and Supplementary Table 1) that relates the ratio of the right and left half widths at full maximum of the peak in daily identified cases X(t) to the effective pandemic R-parameter (r_eff_). Within the limits of the SIR model, our results are general and apply to any pandemic. We apply our method to worldwide country specific data to find r_eff_, L_eff_ and α for the first phase (first peak in daily cases) for the SARS-COV-2 pandemic for twenty-one countries which had a clear, well separated peak in daily cases (Table 1, Supplementary Figures 1).

A novel feature of this analysis is that our result for f_tot_, the fraction of the population that was infected, includes both those who were identified/counted as infected as well as those that were not identified/counted. Our results for f_tot_ suggest that in the 2019-2020 SARS-COV-2 pandemic, the original variant of the virus was highly effective in transmission. In some of the developed countries, we find that almost all exposed individuals were infected in the first phase of the pandemic (Table 1). The fact that f_tot_ is close to unity for many countries means there was a large fraction of infected in the population who were not identified.

Using data from https://www.worldometers.info/world-population/population-by-country and https://en.wikipedia.org/wiki/List_of_cities_by_average_temperature (Supplementary Table 3), we explored possible relationships among the values of the R parameter, the α coefficient and the latency period L_eff_ with the population density, mean temperature during the first peak and the median age of the population (Sup. Figs. 3a-i). Although the results are too noisy to make definitive statements about these relationships, overall, across all the countries analyzed, from the linear fits, the weighted average values of r_eff_, α, and L_eff_ are 2.18, 0.23, and 15.35 days respectively.

Several countries, notably the United States, Canada, The Russian Federation, India, and Pakistan did not meet our criterion of a clear, well separated first peak in daily cases in 2020. This is most likely because they cannot be thought of as homogeneous in the sense of response from local authorities regarding the use of masks, quarantine etc. In the United States for example, the response was state and/or county specific. In principle, our method could be applied at the county or state level in the US or the province or sub-province level in Canada to determine parameters from recorded case data, wherever these compartments had uniform rules for containment of the virus.

It would be interesting to apply our method to subsequent peaks in daily cases for the SARS-COV-2 pandemic, as the virus evolved into different variants across the globe. Comparing changes in the inferred parameters across countries would provide a country specific estimate of the effects of preventive measures such as the effectiveness/efficacy of vaccination, changes in behavior (mask use, testing/quarantine, work-from-home, social distancing, travel restrictions) etc. Furthermore, this method could be applied to other viral pandemics, such as the SARS-COV pandemic of 2003, and Influenza pandemics of the past, such as the H1N1 Spanish Flu pandemic of 1918-19 which recurred in 1950 and 1977, the H2N2 Asian Flu pandemic of 1957, the H3N2 Hongkong pandemic of 1968 and the more deadly H5N1 East Asian pandemic of 1997.

### Summary

In this paper, we propose a simple method to estimate the R parameter of epidemics using the asymmetry around the peak in daily reported cases. We quantify this asymmetry using the ratio W_R_/W_L_ of the right half width W_R_ to the left half width W_L_ and show how this dimensionless ratio can be used to estimate pandemic parameters. The function that makes this possible is shown in Fig. 1 and Supplementary Table 1. Within the SIR model, this relationship between r_eff_ and W_R_/W_L_ is universal and can be used for any epidemic. The value of such estimates is of necessity limited by the accuracy and reliability of the available data. However, as we show using a simulated example (Fig. 2), given data of sufficient accuracy, our method is an extremely simple and accurate way to estimate the R parameter, α, and the effective latency.

## Data Availability

All data produced in the present work are contained in the manuscript and supplementary material.

## Supplementary Figure Captions

Supplementary Figures 1: Raw data, i(τ) and fits of our model to data for X(t) for twenty-one countries. In addition to figures similar to Figs 3-5, we also include the fits to 5,7,9 day averaged data. These show that the r_eff_ values for these various averagings are in good agreement.

Supplementary Figures 2: Data for total number of tests carried out in 3 countries of interest for the period of interest. Data for tests is from https://www.ecdc.europa.eu/en/publications-data/covid-19-testing. The blue dots are the number of positive tests and the red dots are the total number of tests carried out (including both positive and negative cases). The positive fraction in all cases was sufficiently low for the testing to be considered adequate. This means that the X(t) data we used here should be reliable.

Supplementary Figures 3: Exploring possible relationships among r_eff_, α and L_eff_ (in days) and a the population density, average temperature and median age of population. Demographic data from https://www.worldometers.info/world-population/population-by-country and temperature data from https://en.wikipedia.org/wiki/List_of_cities_by_average_temperature.

## Supplementary Table Captions

Supplementary Table 1 : Detailed results from numerical solutions of Eq 6-9 for different values of r_eff_. These were used in Figure 1 and in inferring r_eff_ from X(t) data. Within the context of the SIR model, the relationship between r_eff_ and W_R_/W_L_ in this Table is universal and applies to any pandemic.

Supplementary Table 2 : Data on recorded cases for the SARS-Cov2 pandemic from the World Health organization that was used in the analysis (from https://covid19.who.int/WHO-COVID-19-global-data.csv)

Supplementary Table 3: Expanded version of Table 1 including demographic and temperature data from https://www.worldometers.info/world-population/population-by-country and https://en.wikipedia.org/wiki/List_of_cities_by_average_temperature.

## Author Contributions

Data Analysis and coding: SB, MM, AT, MZ, GB Ideas: SB, GB, SD

Paper writing: SB, GB

### Author Approval

All authors have seen and approved the manuscript.

### Competing Interests

The authors declare there are no competing interests.

## Acknowledgements

GB was partly supported by grants from DoD (KC180159) and NIH (P01CA250957). GB thanks Professor Charles DeLisi for many discussions and collaboration on earlier (unpublished) work on the SARS-CoV-2 pandemic using the SIR model. GB also thanks the Aspen Center for Physics, which is supported by National Science Foundation grant PHY-1607611, the Geballe Lab for Advanced Materials at Stanford University, and Professor Gautam Mandal and the Tata Institute of Fundamental Research for their kind hospitality while this work was being done.

## Appendix A: Some useful results

The rescaled equations for the pandemic dynamics are:

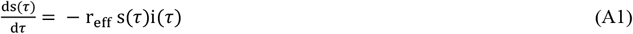

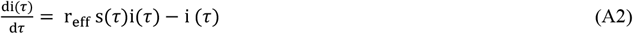

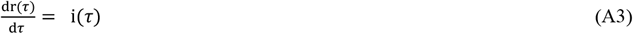

The scaled quantities s(τ), i(τ) are related to S(t), I(t) of the SIR model by:

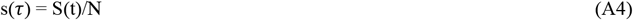

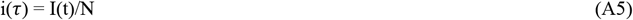

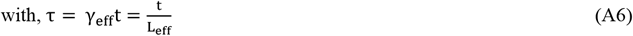

Dividing (A2) by (A1) gives:

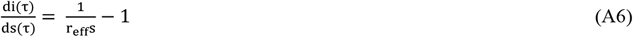

Using the large N boundary conditions s(0) = 1, i(0) = 0 generates the exact result:

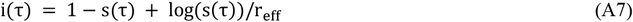

At t=∞, i(τ) = 0. Hence,

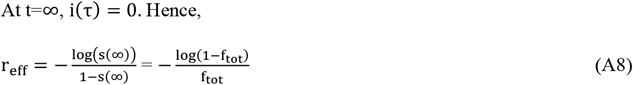

When s(∞) = 1 (no pandemic), L’Hôpital’s rule gives r_eff_(s(∞) = 1) = 1.

It is easy to see that for 0 ≤ s(∞) < 1, r_eff_> 1. Hence, a pandemic requires r_eff_> 1.

From (A2), the maximum in i(τ) happens when s(τ) = 1/r_eff_. Hence:

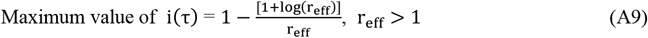

Note that because of (A5), this quantity is the same as the maximum of I(t)/N which is the quantity H_I_/N in Eq. 12c in the main text. Hence,

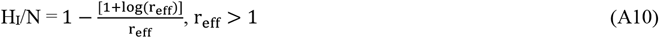

For small τ, s(τ)∼1. Hence, we can expand the right-hand side of (A7) in powers of (1-s(τ)). To lowest order,

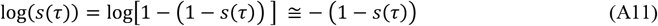

Substituted into (A7) this gives,

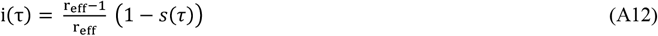

Substituting from (A12) into (A1) gives the Logistic Equation:

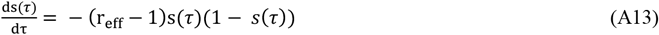

whose solution, with the boundary condition s(0) = 1-ε is:

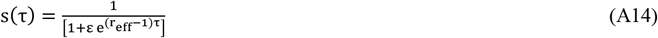

Hence, for 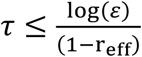,

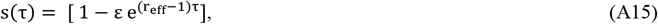

Combining (A12) and A(15) shows that for 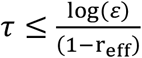,

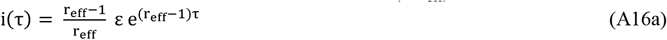

Hence, from Eq. 9,

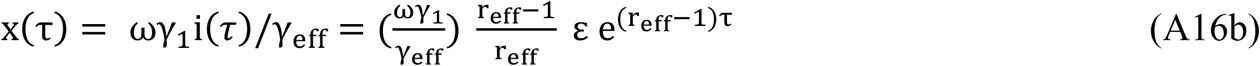

This shows that,

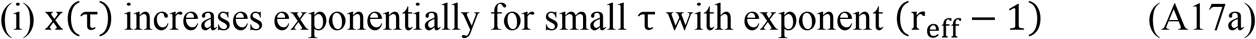

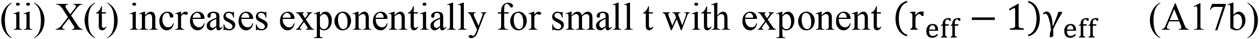

Finally, using similar arguments, it is easy to show that,

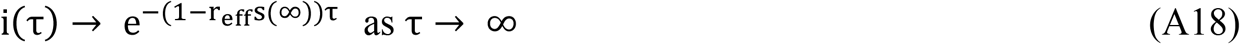

Hence,

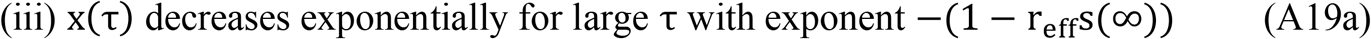

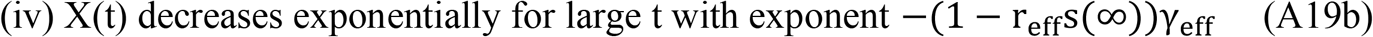

## Notes

### Competing Interest Statement

The authors have declared no competing interest.

### Author Declarations

https://covid19.who.int/WHO-COVID-19-global-data.csv. https://www.ecdc.europa.eu/en/publications-data/covid-19-testing

### Summary of Updates

Earlier version was unreadable. Results unchanged.

