## Supplementary figures and images for "Asymmetry in the peak in Covid-19 daily cases and the pandemic R-parameter"

### Australia_raw_data.png

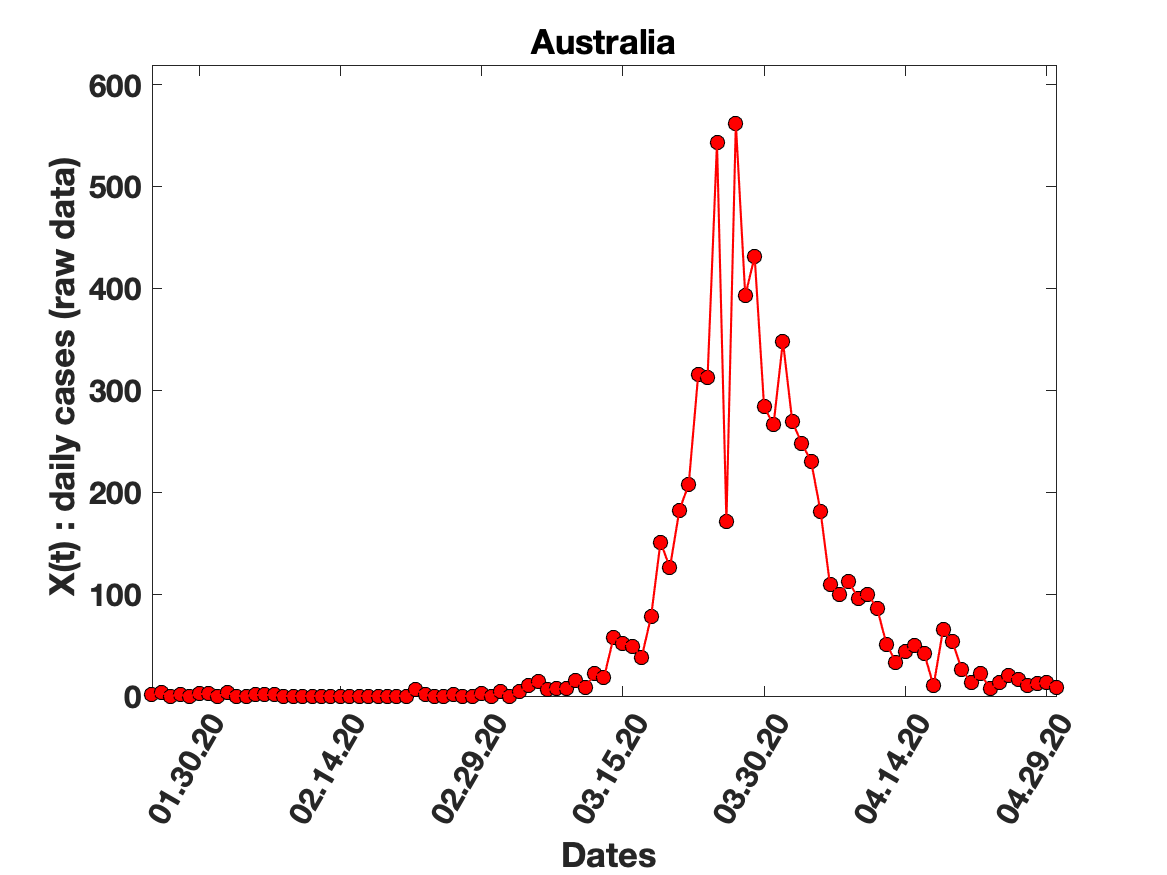

### Austria_solution_fit.png

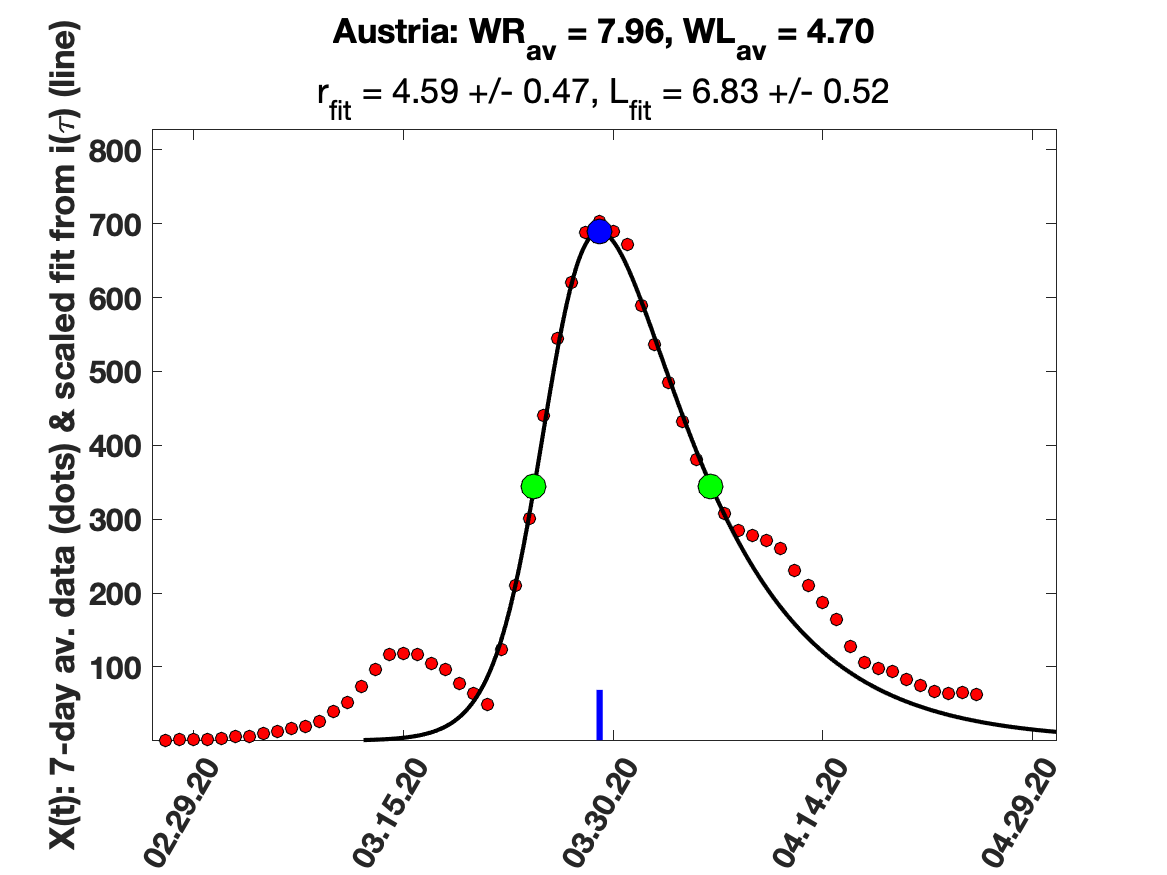

### Belgium_5_day_av_and_fit.png

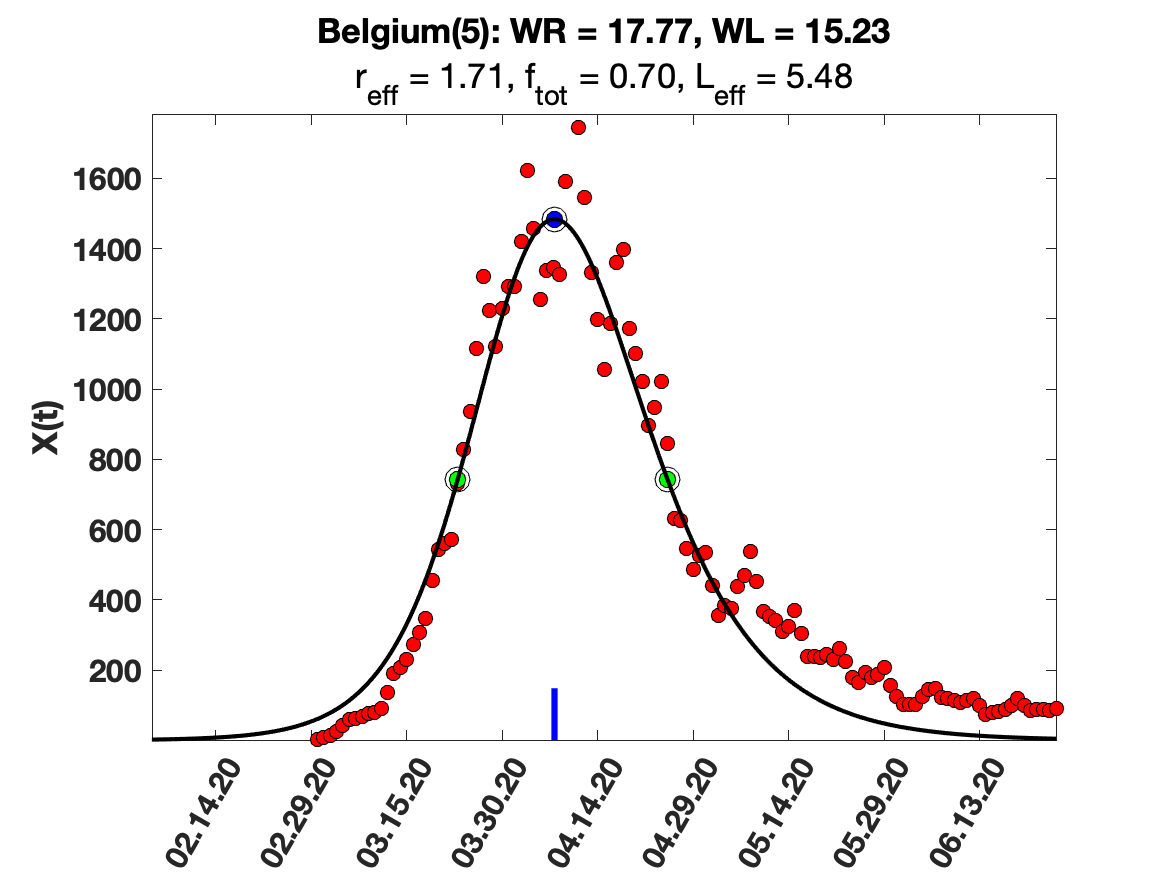

### Belgium_i(tau)_for_average_r.png

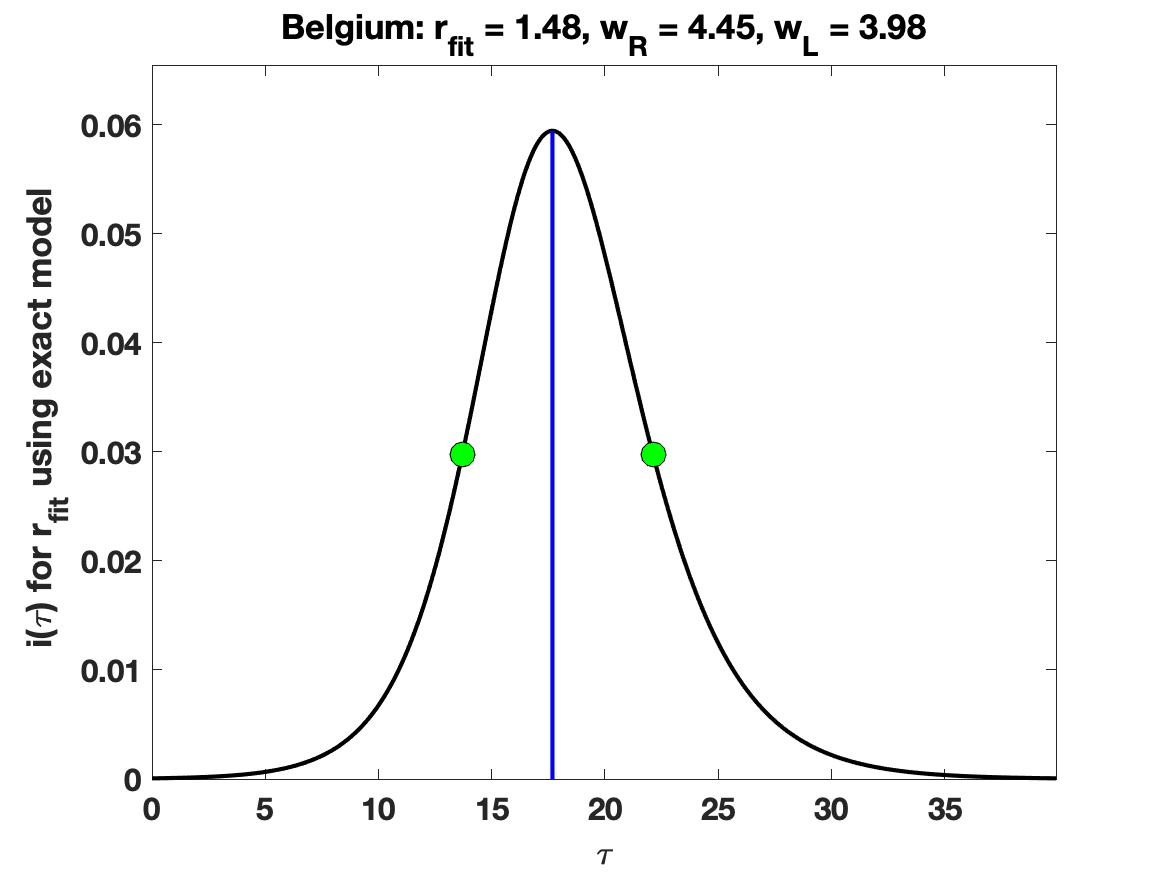

### Egypt_raw_data.png

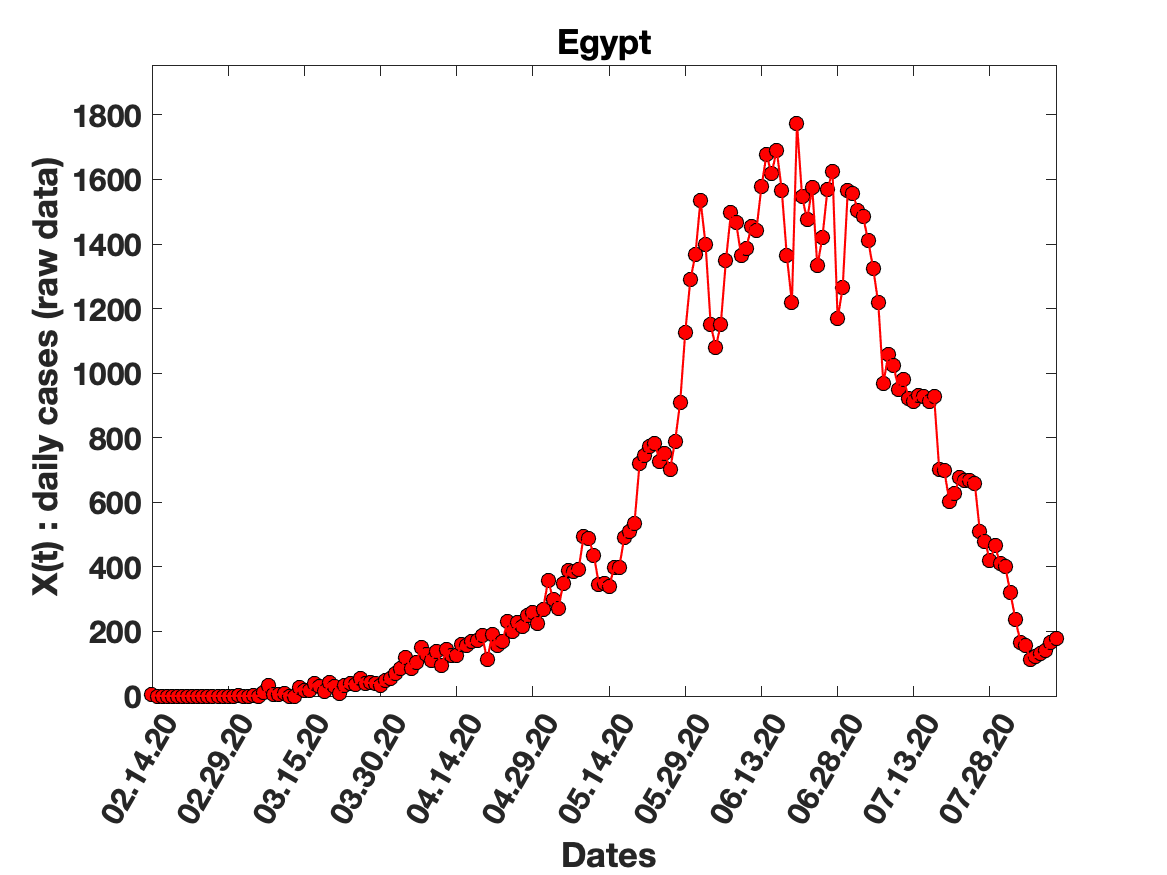

### France Tests .png

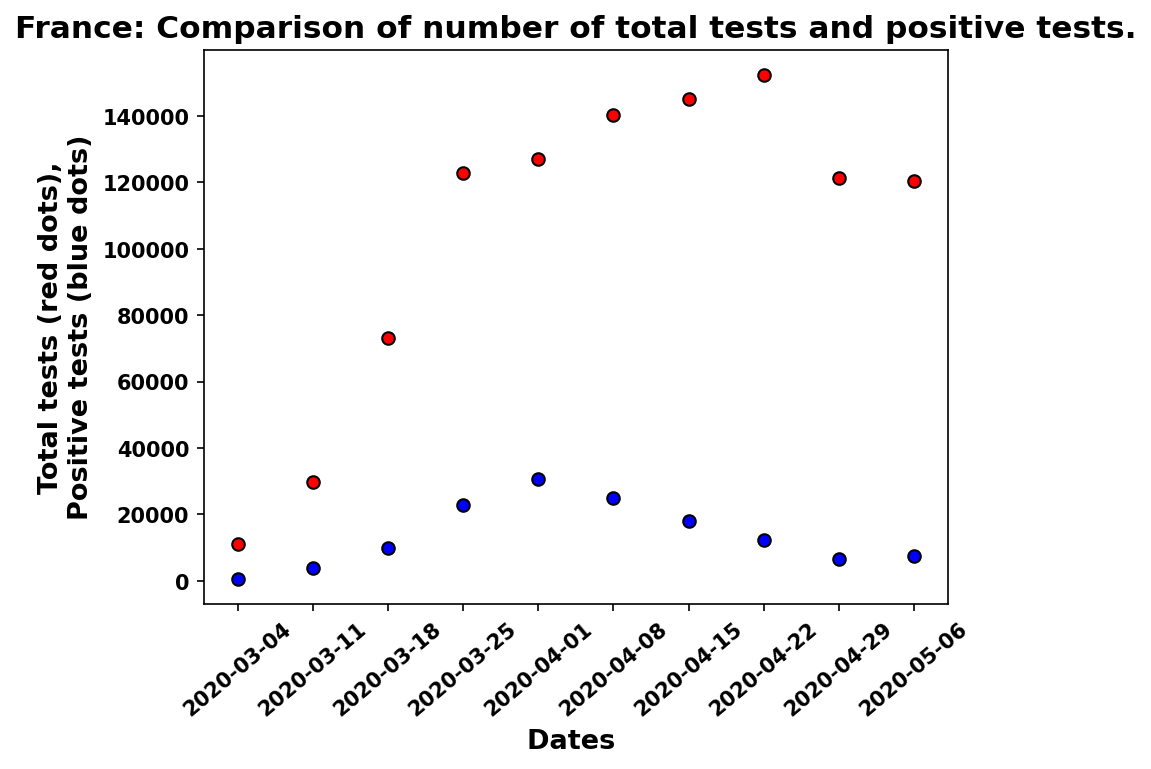

### France_7_day_av_and_fit.png

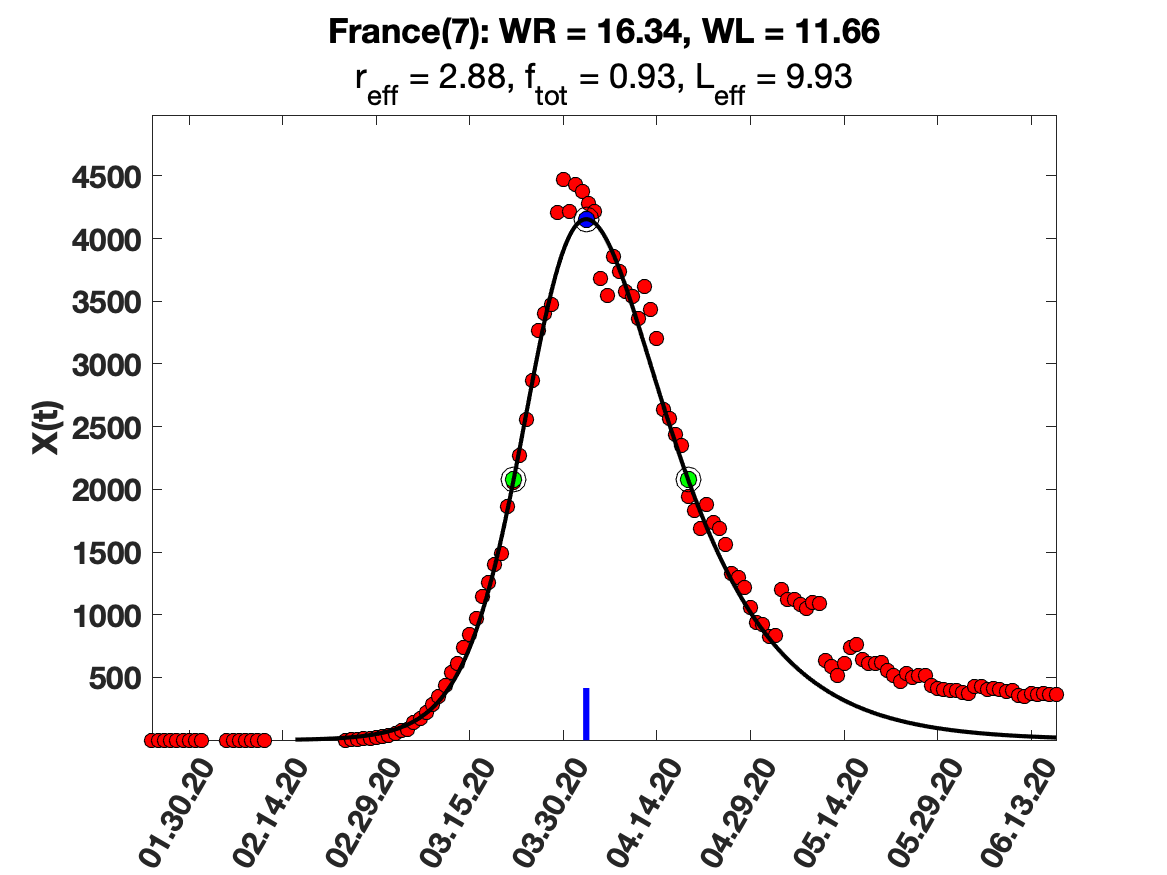

### France_9_day_av_and_fit.png

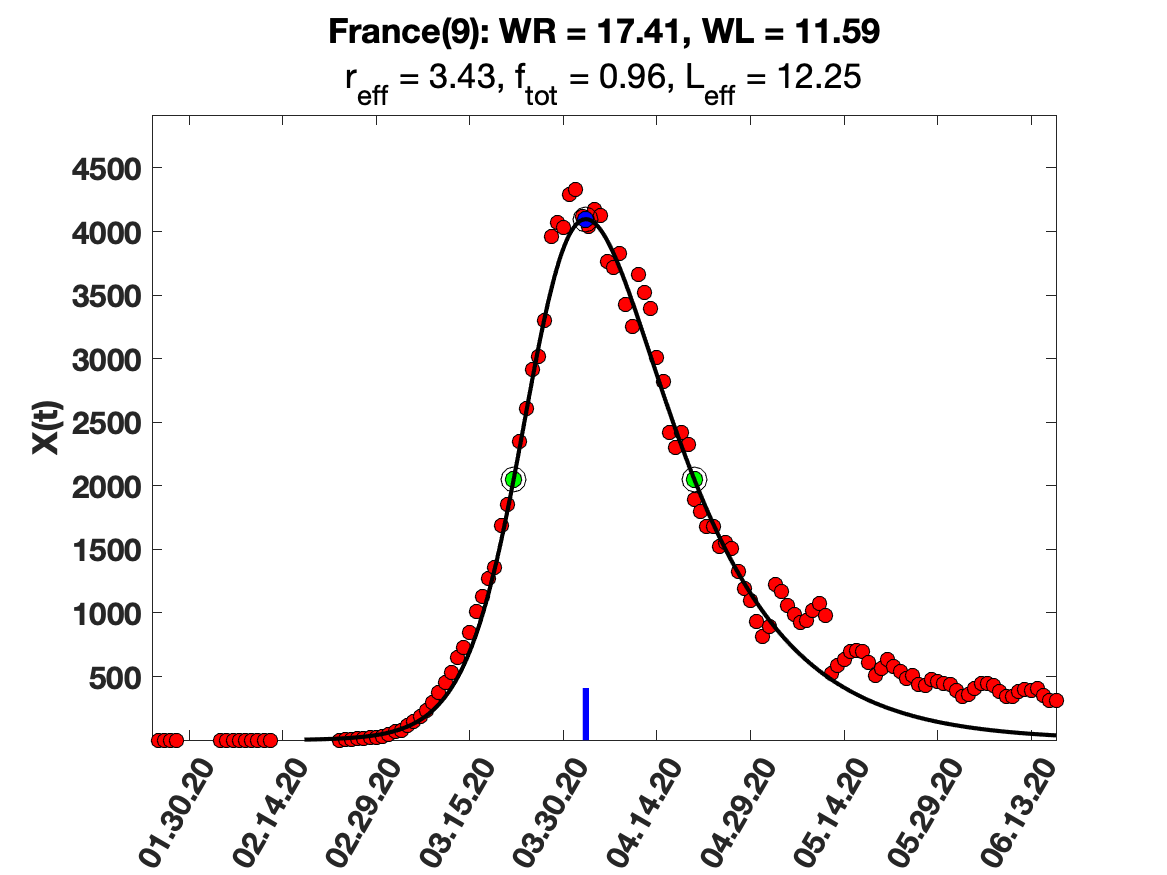

### Germany Tests .png

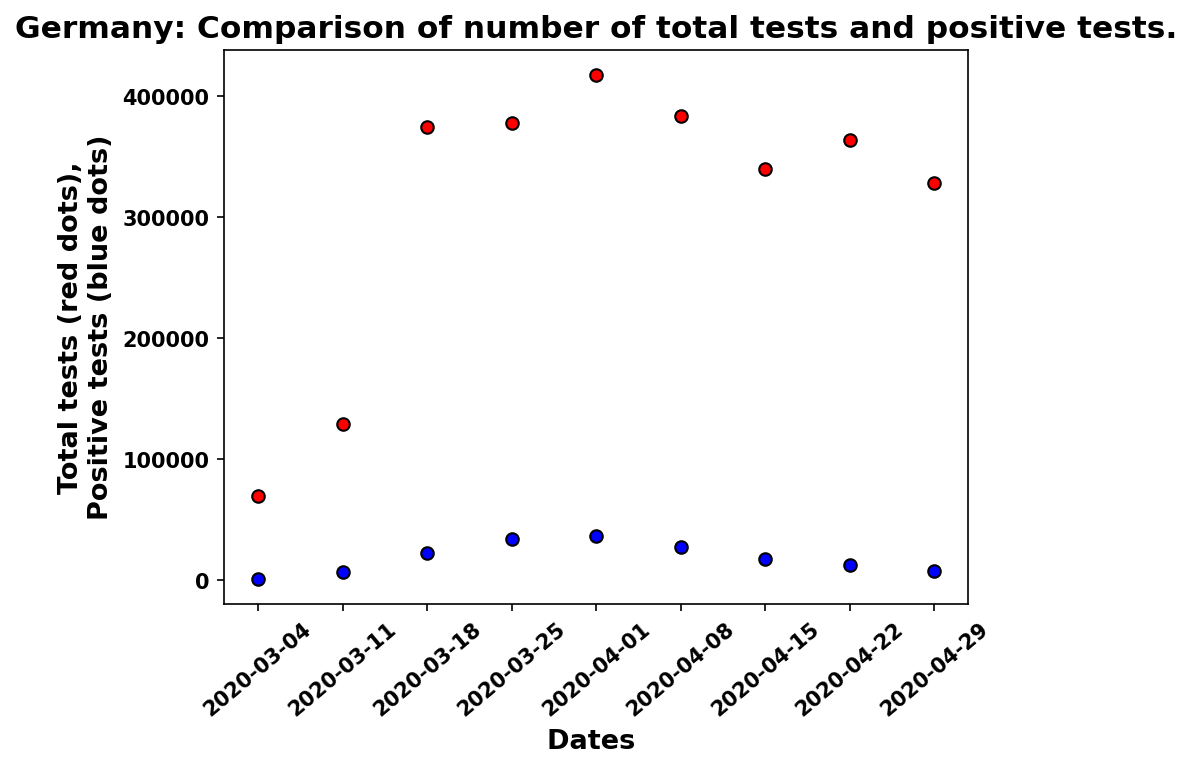

### Germany_7_day_av_and_fit.png

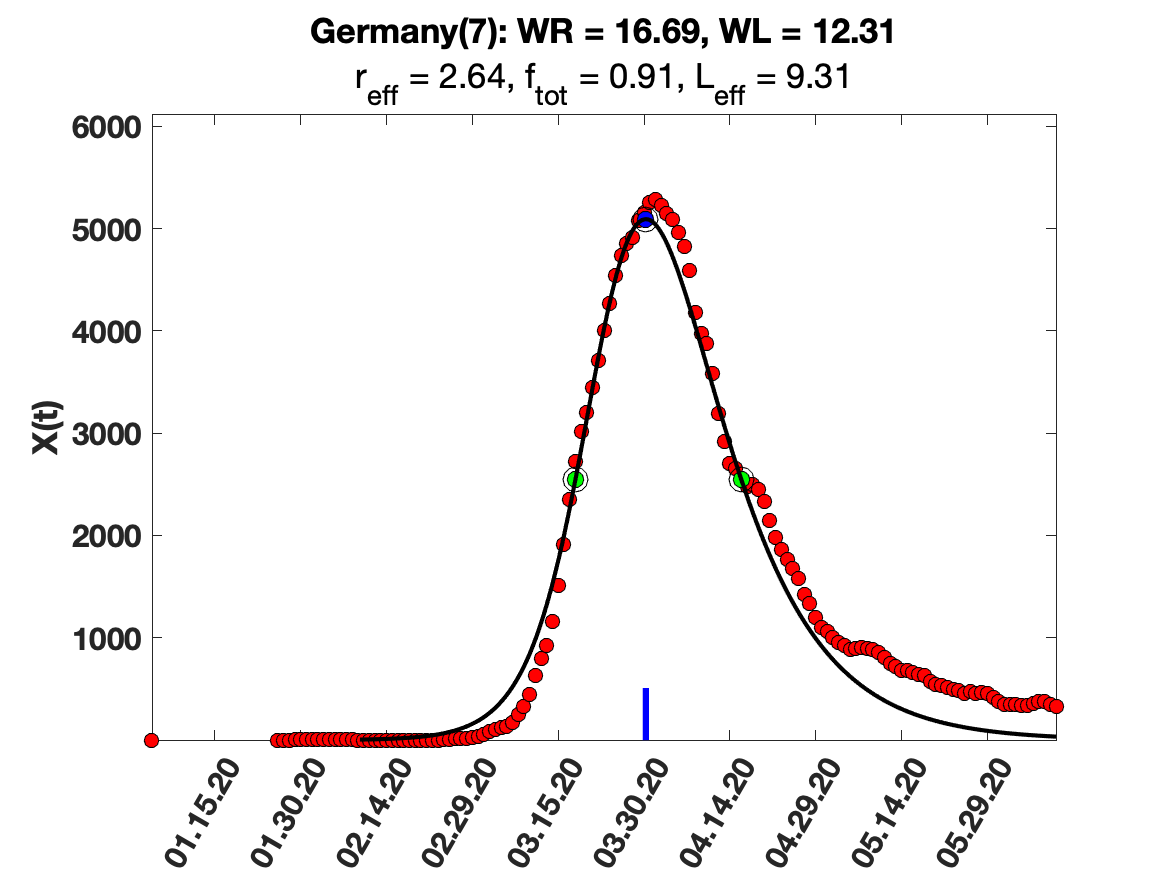

### Israel_raw_data.png

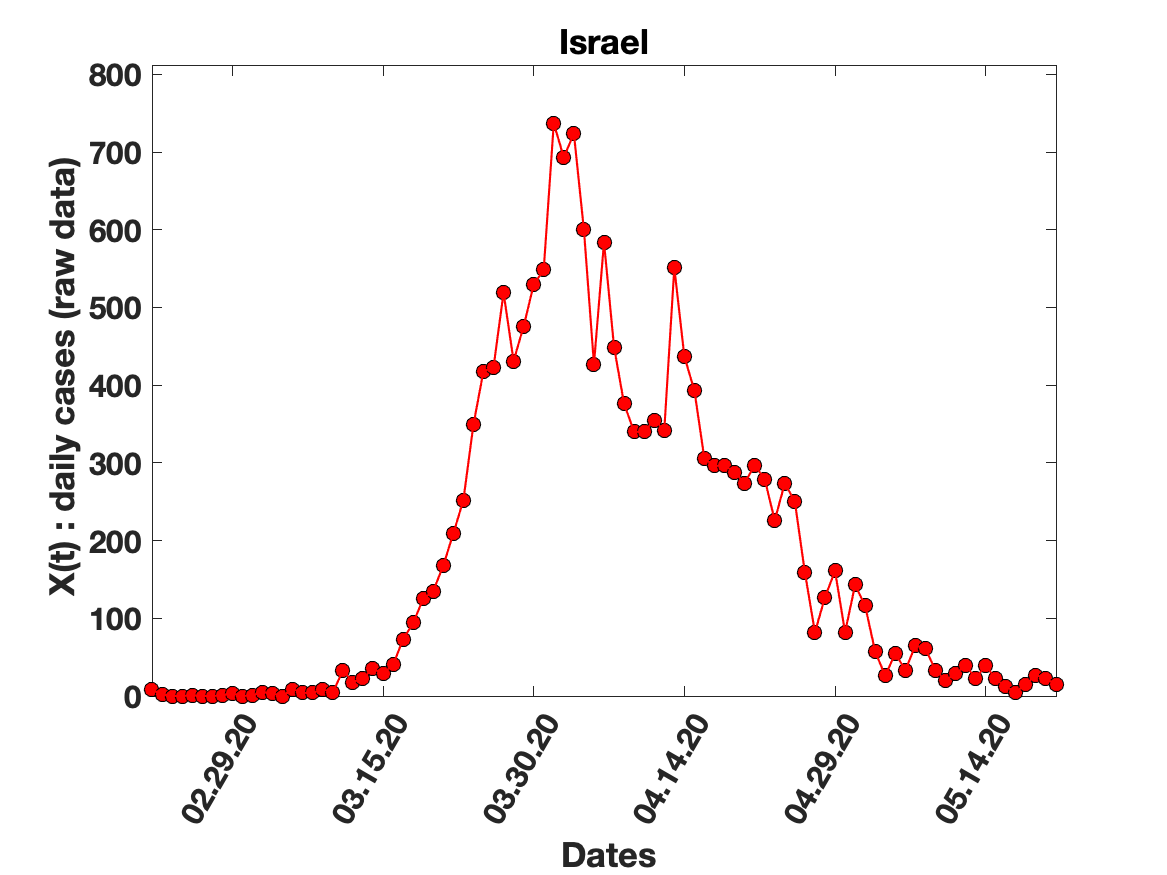

### Italy Tests .png

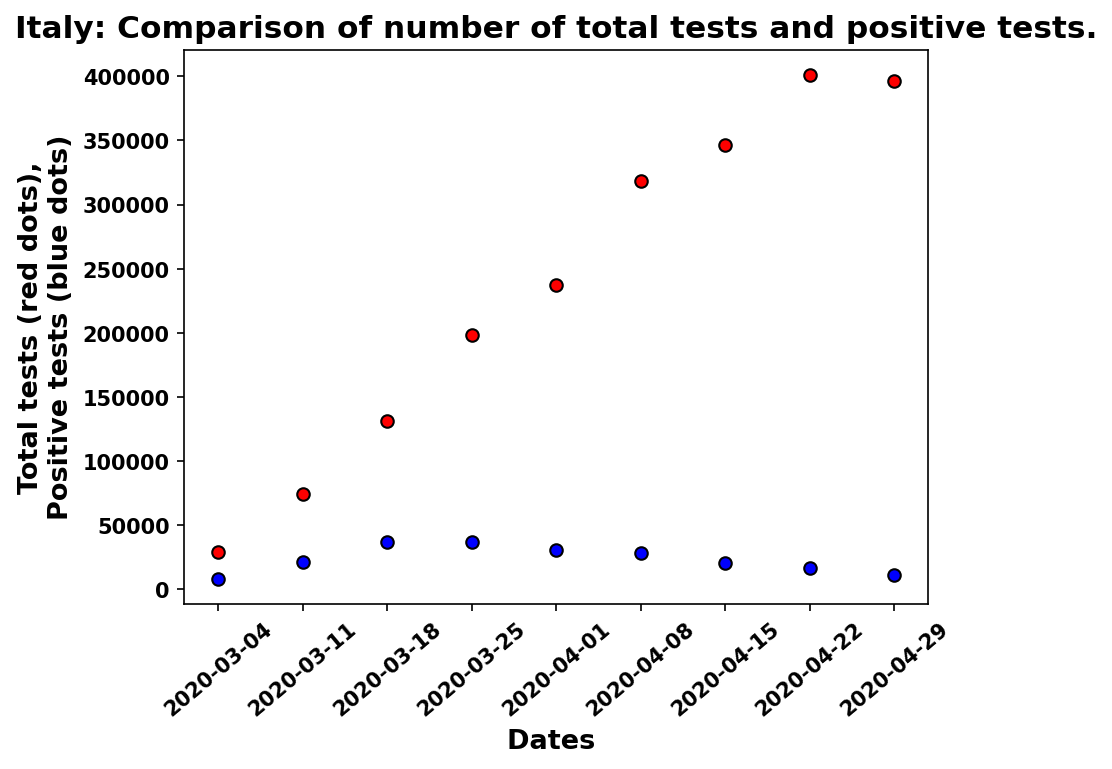

### Japan_5_day_av_and_fit.png

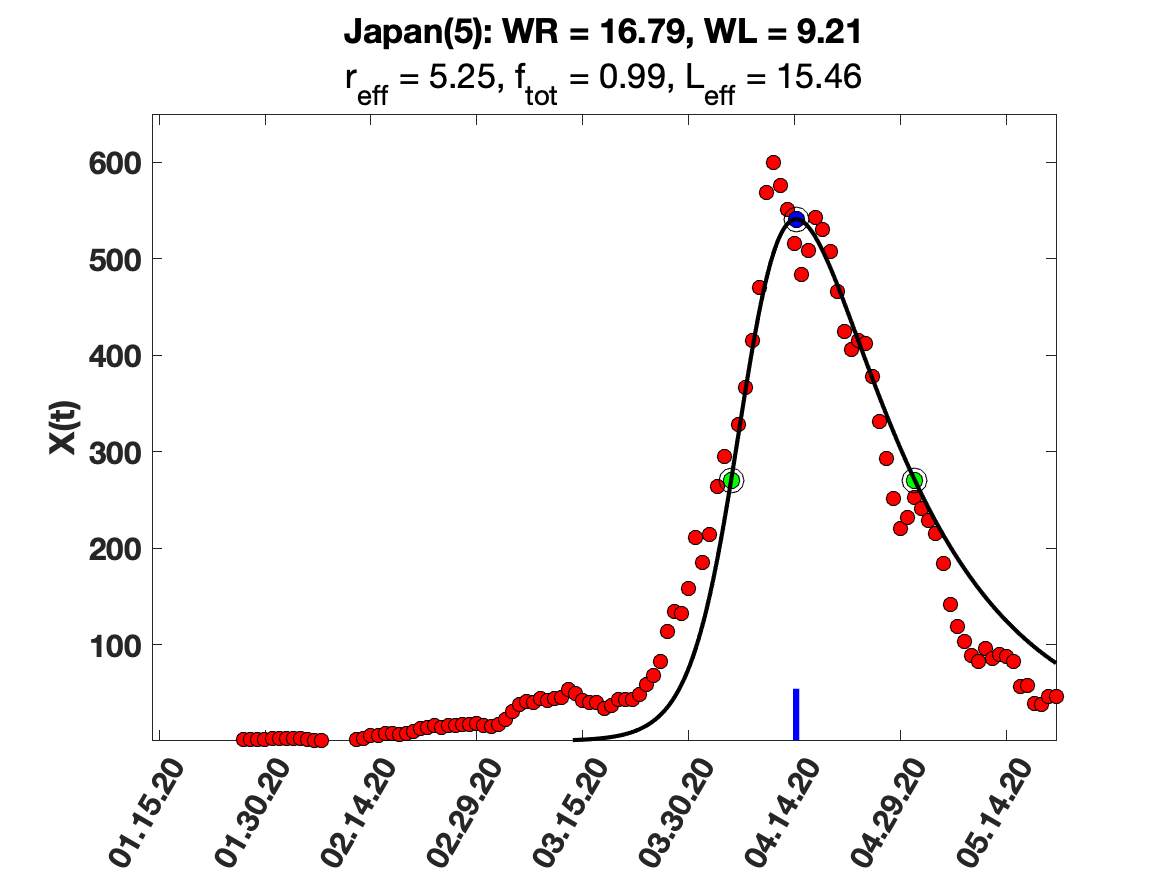

### Netherlands_7_day_av_and_fit.png

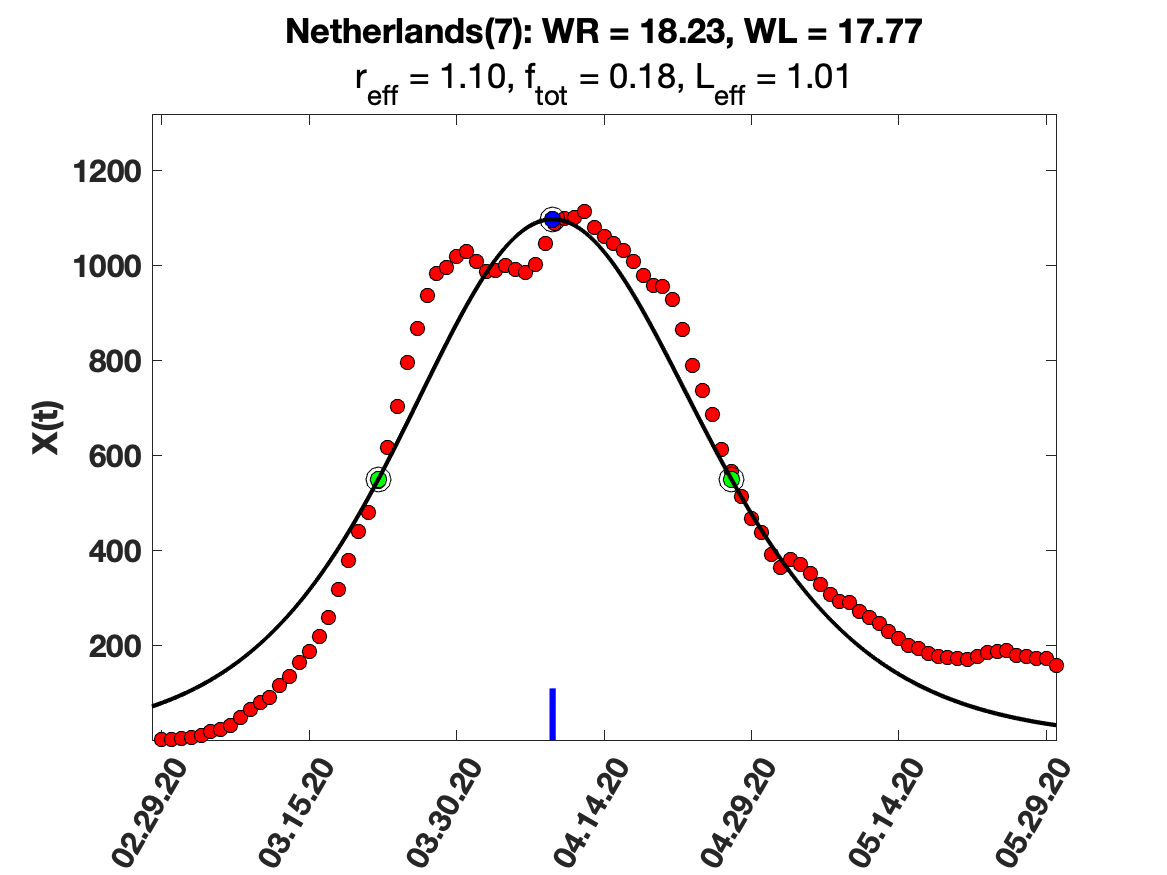

### Netherlands_9_day_av_and_fit.png

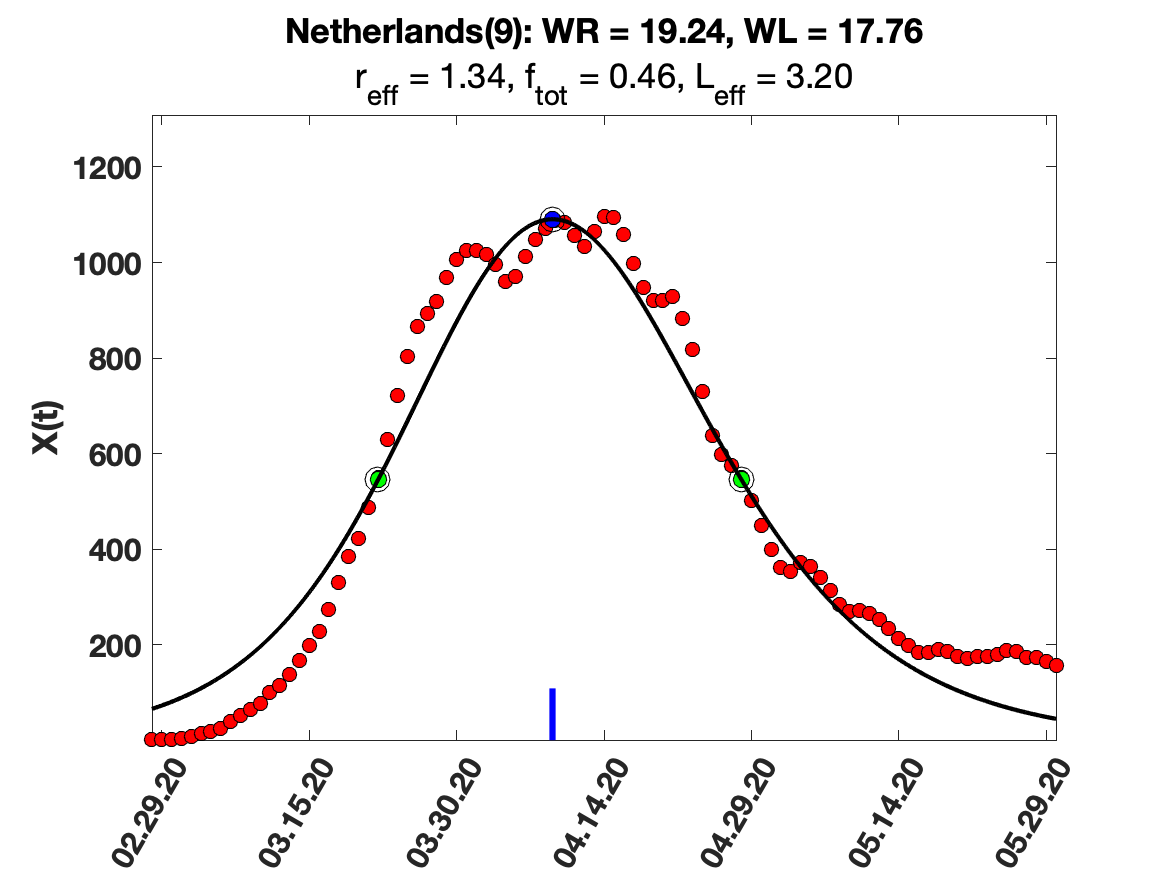

### New_Zealand_solution_fit.png

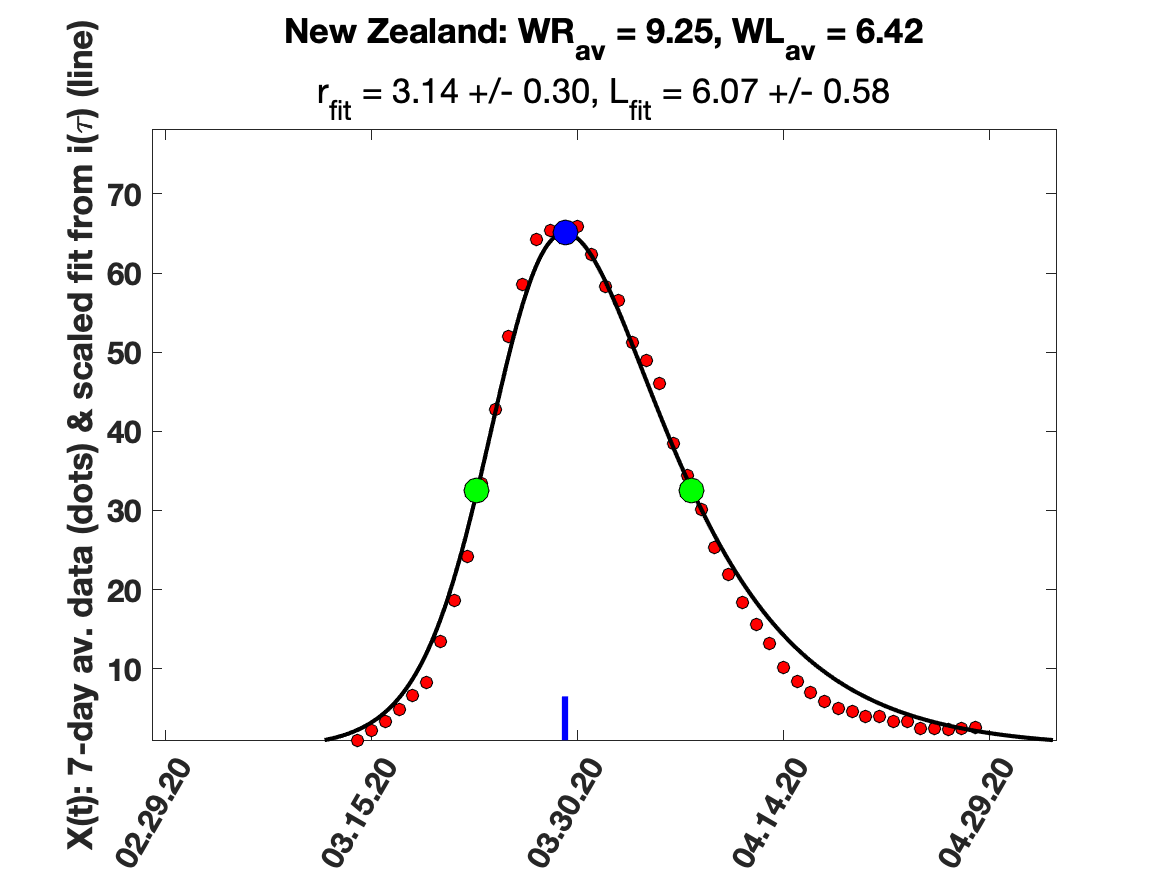

### Portugal_5_day_av_and_fit.png

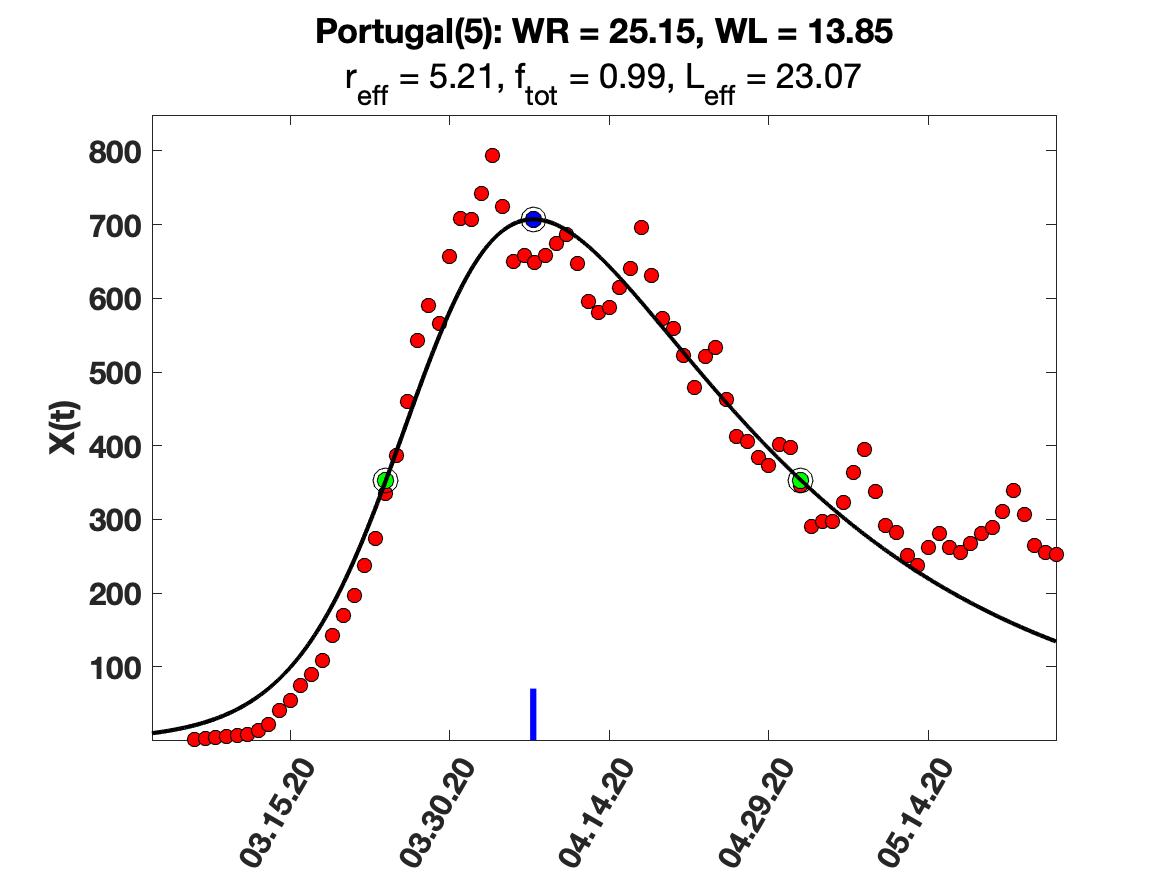

### Qatar_5_day_av_and_fit.png

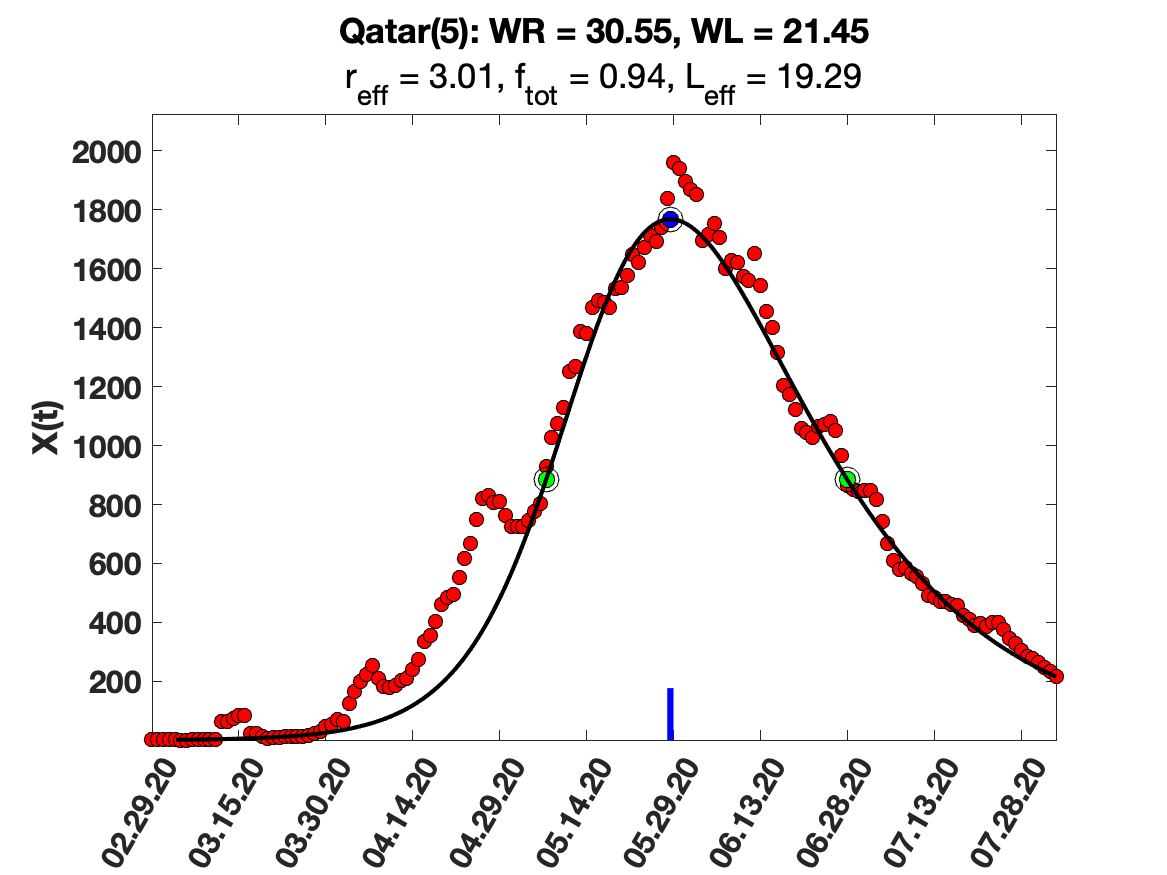

### Qatar_raw_data.png

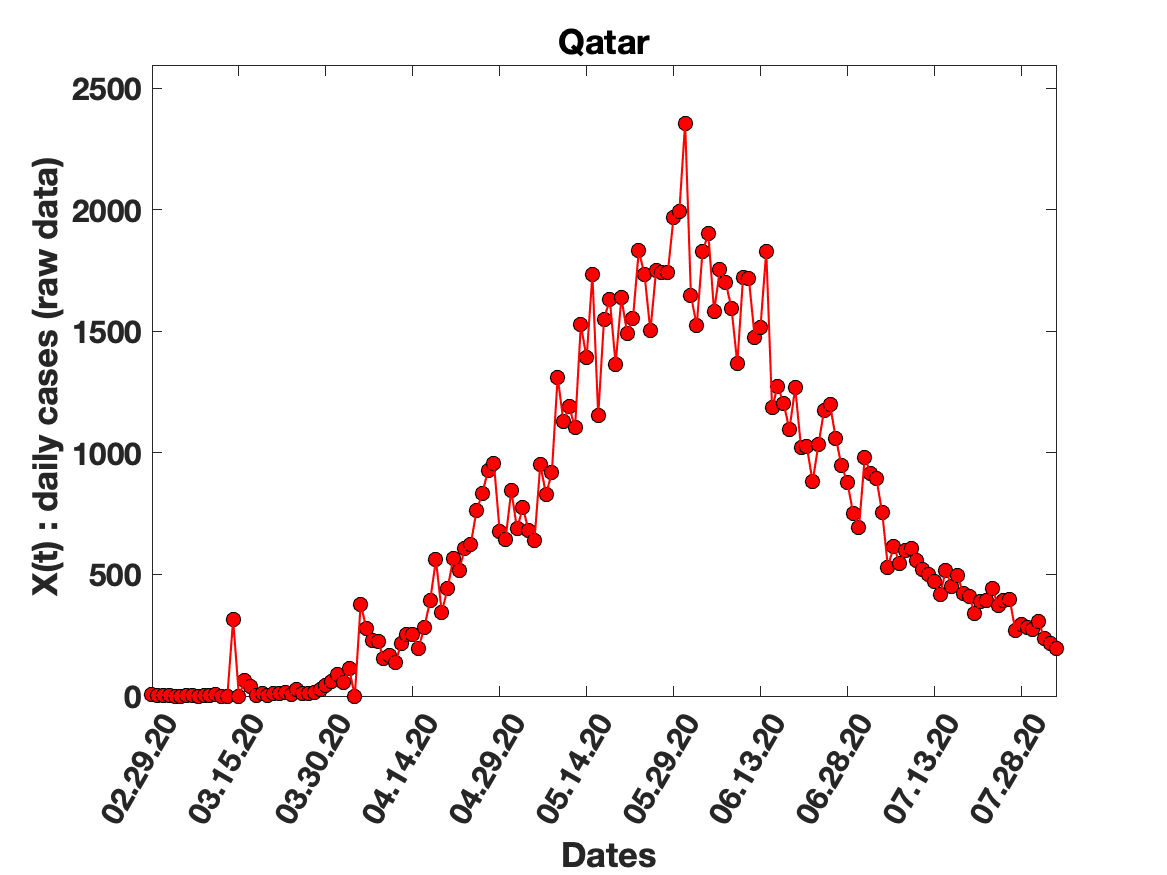

### Qatar_solution_fit.png

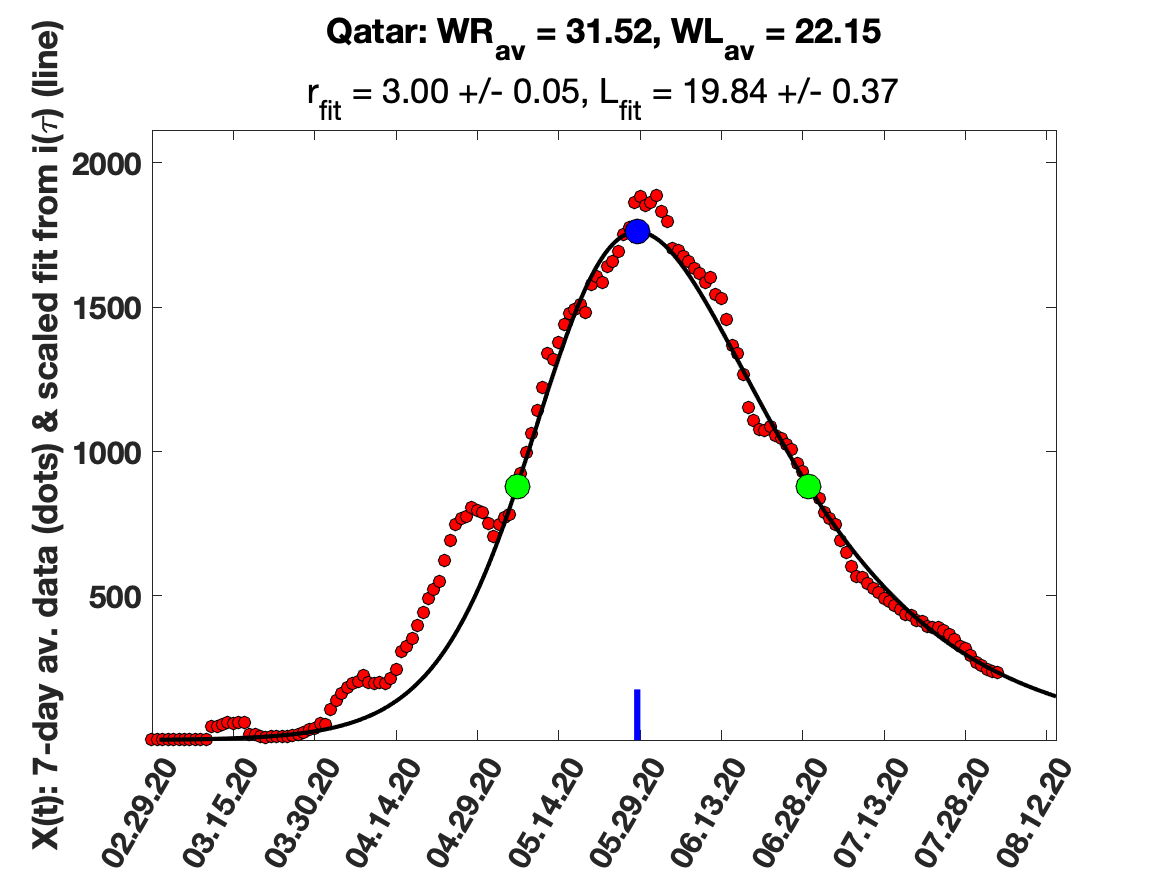

### South_Afrrica_9_day_av_and_fit.png

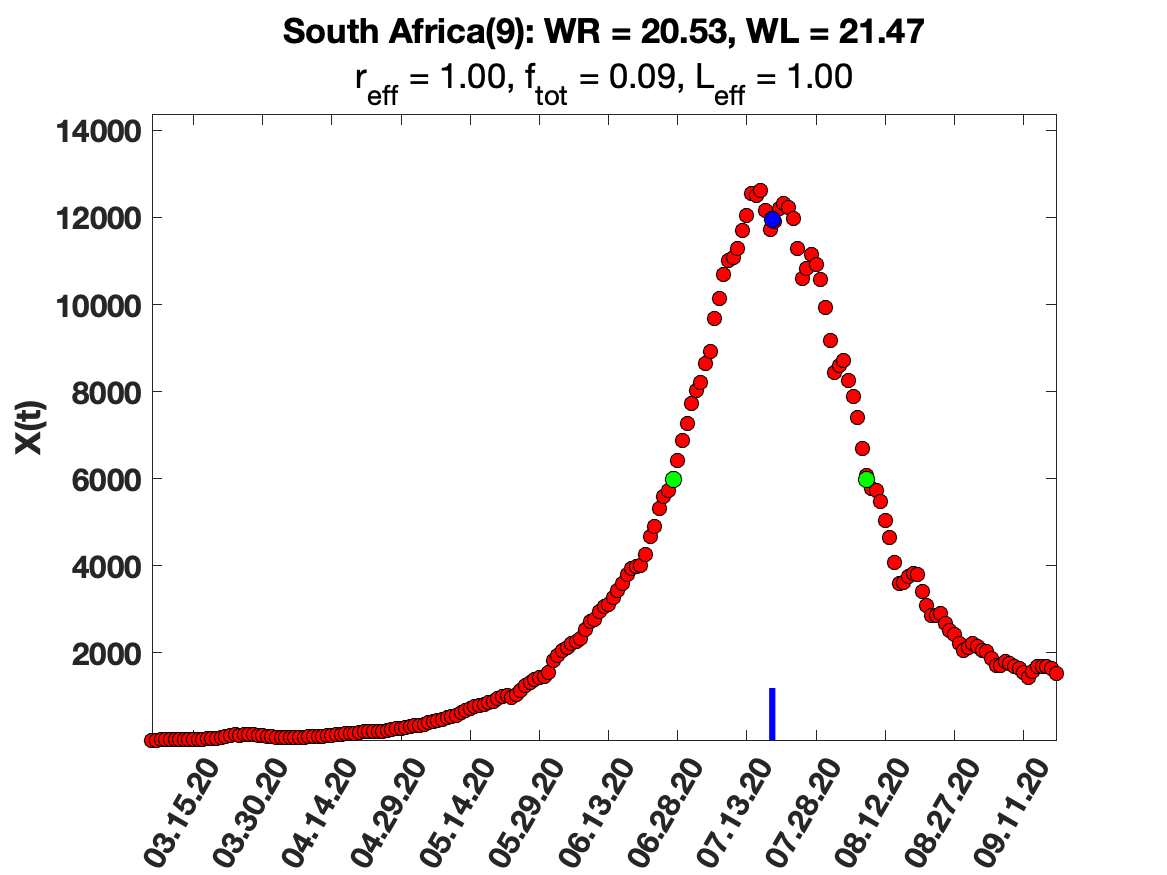

### South_Afrrica_solution_fit.png

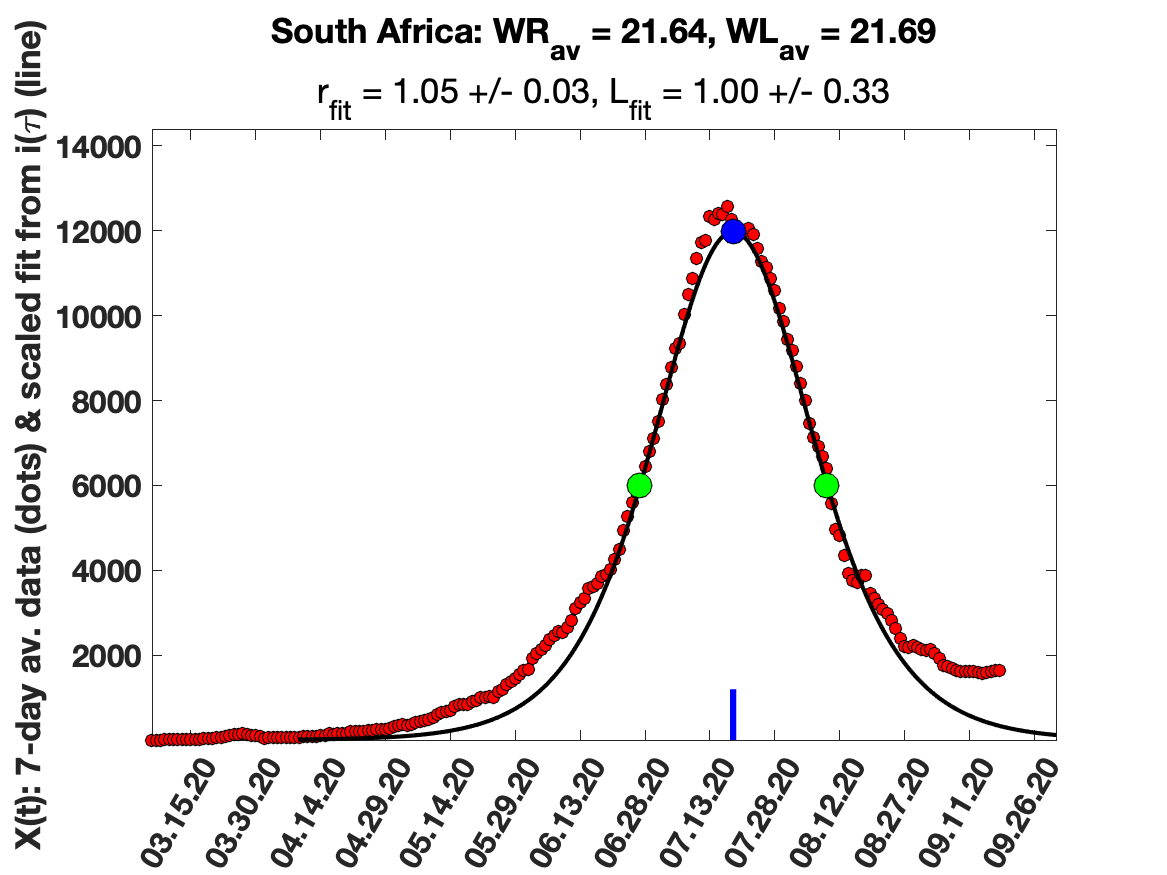

### Spain_i(tau)_for_average_r.png

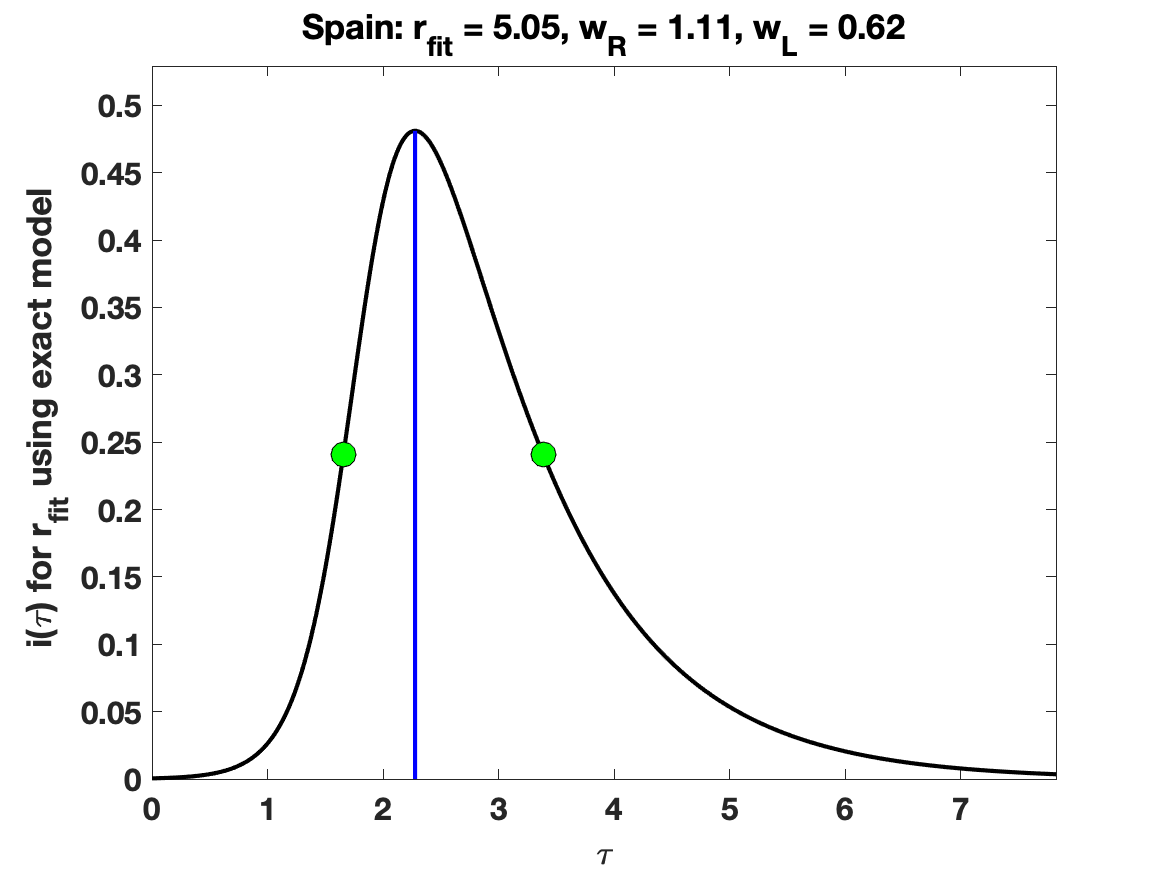

### Sup fig 3a.png

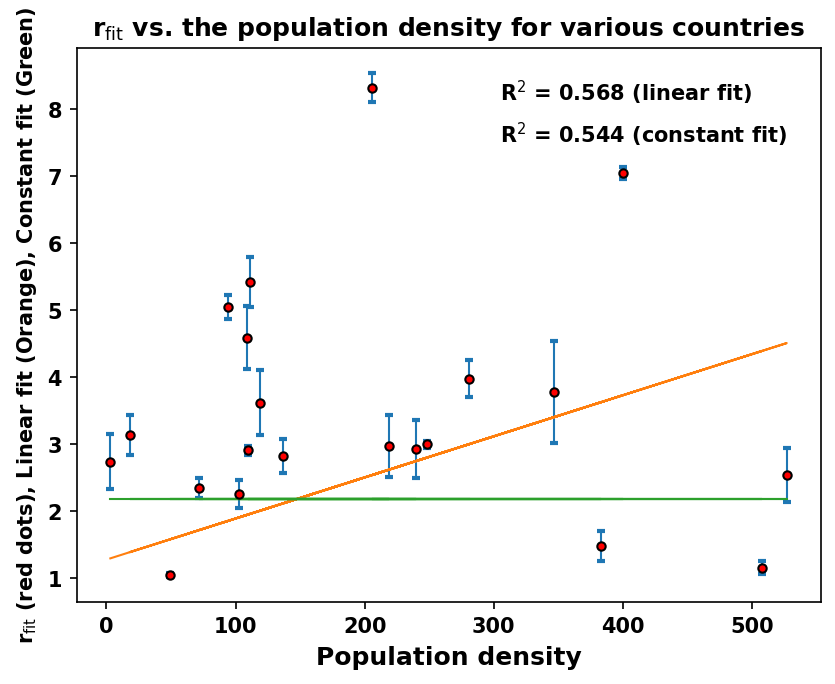

### Sup fig 3b.png

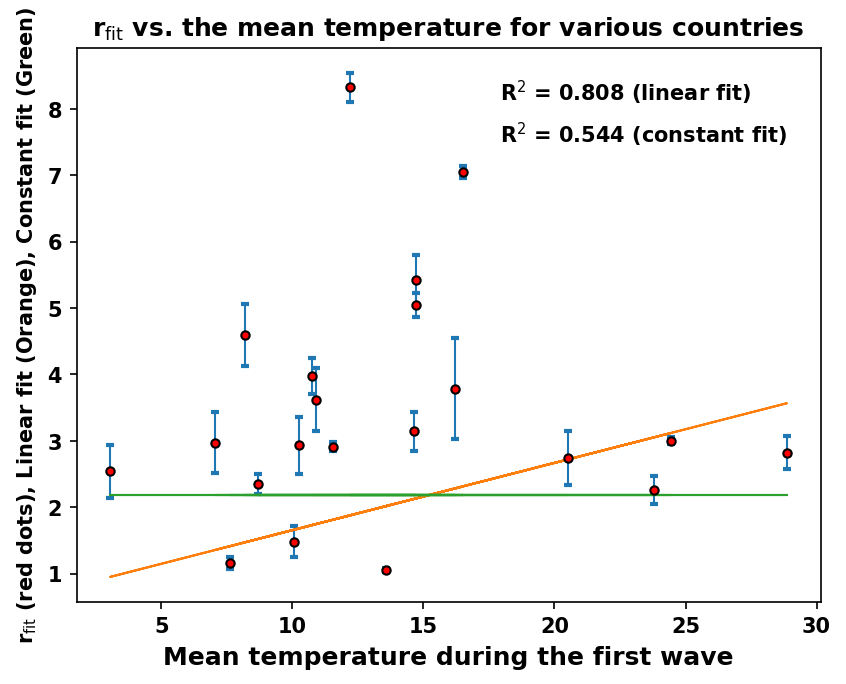

### Sup fig 3c.png

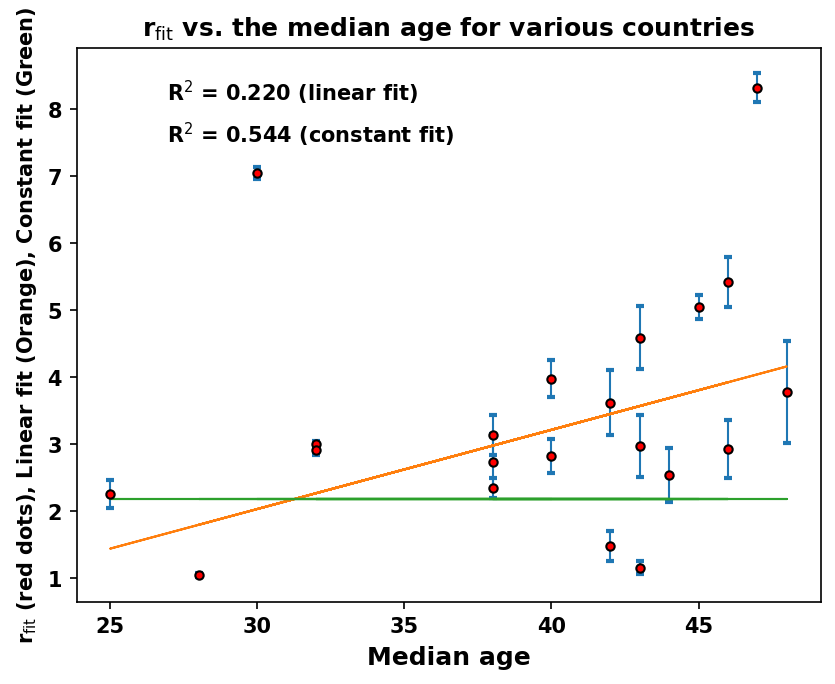

### Sup fig 3d.png

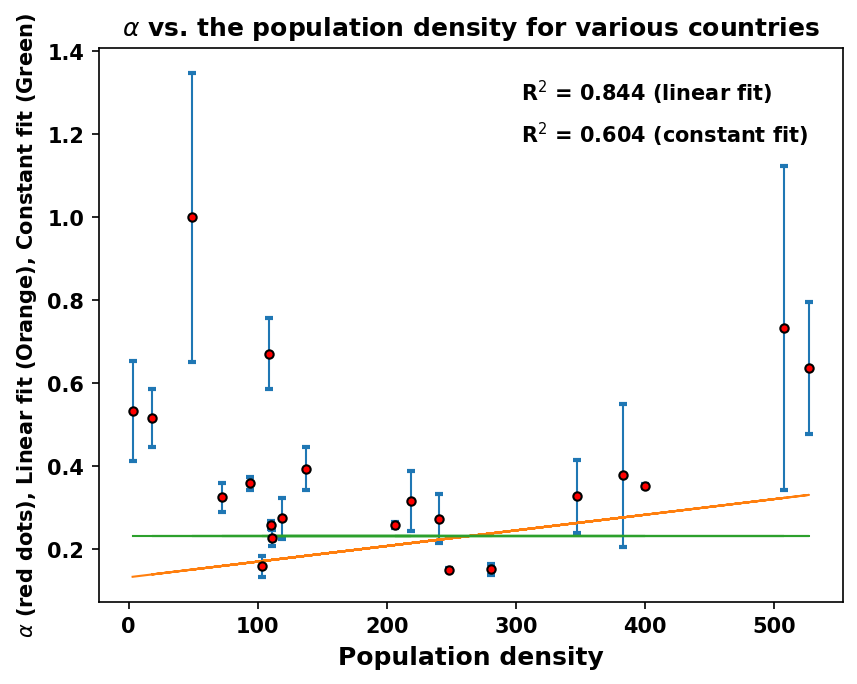

### Sup fig 3e.png

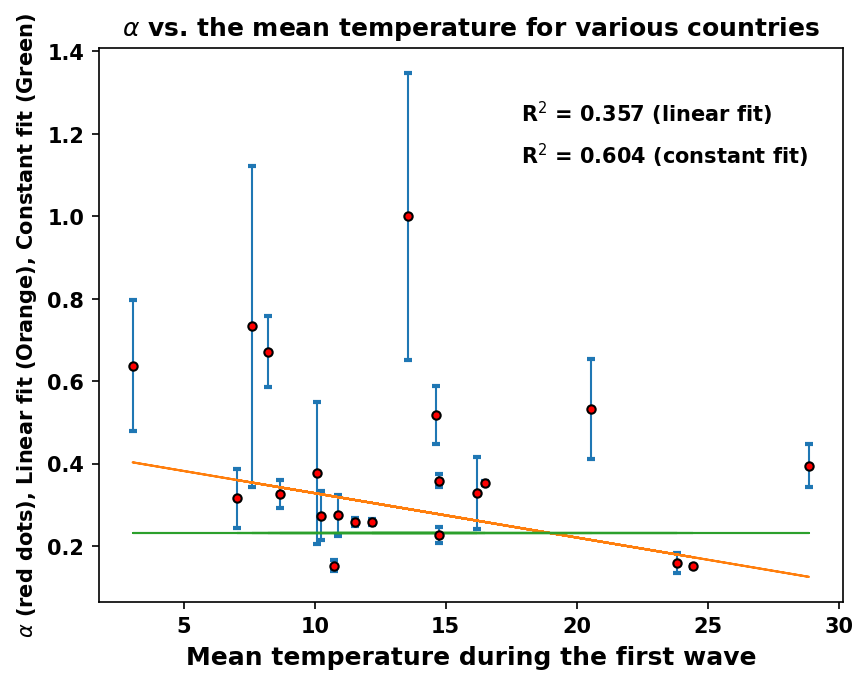

### Sup fig 3f.png

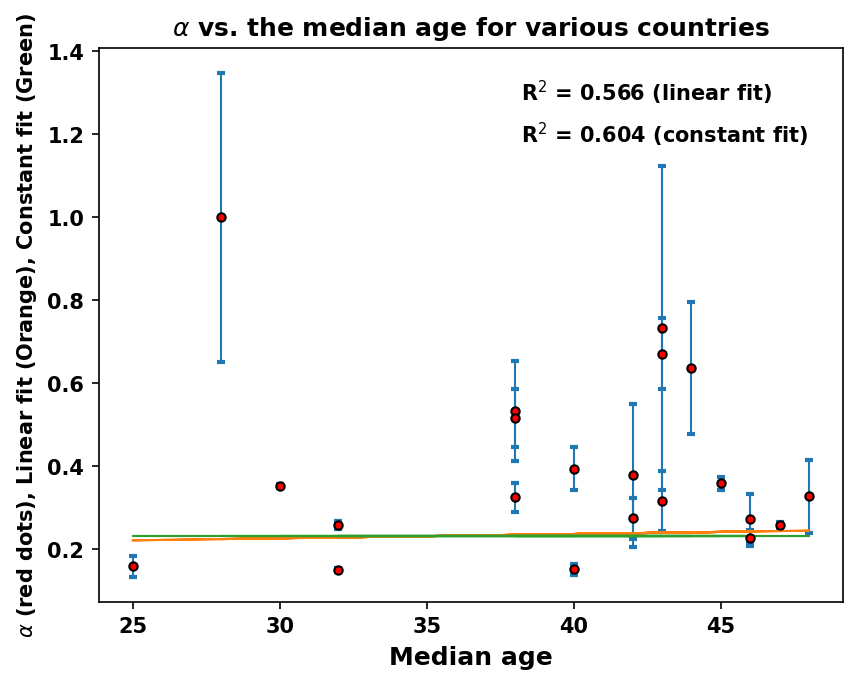

### Sup fig 3g.png

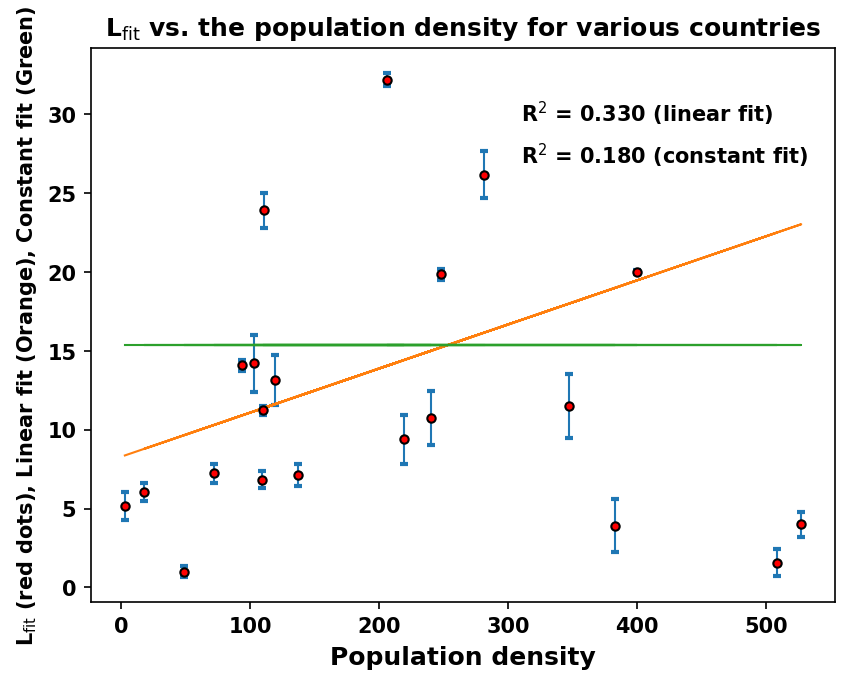

### Sup fig 3h.png

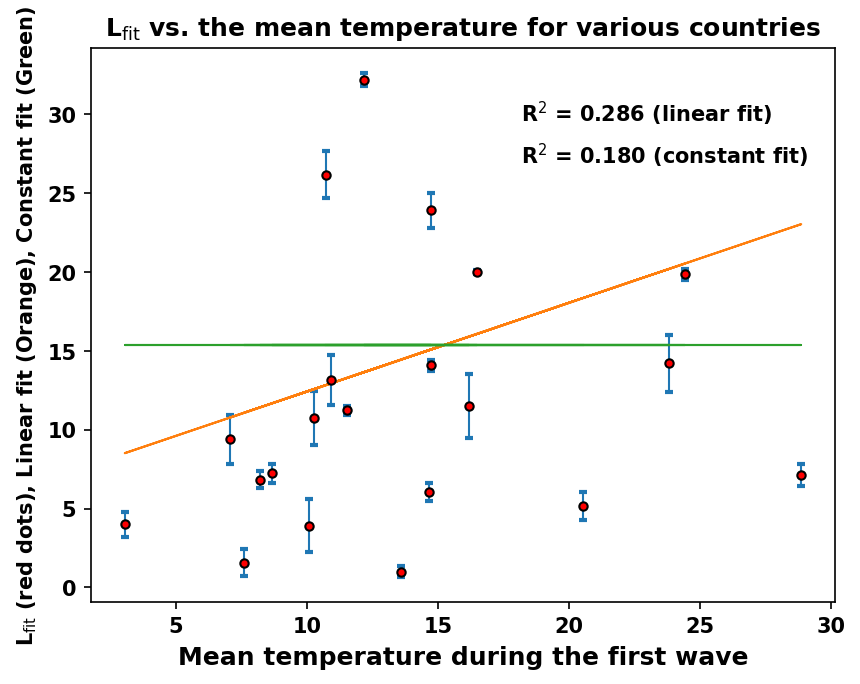

### Sup fig 3i.png

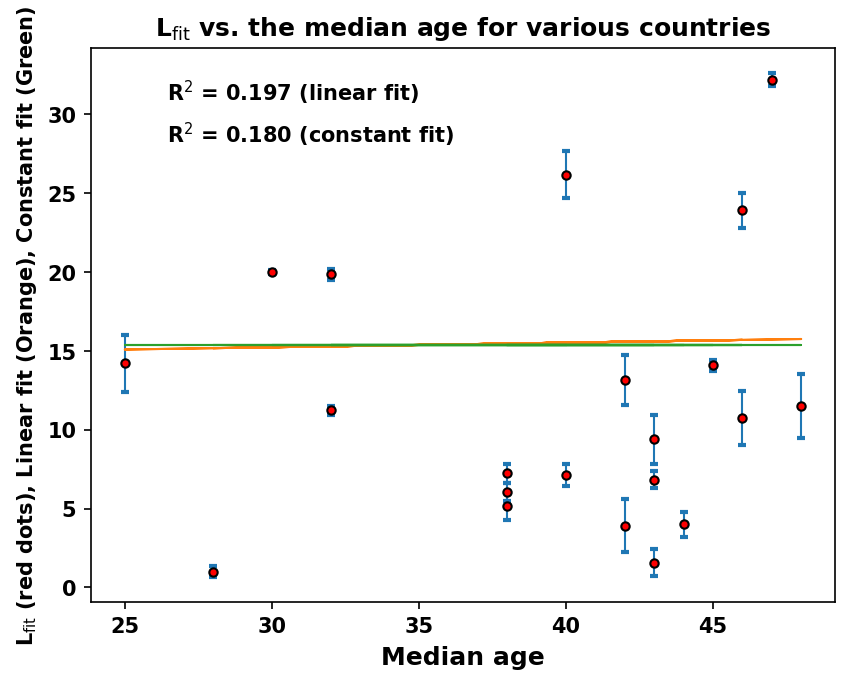

### Switzerland_7_day_av_and_fit.png

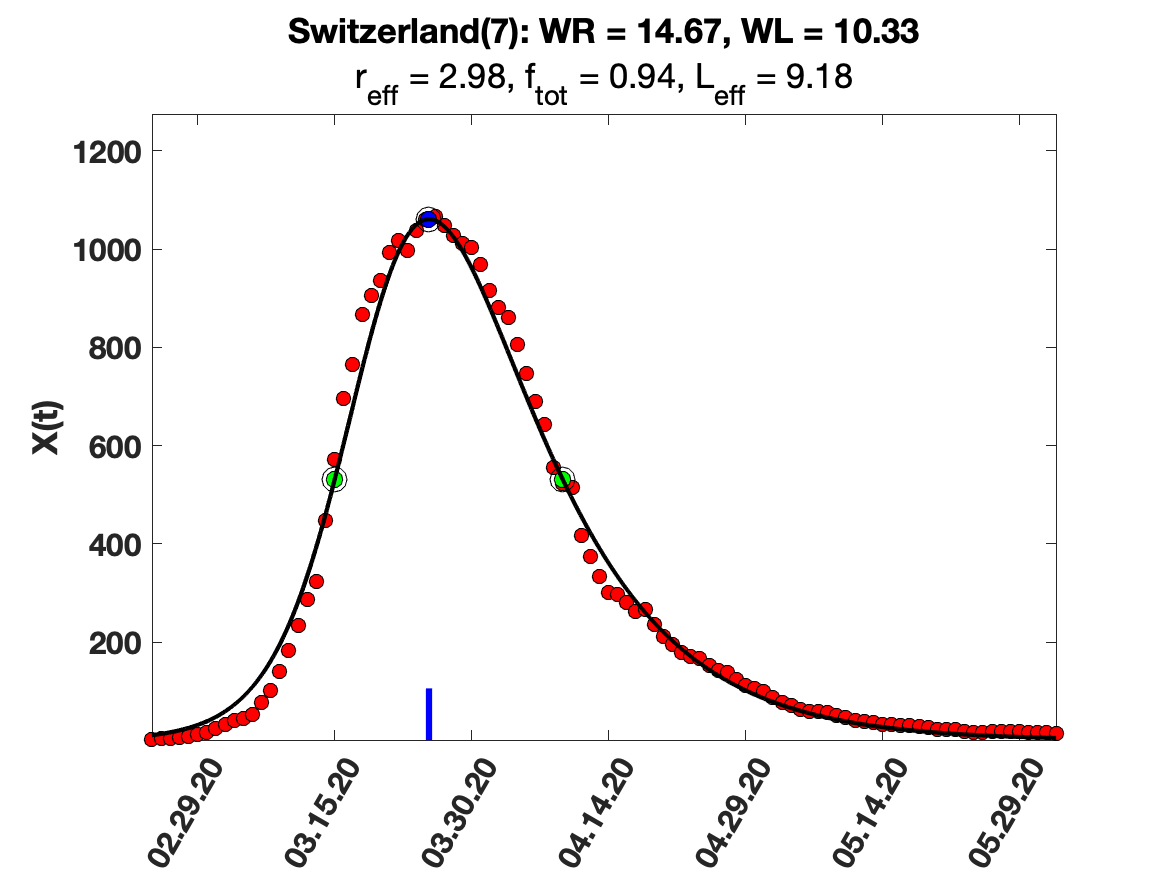

### Switzerland_solution_fit.png

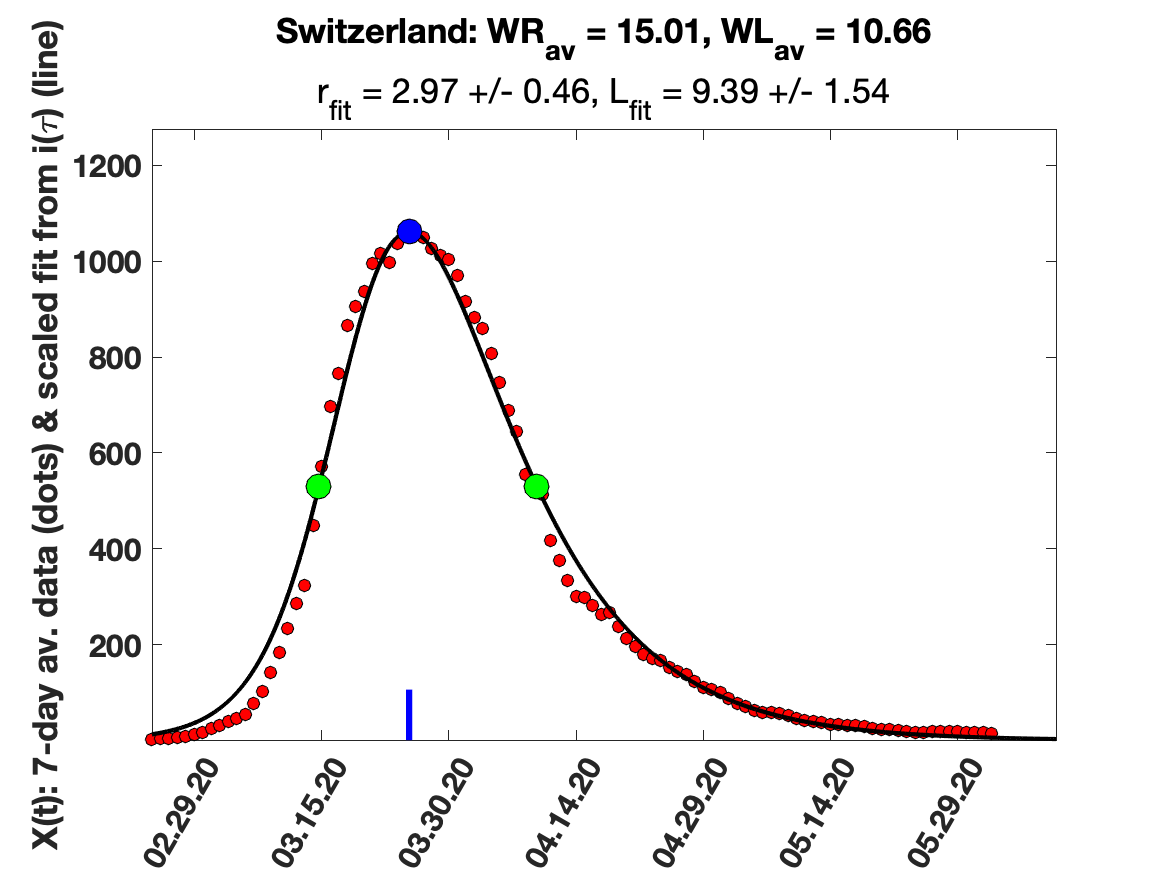

### Thailand_solution_fit.png

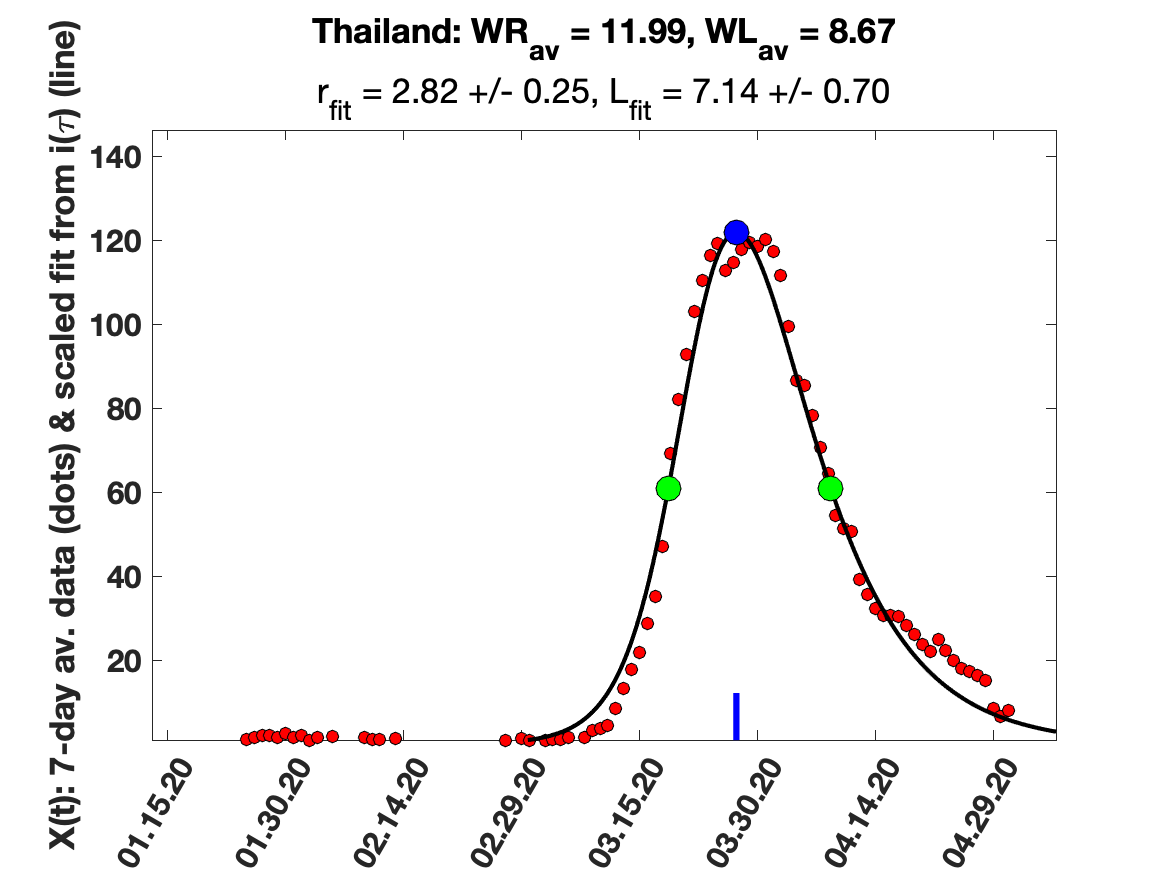

### Turkey_7_day_av_and_fit.png

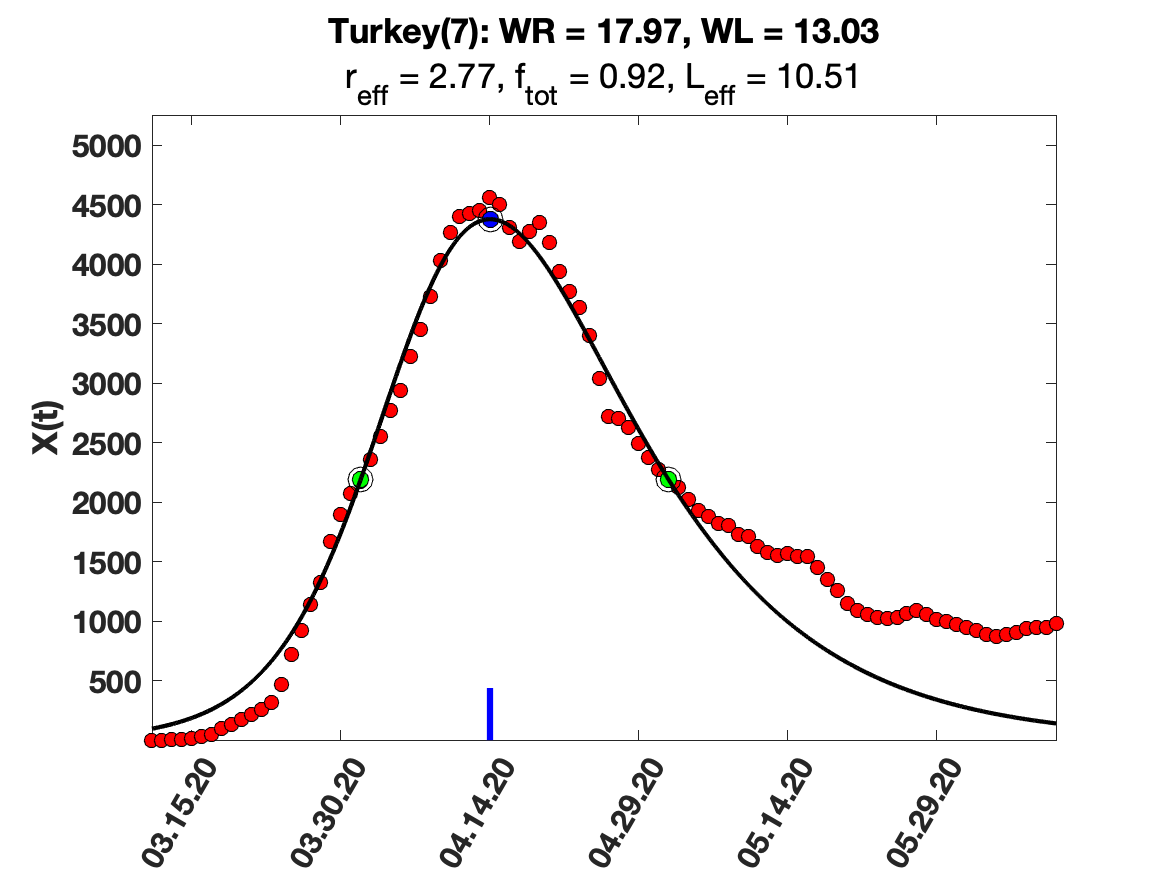

### Turkey_9_day_av_and_fit.png

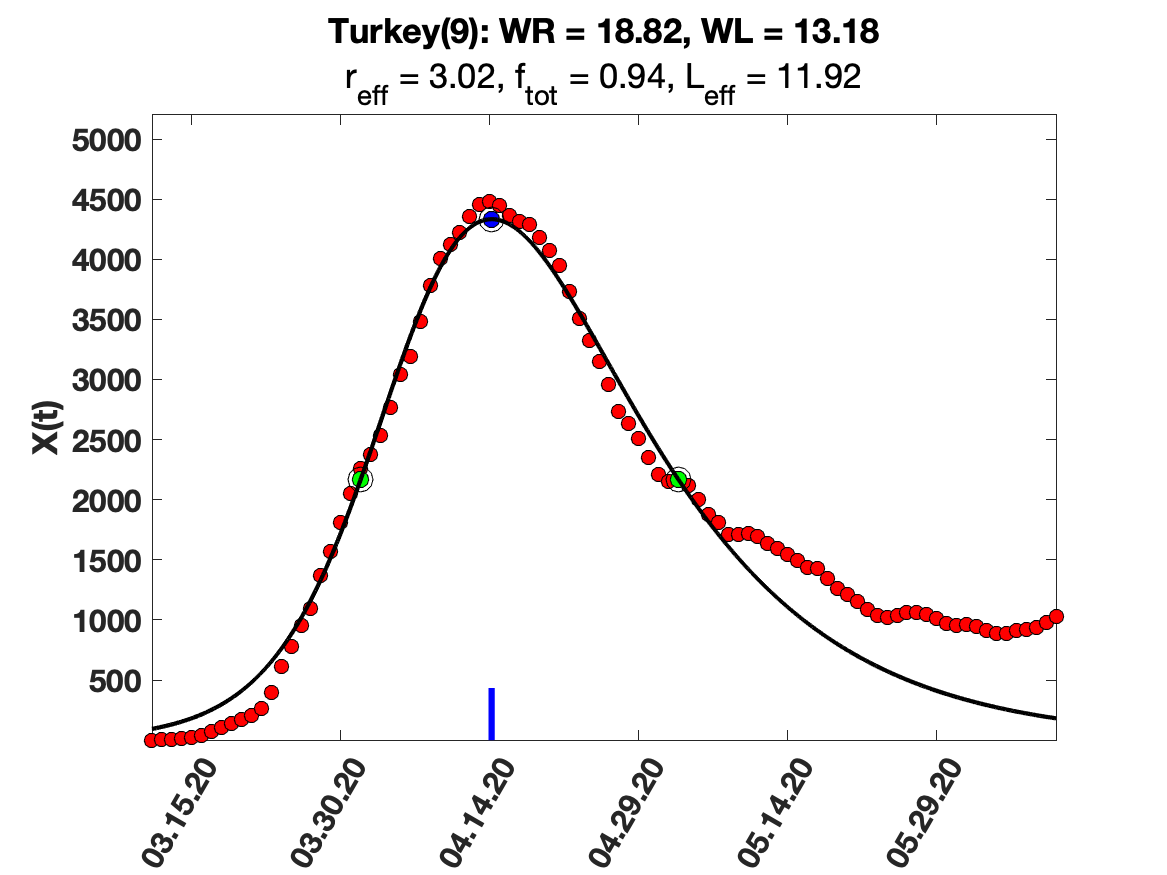

### UK_solution_fit.png

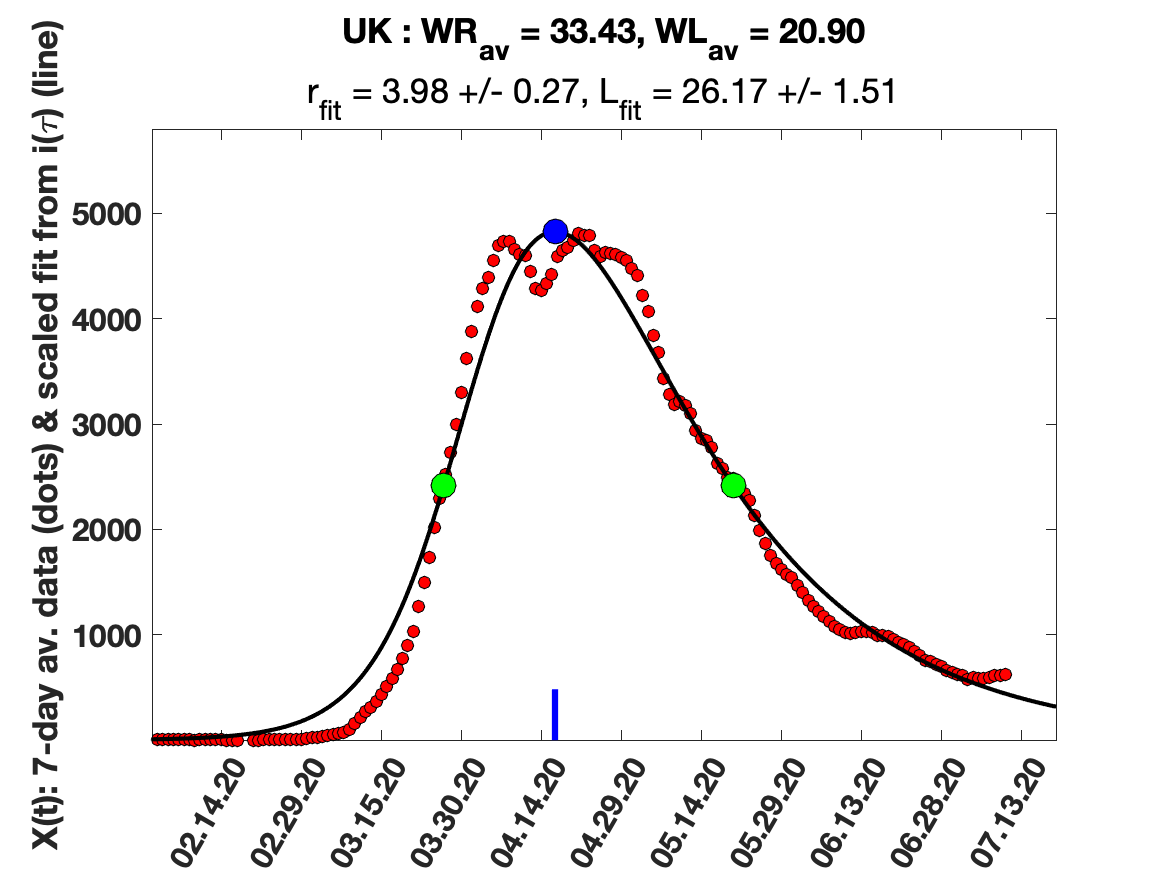
