## Supplementary figures and images for "Asymmetry in the peak in Covid-19 daily cases and the pandemic R-parameter"

### Australia_5_day_av_and_fit.png

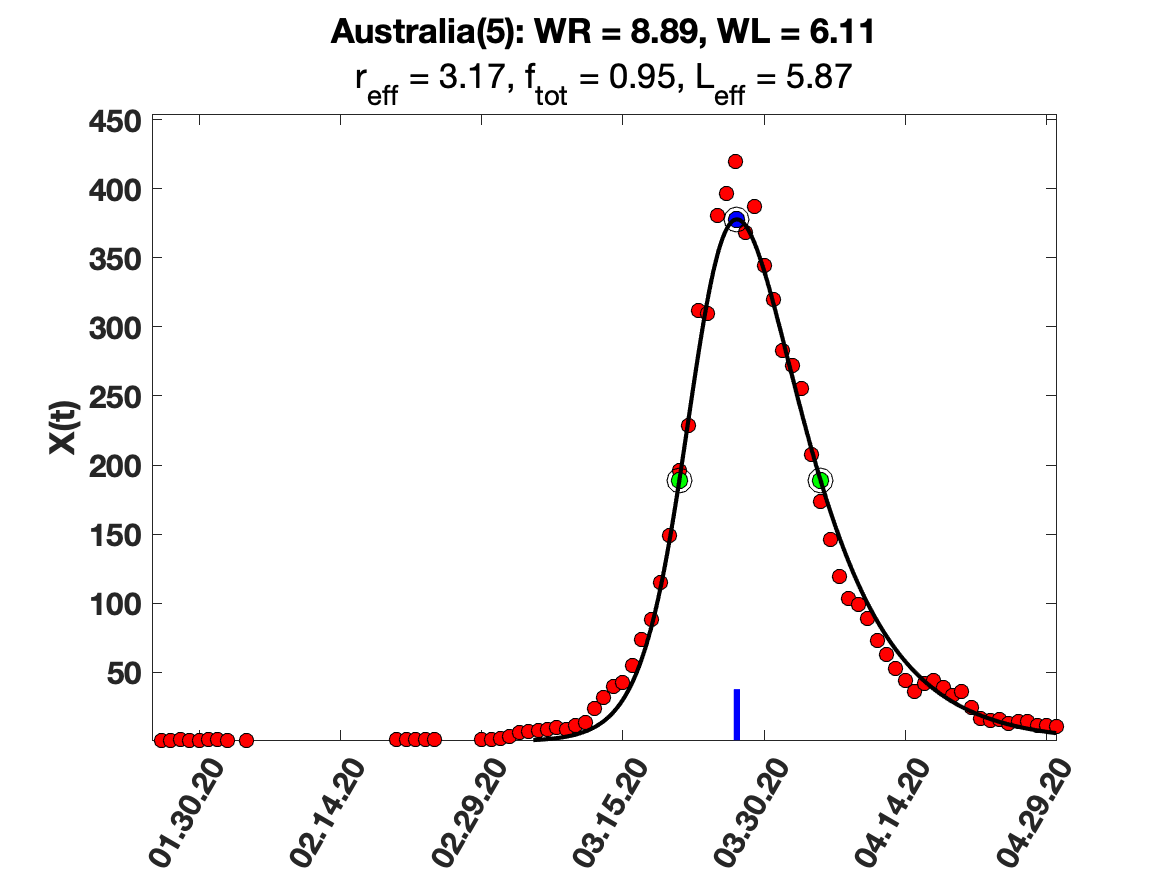

### Australia_7_day_av_and_fit.png

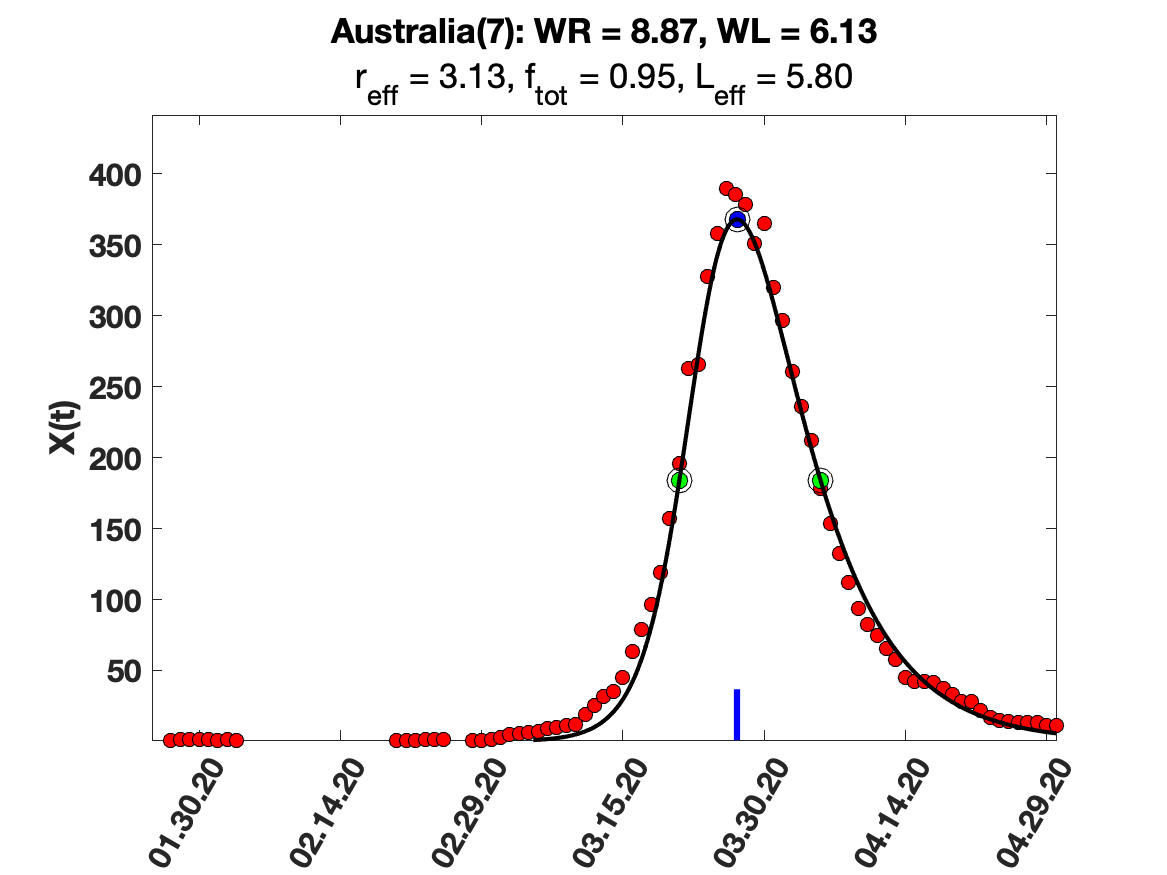

### Australia_9_day_av_and_fit.png

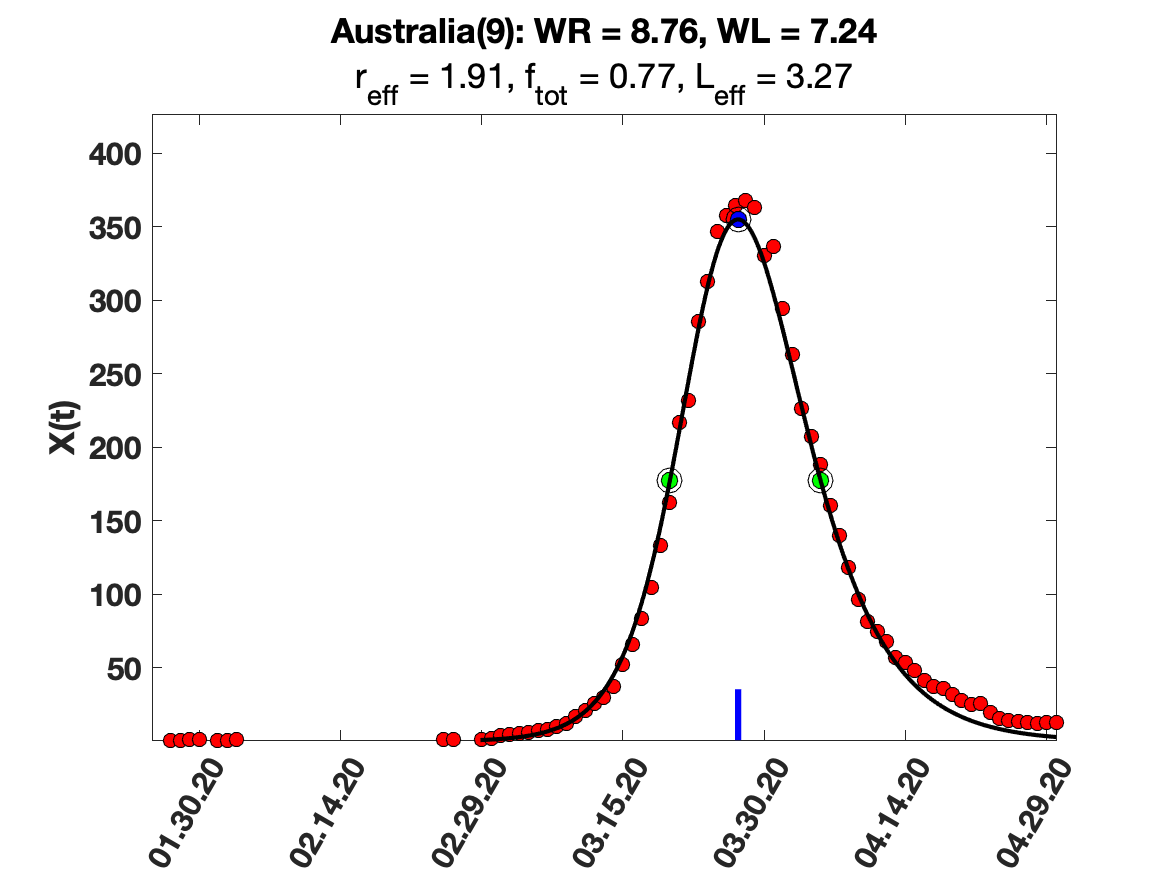

### Australia_i(tau)_for_average_r.png

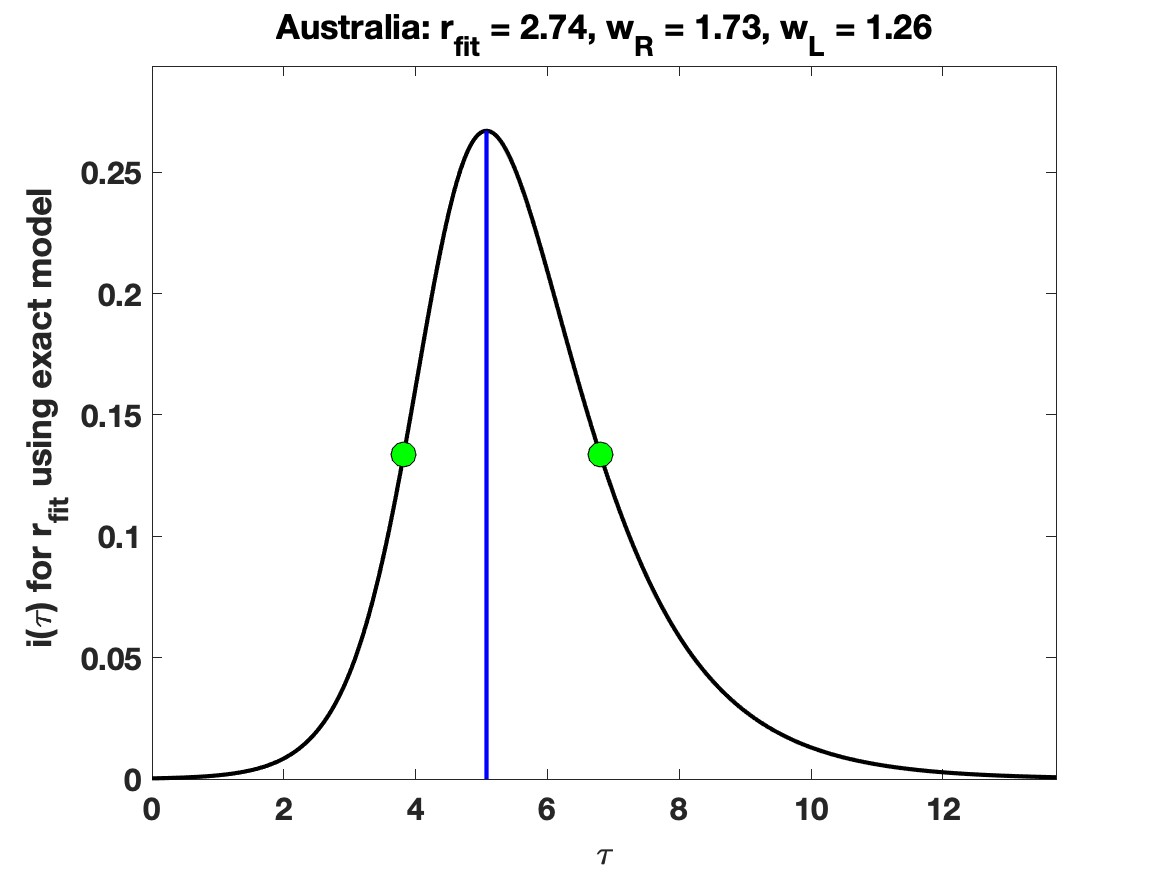

### Australia_solution_fit.png

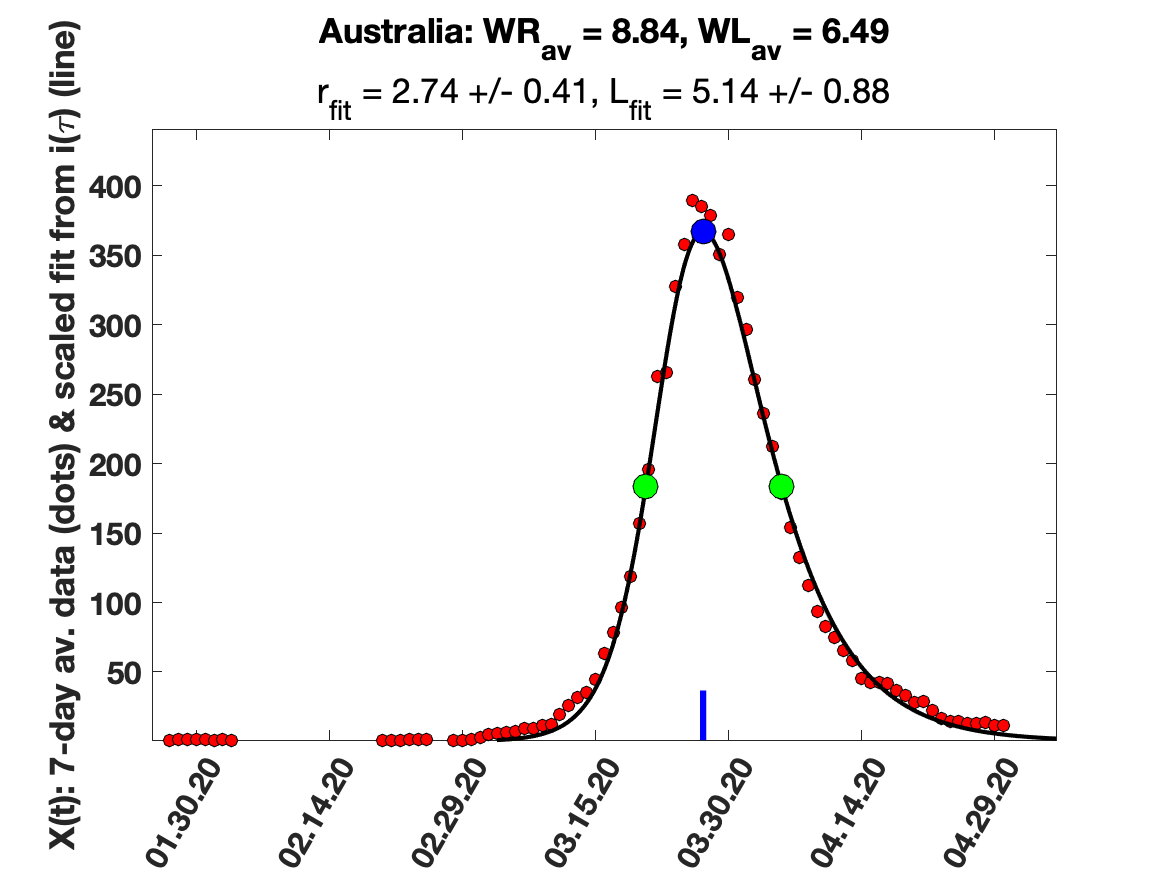

### Austria_5_day_av_and_fit.png

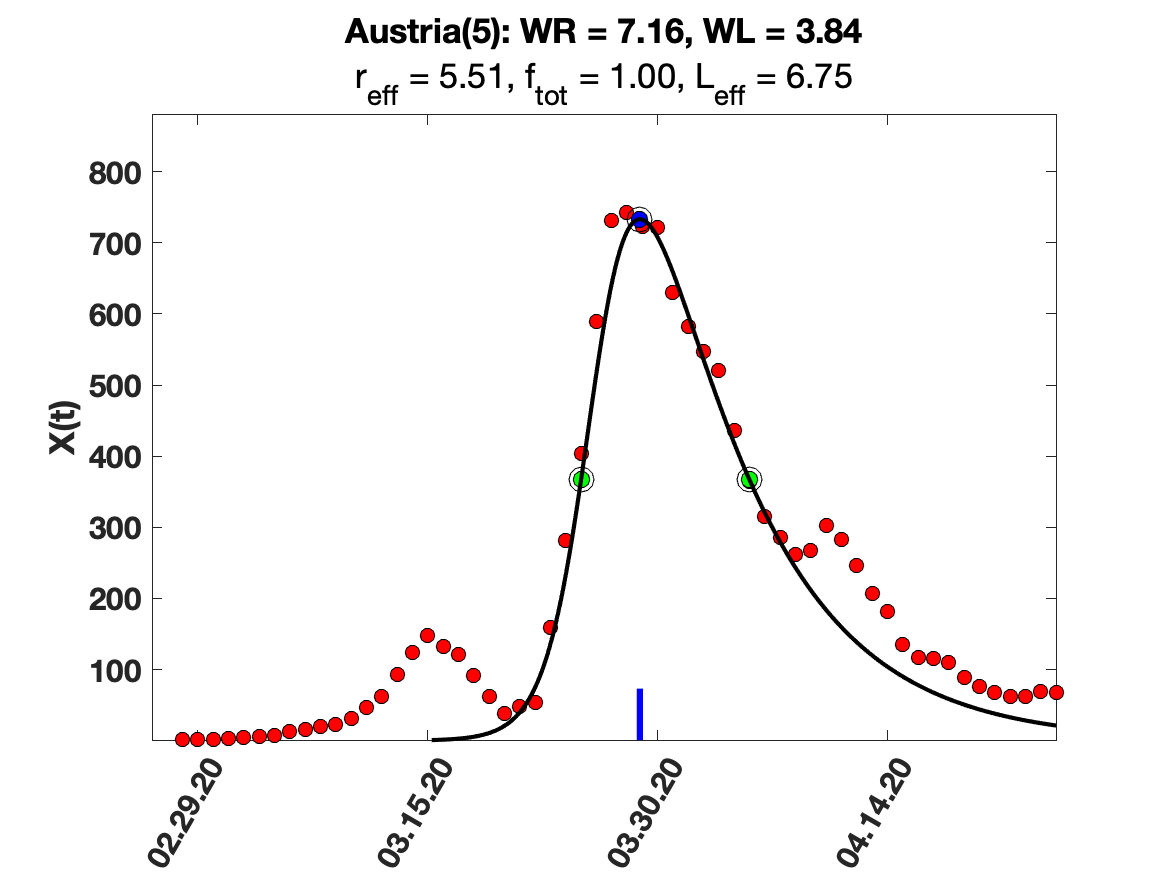

### Austria_7_day_av_and_fit.png

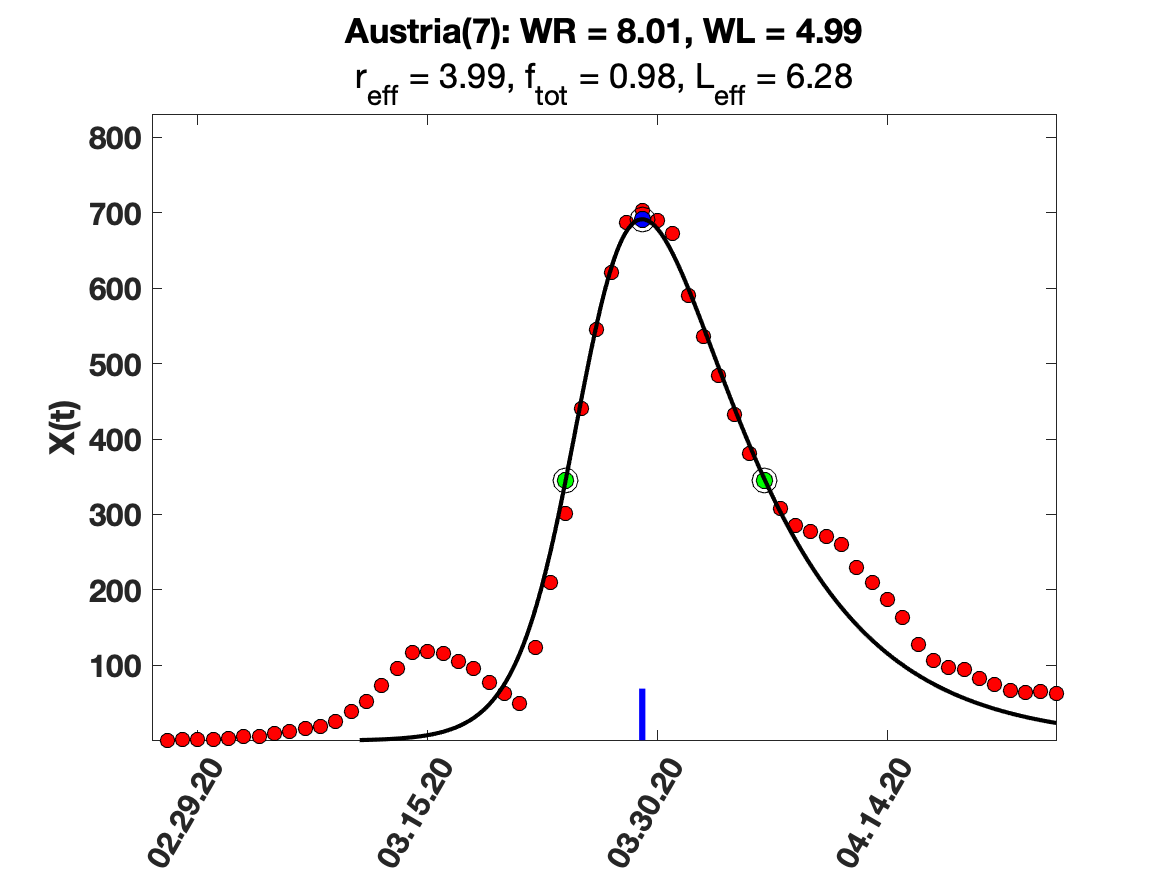

### Austria_9_day_av_and_fit.png

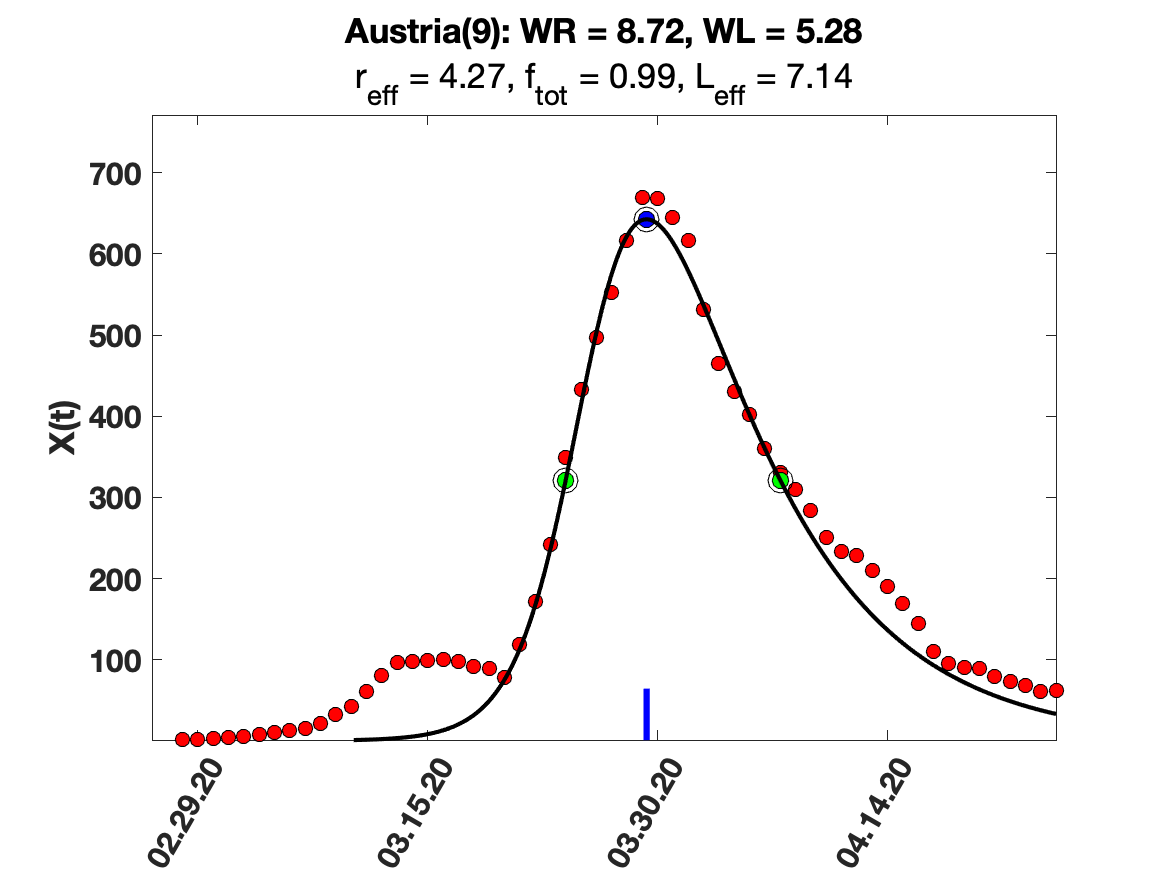

### Austria_i(tau)_for_average_r.png

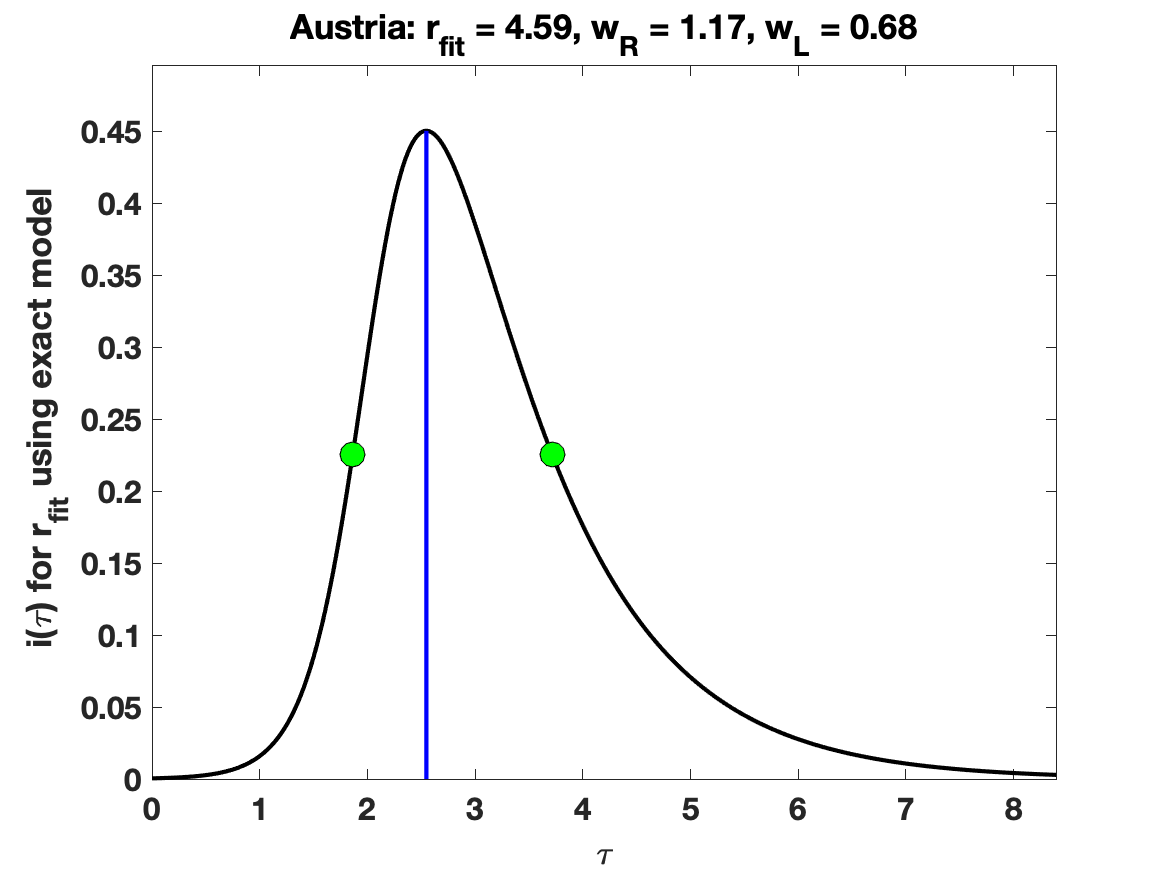

### Austria_raw_data.png

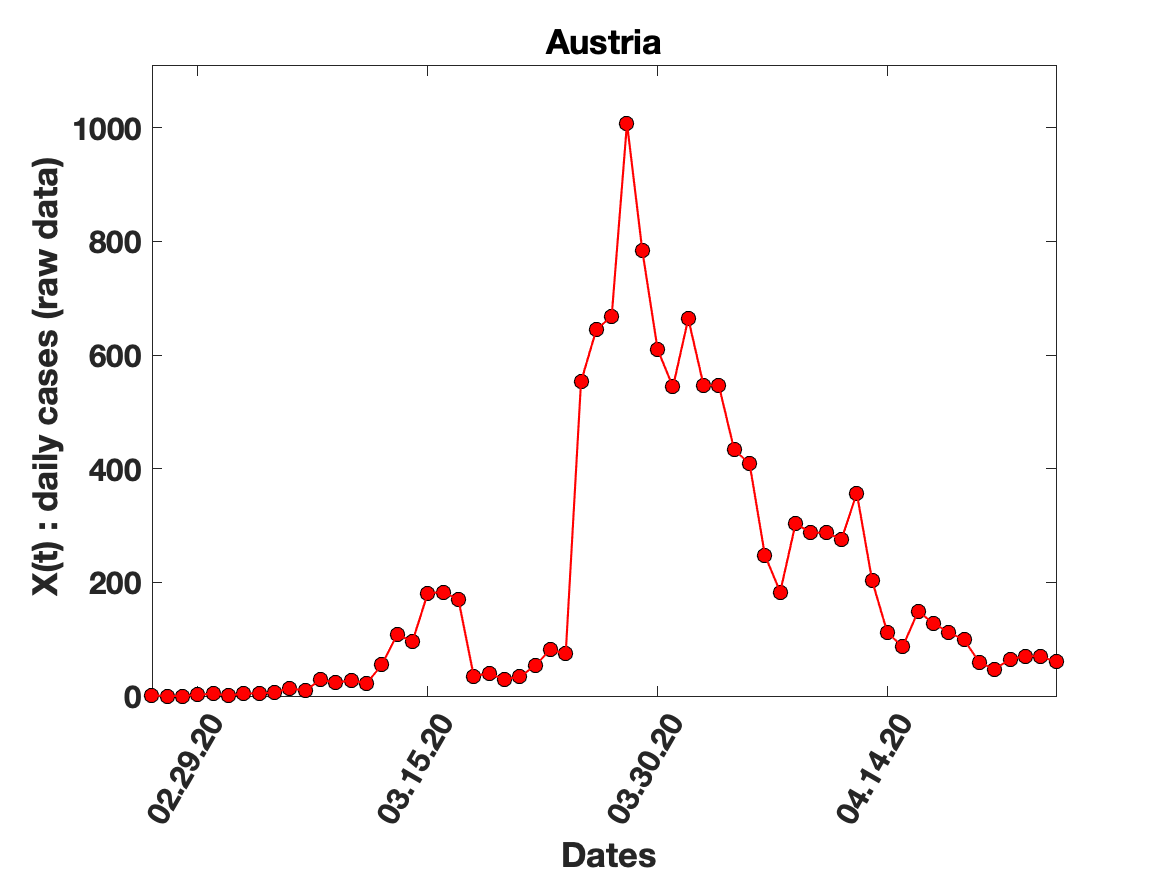

### Belgium_7_day_av_and_fit.png

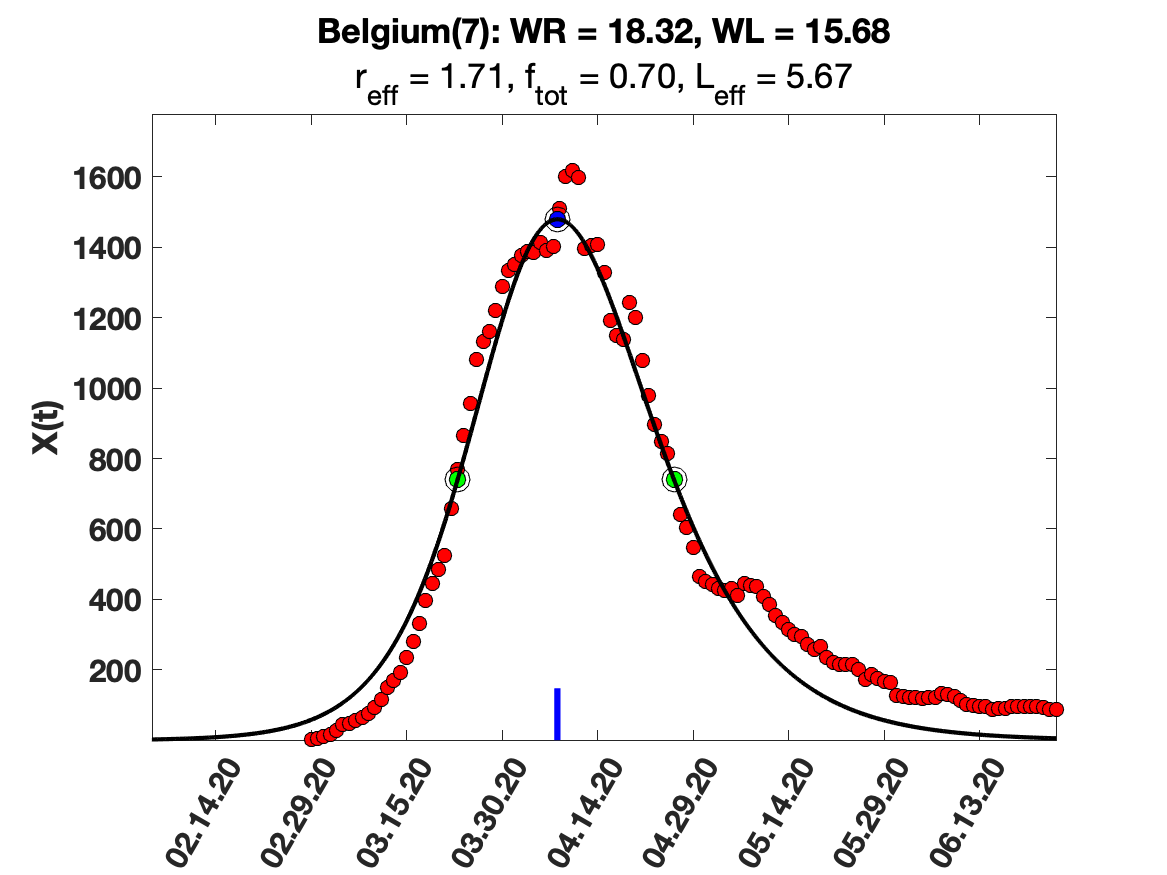

### Belgium_9_day_av_and_fit.png

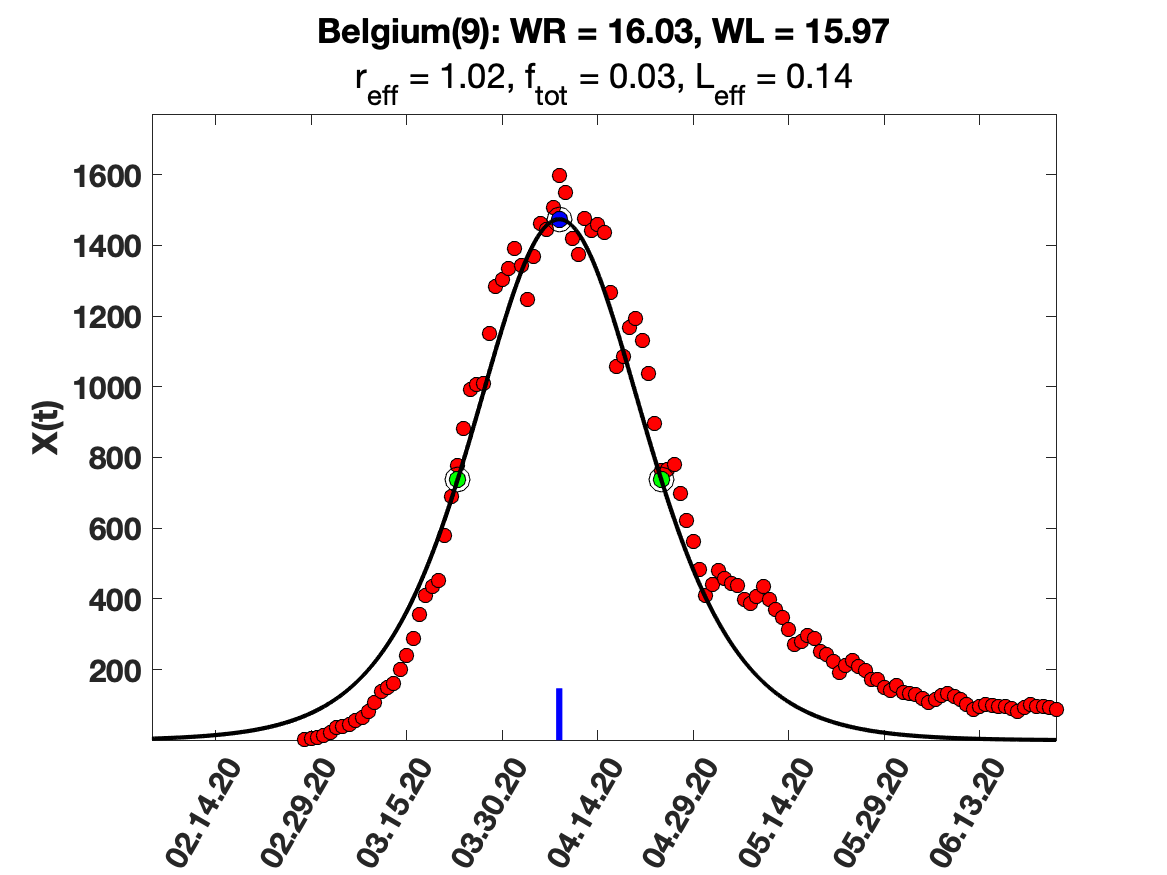

### Belgium_raw_data.png

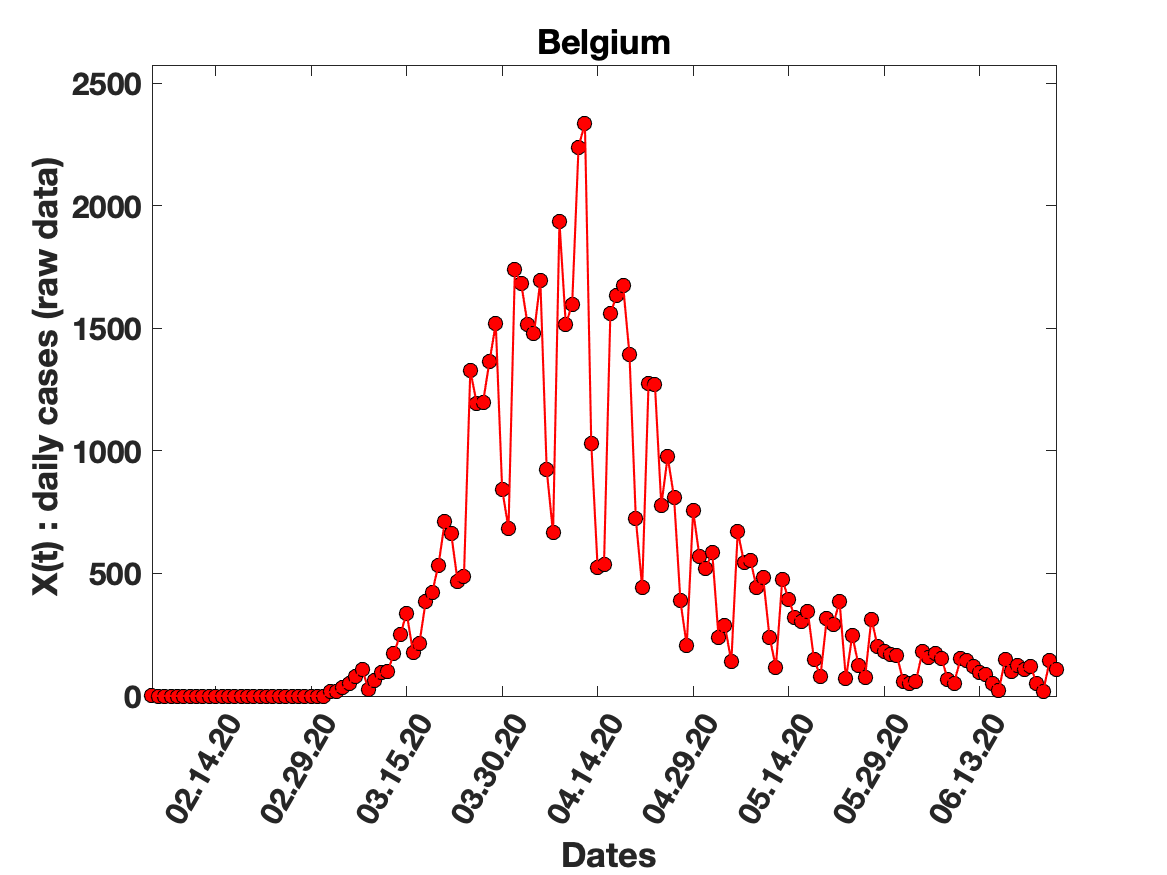

### Belgium_solution_fit.png

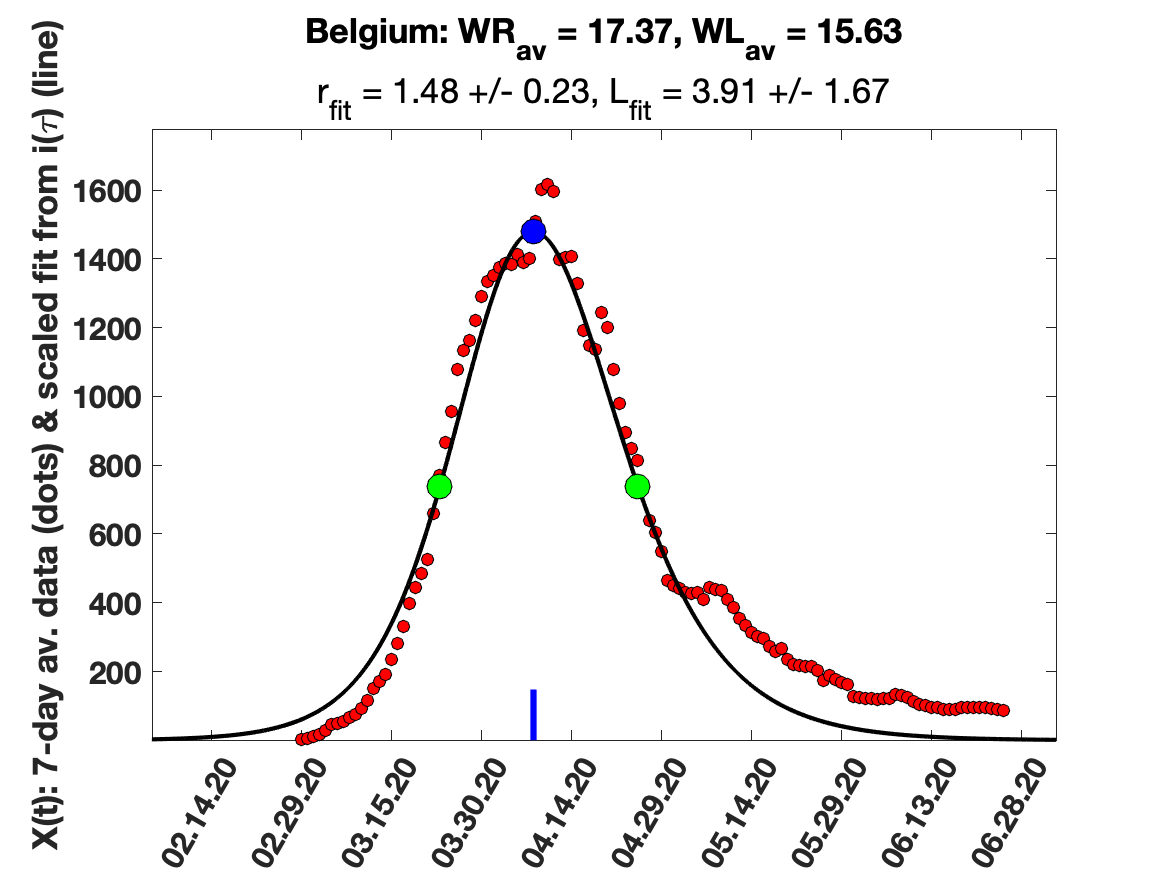

### Egypt_5_day_av_and_fit.png

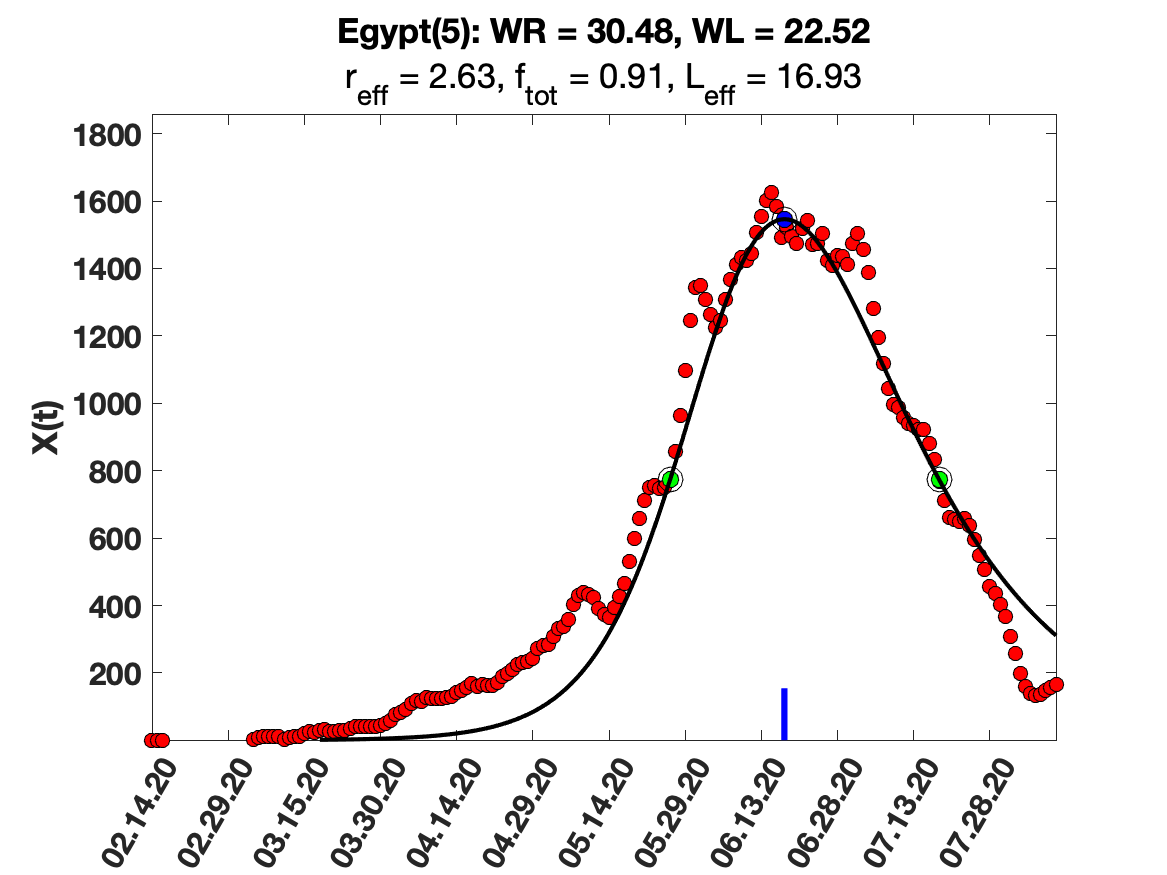

### Egypt_7_day_av_and_fit.png

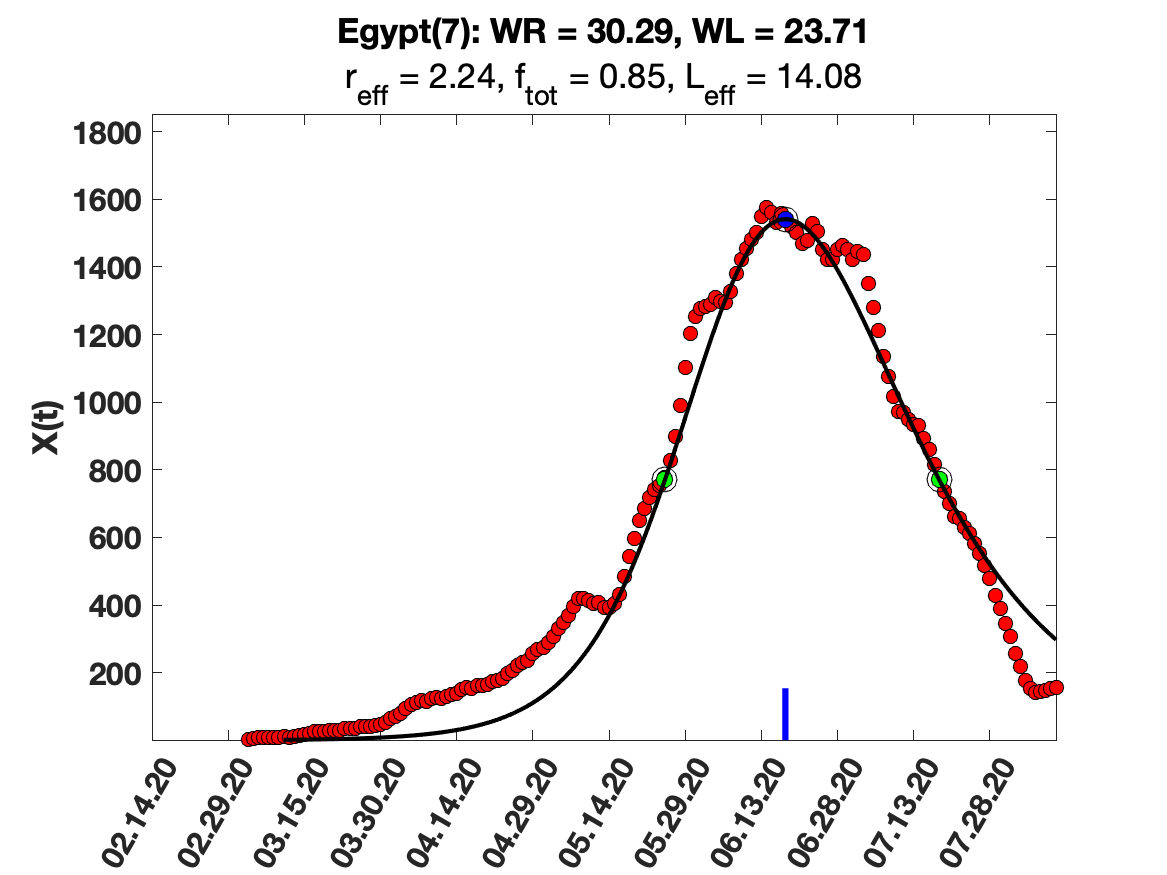

### Egypt_9_day_av_and_fit.png

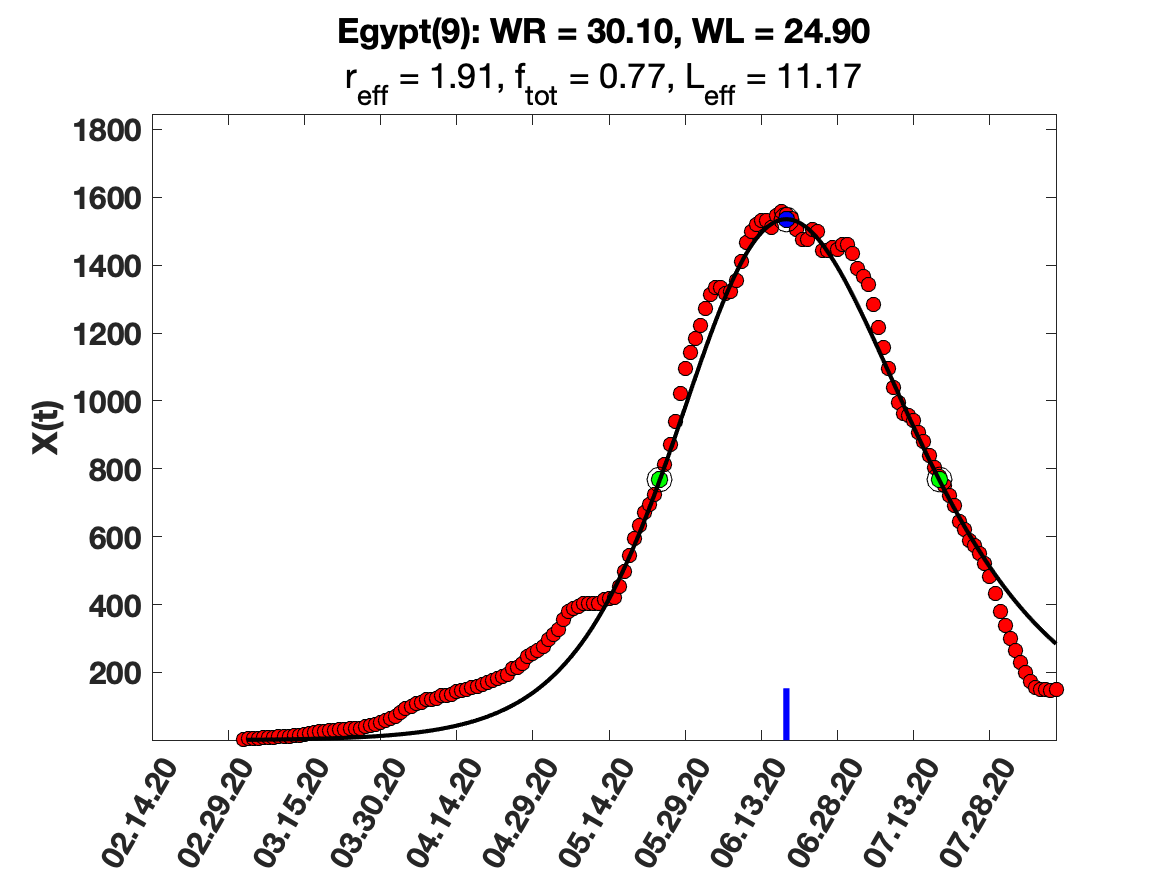

### Egypt_i(tau)_for_average_r.png

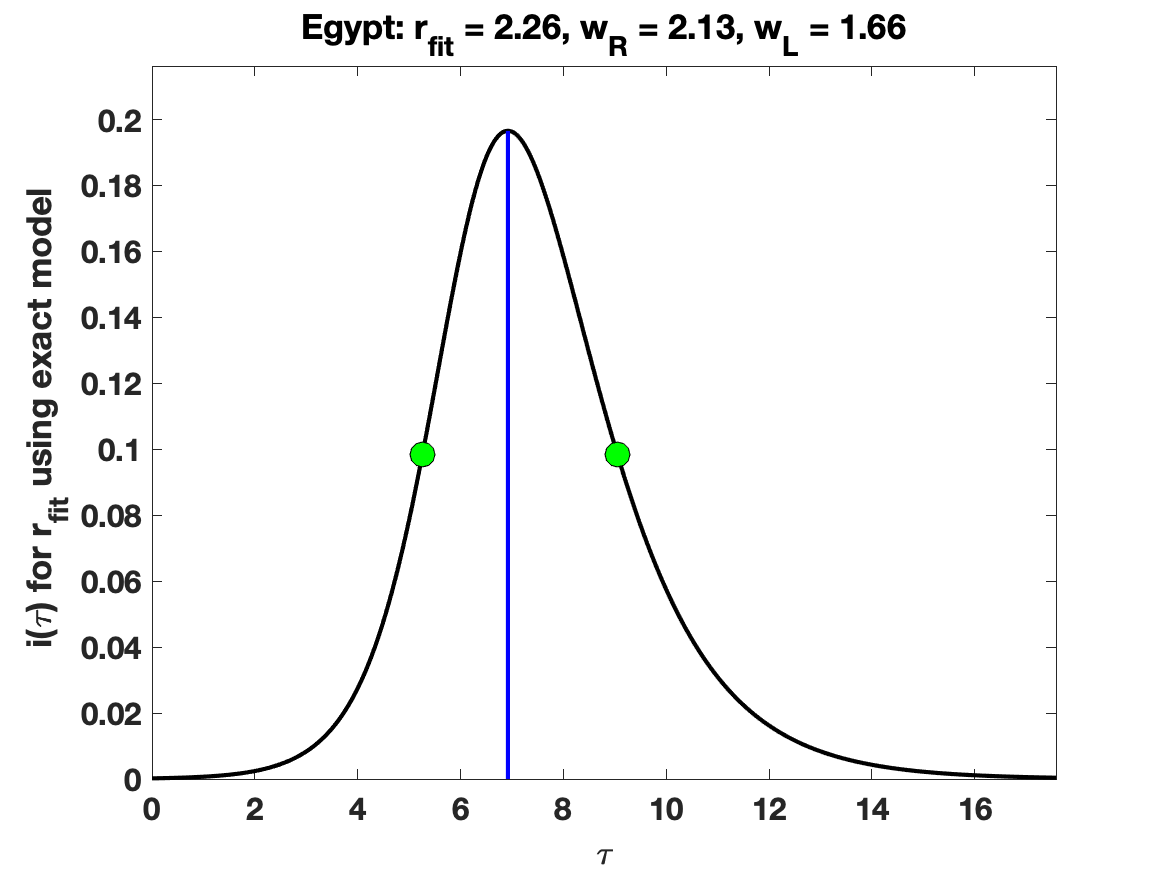

### Egypt_solution_fit.png

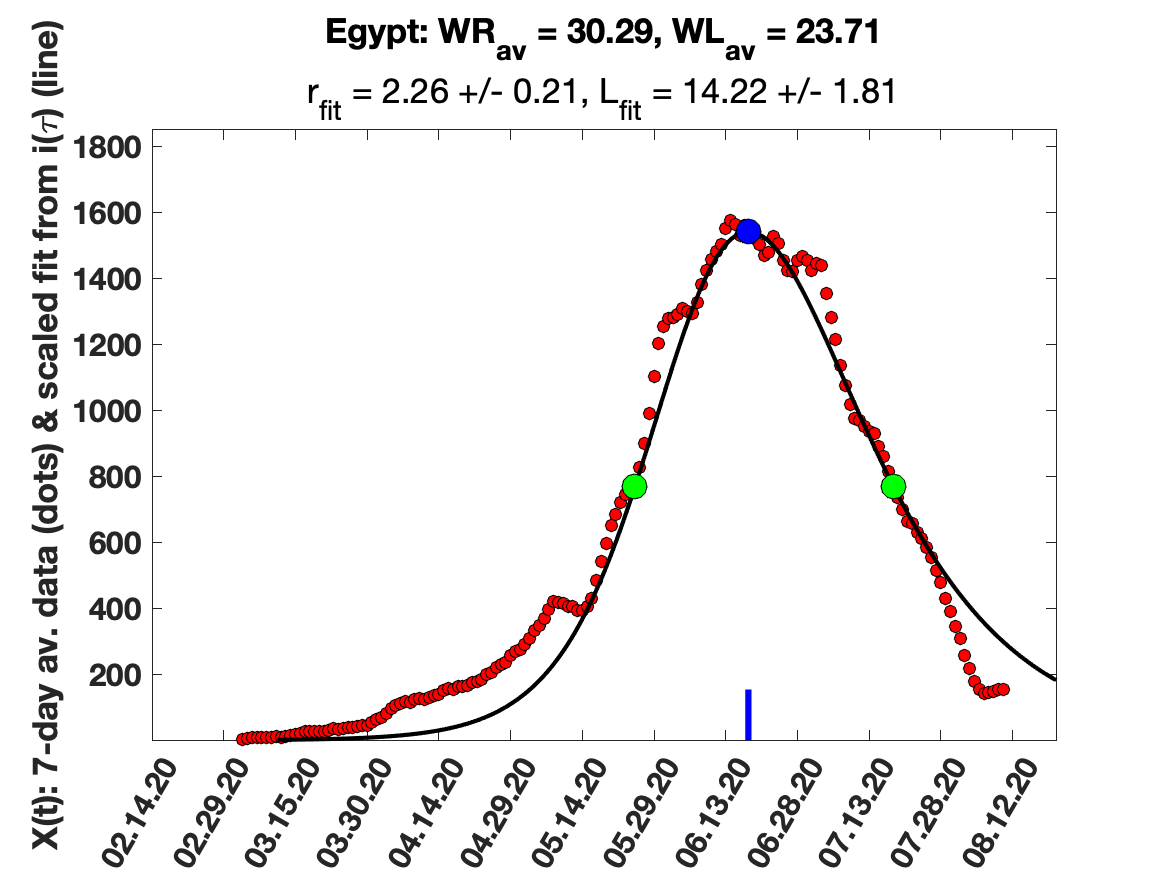

### France_5_day_av_and_fit.png

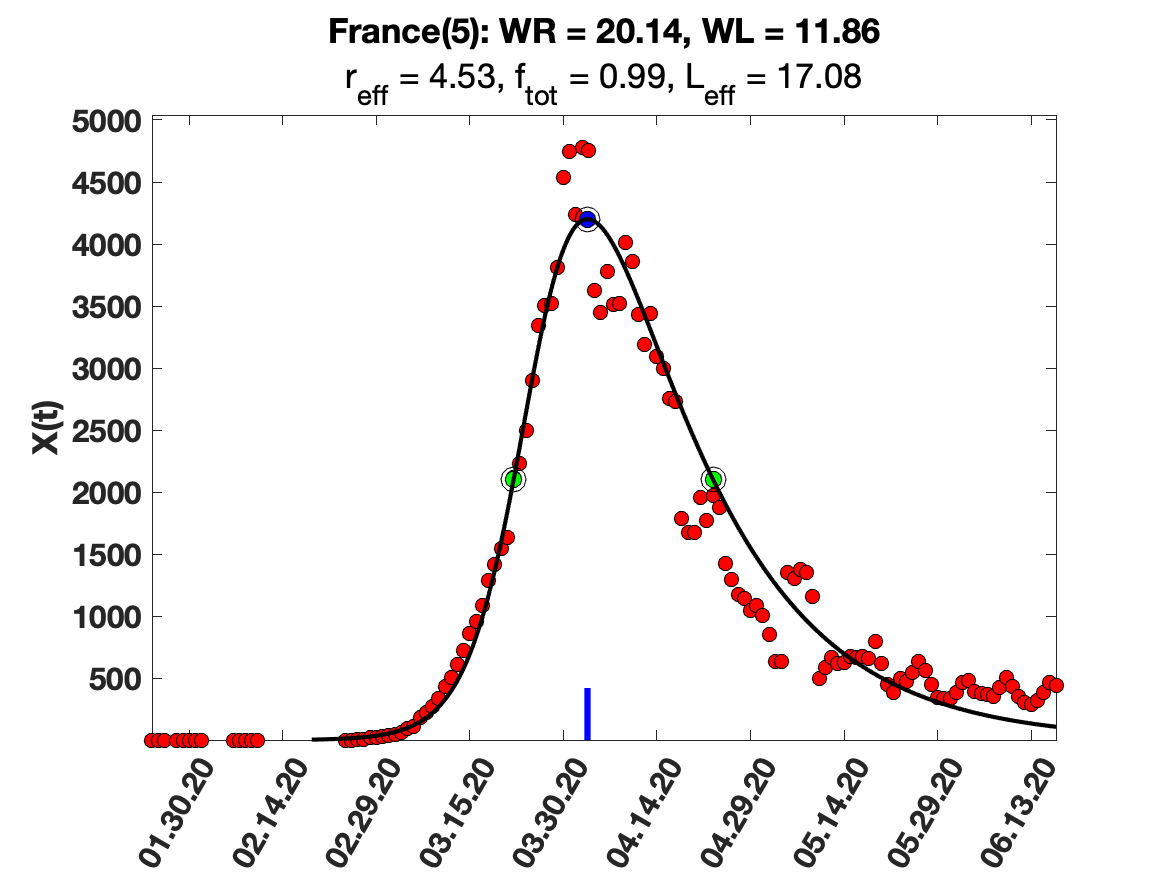

### France_i(tau)_for_average_r.png

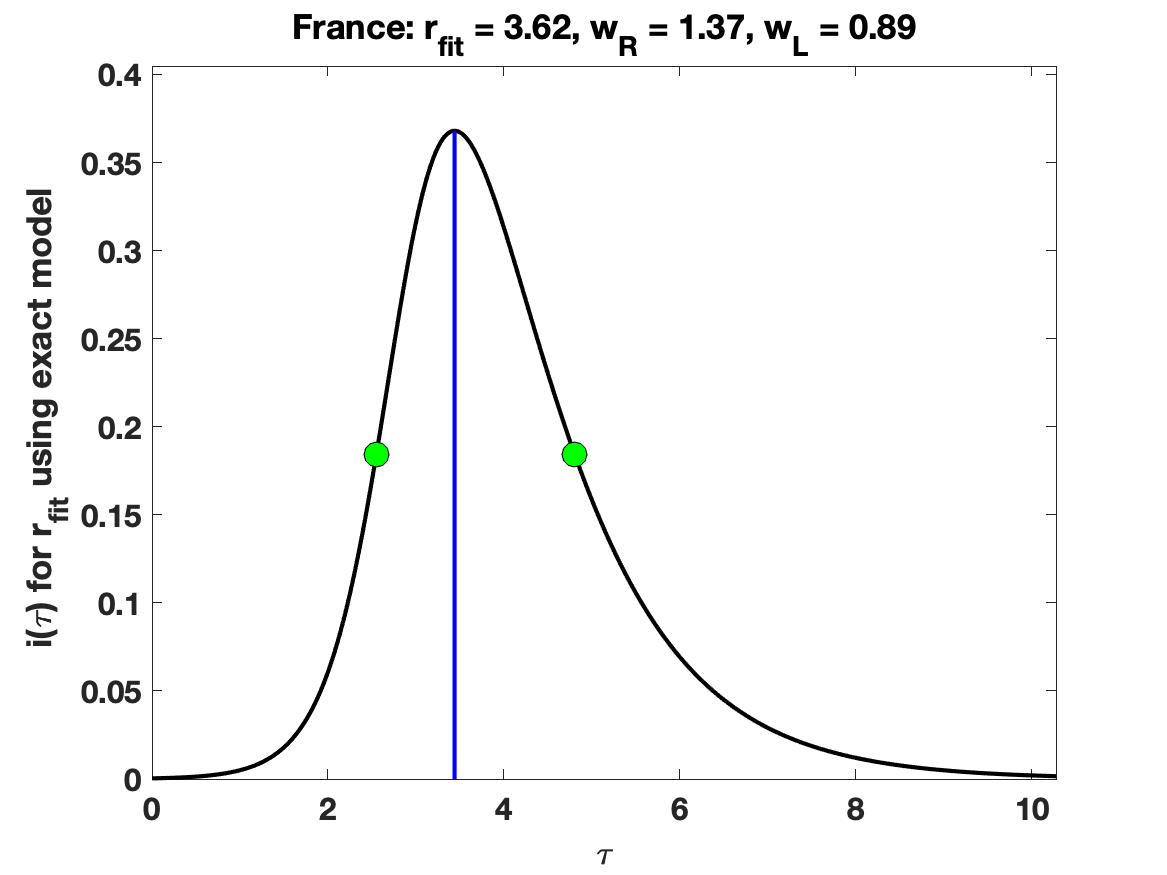

### France_raw_data.png

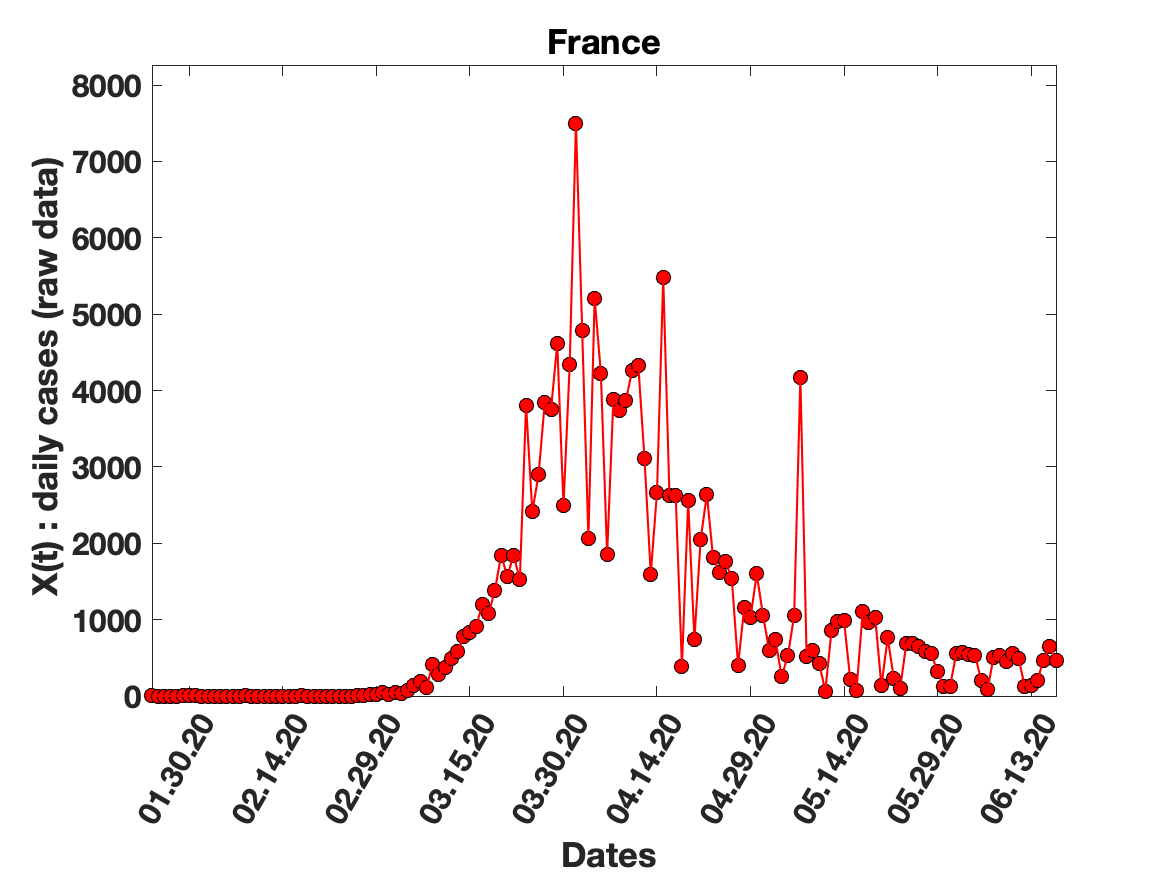

### France_solution_fit.png

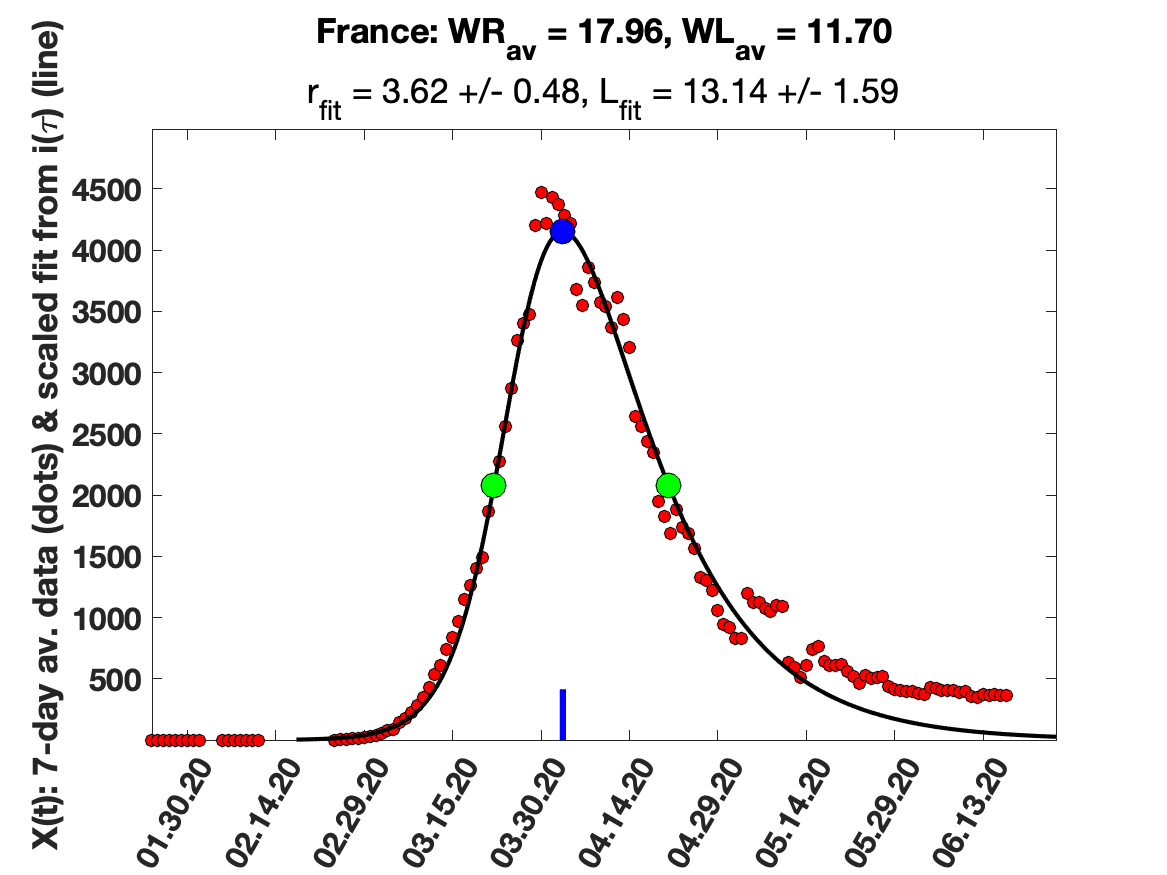

### Germany_5_day_av_and_fit.png

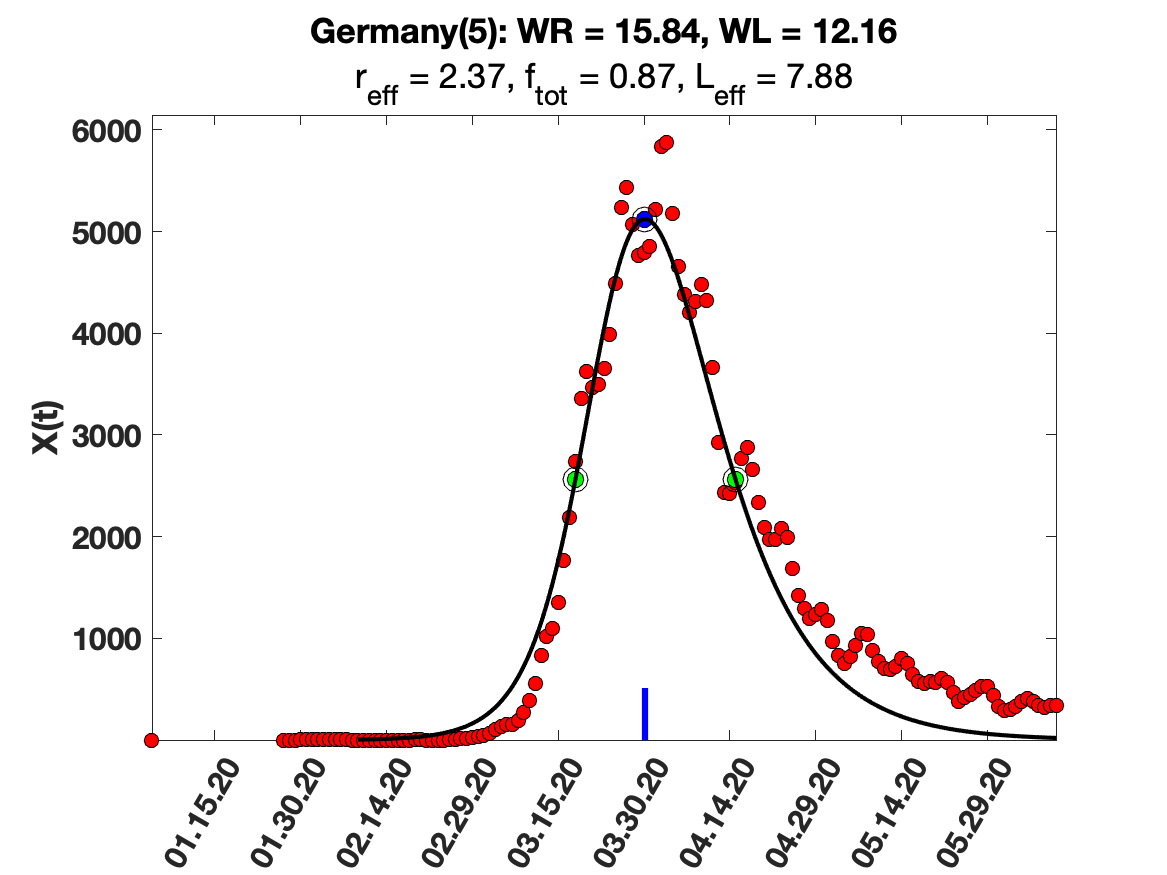

### Germany_9_day_av_and_fit.png

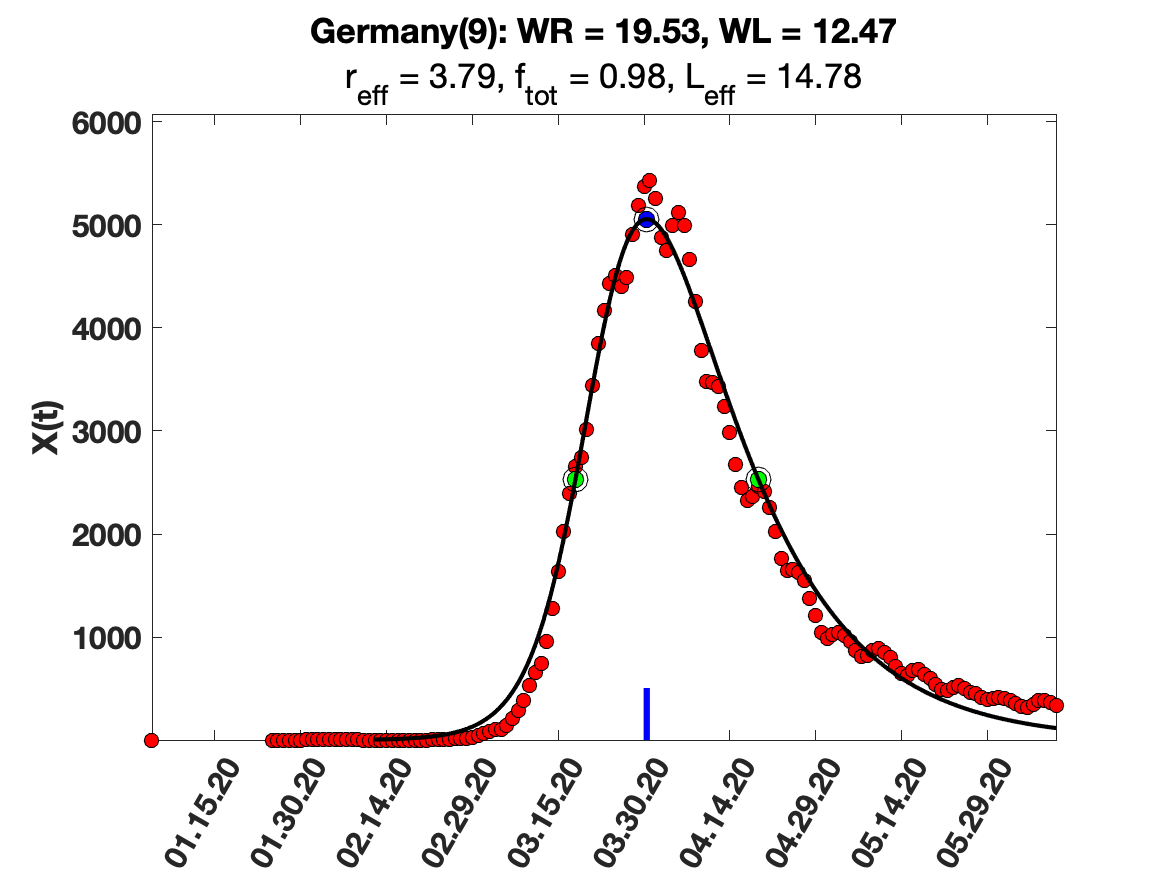

### Germany_i(tau)_for_average_r.png

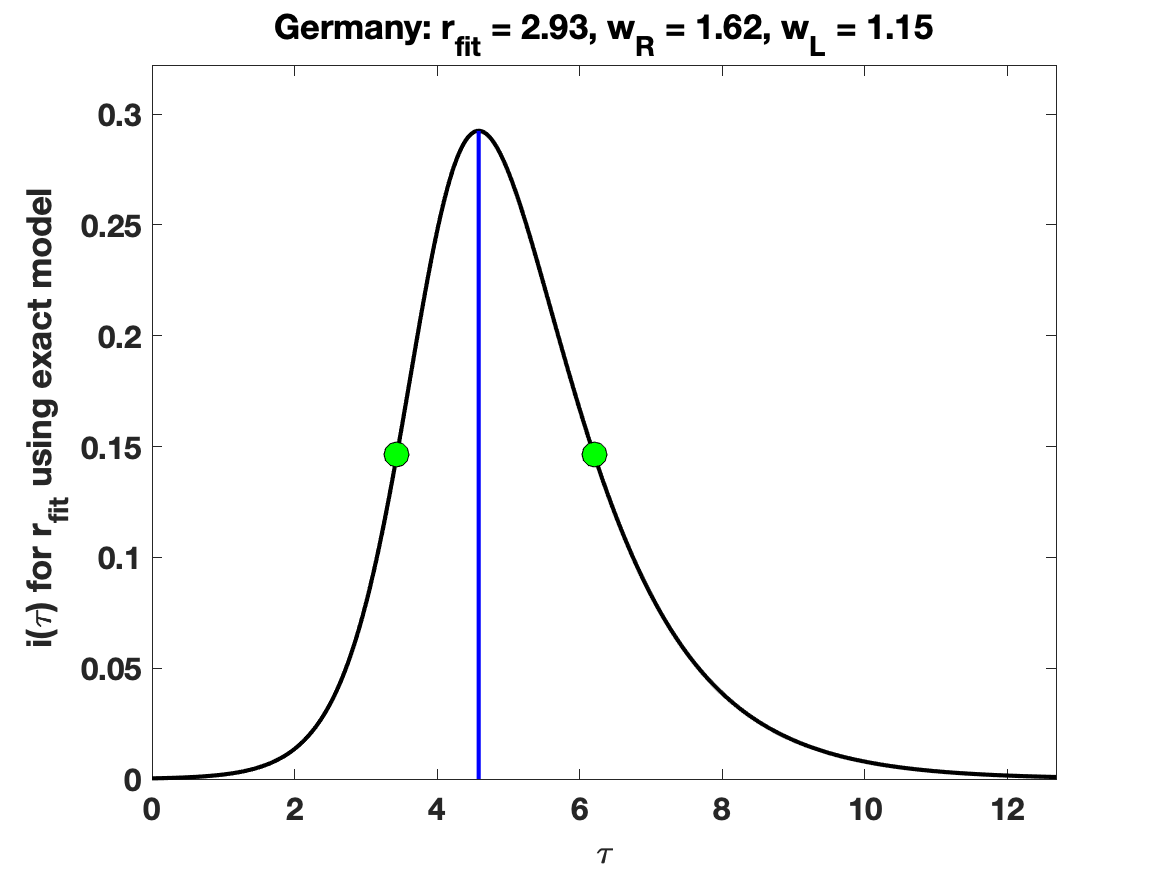

### Germany_raw_data.png

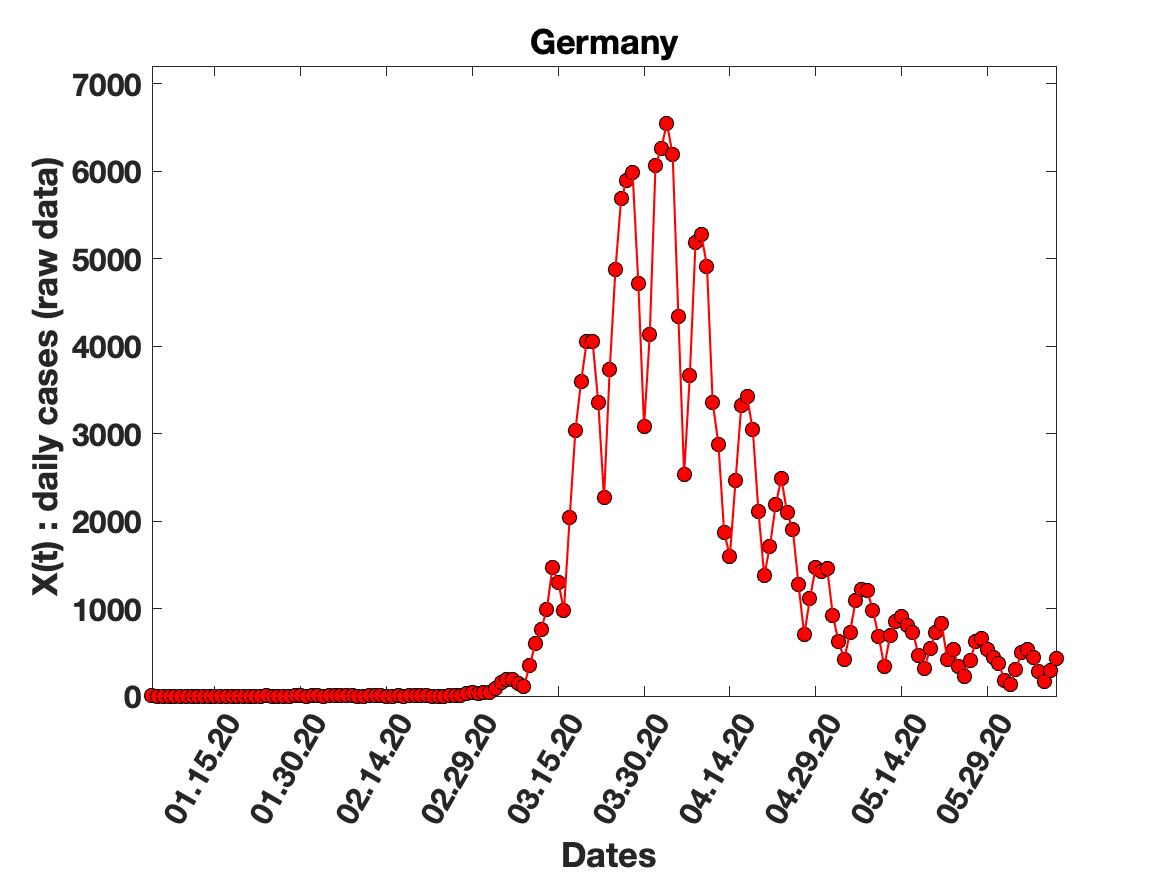

### Germany_solution_fit.png

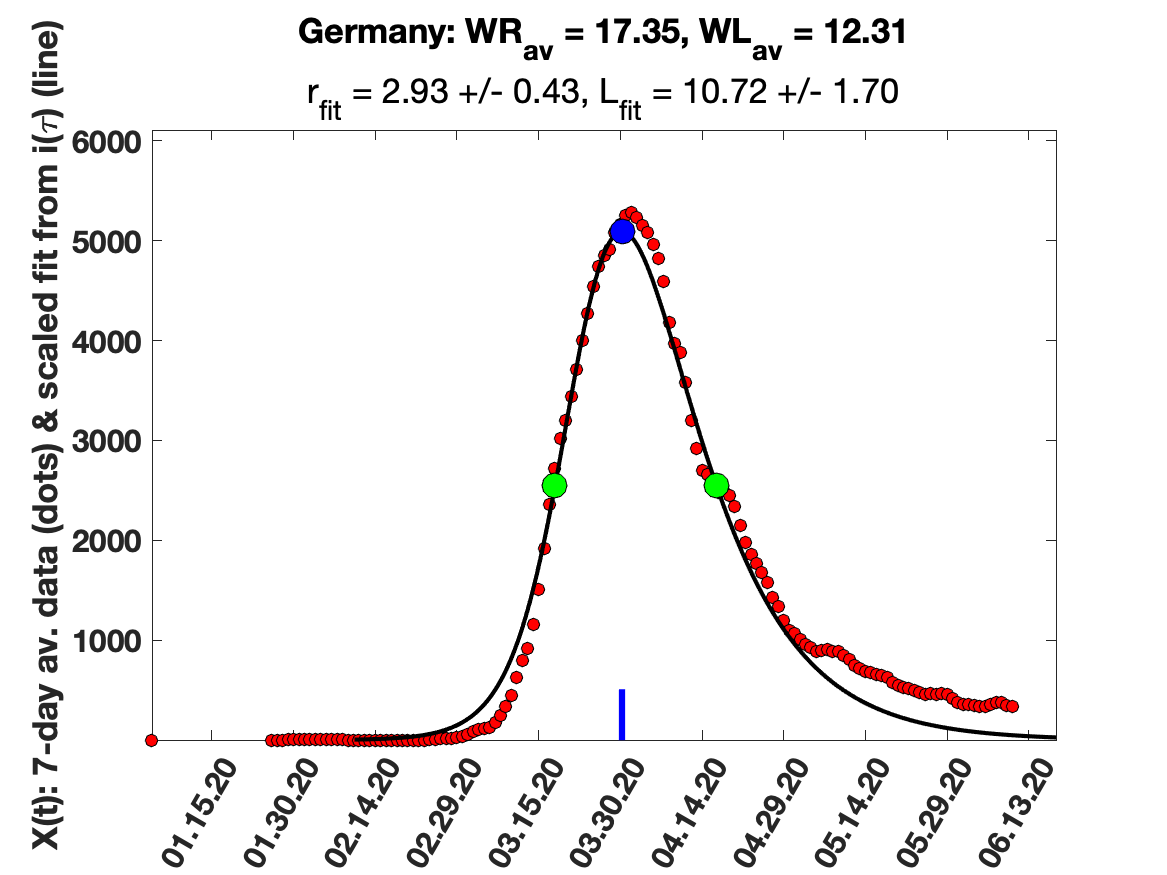

### Ireland_5_day_av_and_fit.png

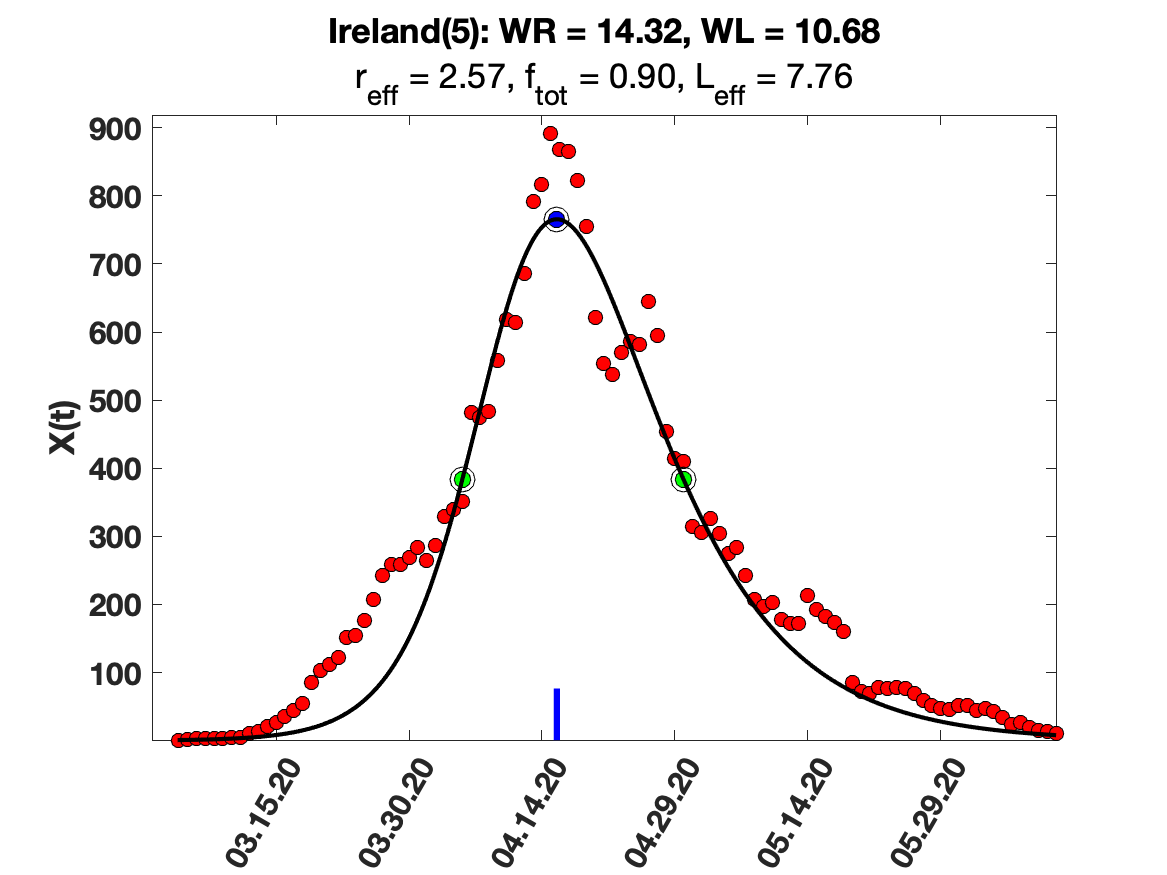

### Ireland_7_day_av_and_fit.png

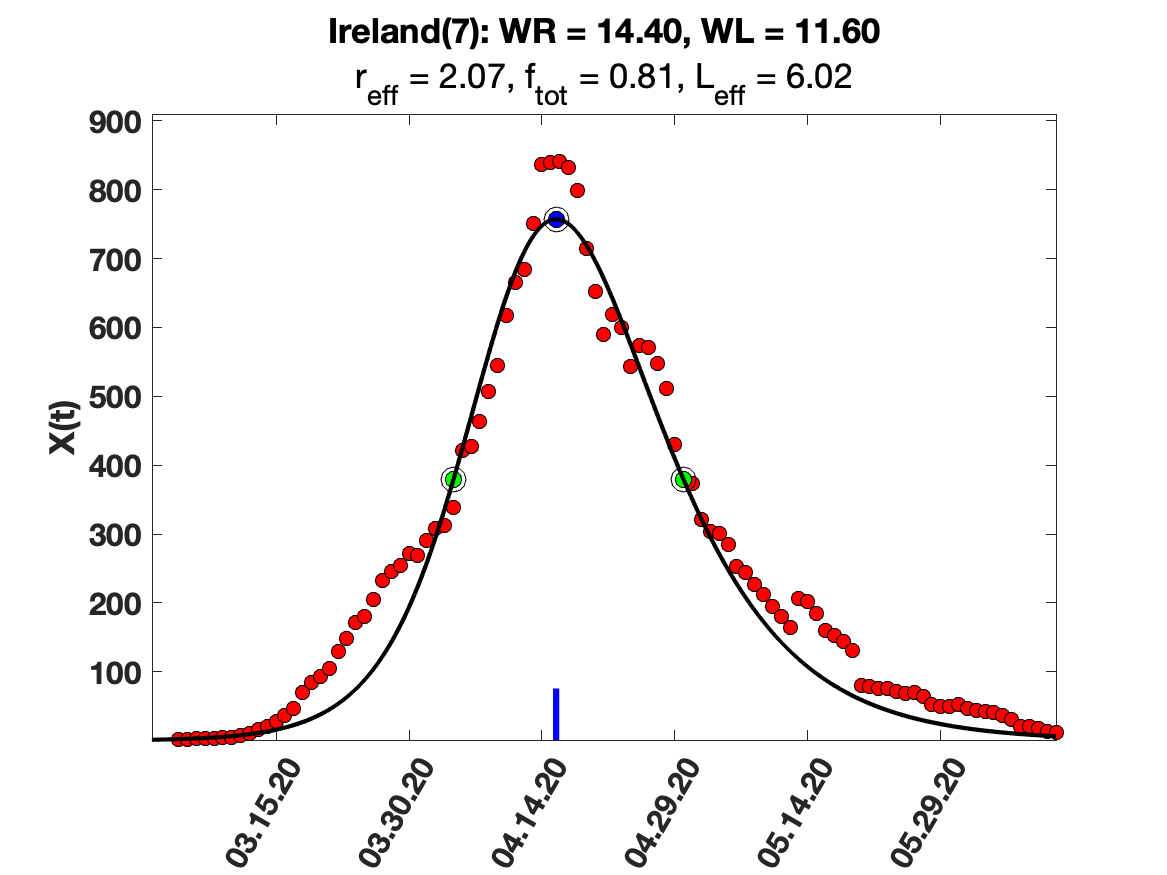

### Ireland_9_day_av_and_fit.png

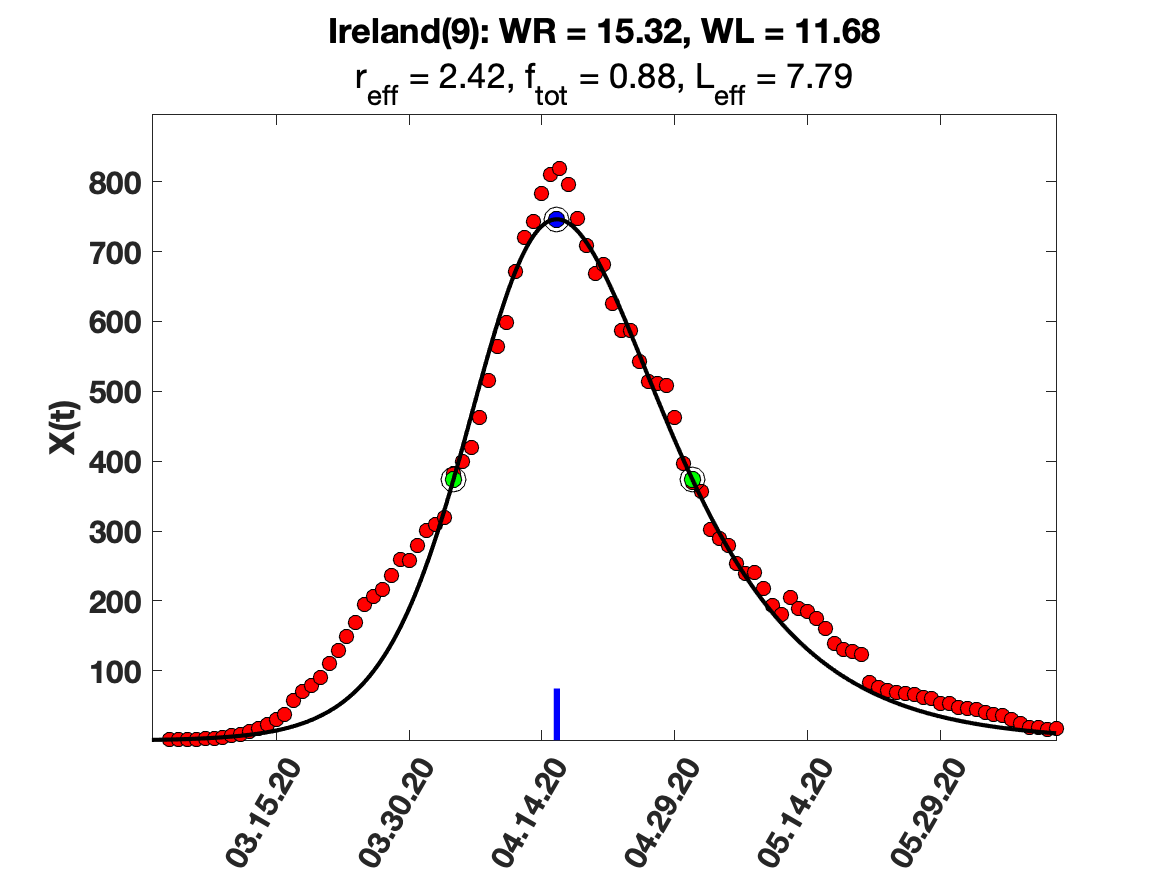

### Ireland_i(tau)_for_average_r.png

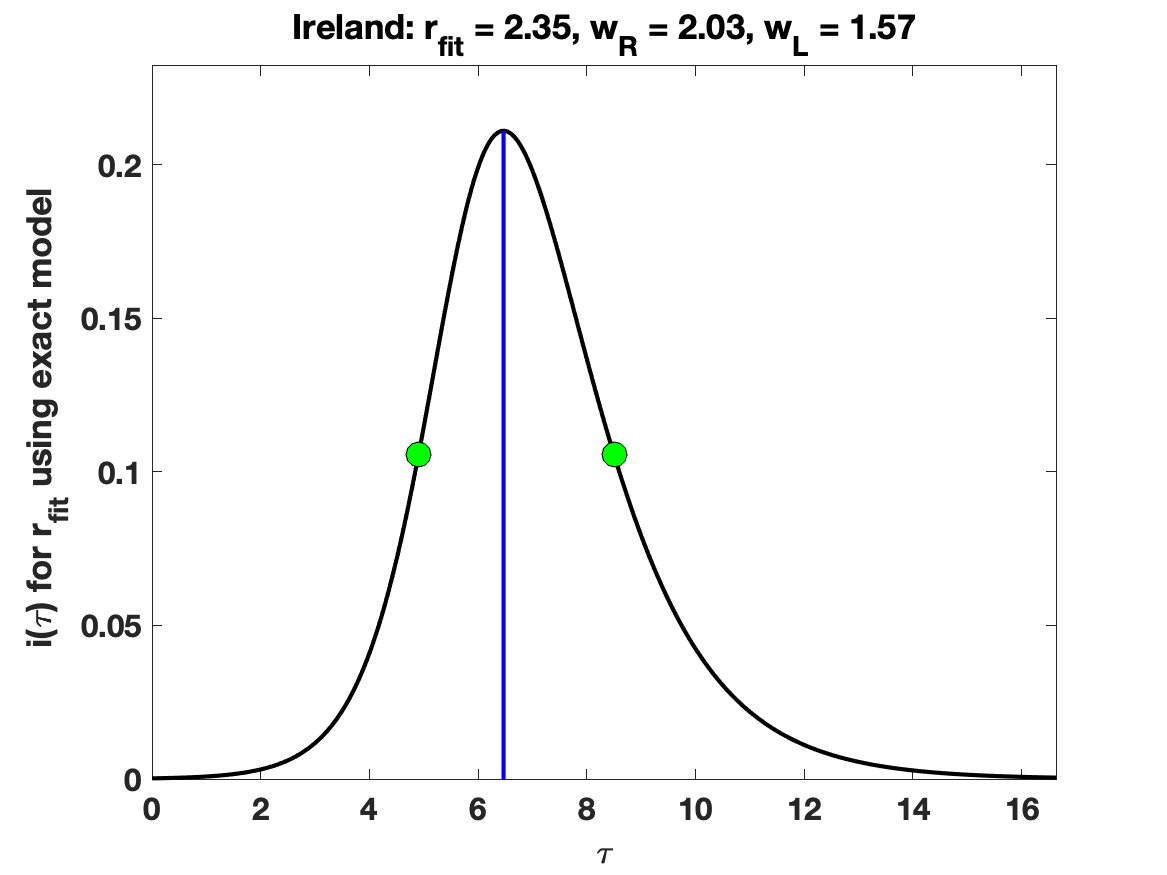

### Ireland_raw_data.png

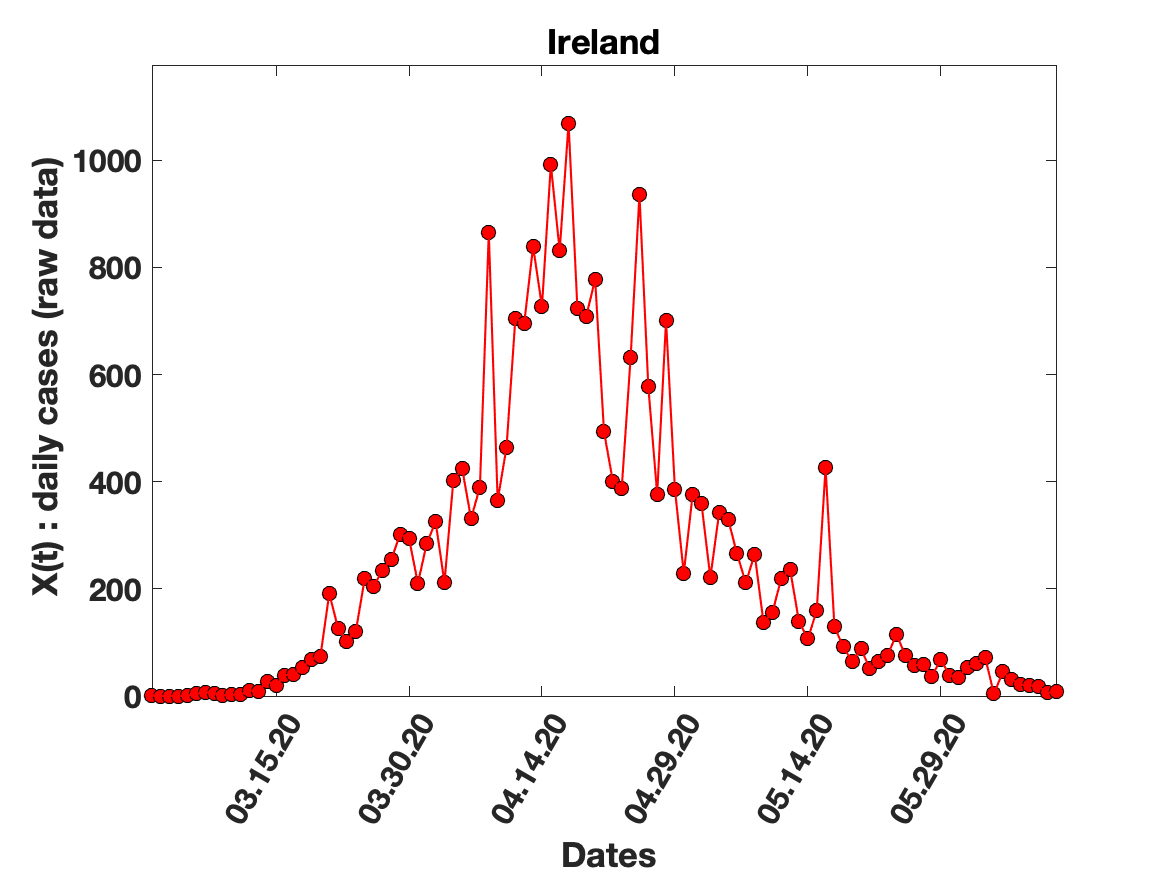

### Ireland_solution_fit.png

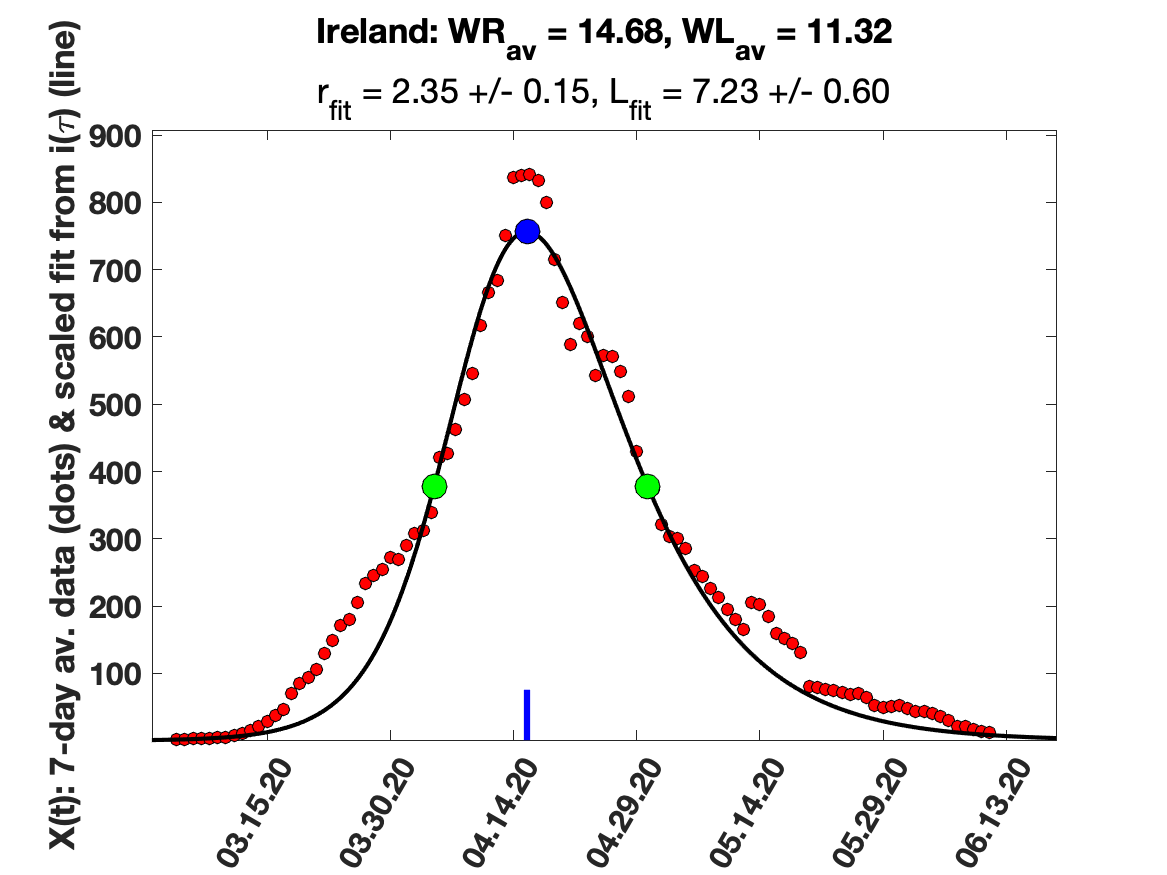

### Israel_5_day_av_and_fit.png

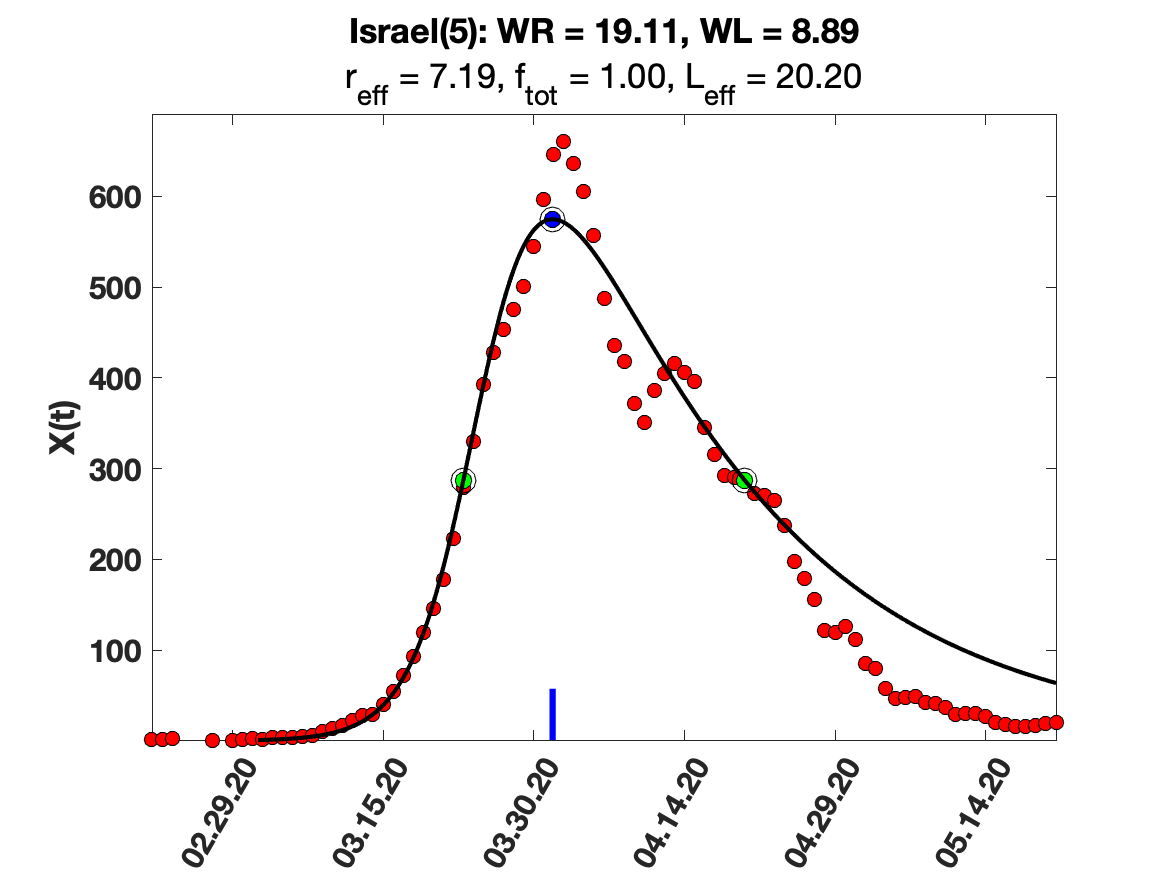

### Israel_7_day_av_and_fit.png

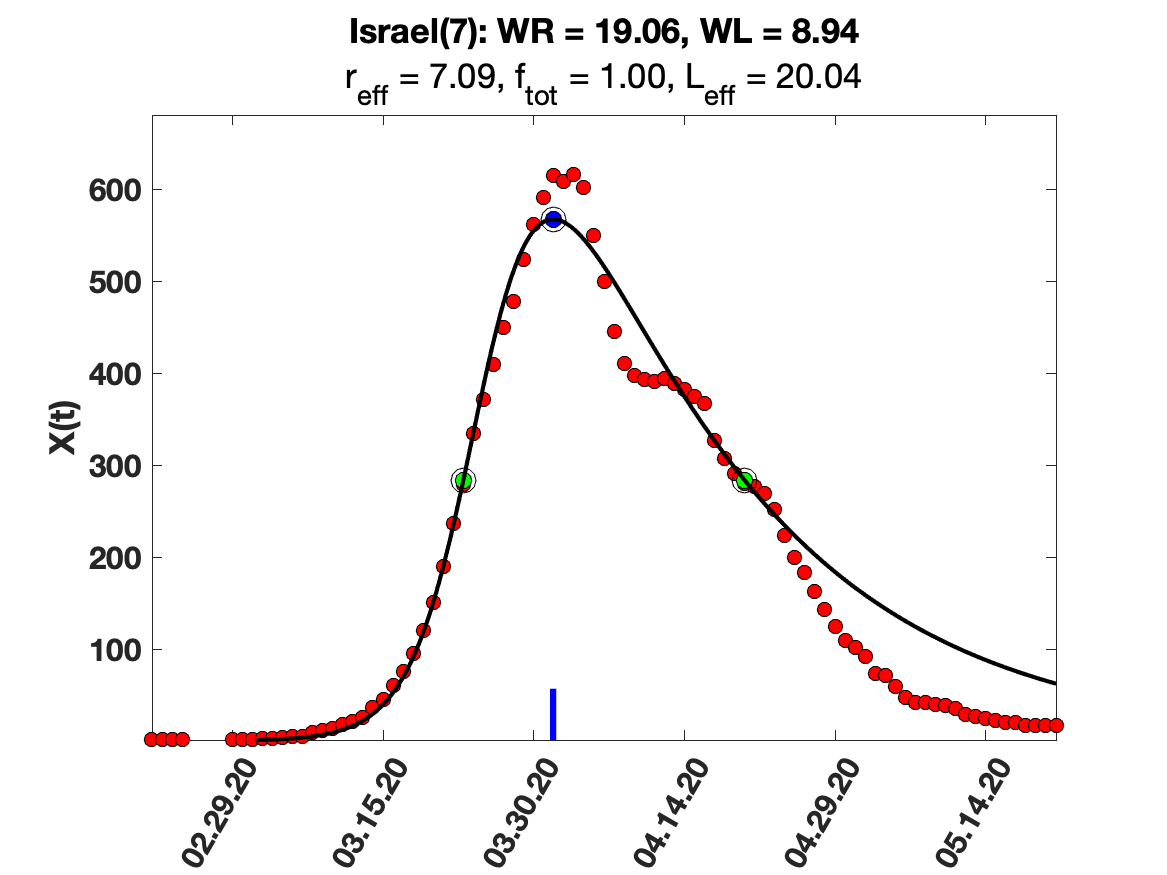

### Israel_9_day_av_and_fit.png

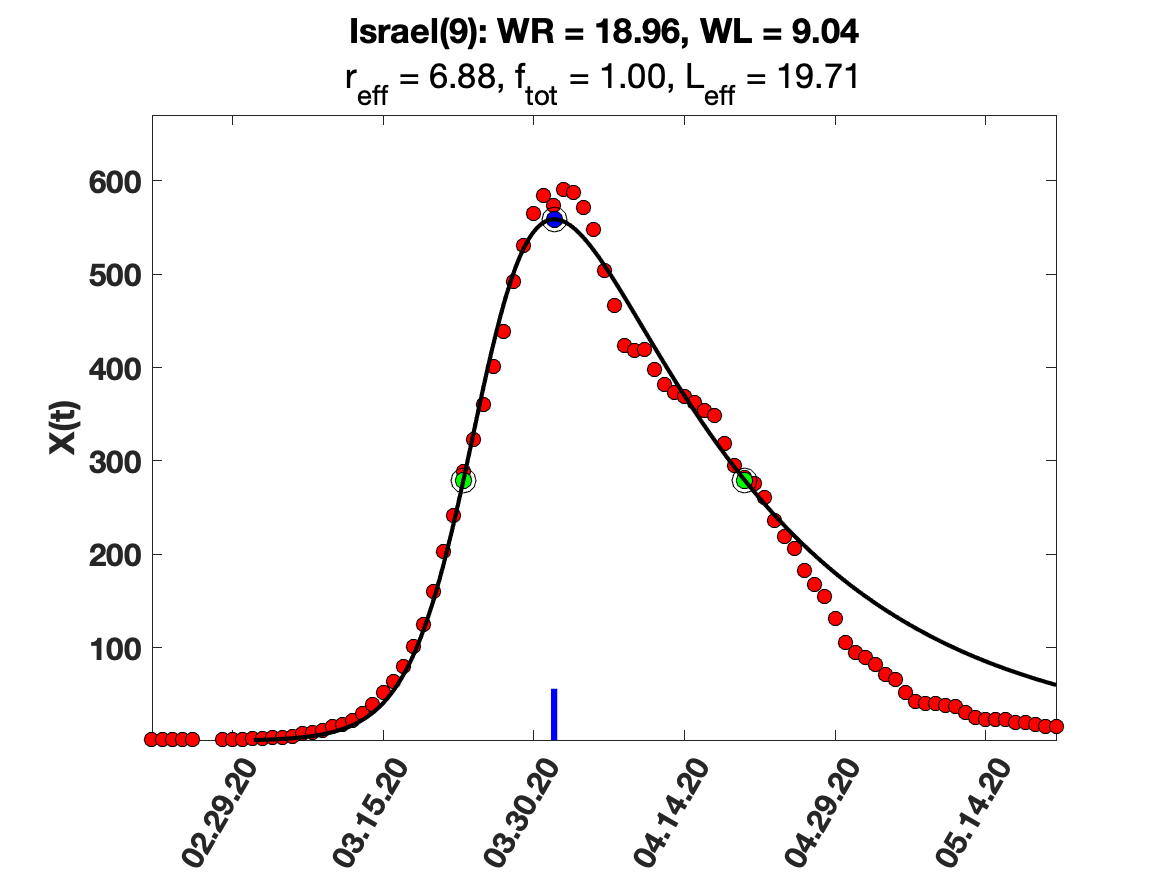

### Israel_i(tau)_for_average_r.png

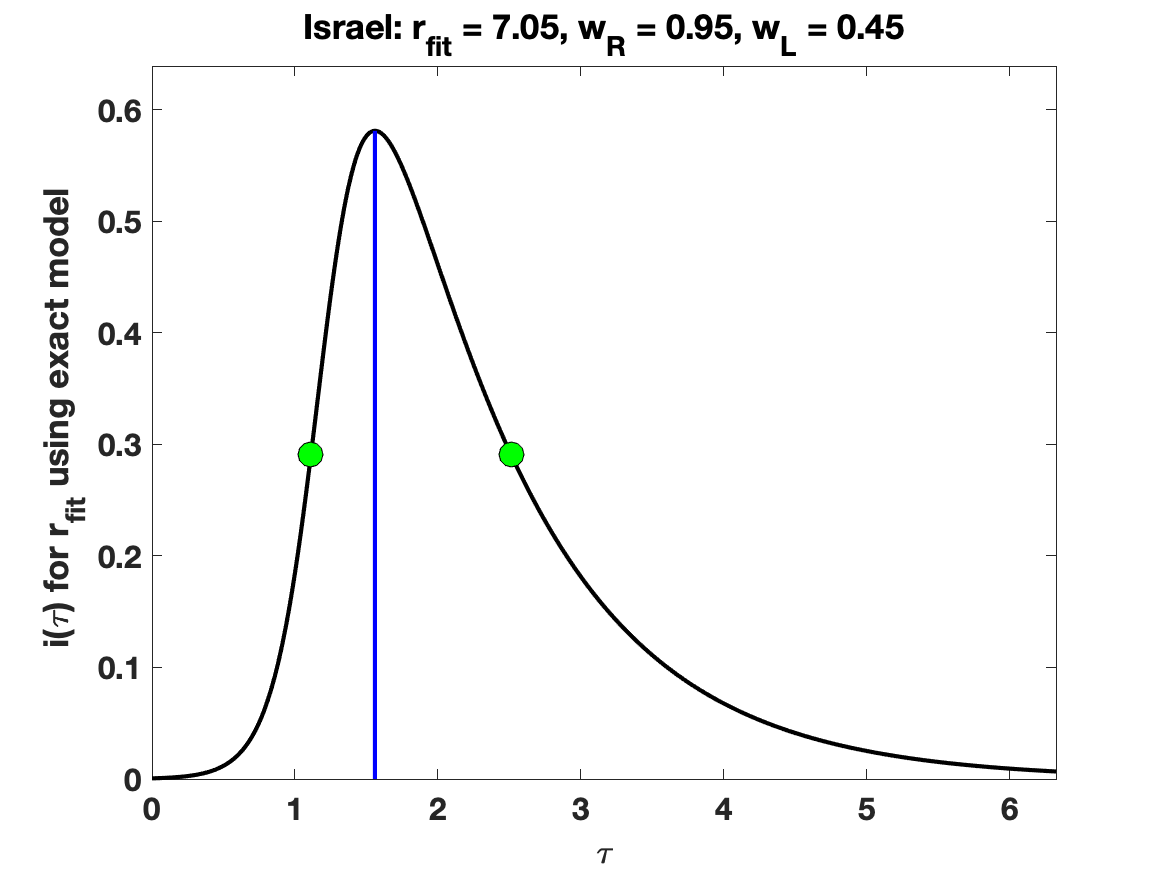

### Israel_solution_fit.png

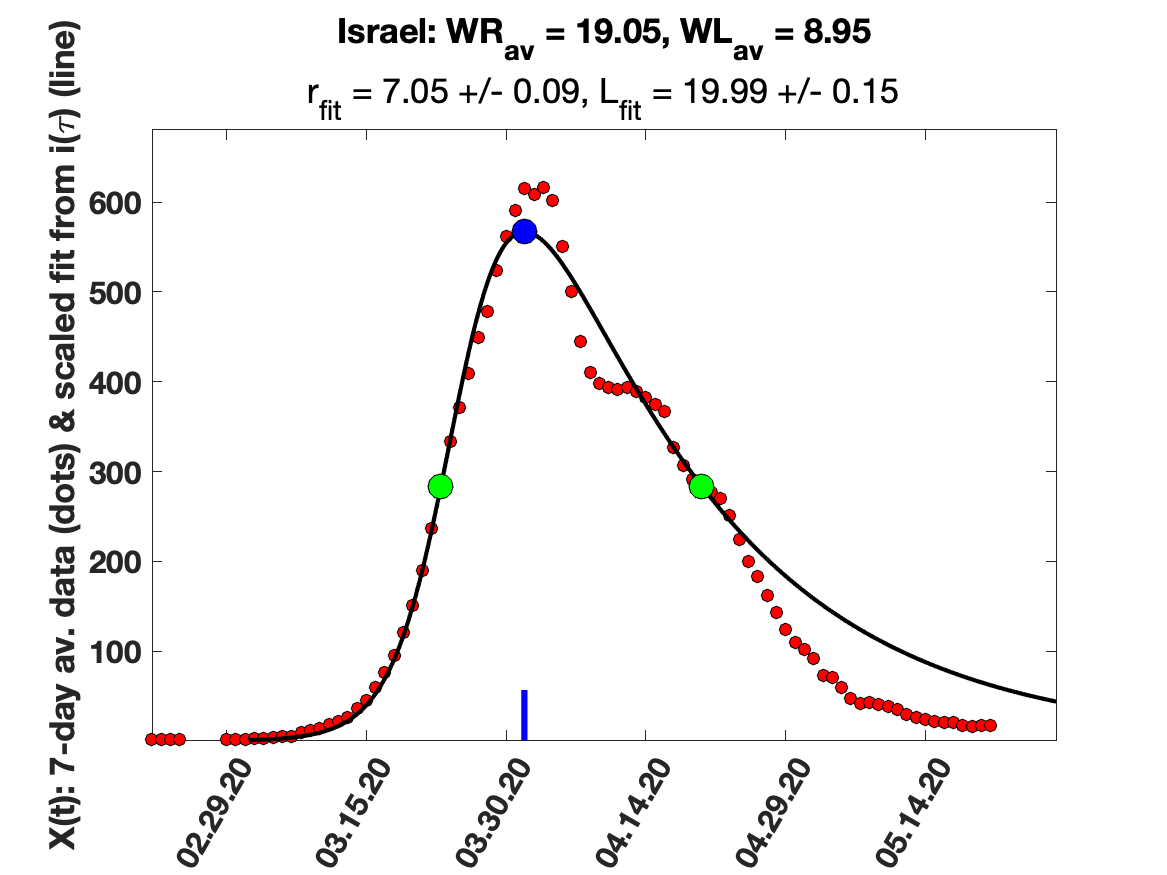

### Italy_5_day_av_and_fit.png

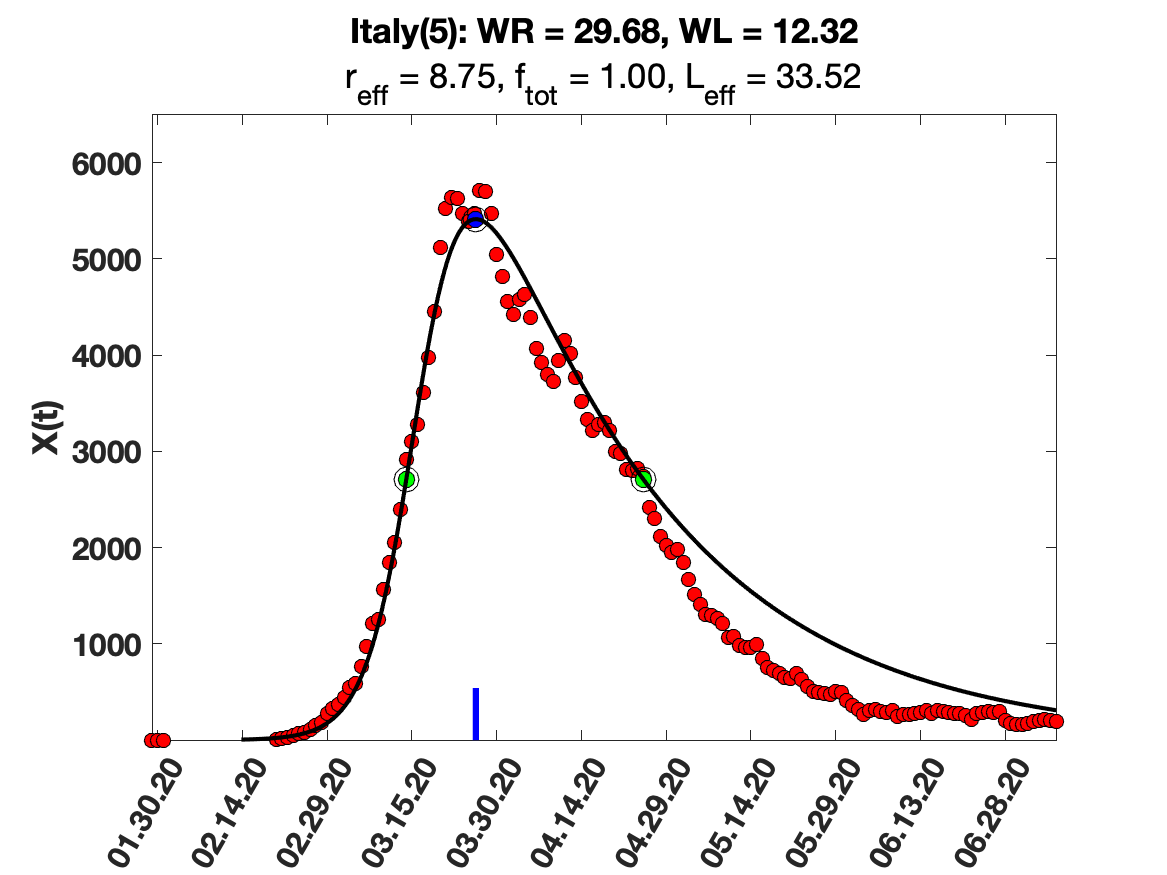

### Italy_7_day_av_and_fit.png

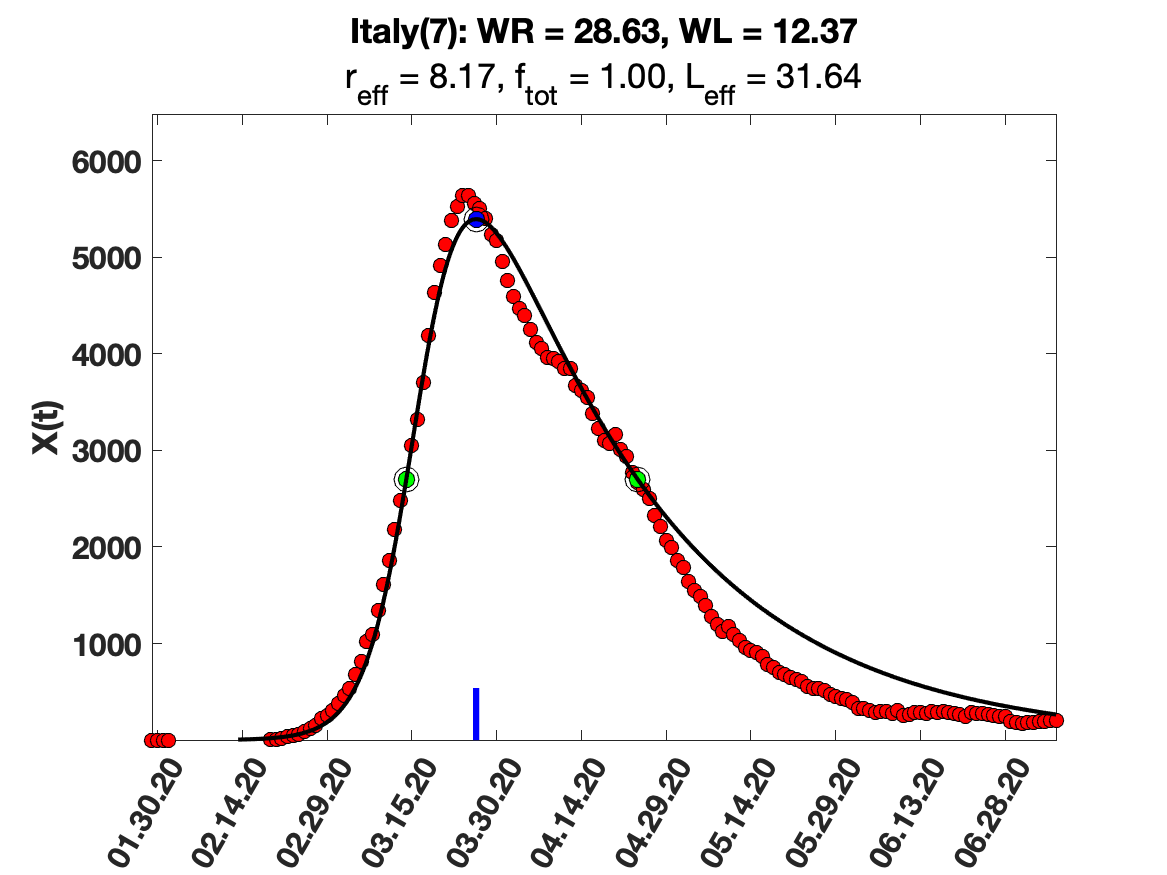

### Italy_9_day_av_and_fit.png

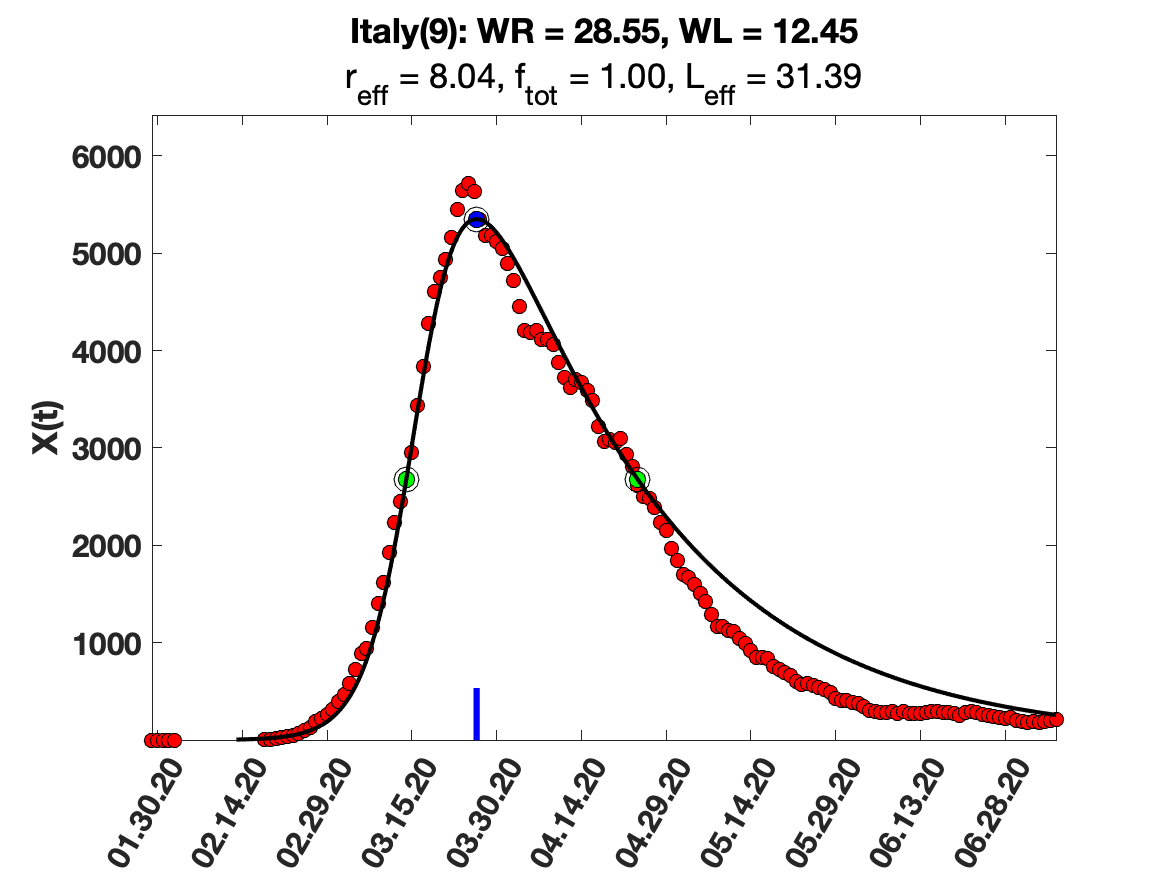

### Italy_i(tau)_for_average_r.png

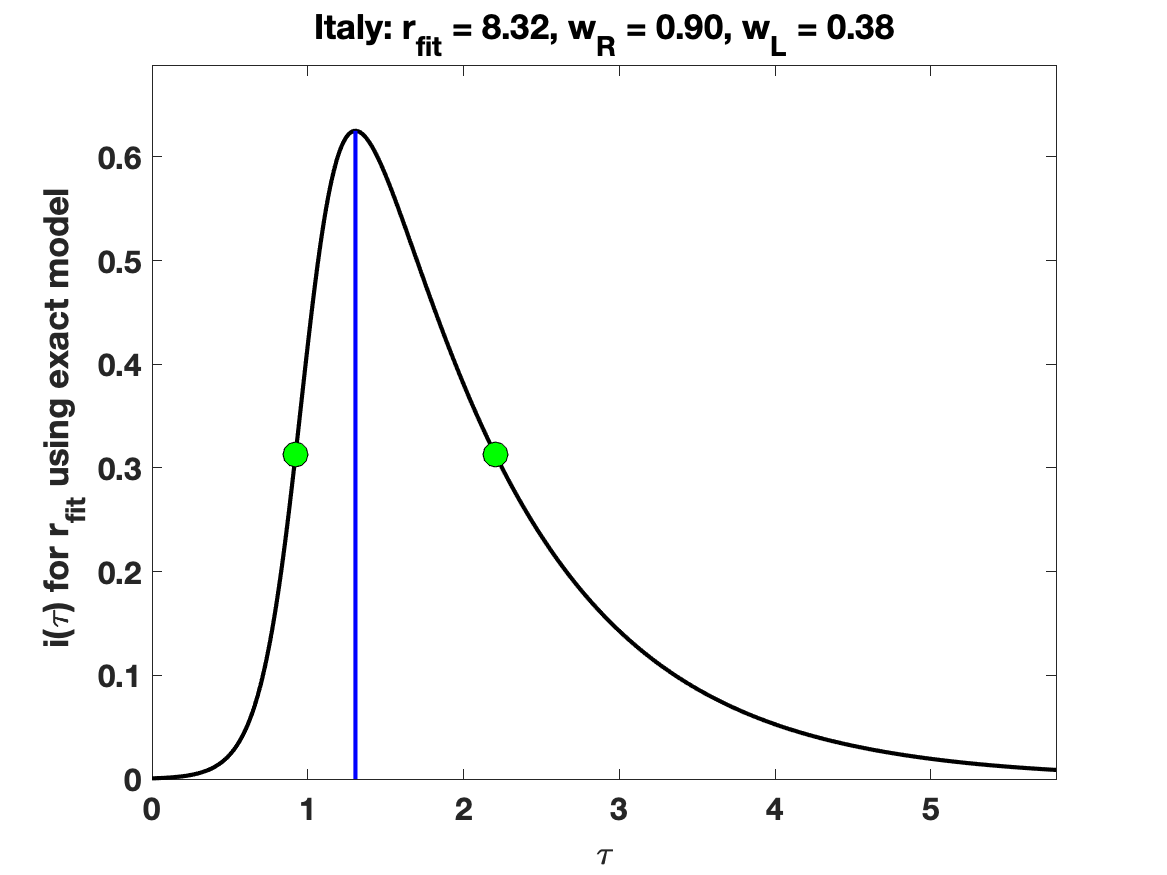

### Italy_raw_data.png

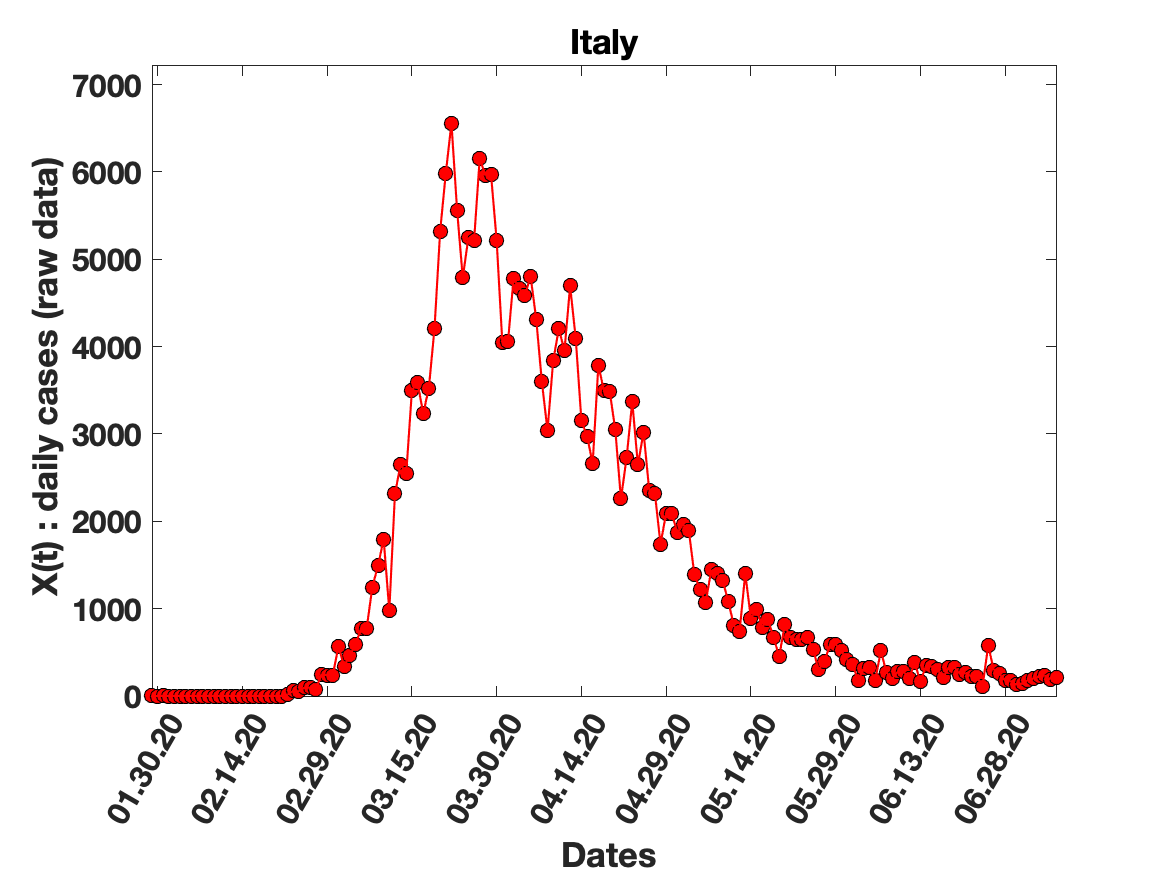

### Italy_solution_fit.png

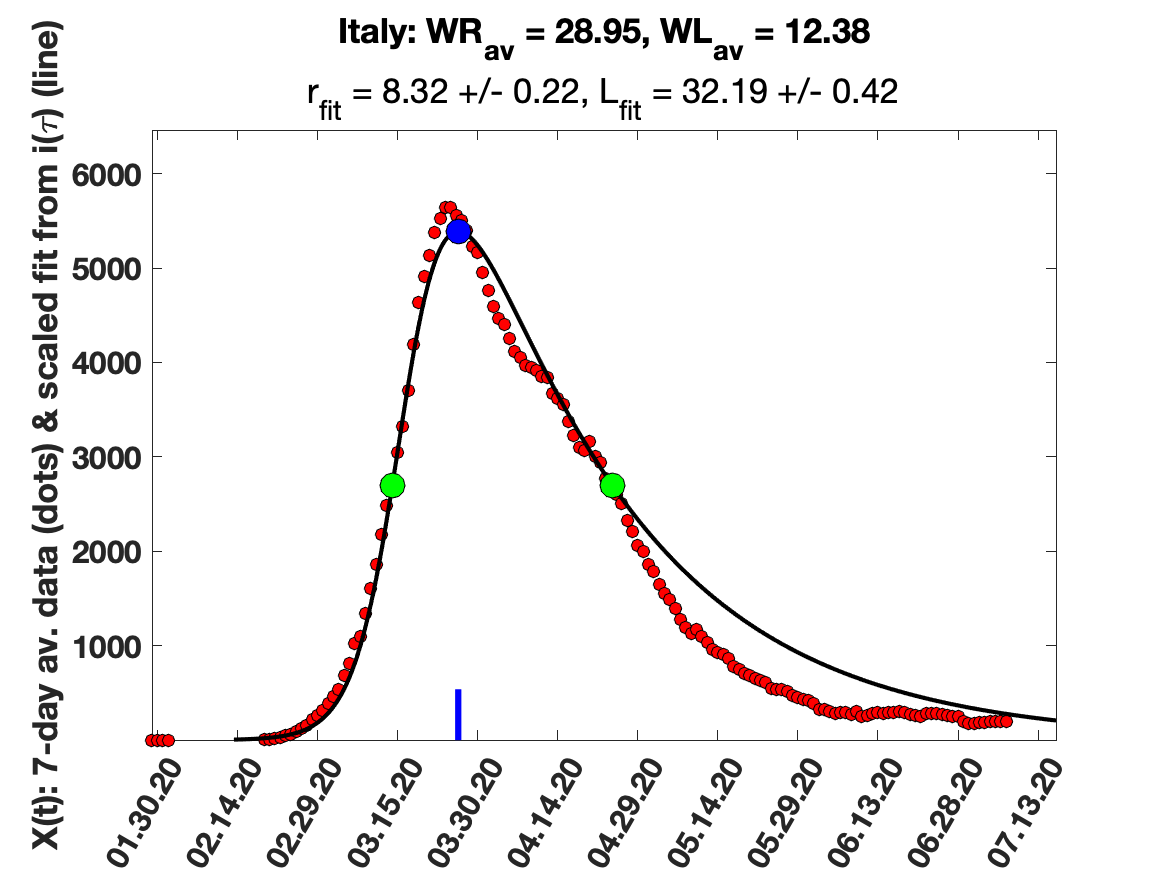

### Japan_7_day_av_and_fit.png

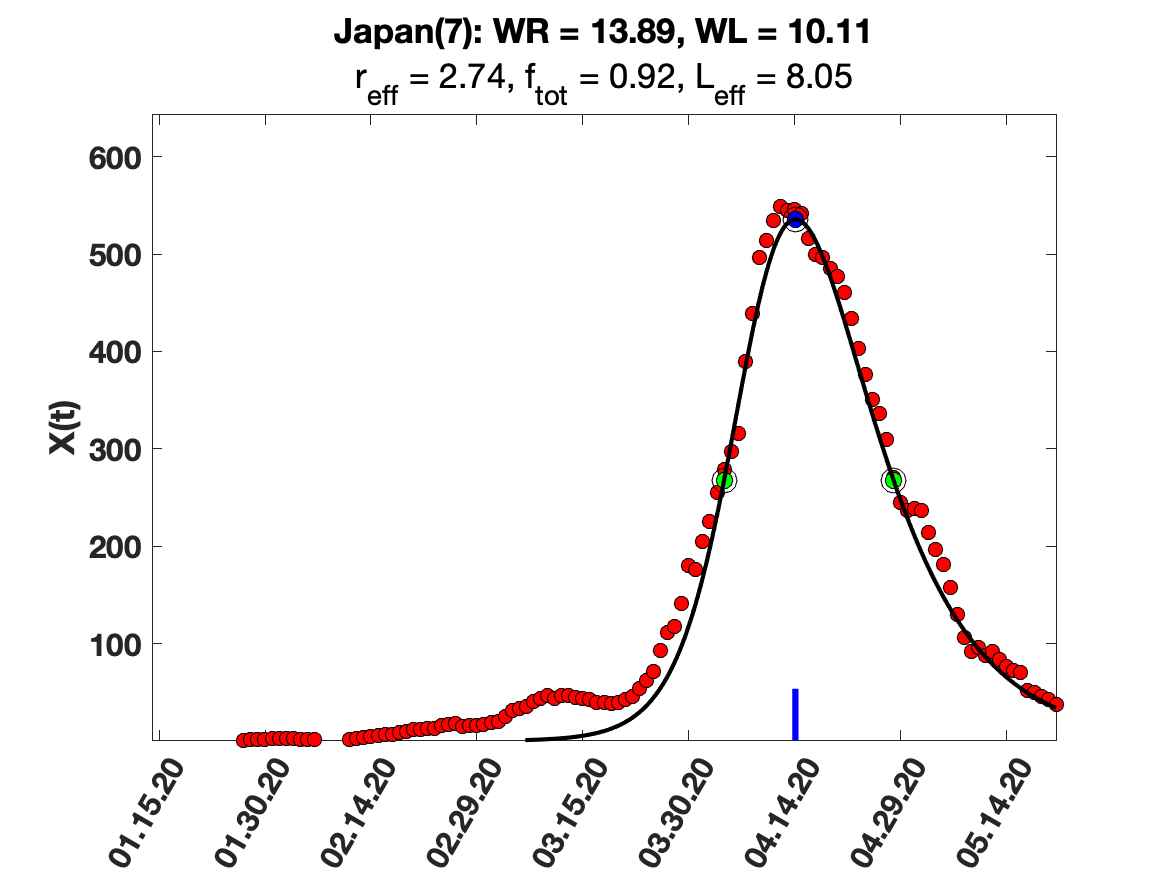

### Japan_9_day_av_and_fit.png

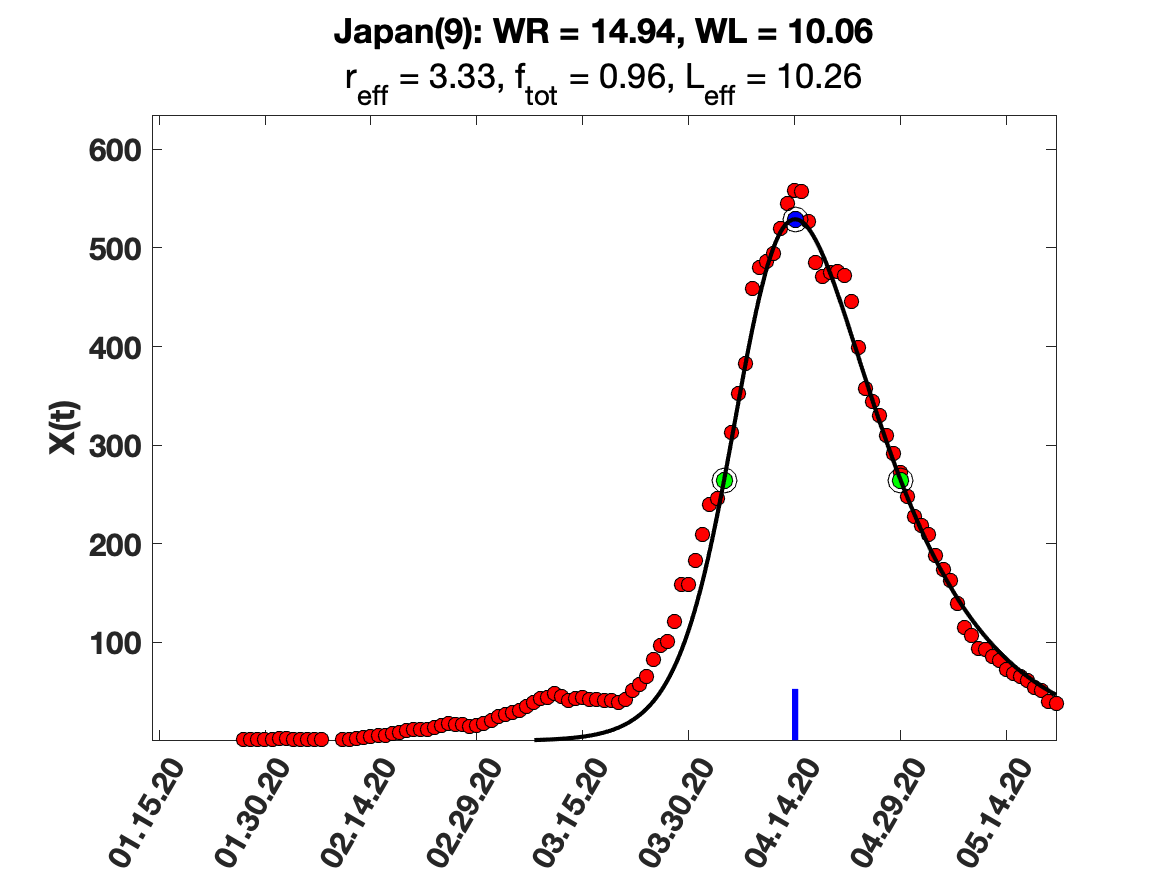

### Japan_i(tau)_for_average_r.png

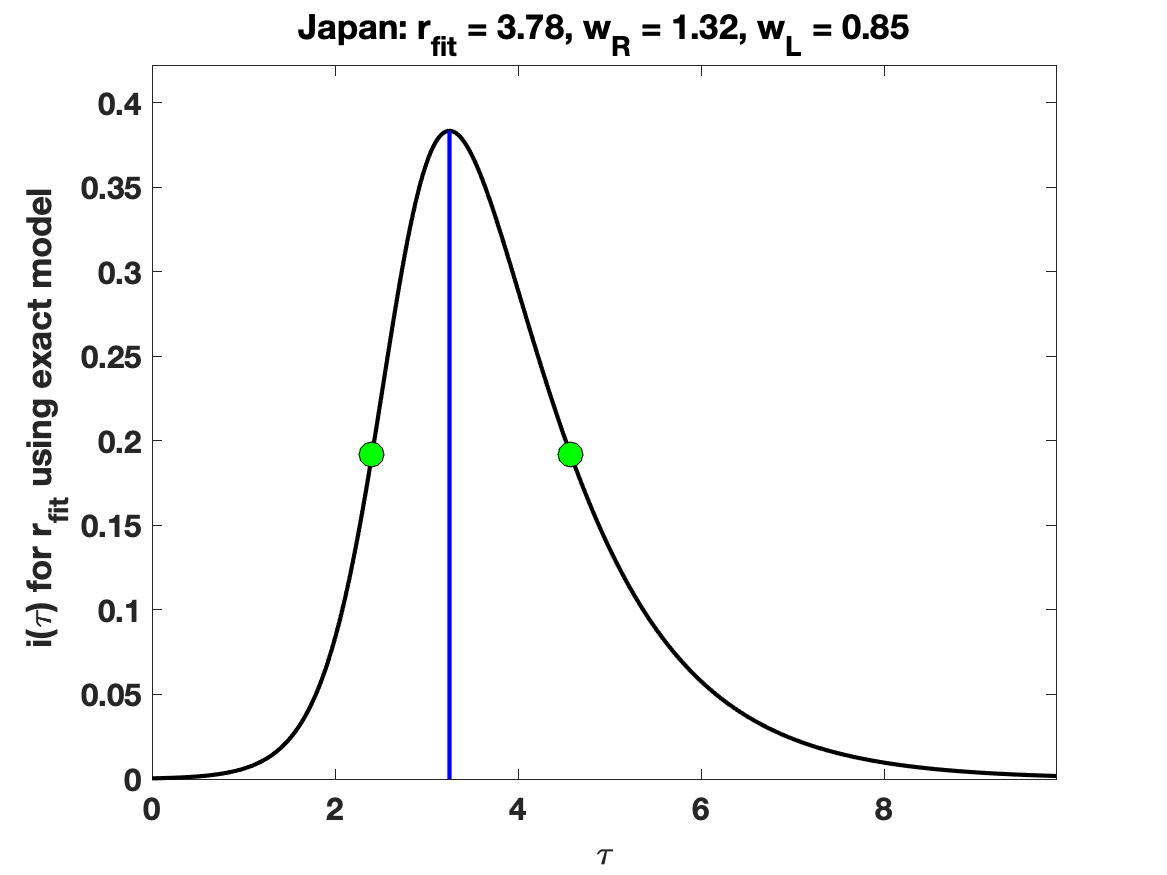

### Japan_raw_data.png

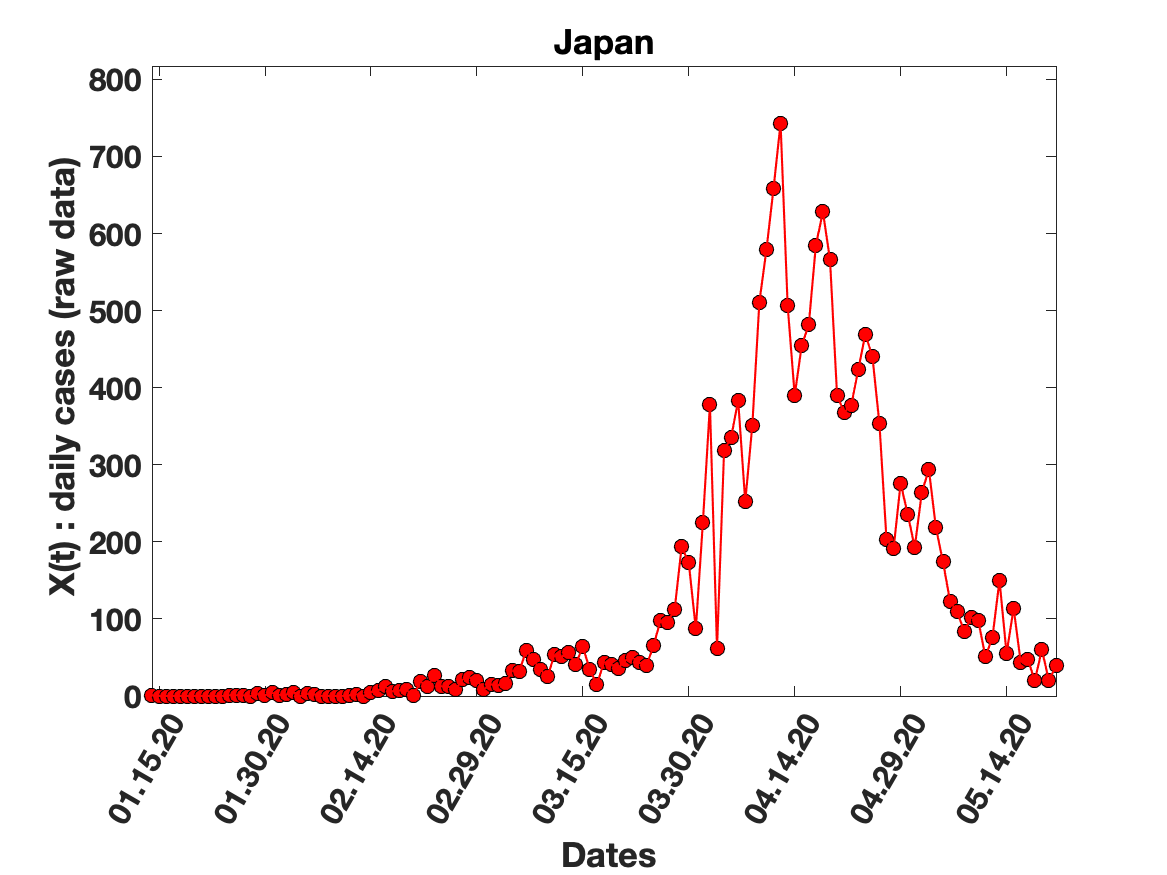

### Japan_solution_fit.png

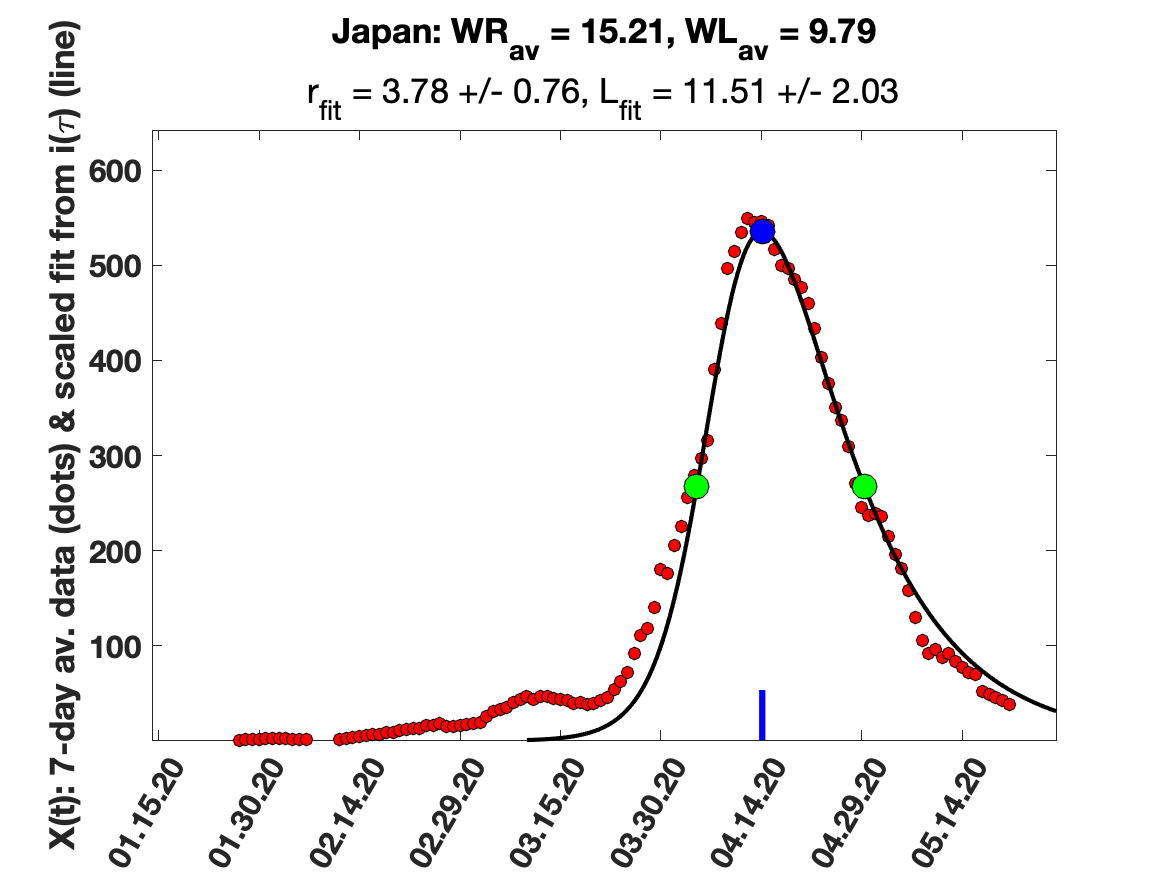

### Netherlands_5_day_av_and_fit.png

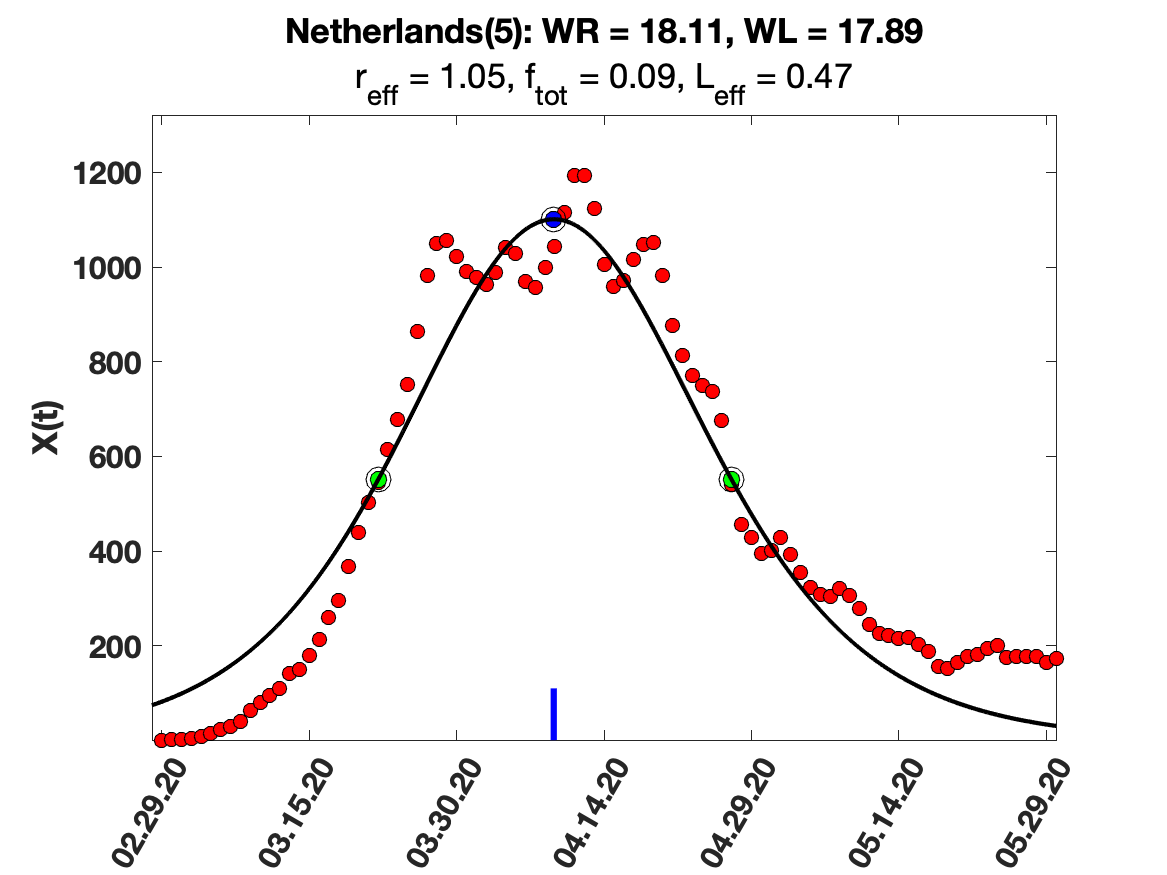

### Netherlands_i(tau)_for_average_r.png

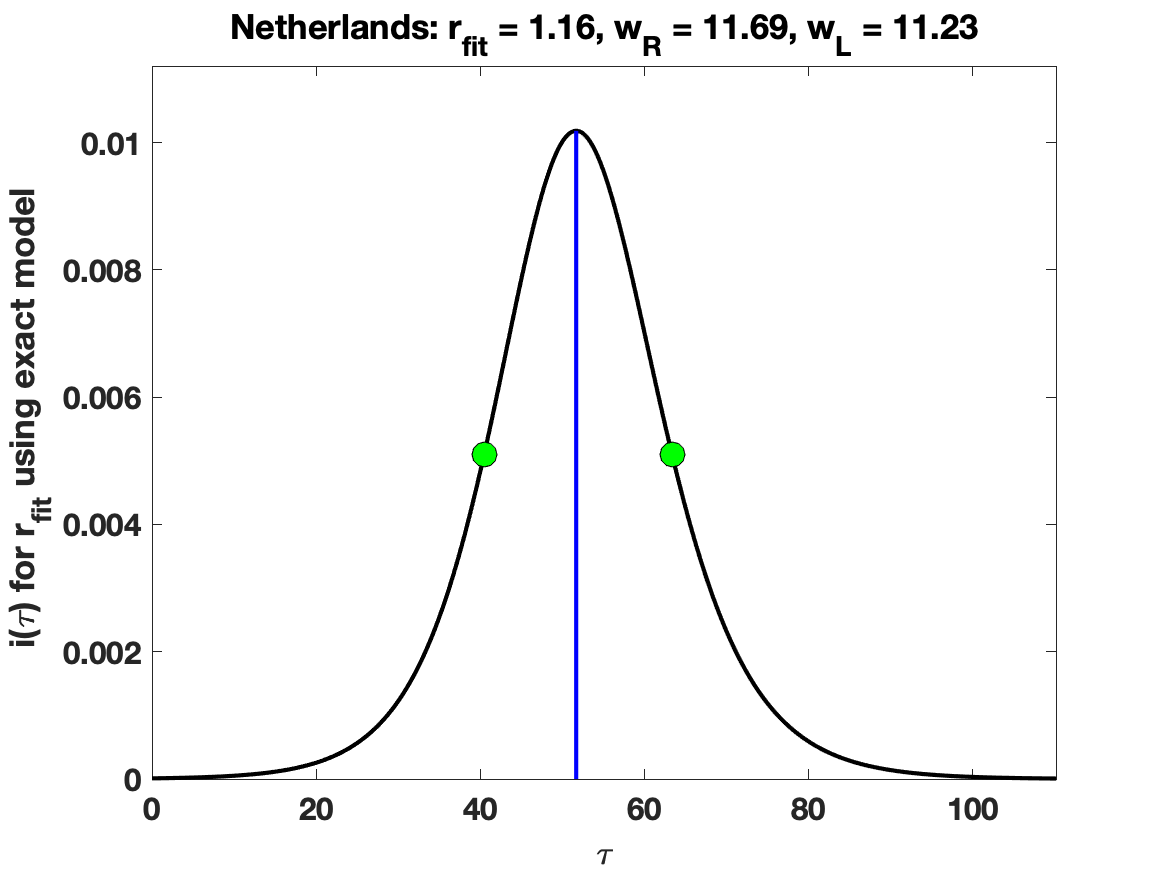

### Netherlands_raw_data.png

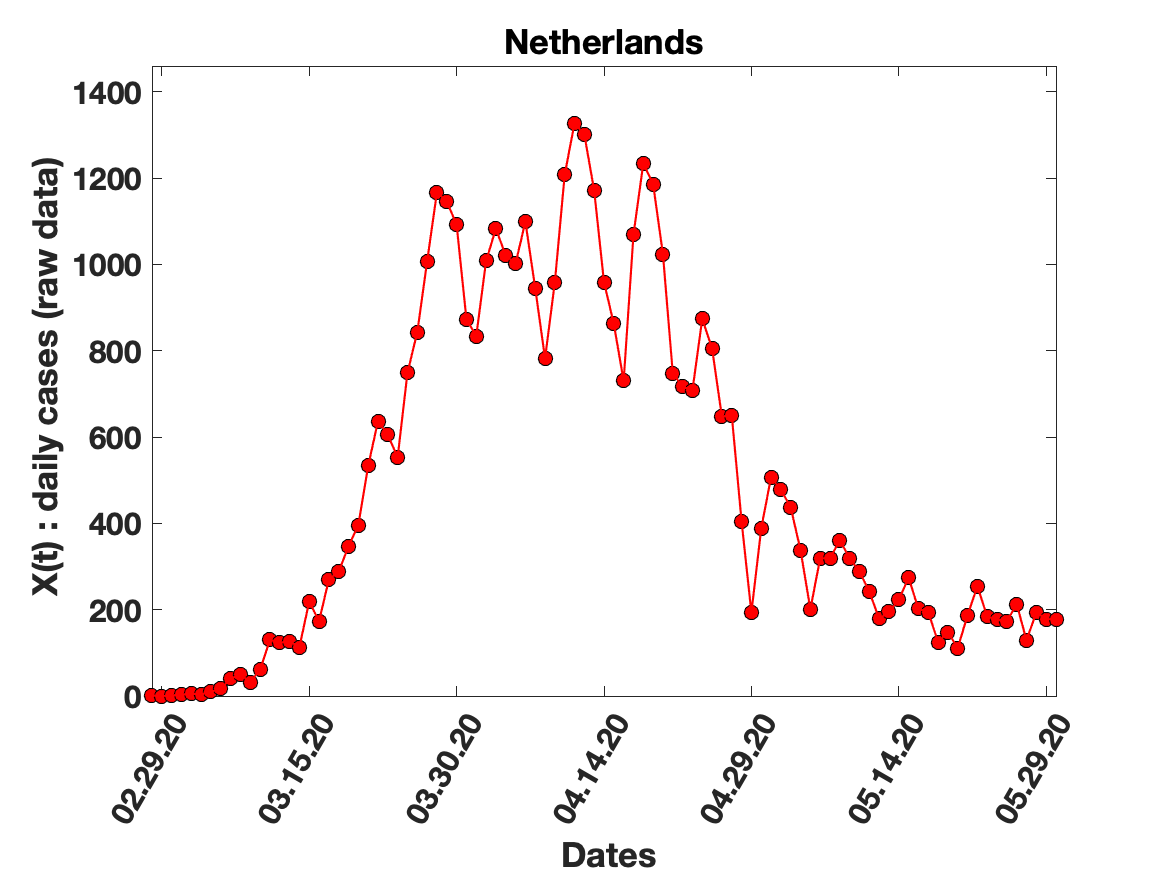

### Netherlands_solution_fit.png

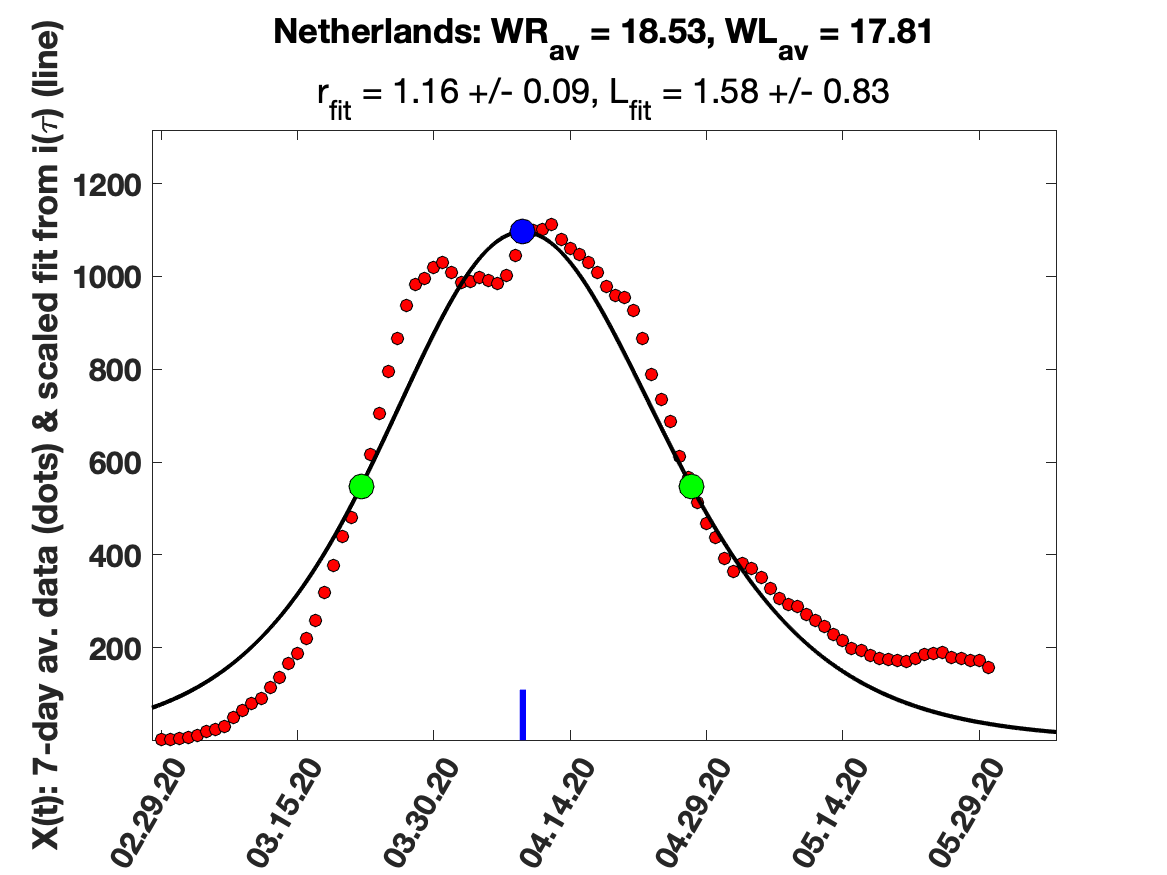

### New_Zealand_5_day_av_and_fit.png

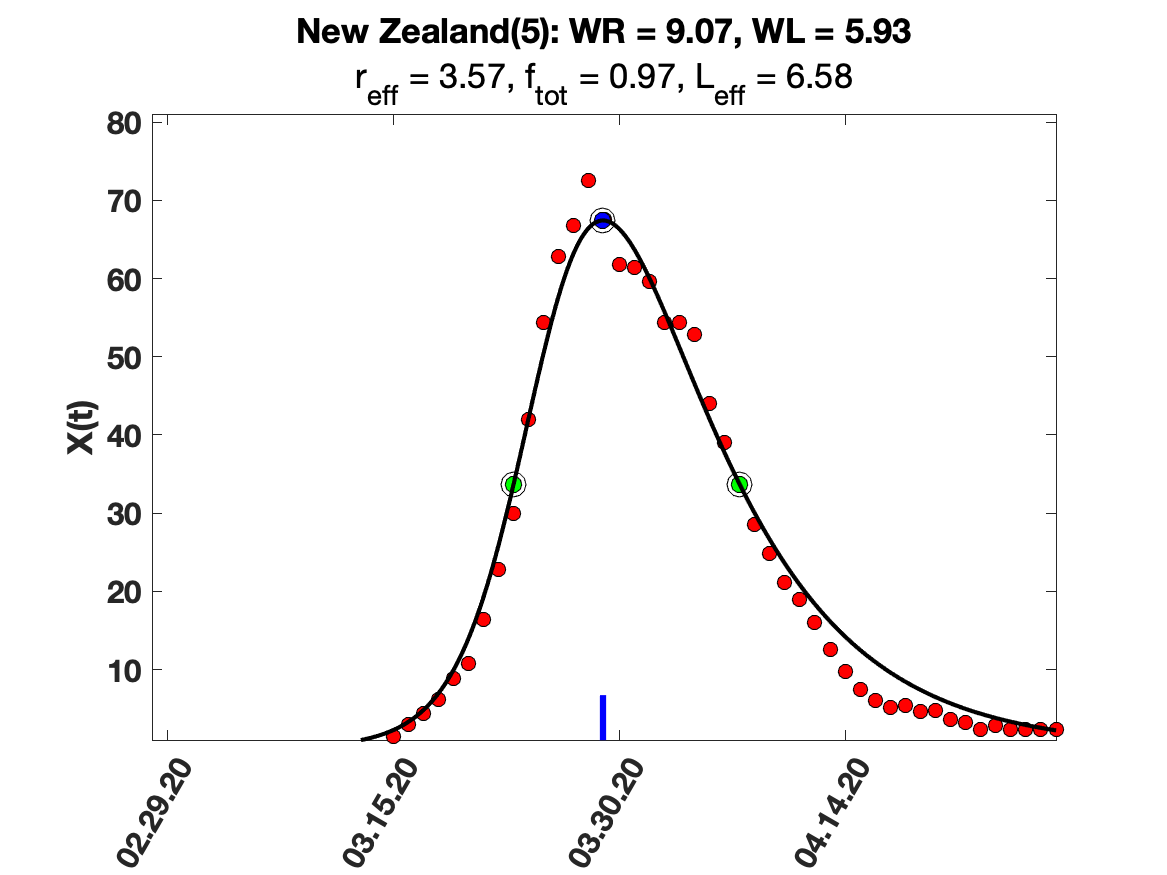

### New_Zealand_7_day_av_and_fit.png

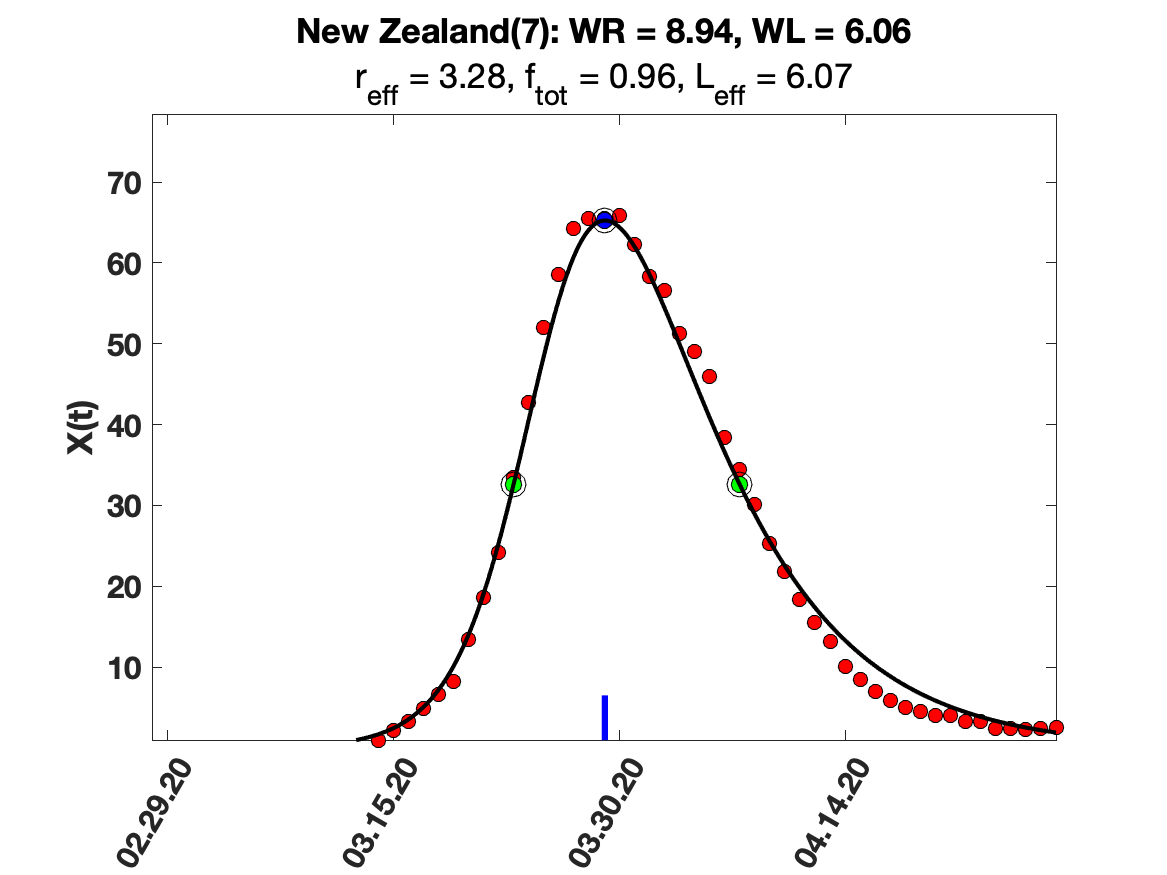

### New_Zealand_9_day_av_and_fit.png

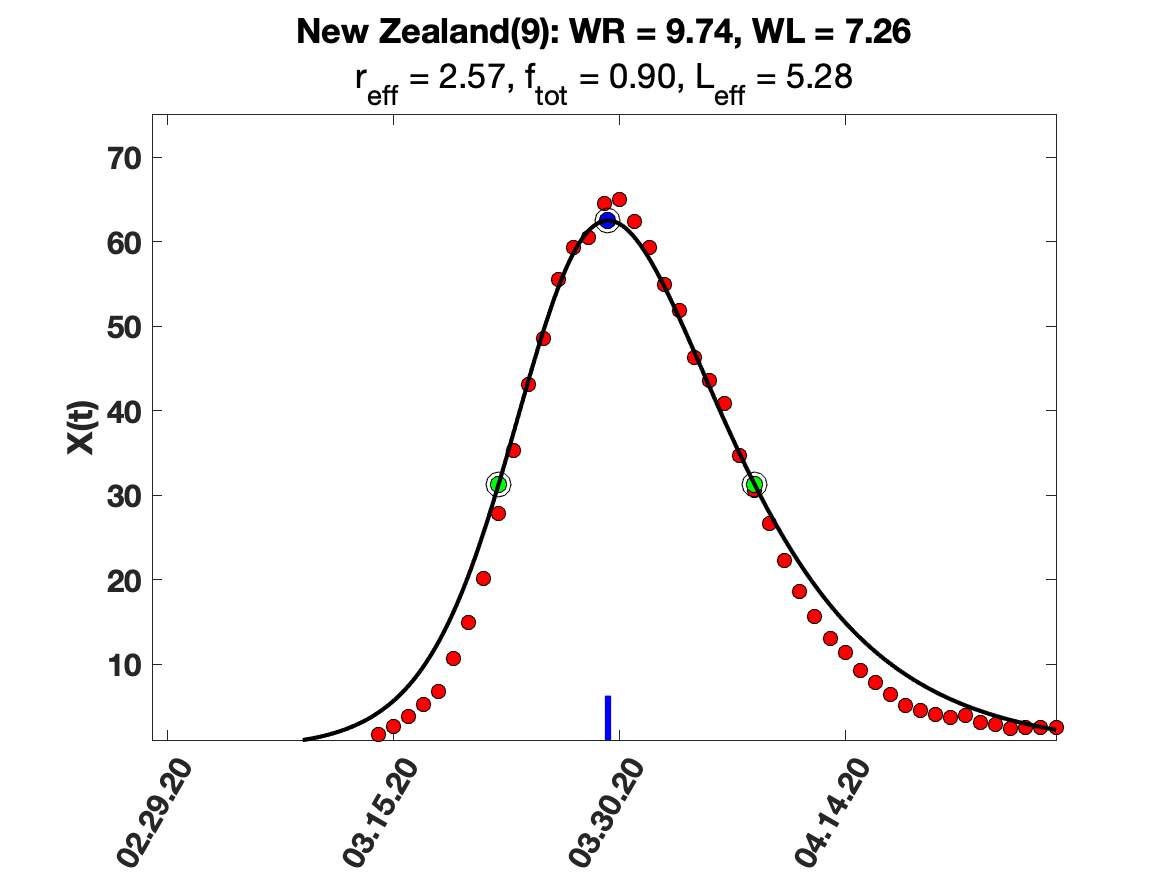

### New_Zealand_i(tau)_for_average_r.png

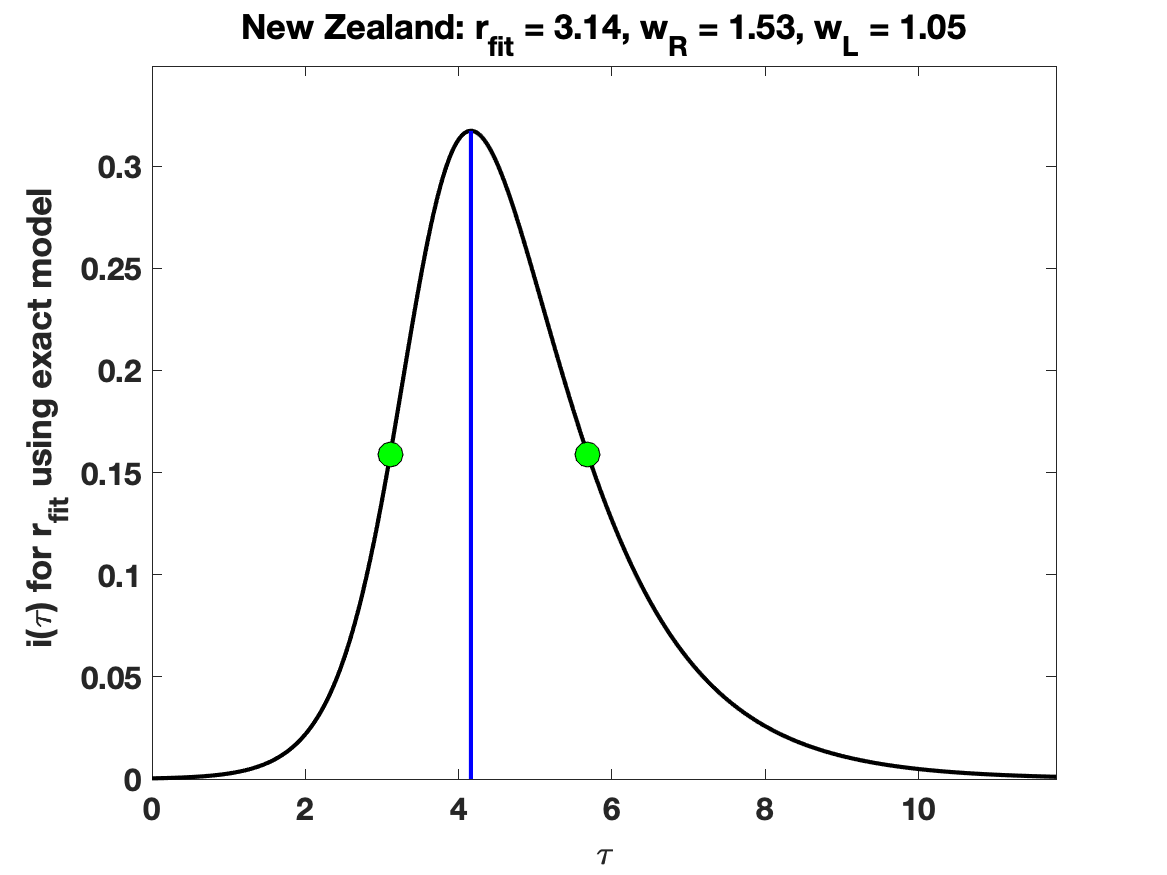

### New_Zealand_raw_data.png

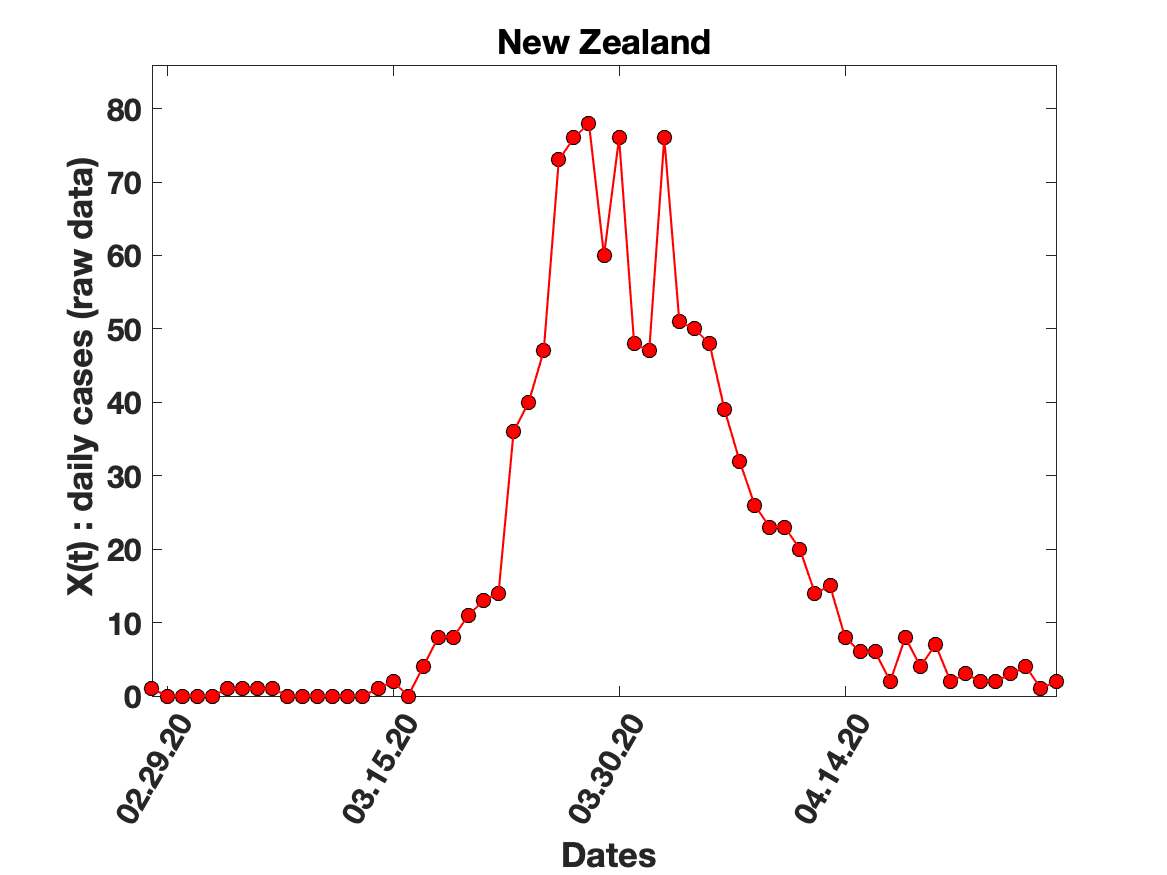

### Portugal_7_day_av_and_fit.png

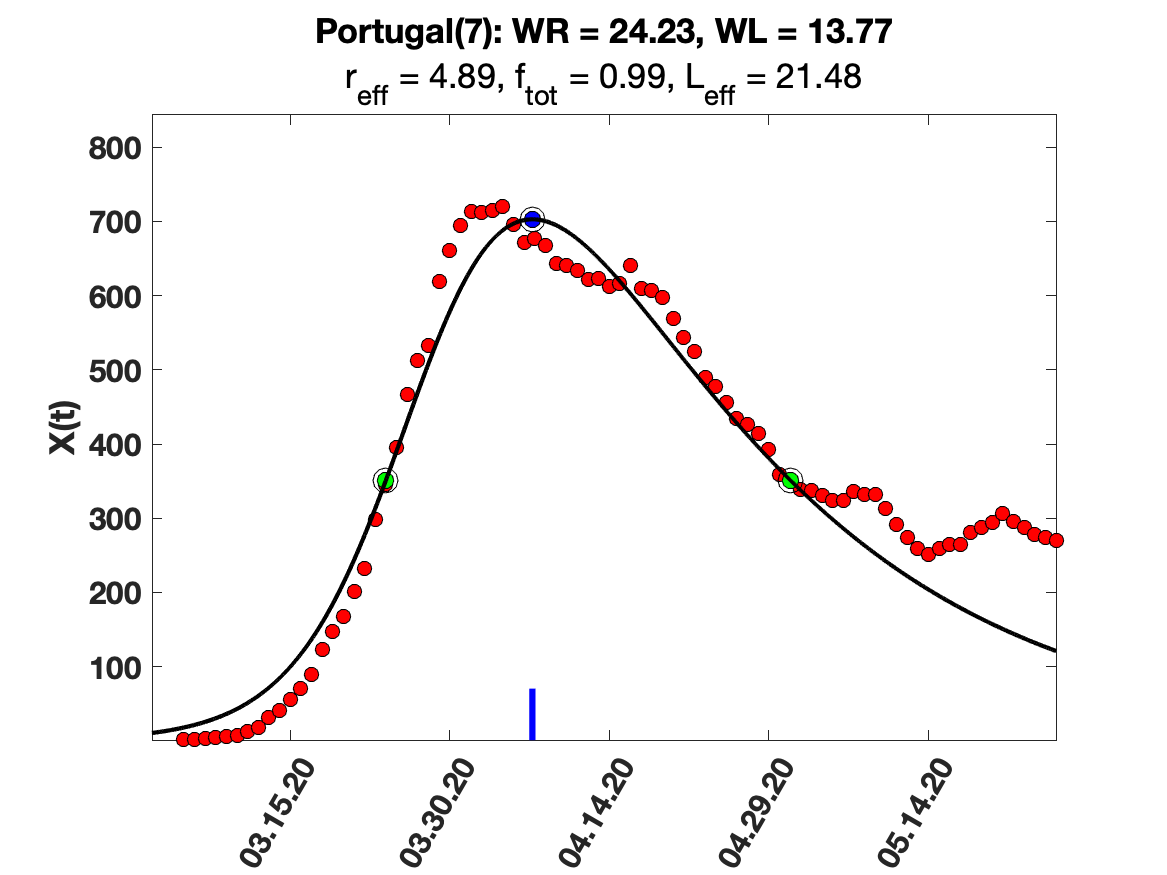

### Portugal_9_day_av_and_fit.png

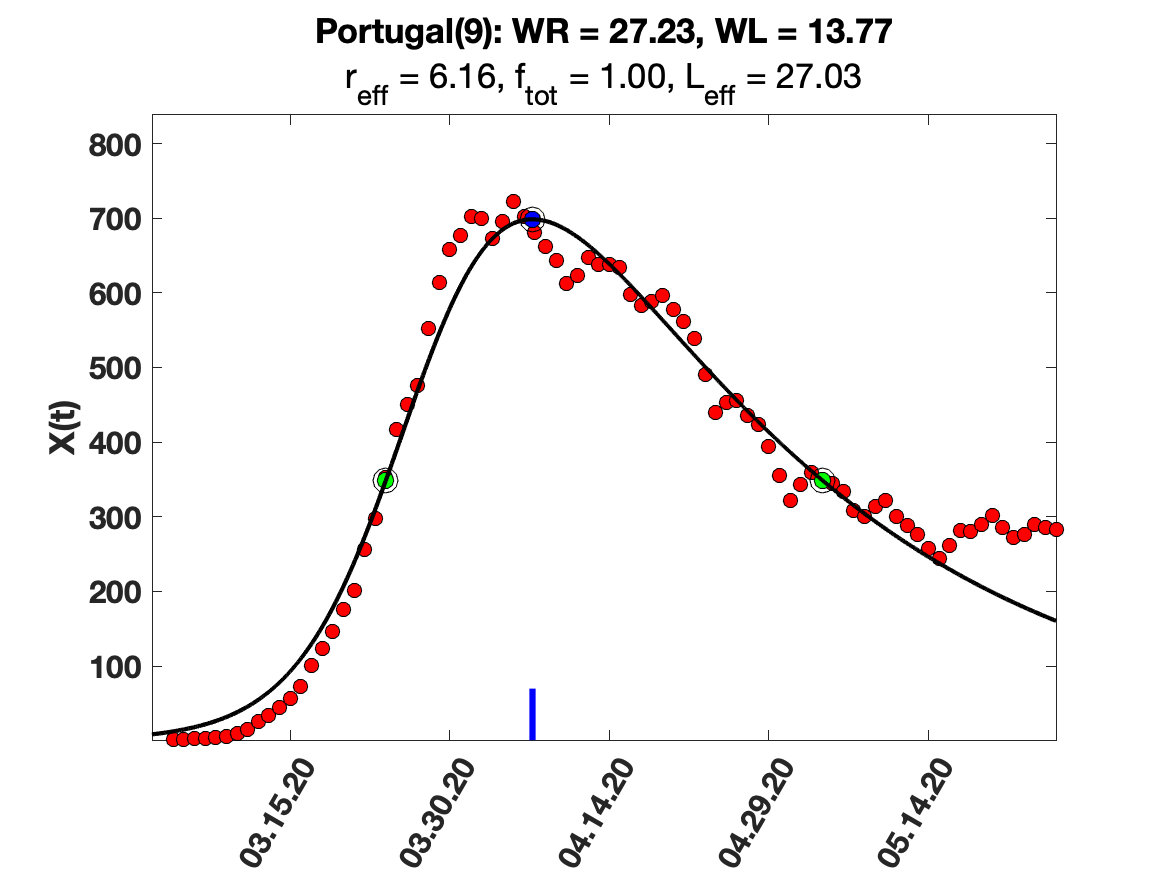

### Portugal_i(tau)_for_average_r.png

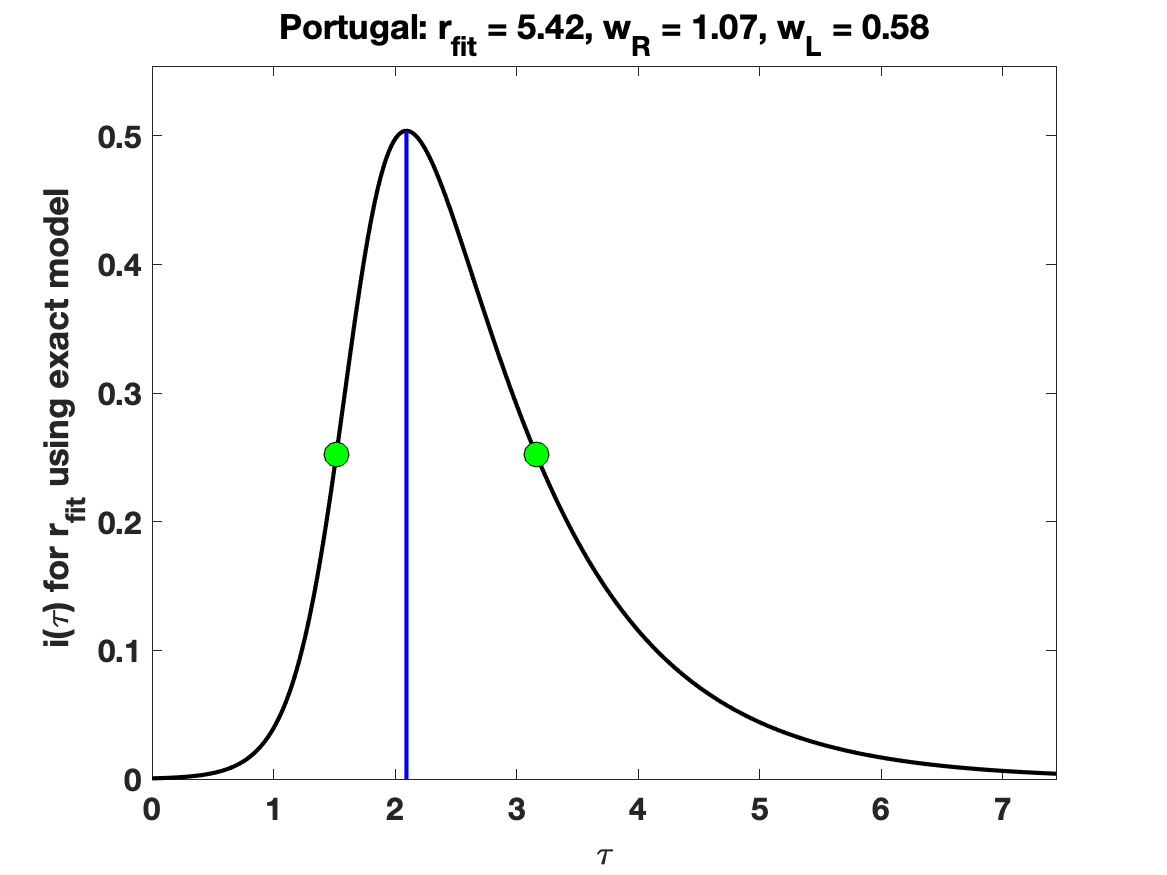

### Portugal_raw_data.png

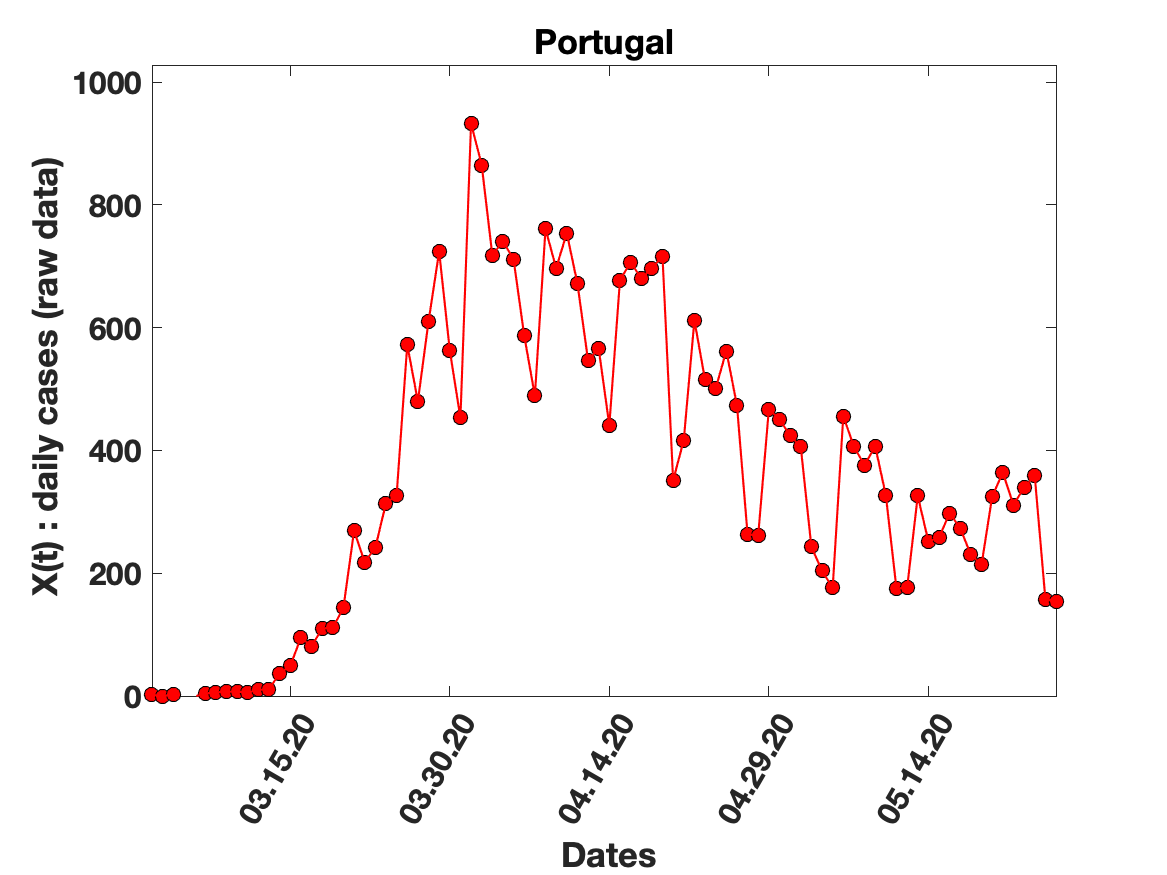

### Portugal_solution_fit.png

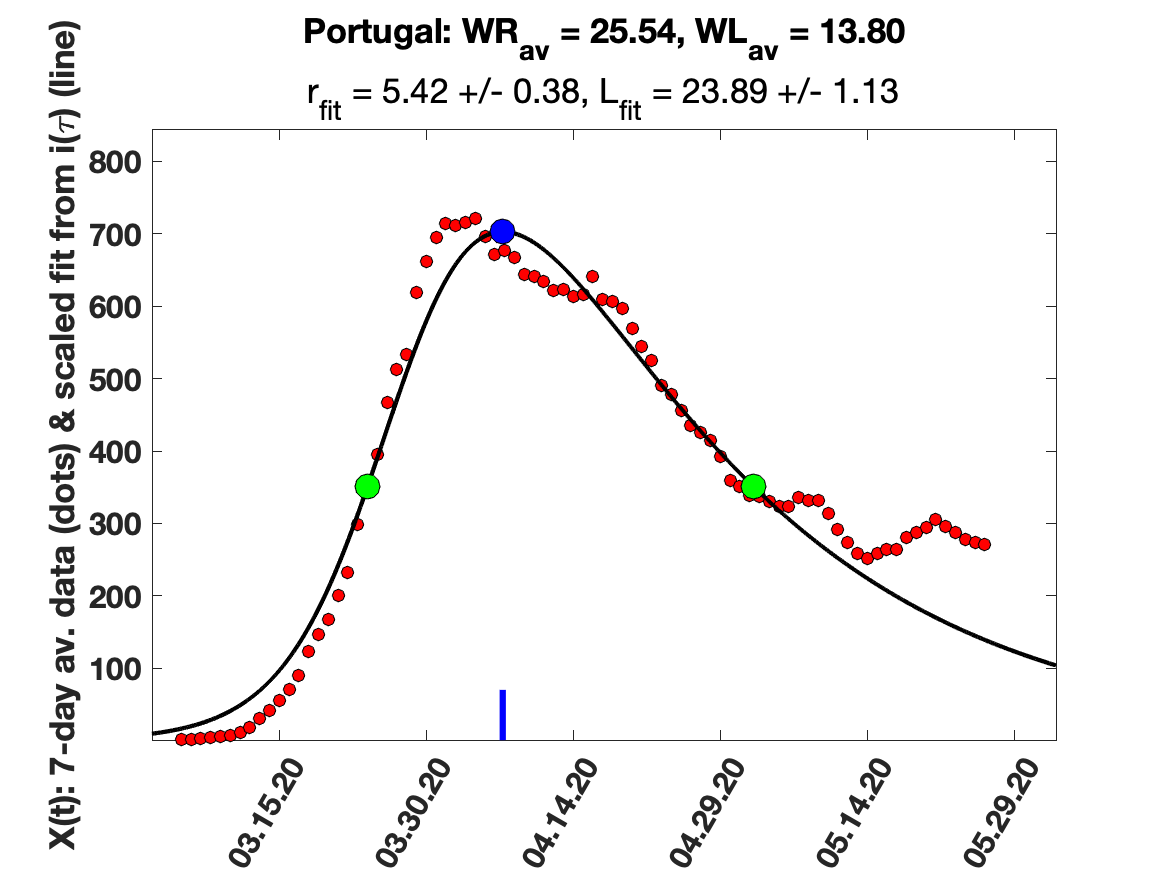

### Qatar_7_day_av_and_fit.png

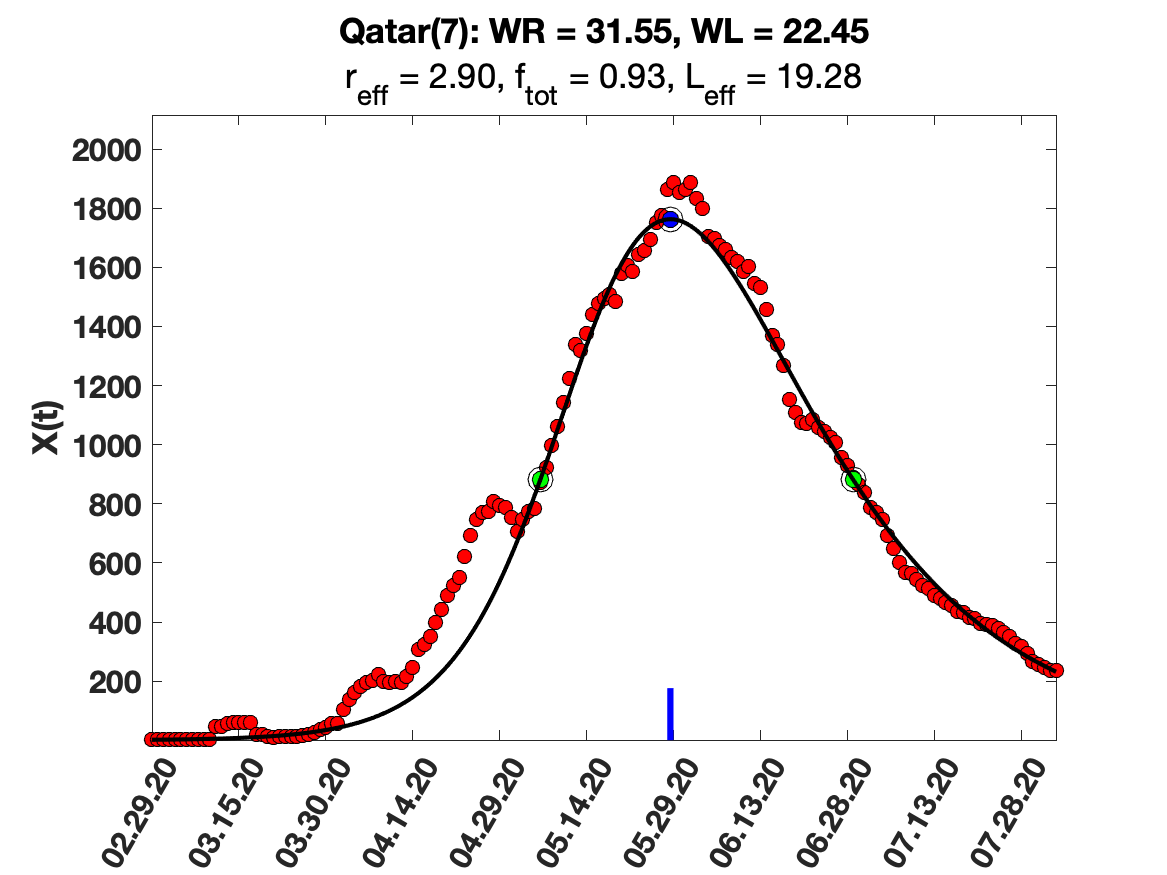

### Qatar_9_day_av_and_fit.png

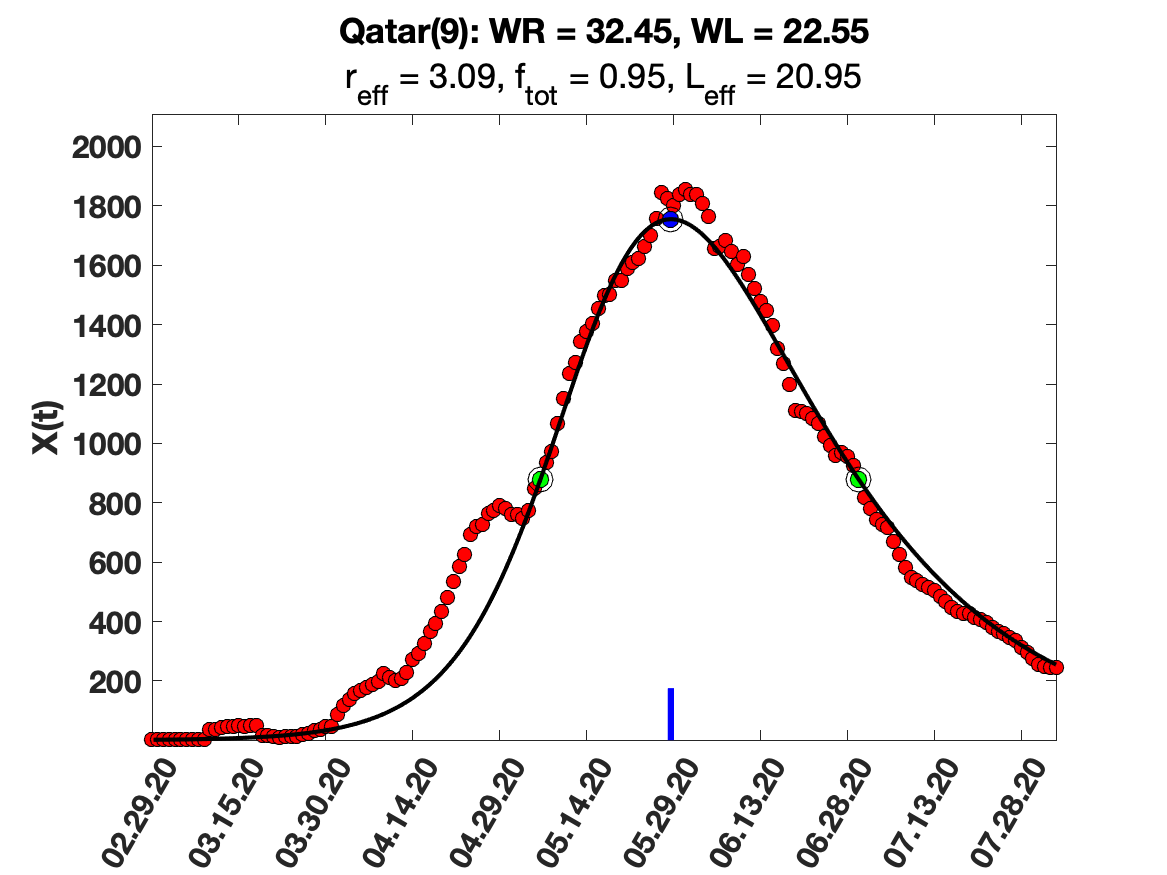

### Qatar_i(tau)_for_average_r.png

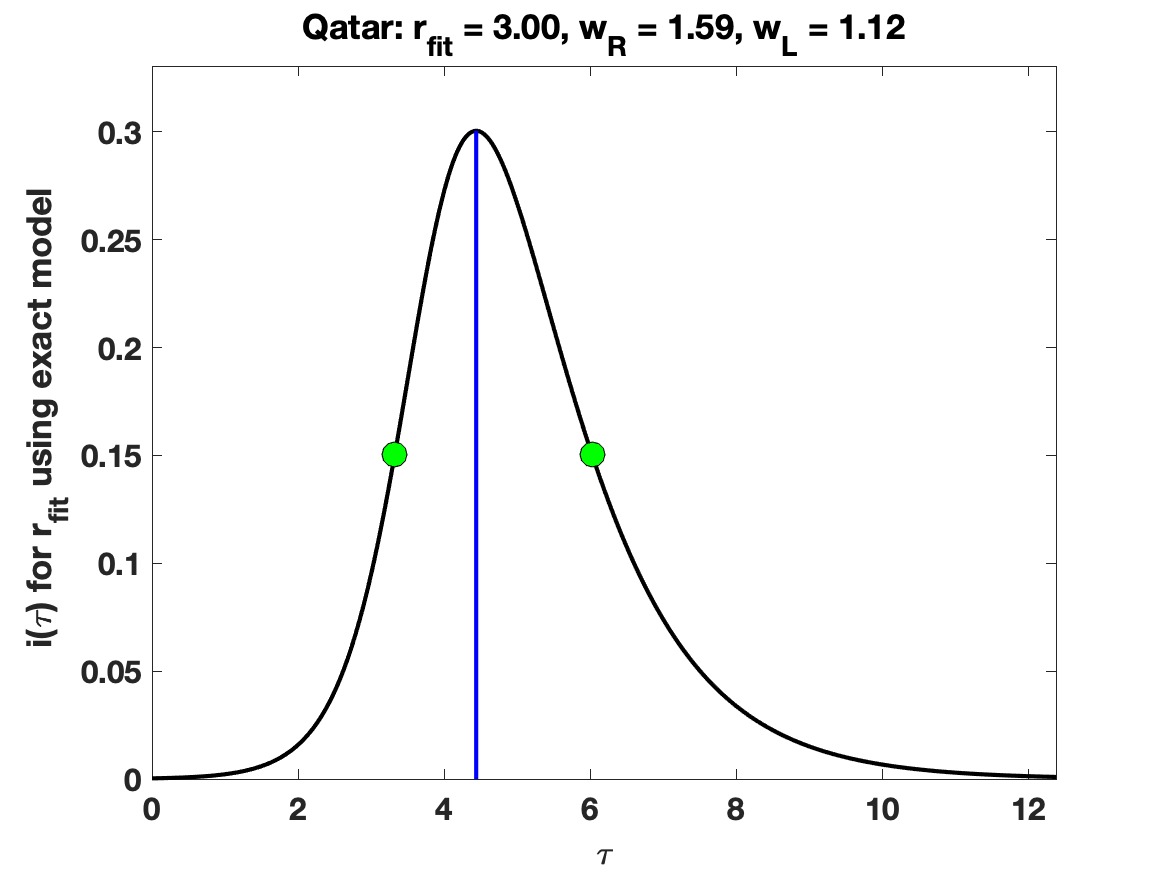

### Republic_of_Korea_5_day_av_and_fit.png

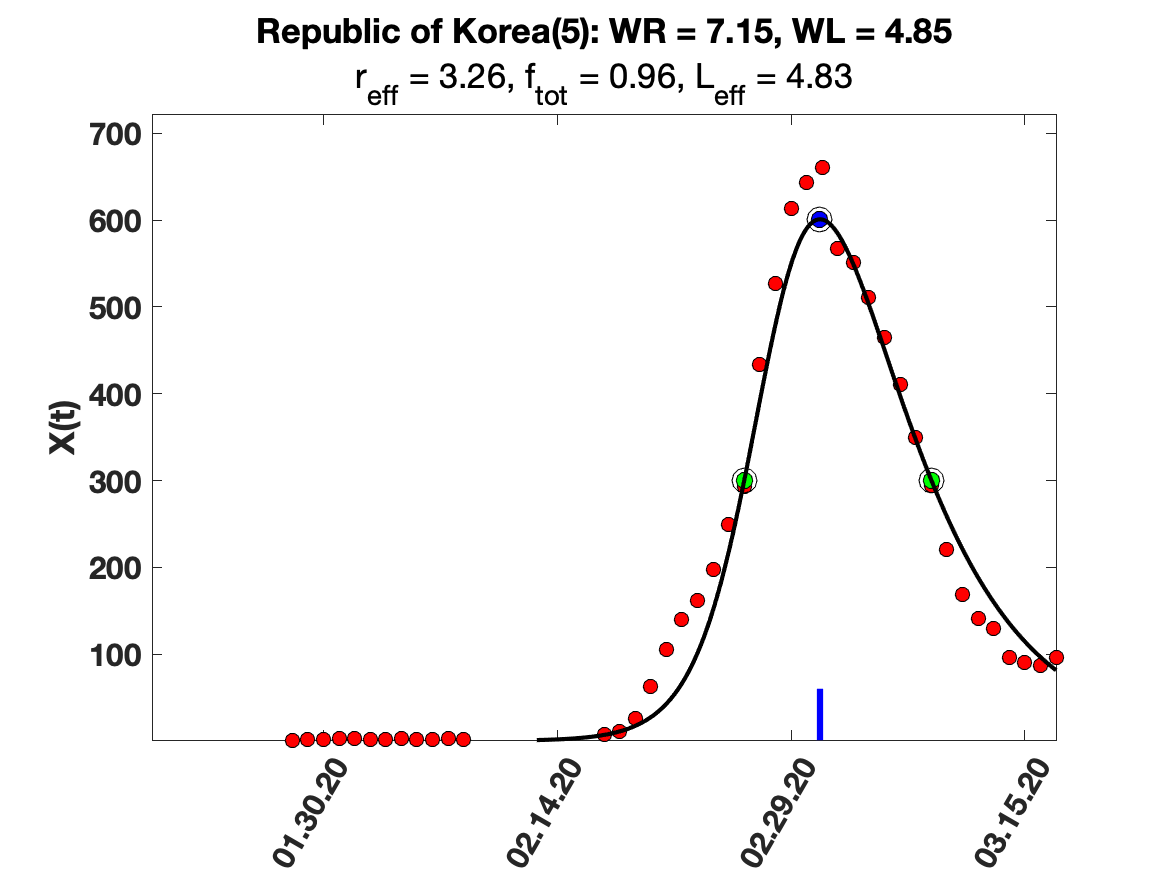

### Republic_of_Korea_7_day_av_and_fit.png

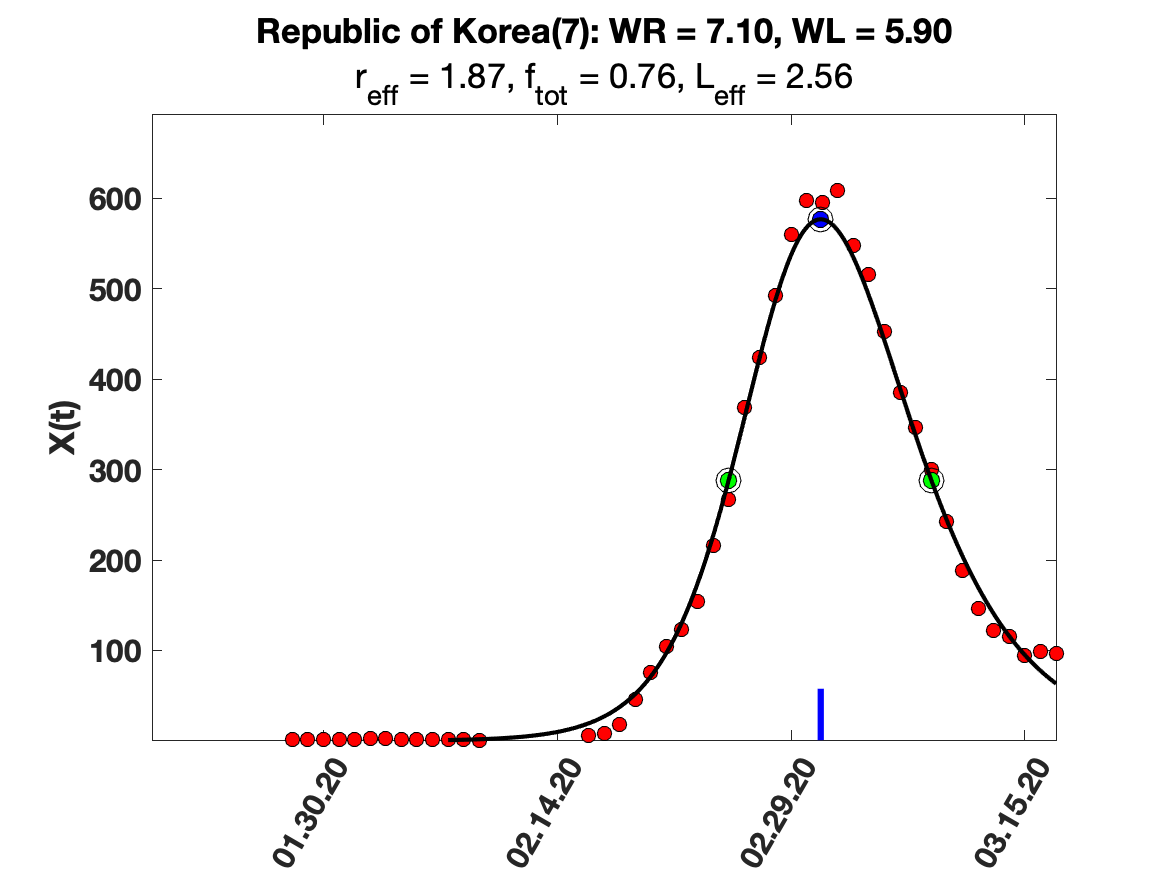

### Republic_of_Korea_9_day_av_and_fit.png

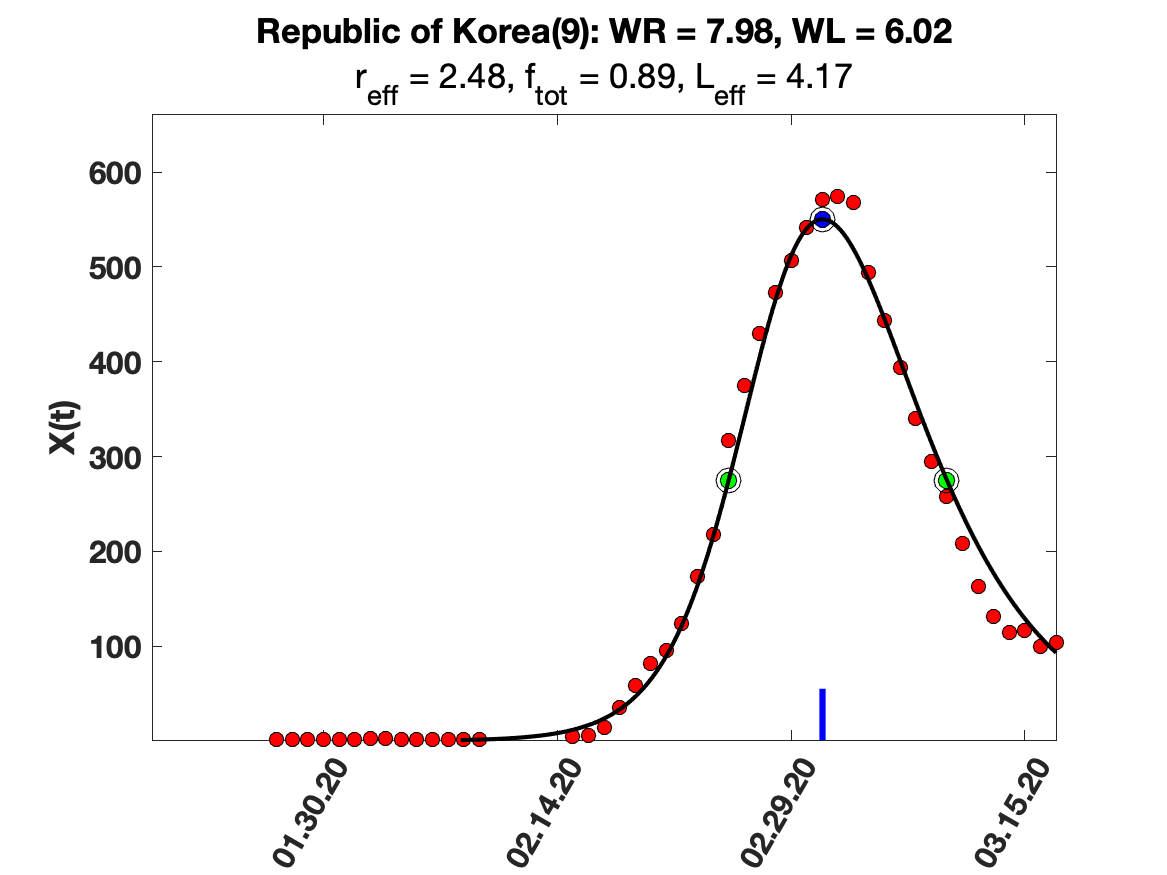

### Republic_of_Korea_i(tau)_for_average_r.png

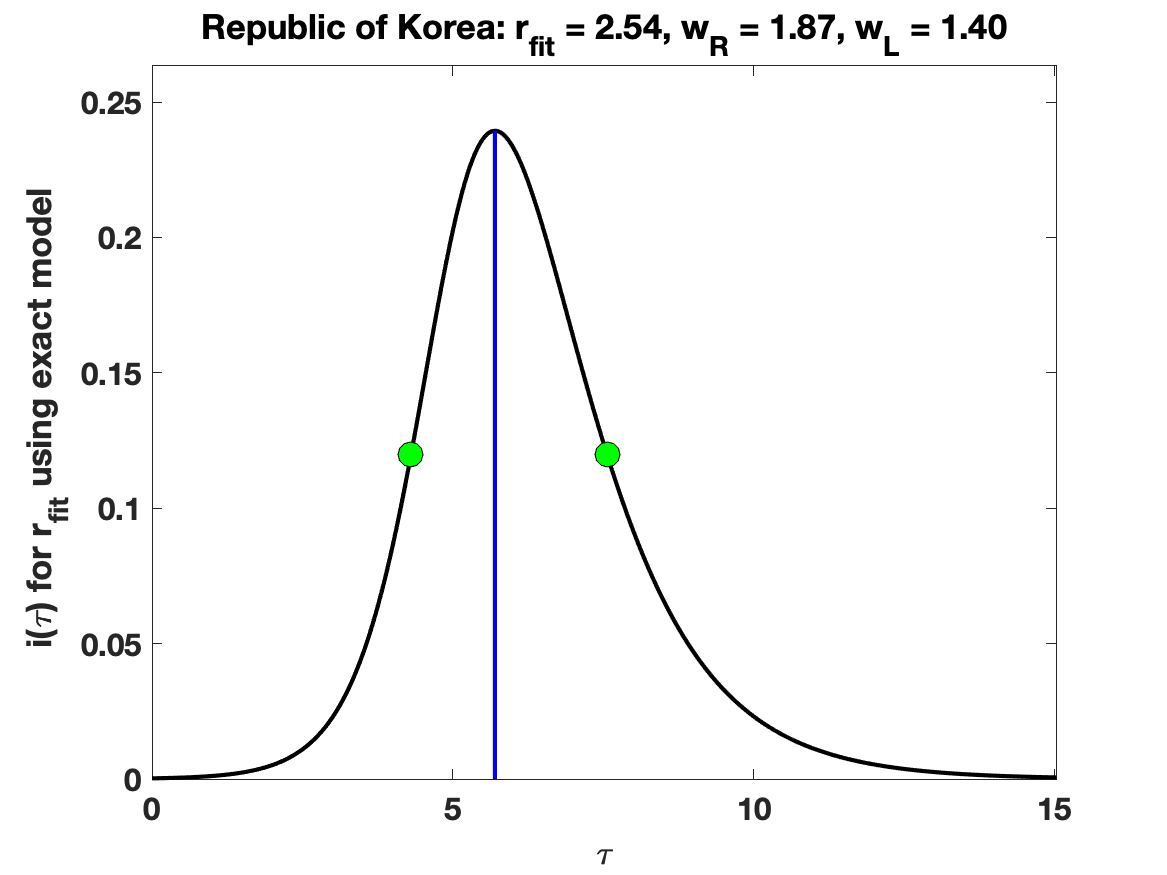

### Republic_of_Korea_raw_data.png

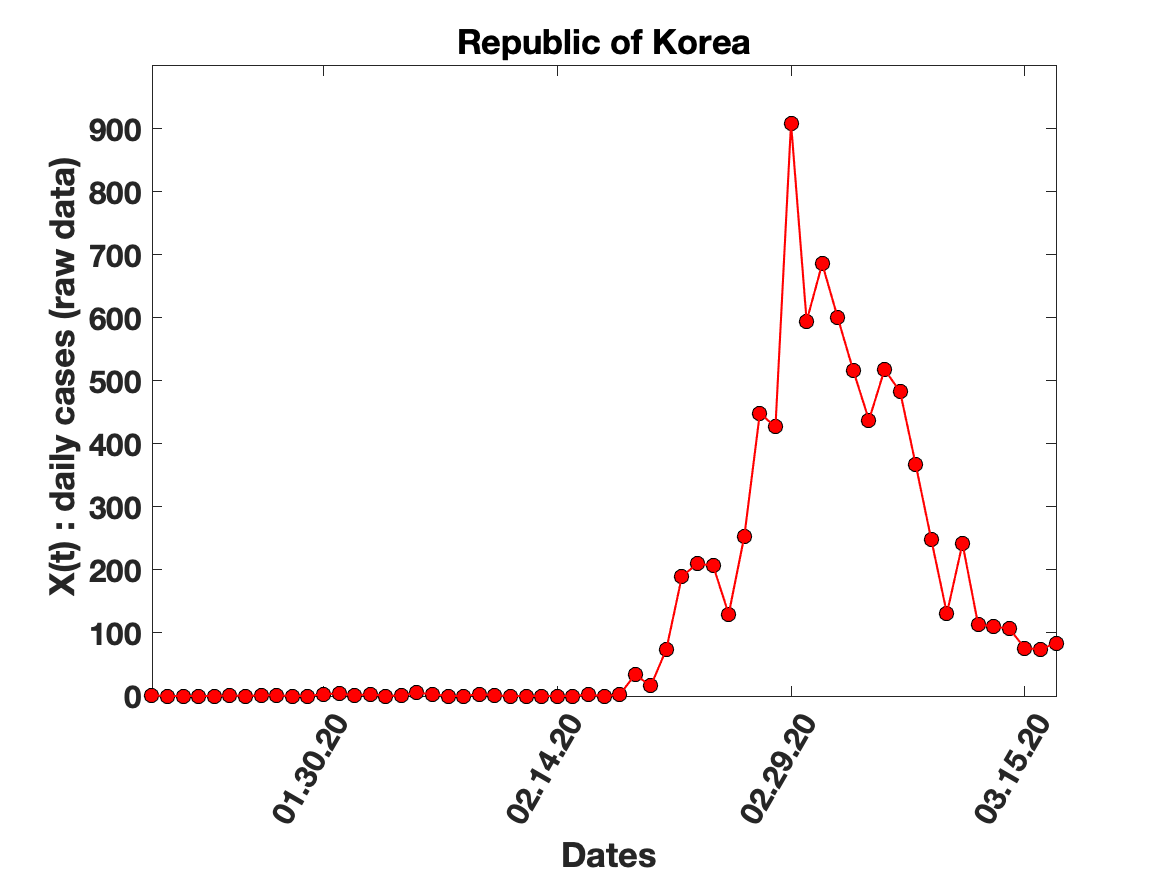

### Republic_of_Korea_solution_fit.png

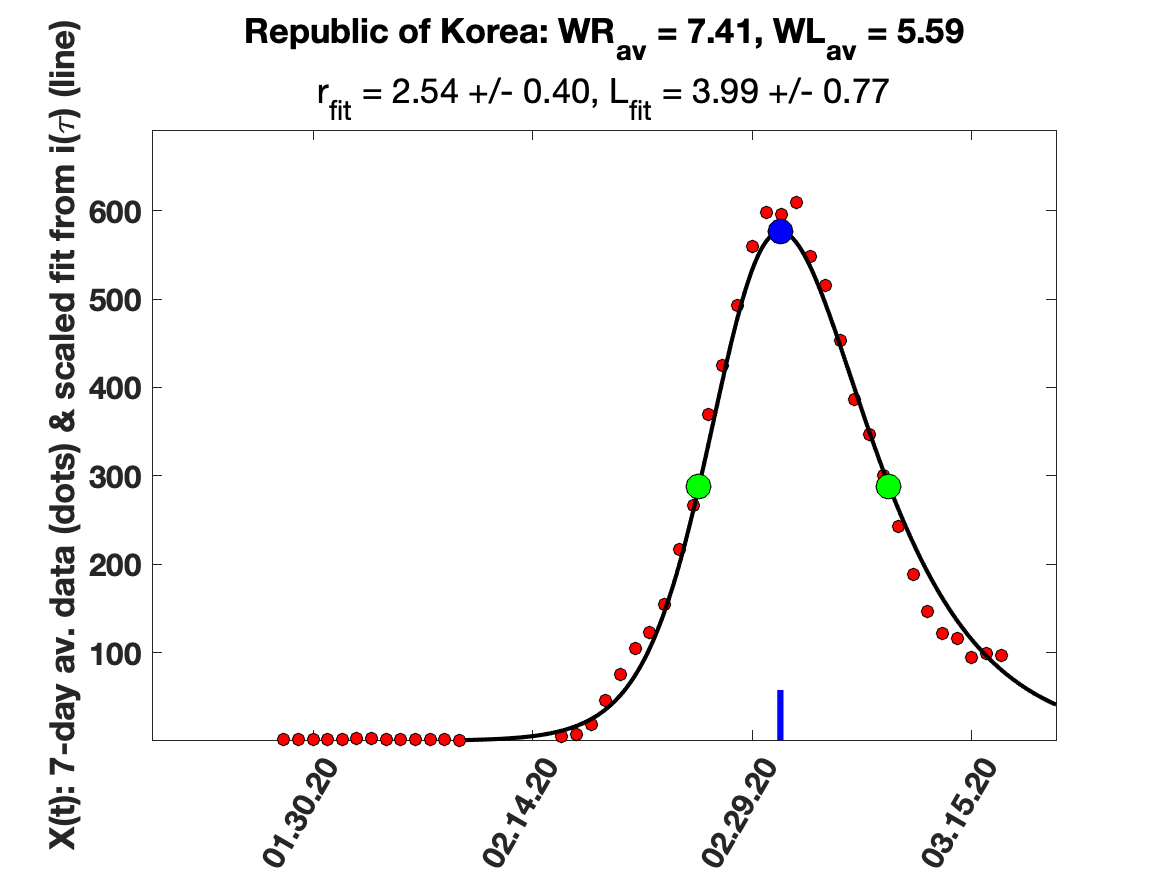

### South_Afrrica_5_day_av_and_fit.png

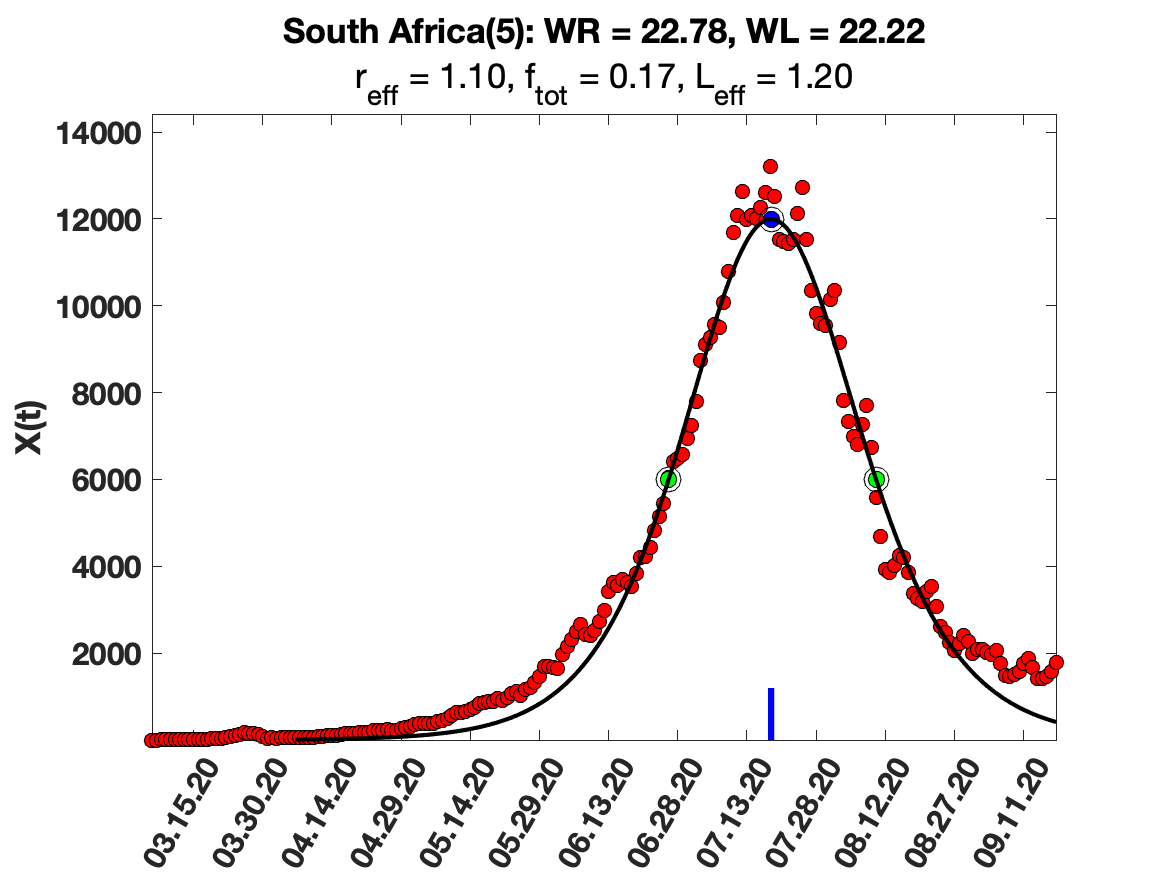

### South_Afrrica_7_day_av_and_fit.png

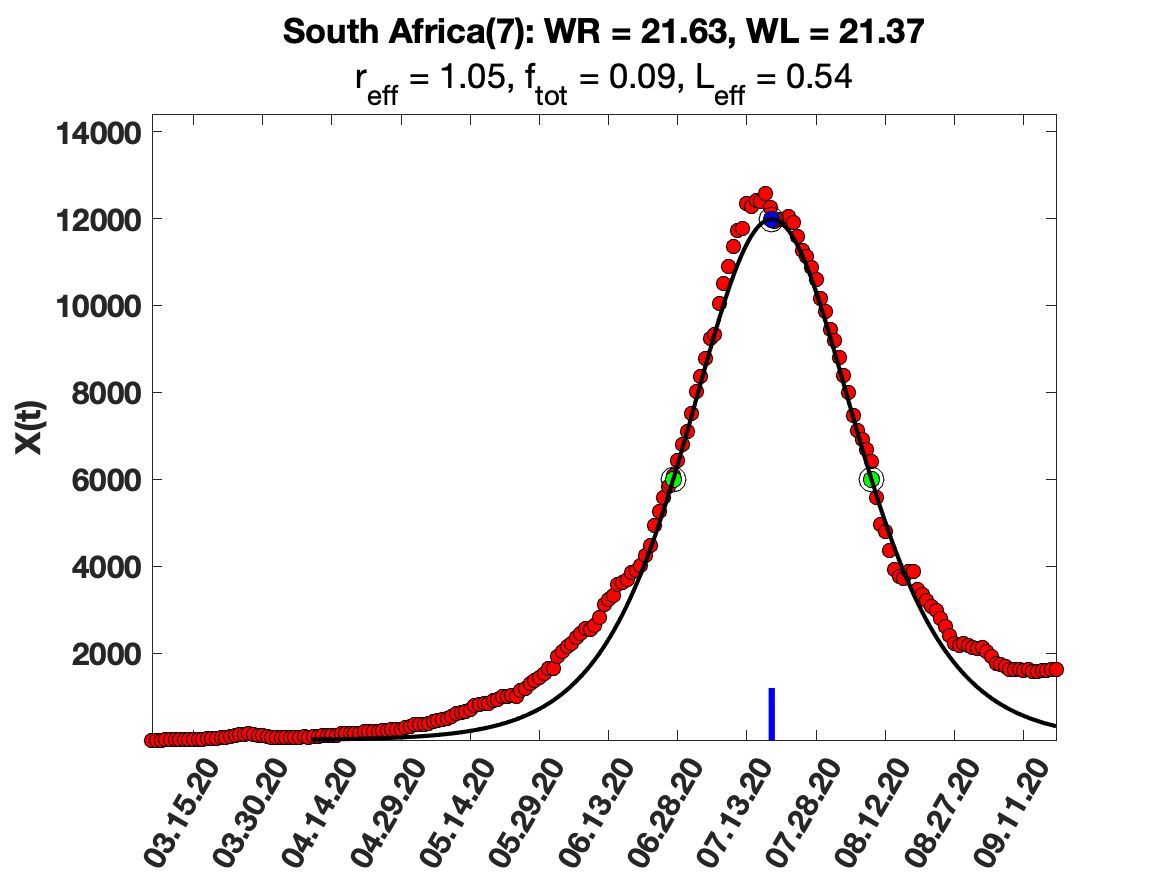

### South_Afrrica_i(tau)_for_average_r.png

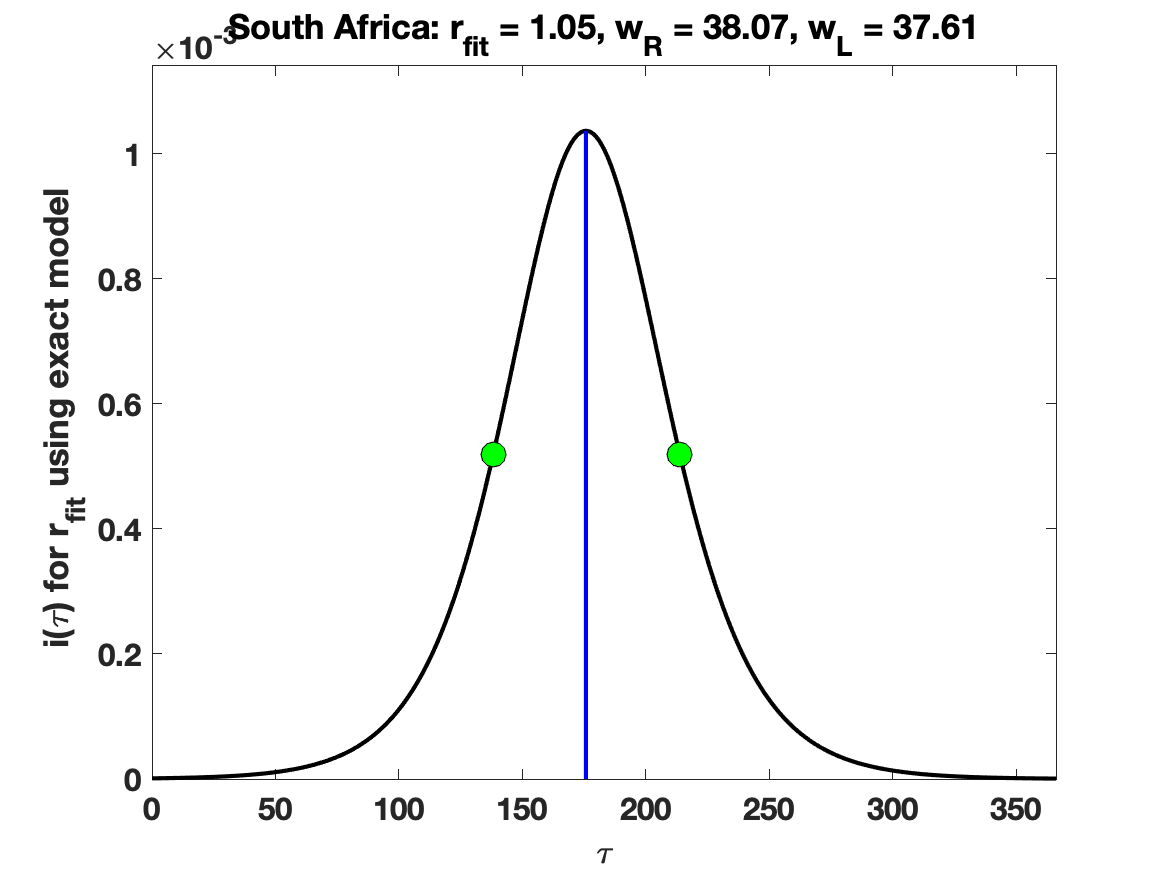

### South_Afrrica_raw_data.png

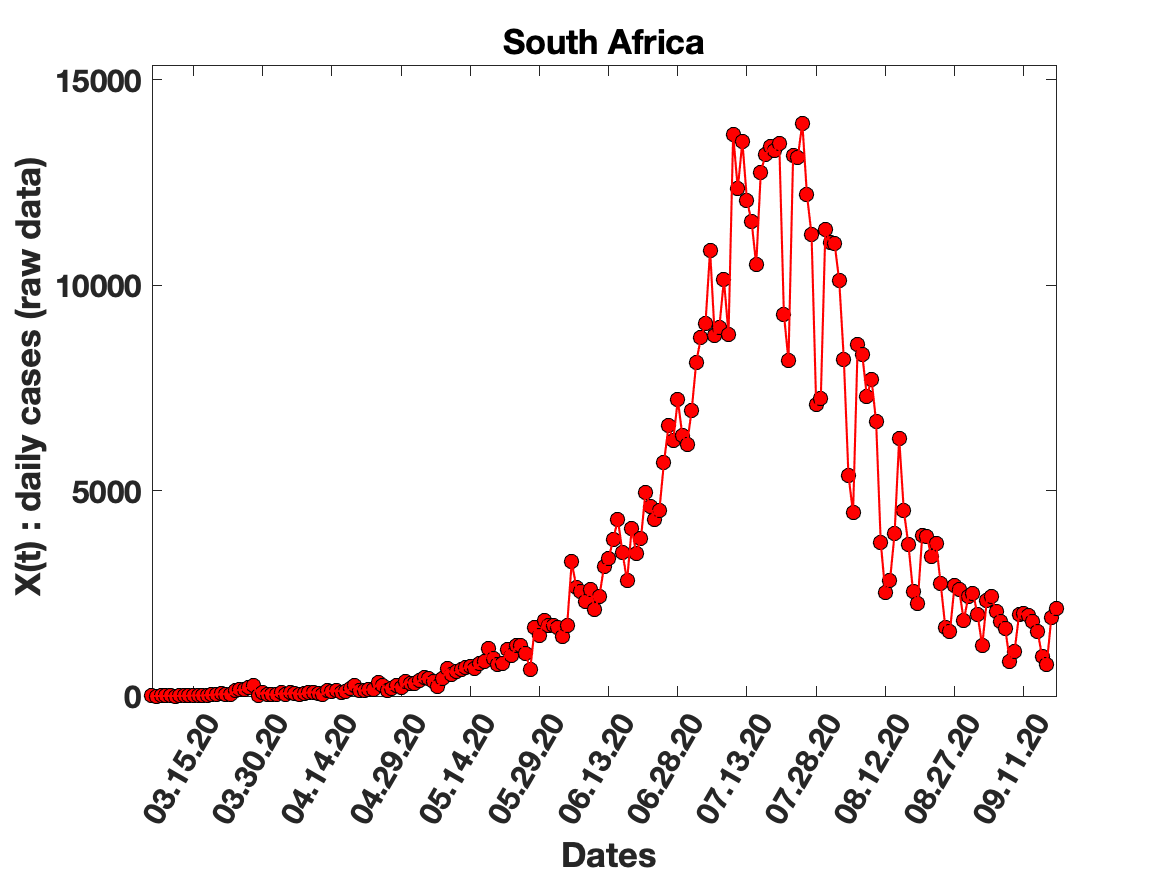

### Spain_5_day_av_and_fit.png

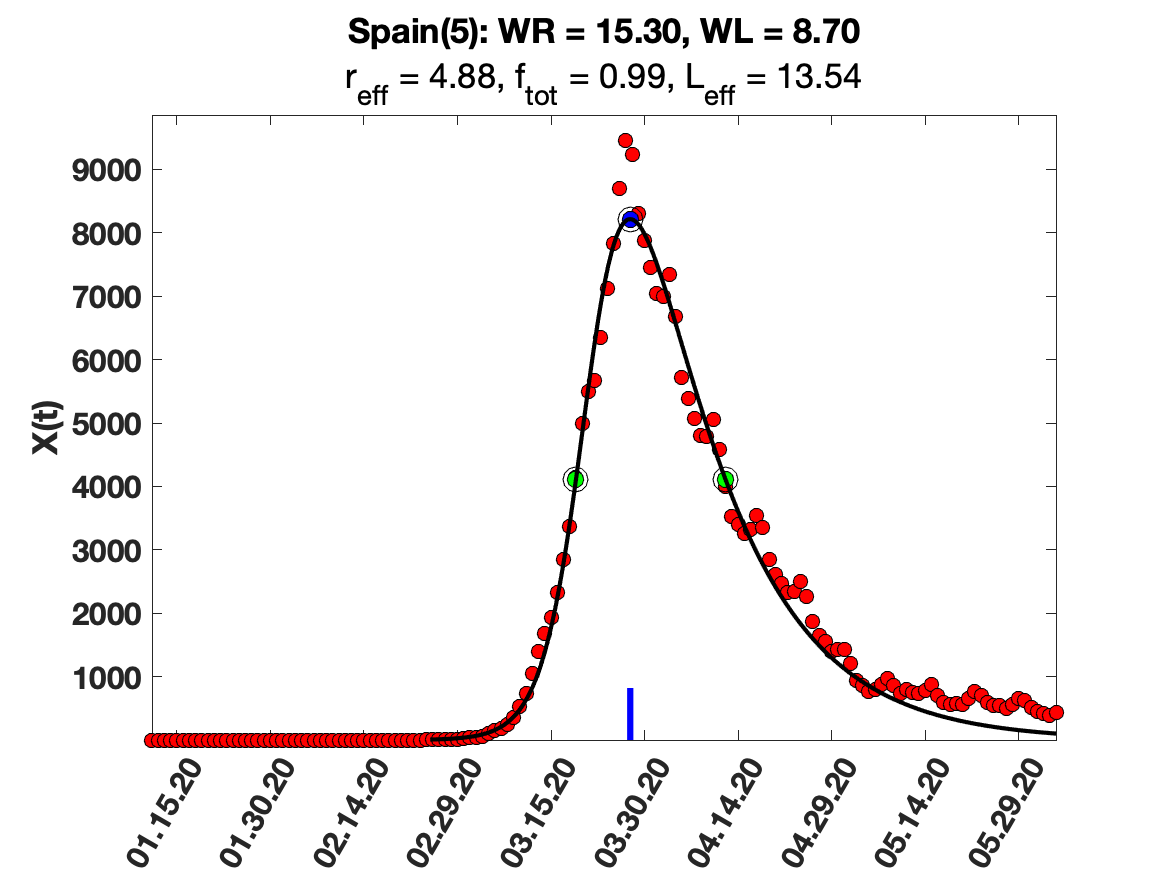

### Spain_7_day_av_and_fit.png

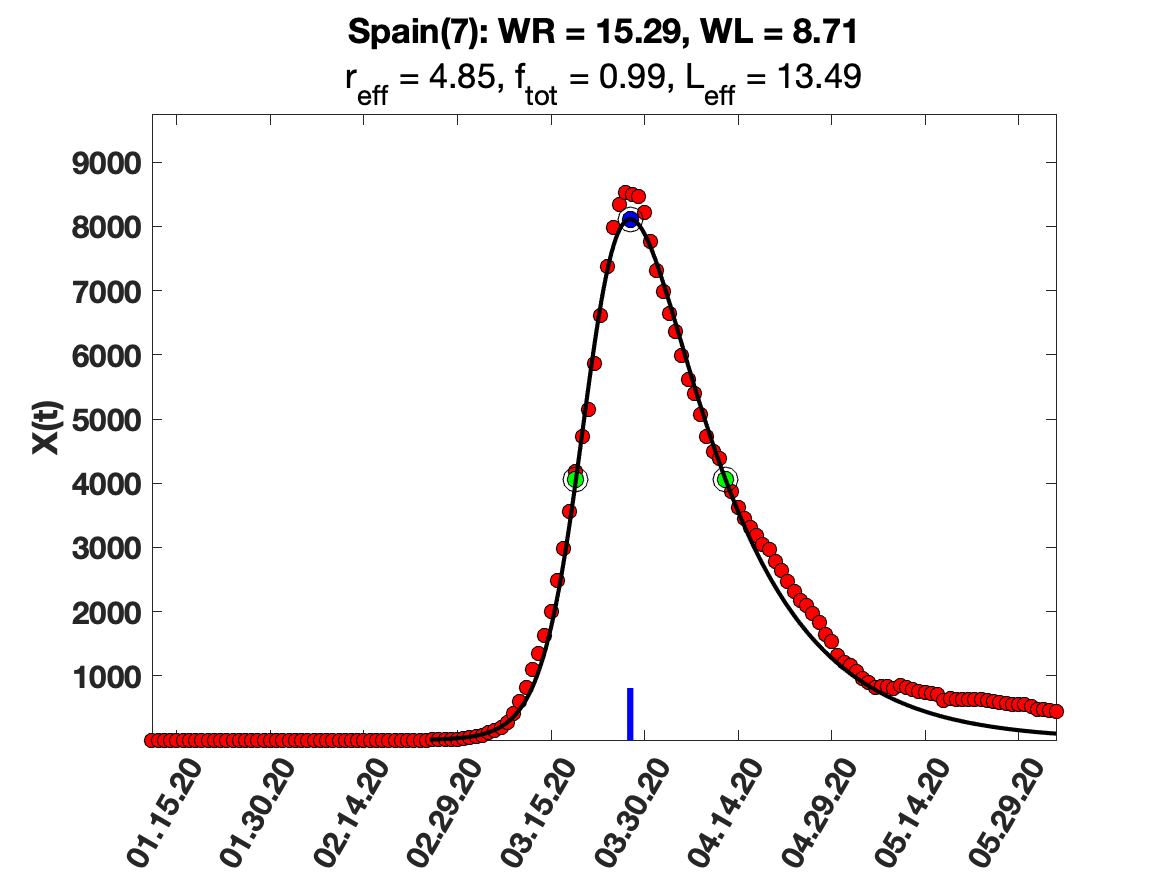

### Spain_9_day_av_and_fit.png

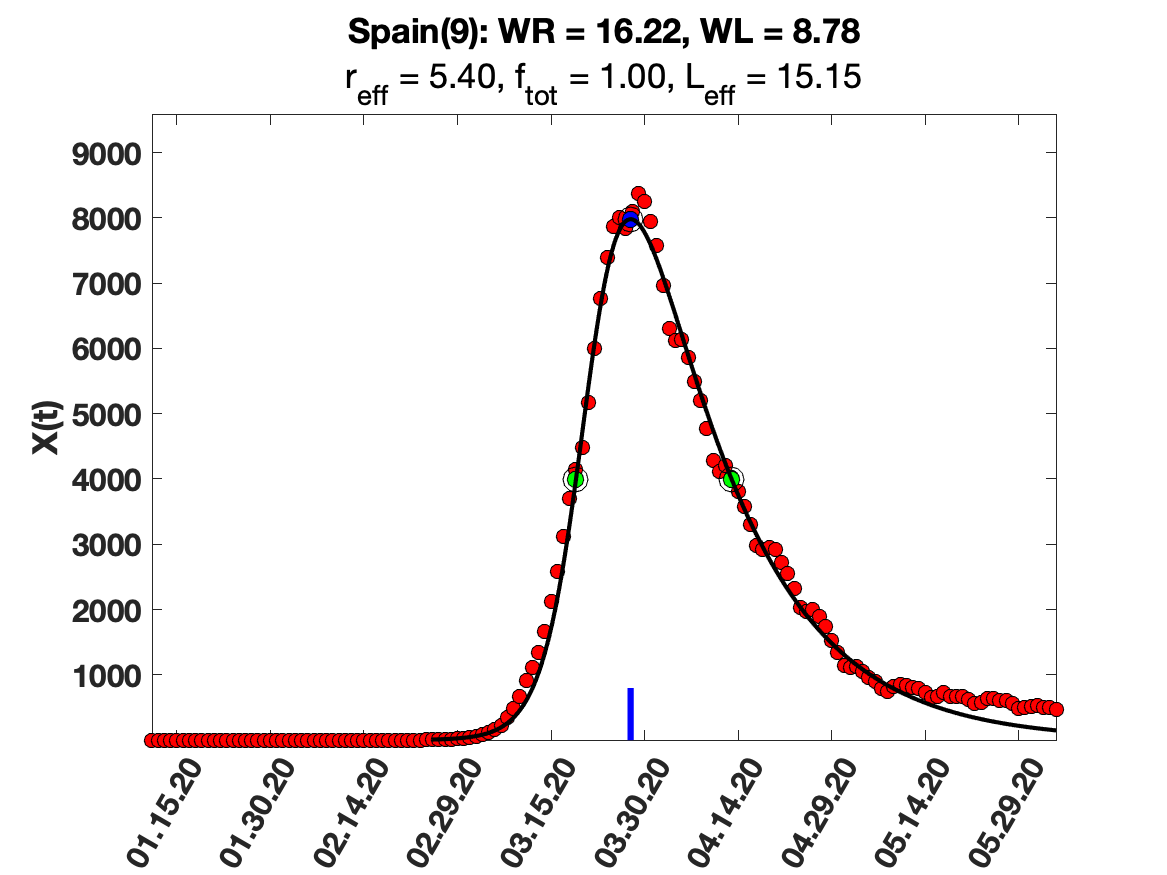

### Spain_raw_data.png

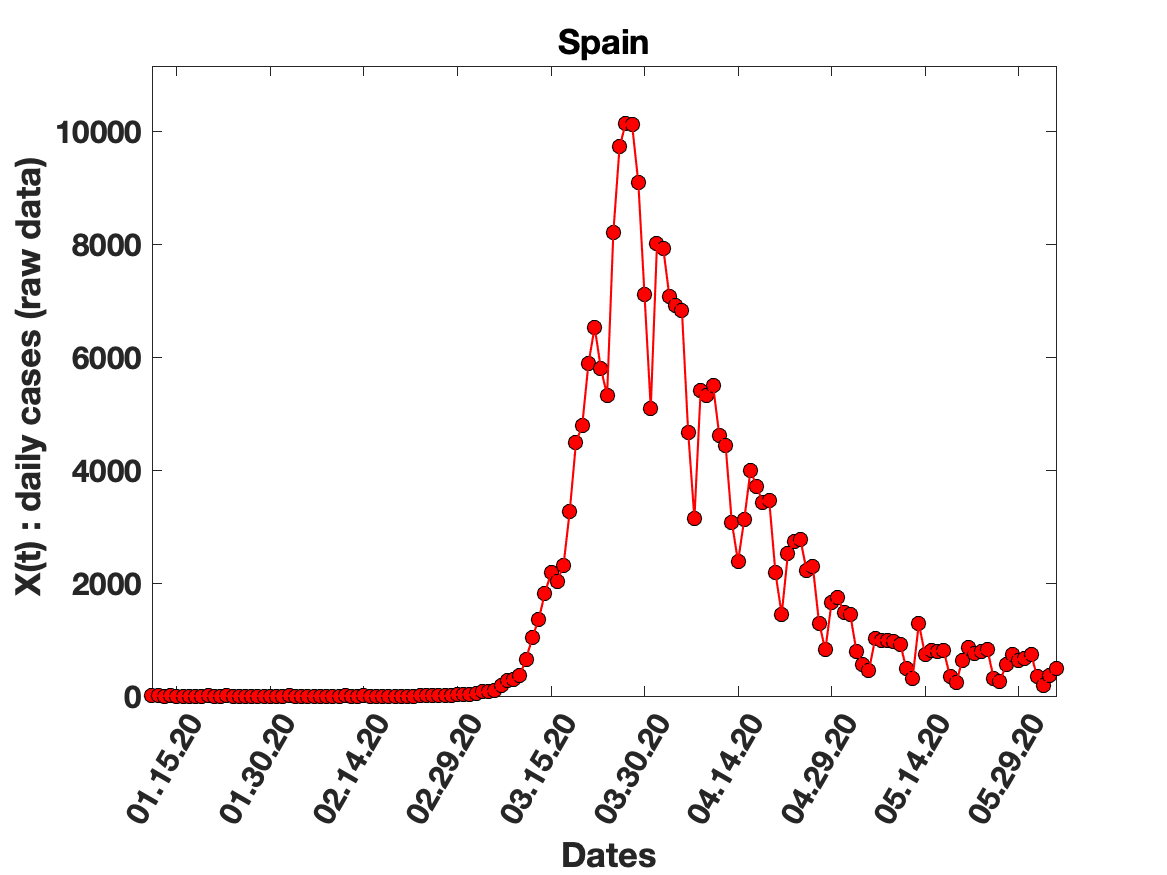

### Spain_solution_fit.png

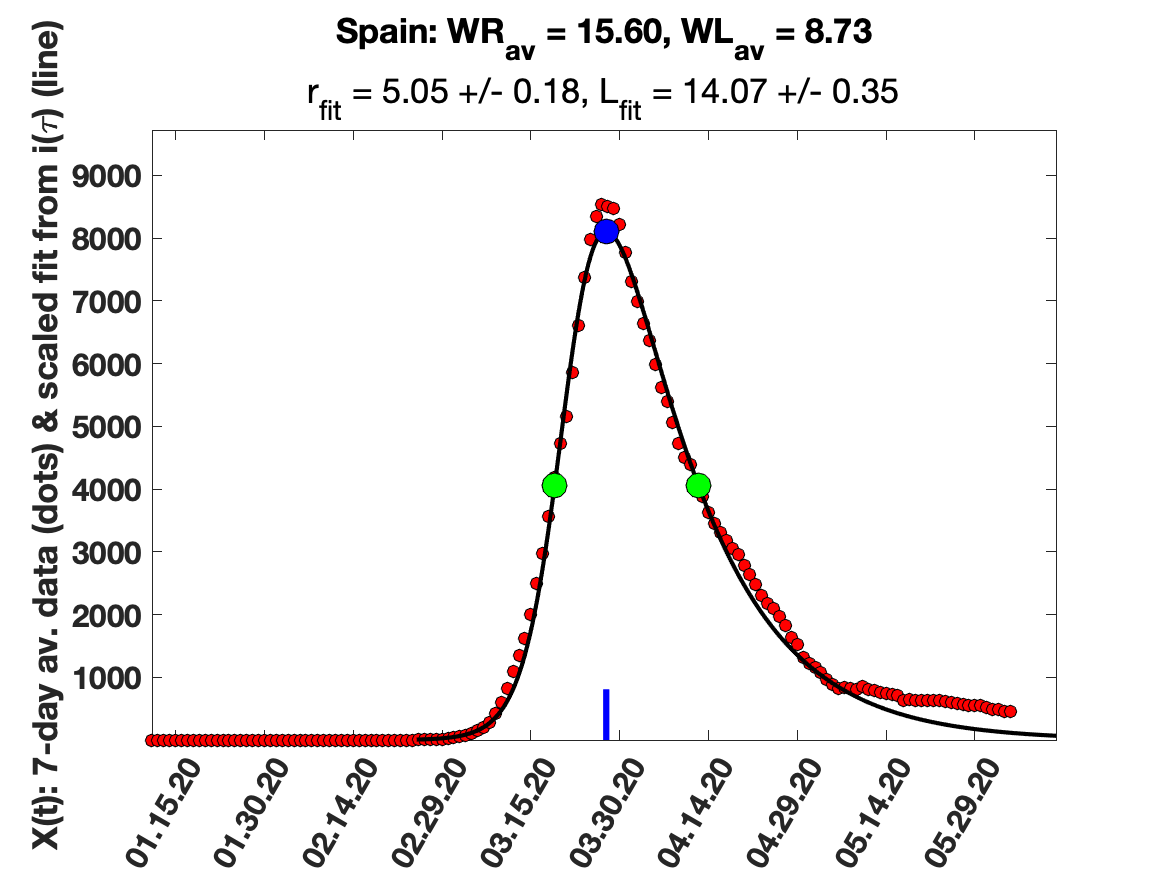

### Switzerland_5_day_av_and_fit.png

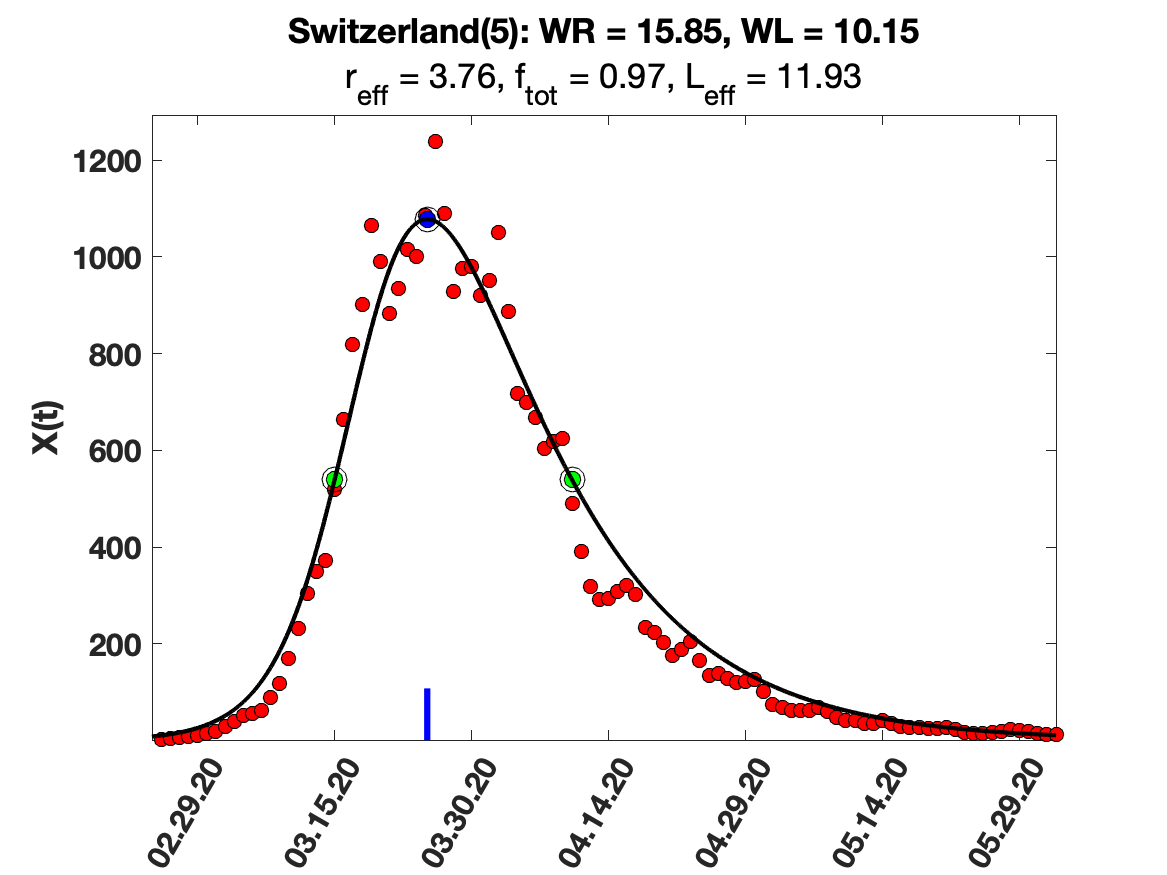

### Switzerland_9_day_av_and_fit.png

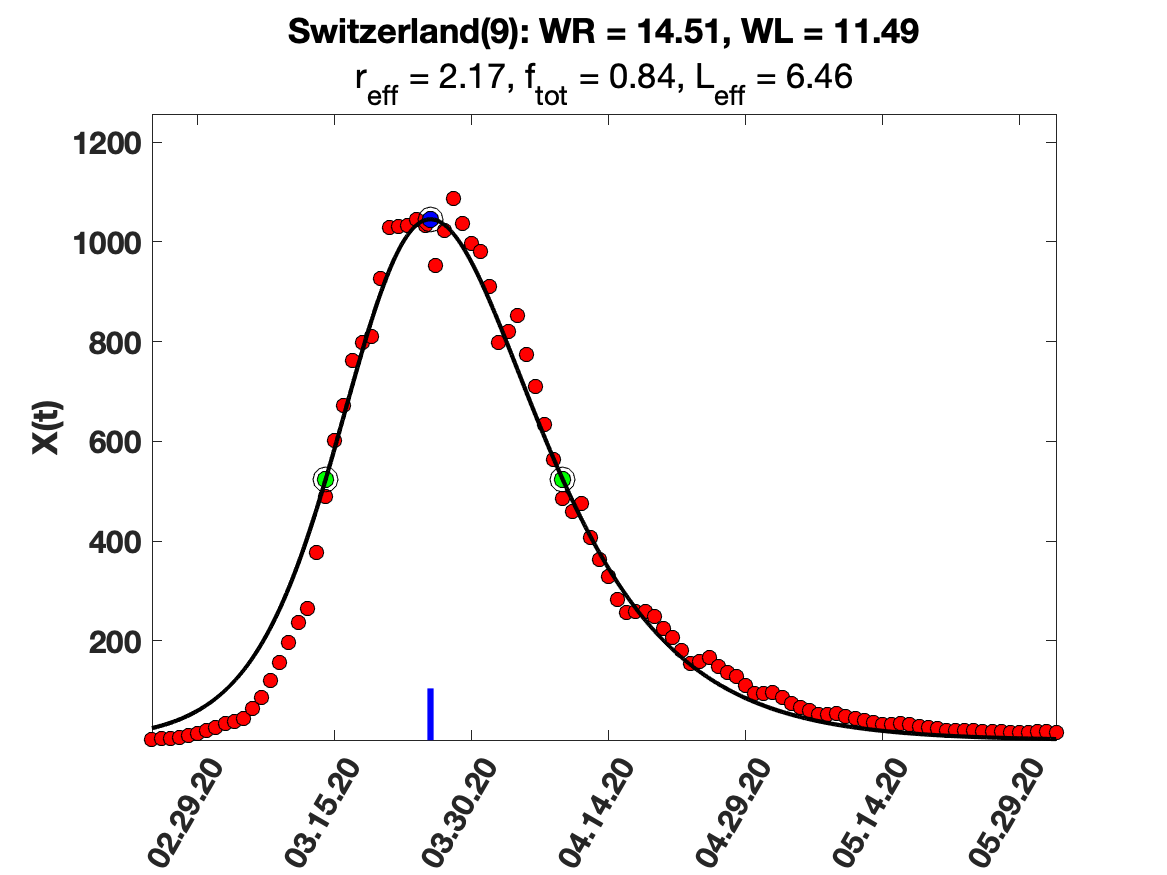

### Switzerland_i(tau)_for_average_r.png

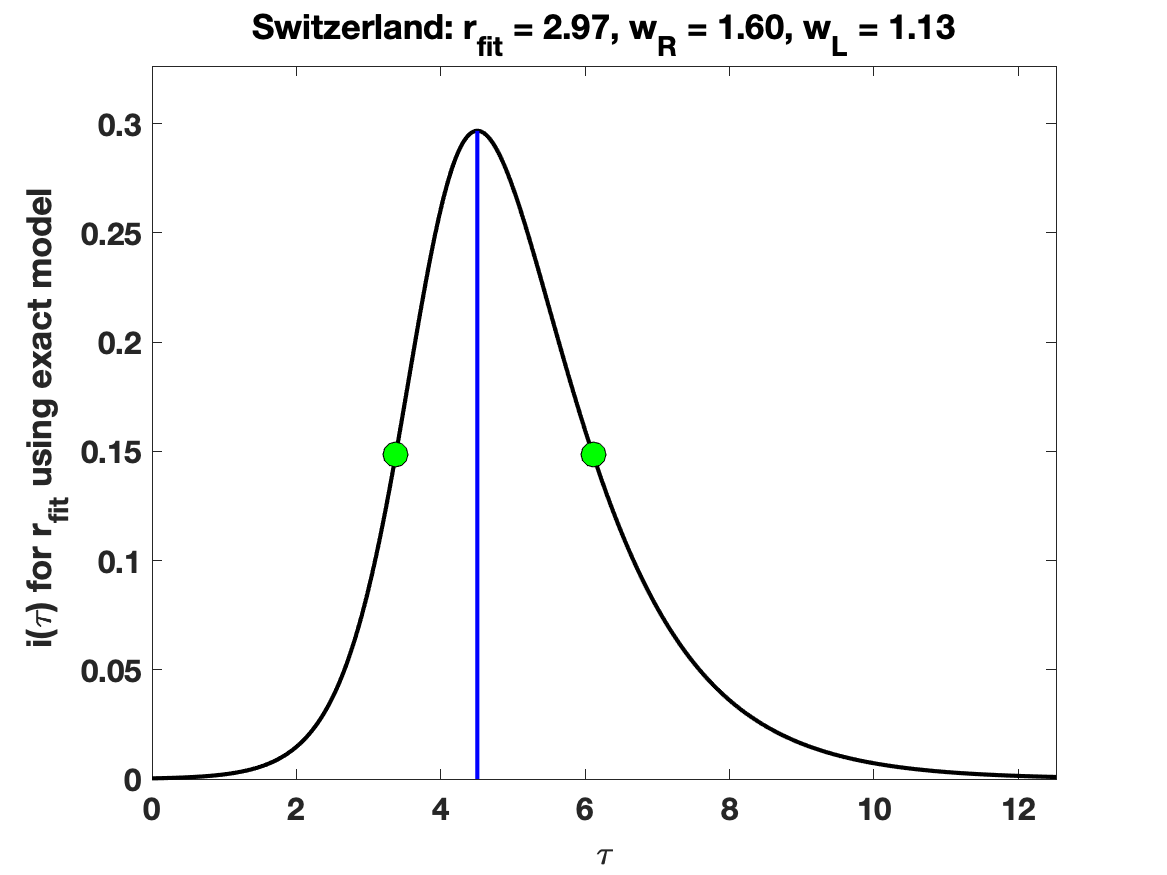

### Switzerland_raw_data.png

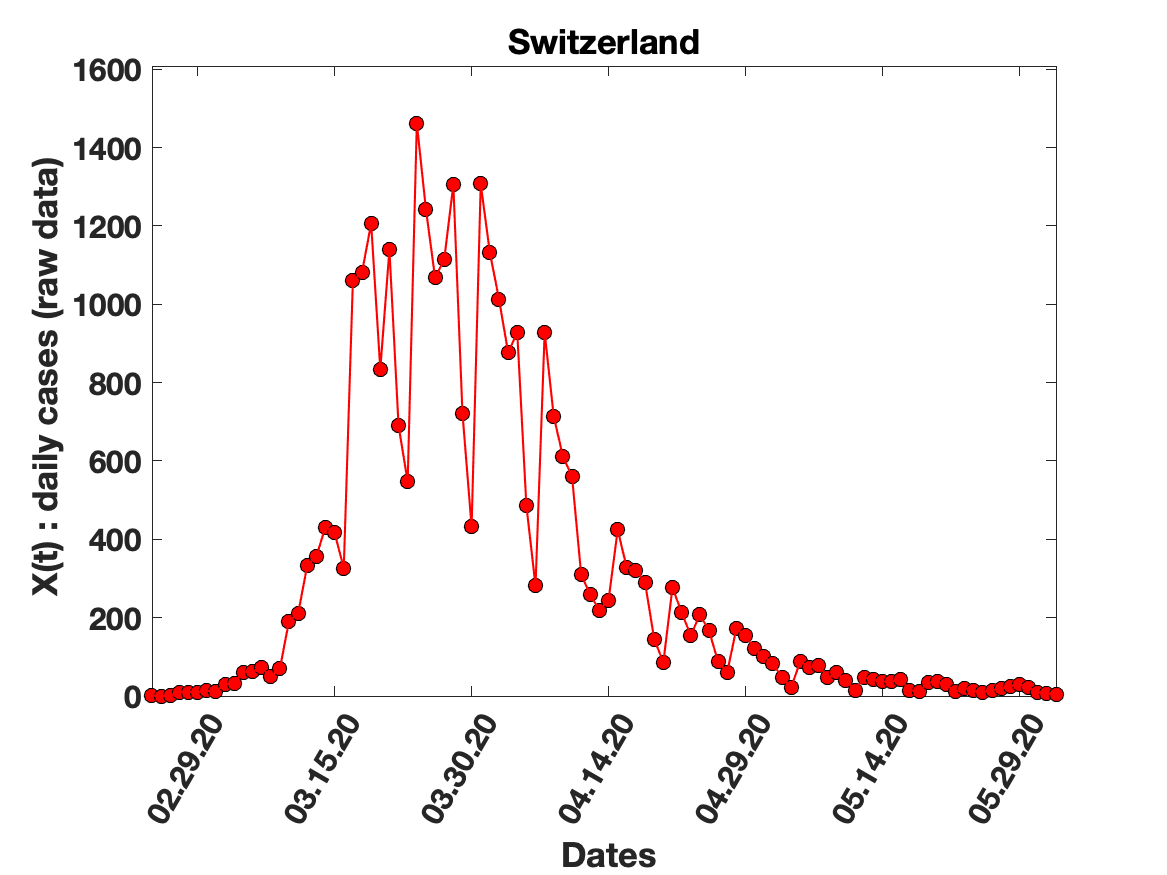

### Thailand_5_day_av_and_fit.png

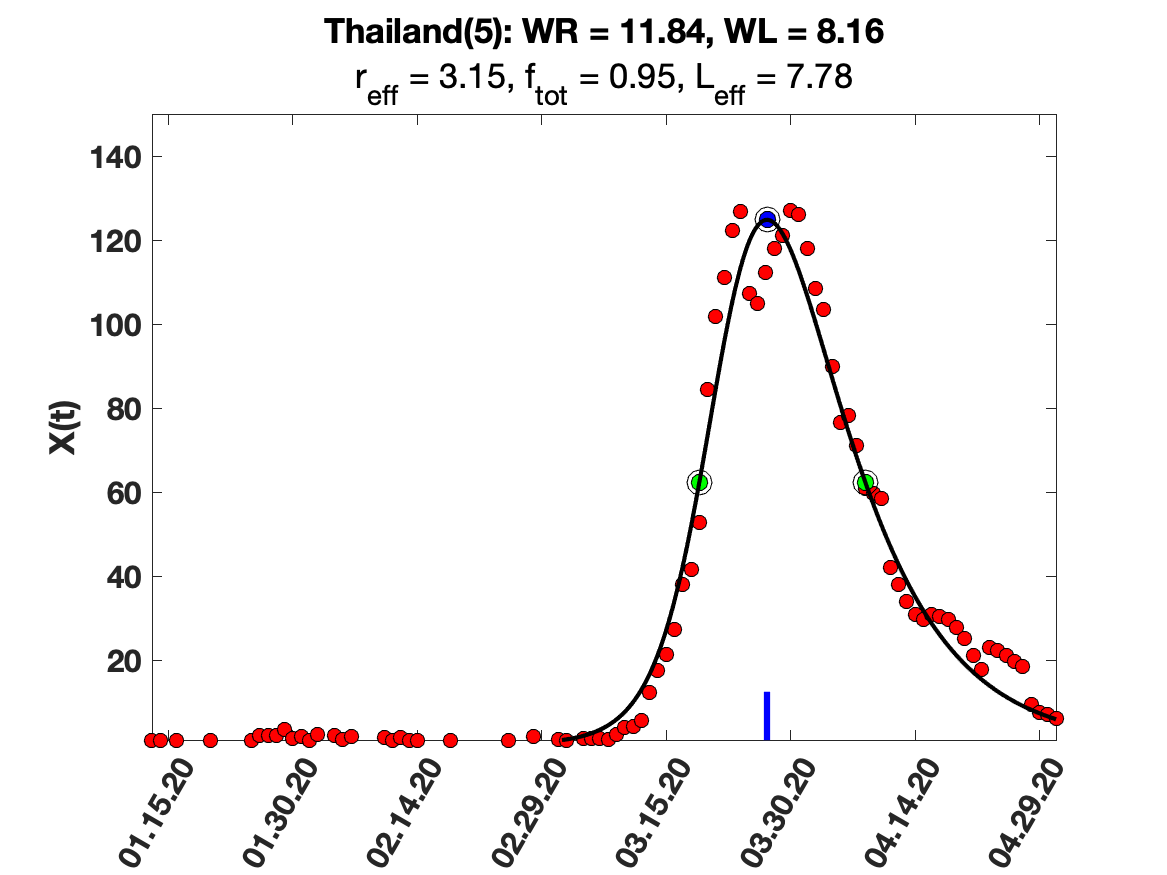

### Thailand_7_day_av_and_fit.png

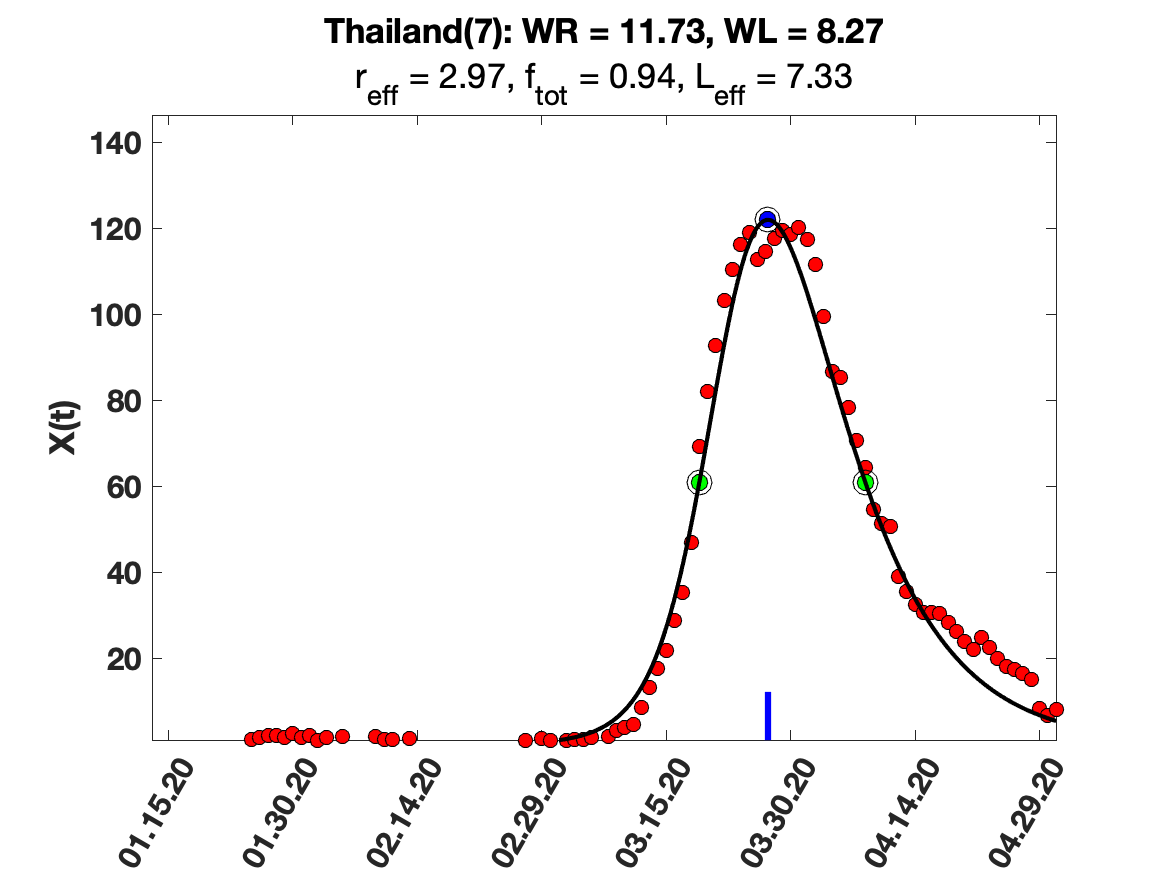

### Thailand_9_day_av_and_fit.png

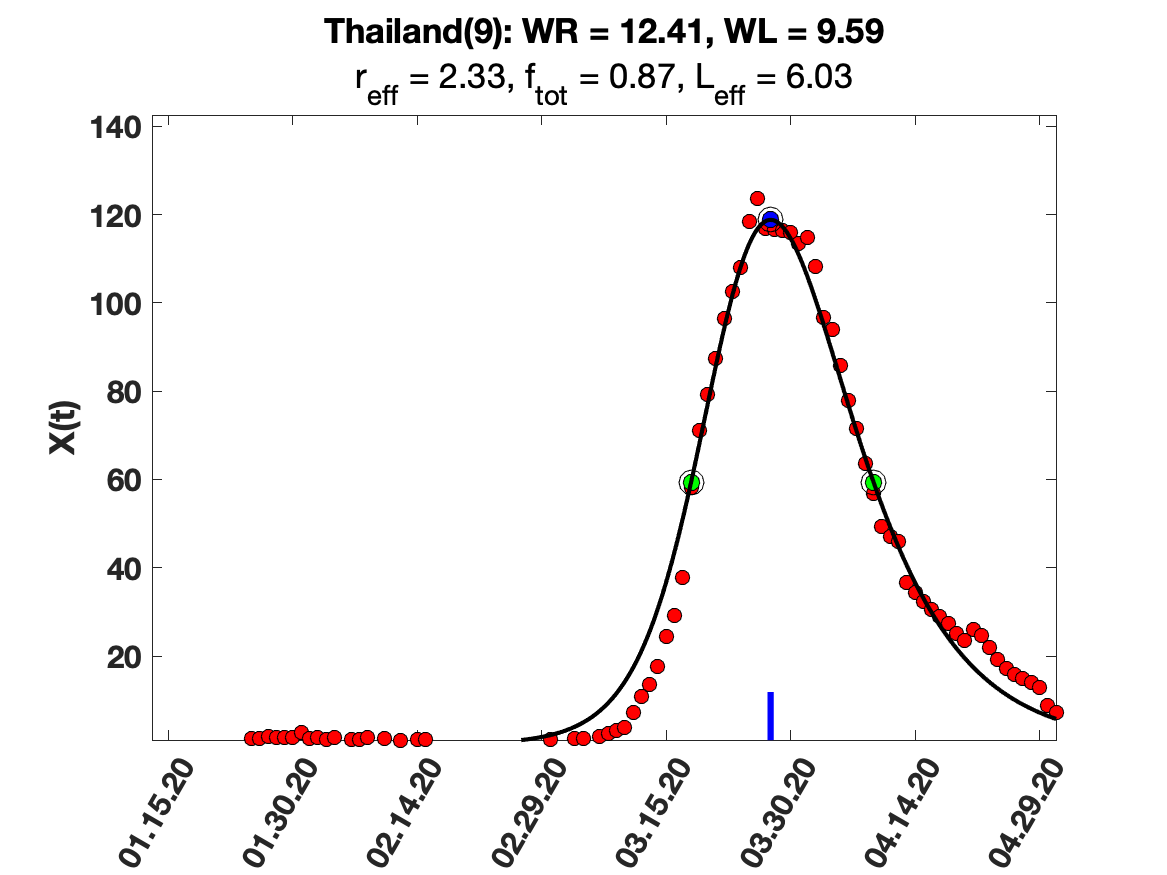

### Thailand_i(tau)_for_average_r.png

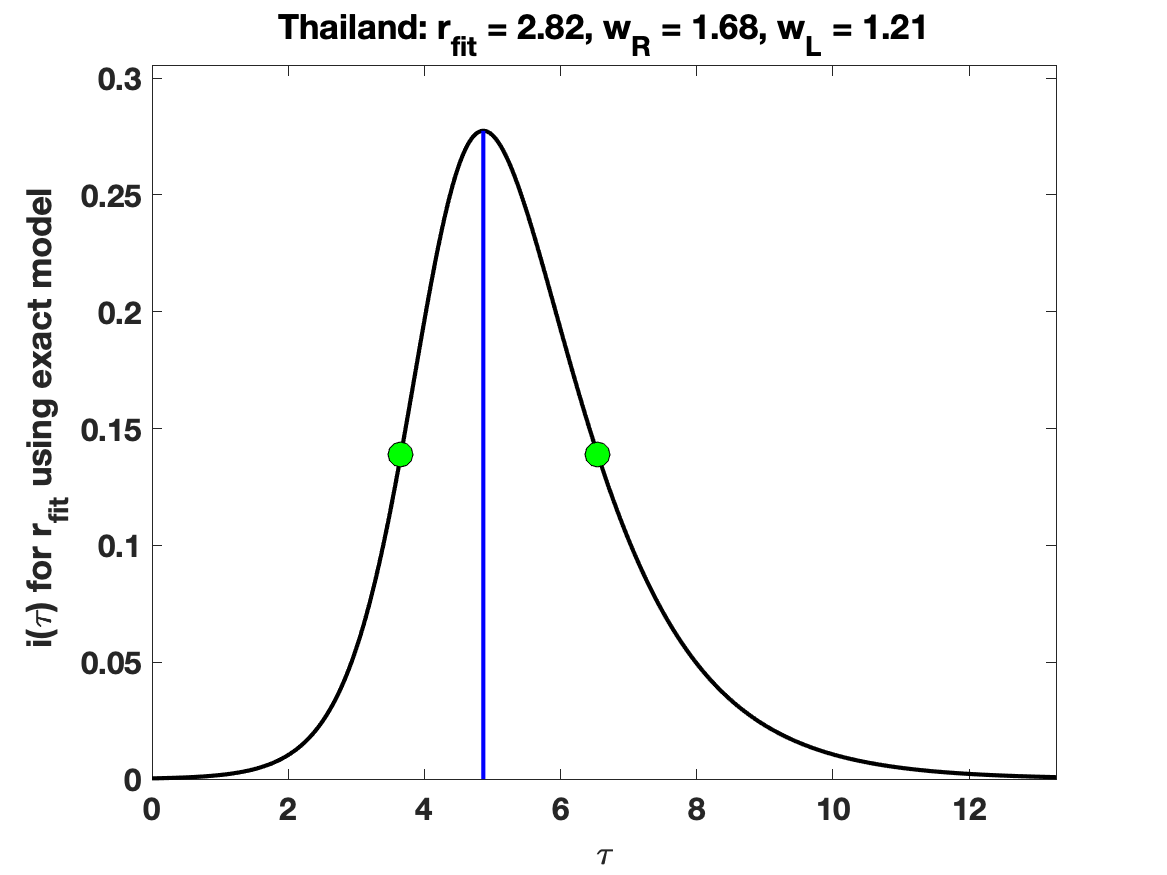

### Thailand_raw_data.png

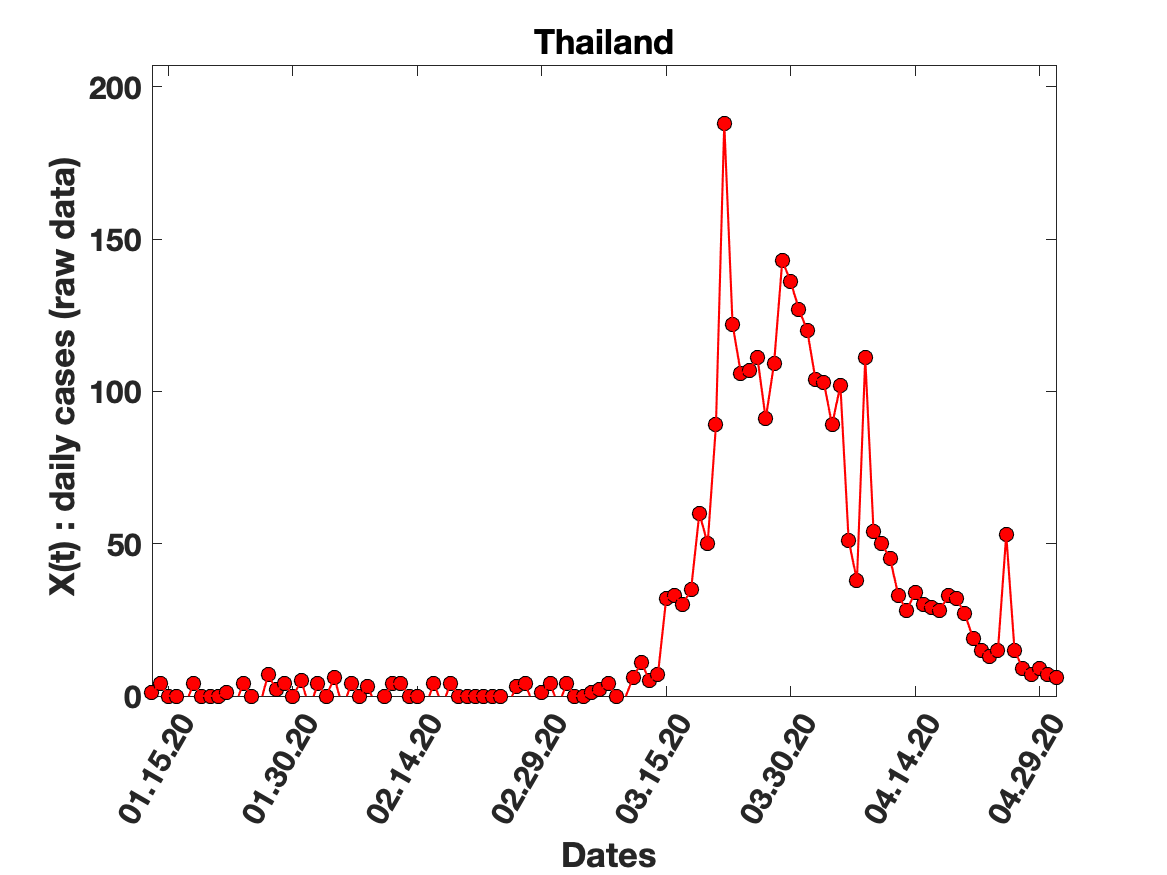

### Turkey_5_day_av_and_fit.png

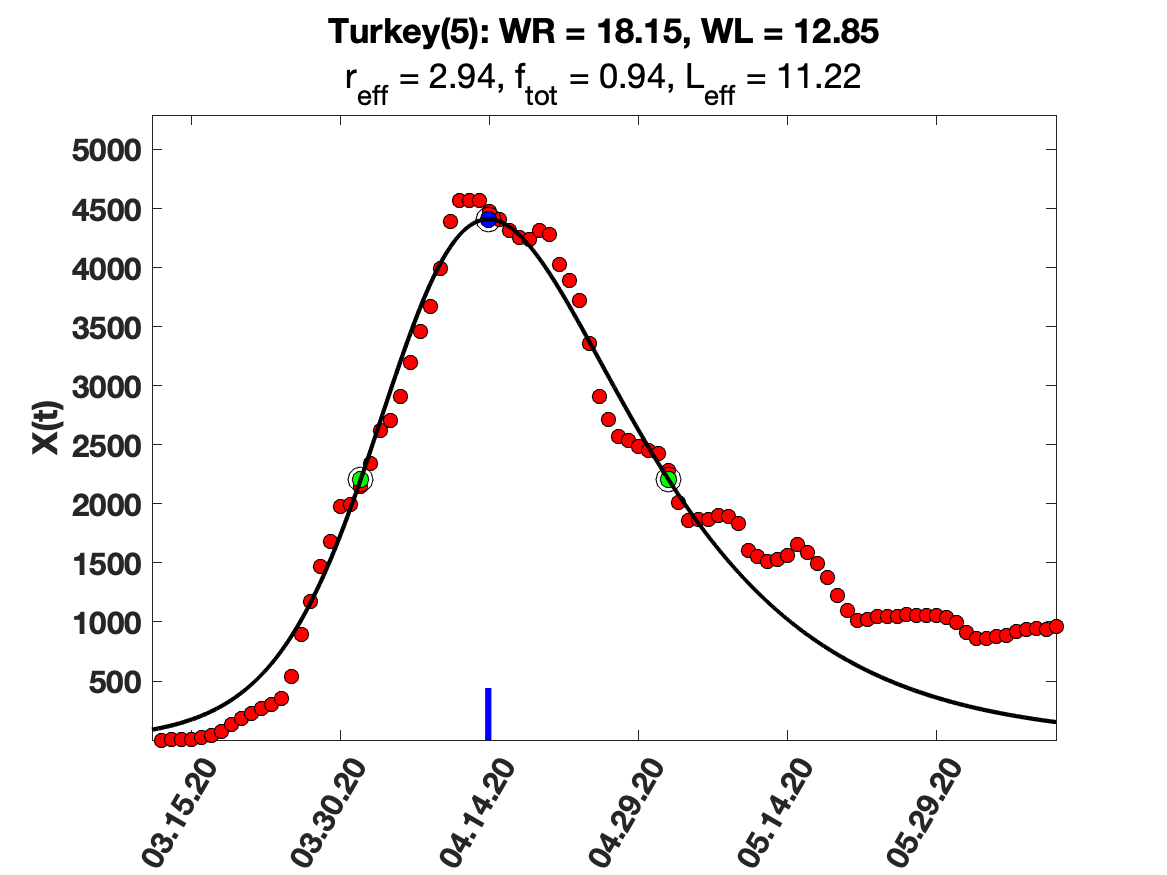

### Turkey_i(tau)_for_average_r.png

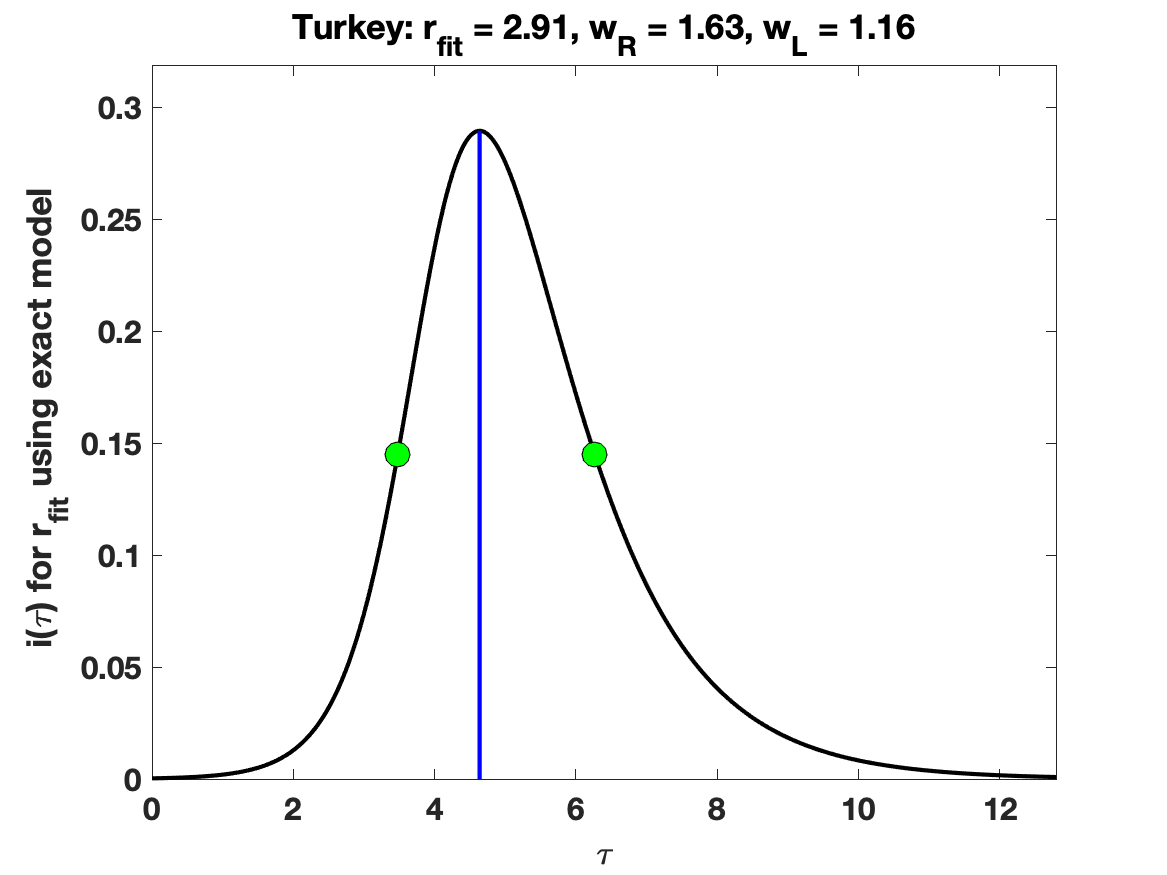

### Turkey_raw_data.png

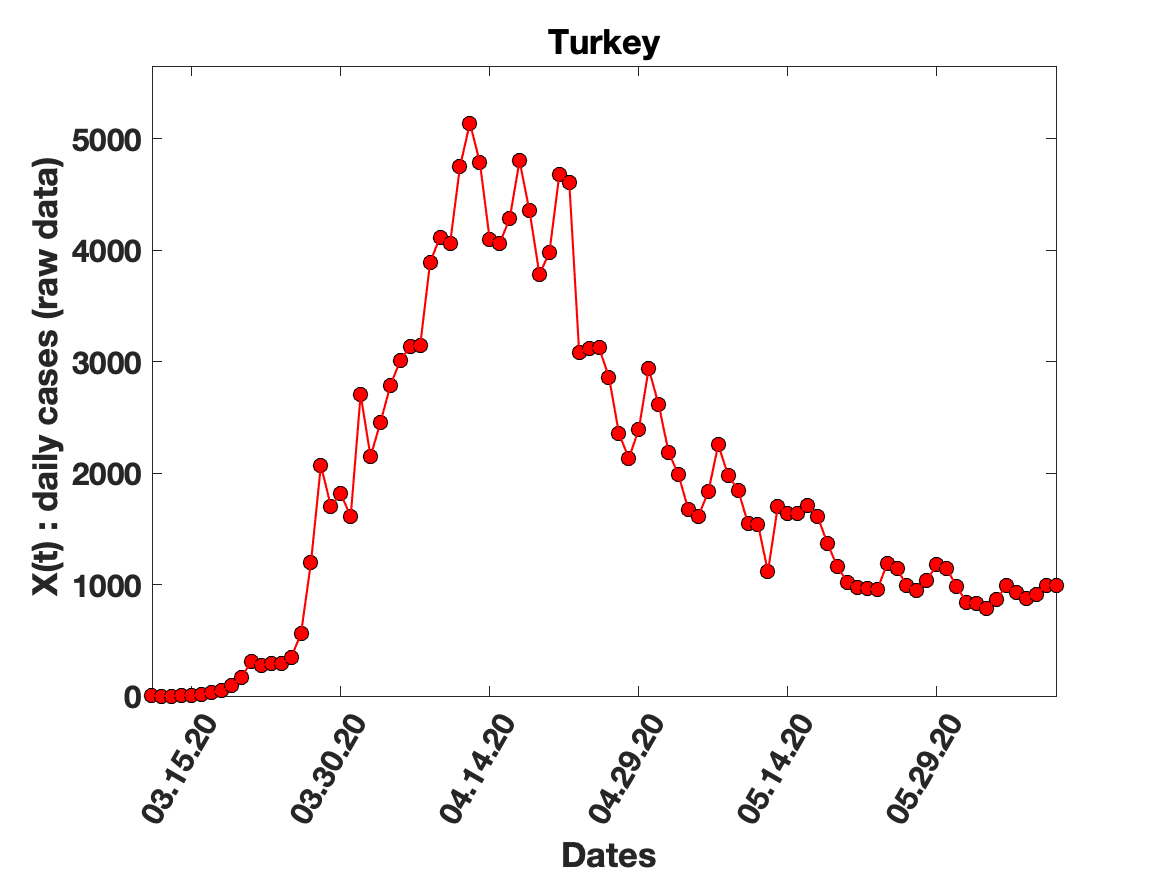

### Turkey_solution_fit.png

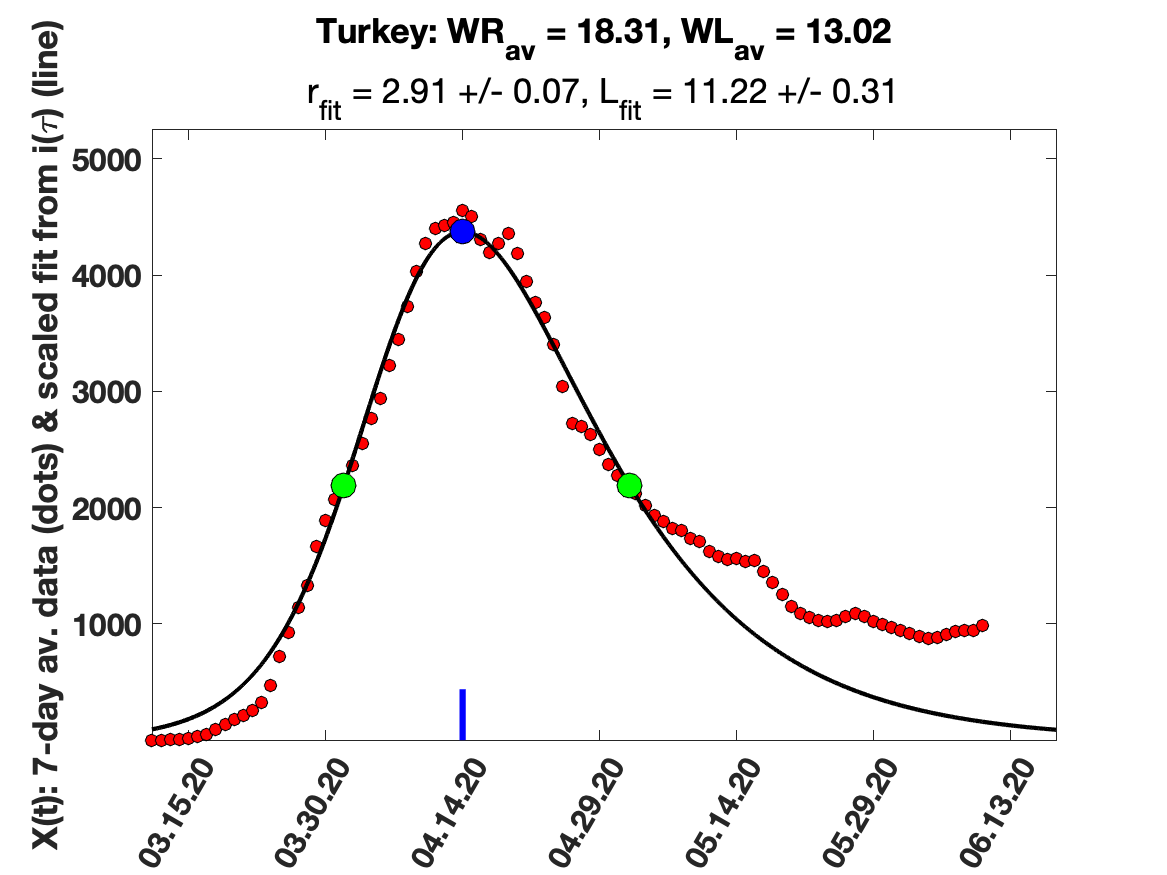

### UK_5_day_av_and_fit.png

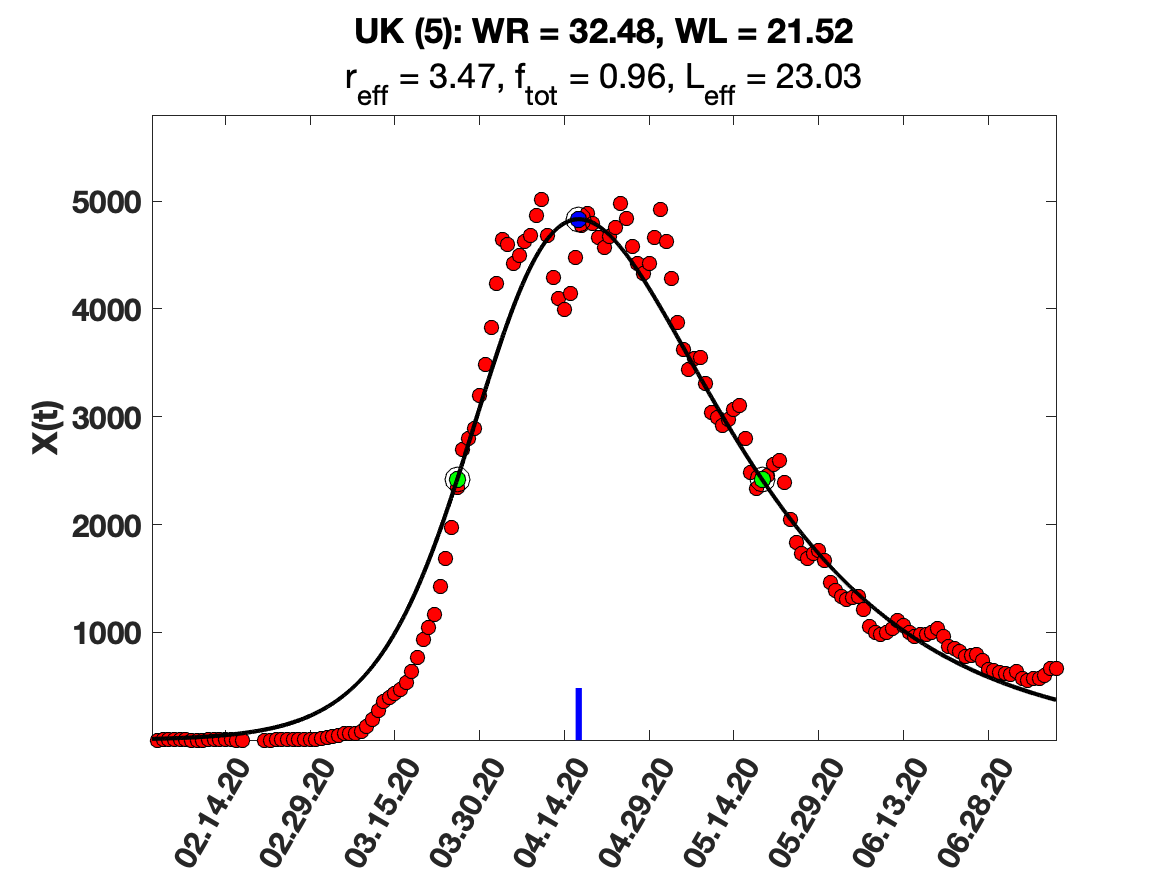

### UK_7_day_av_and_fit.png

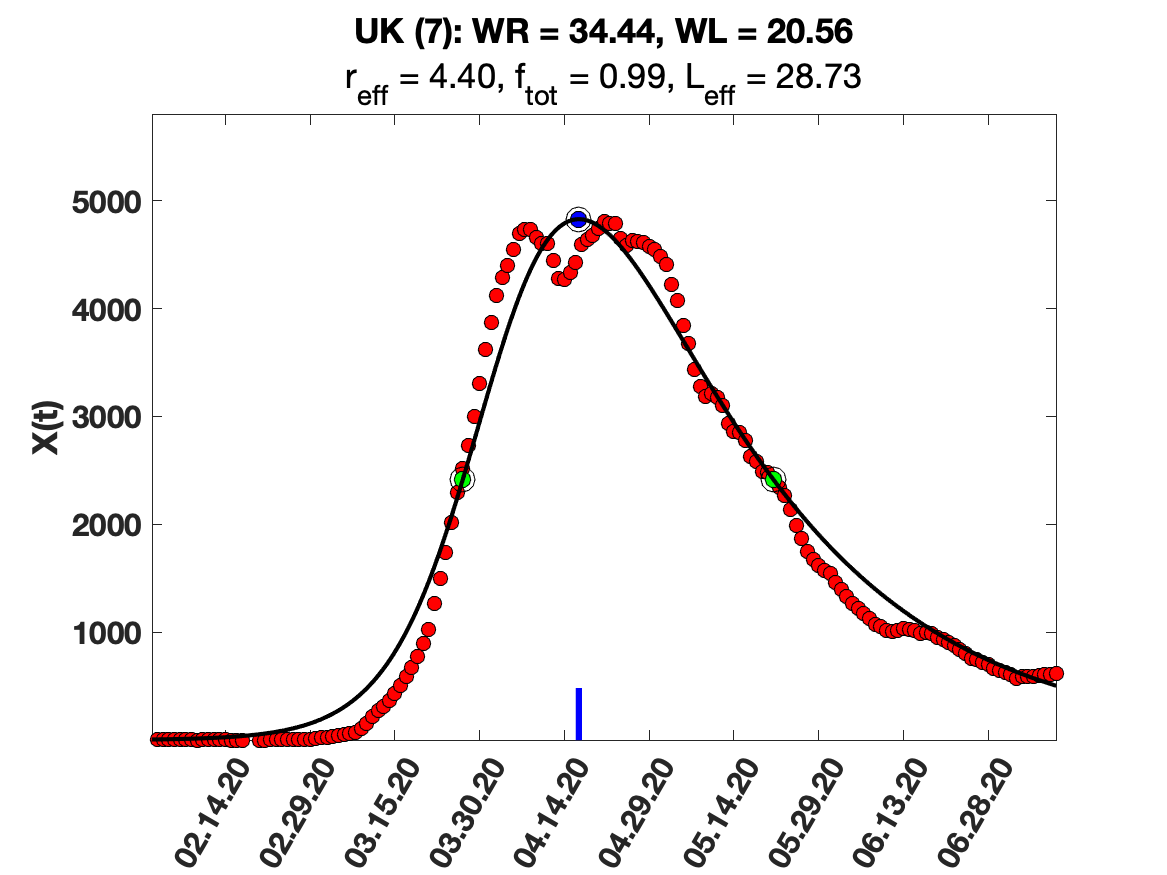

### UK_9_day_av_and_fit.png

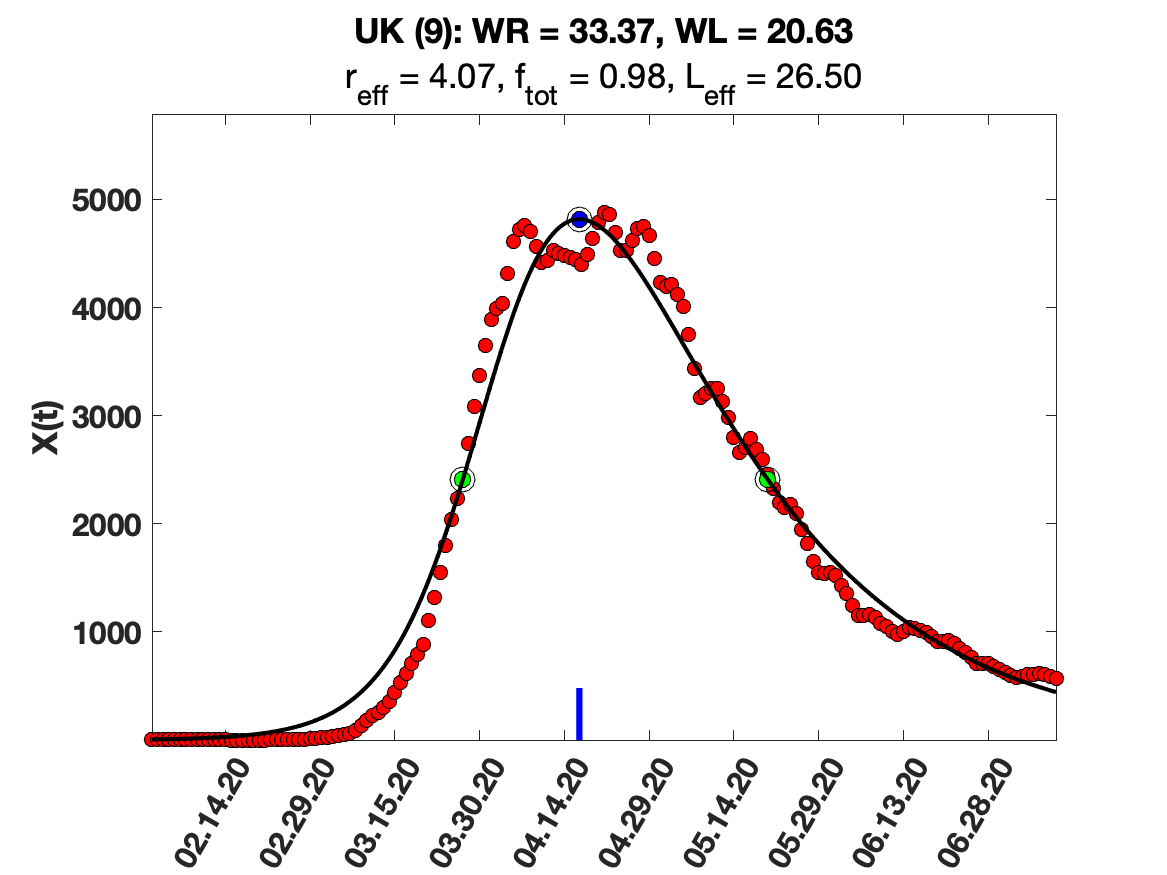

### UK_i(tau)_for_average_r.png

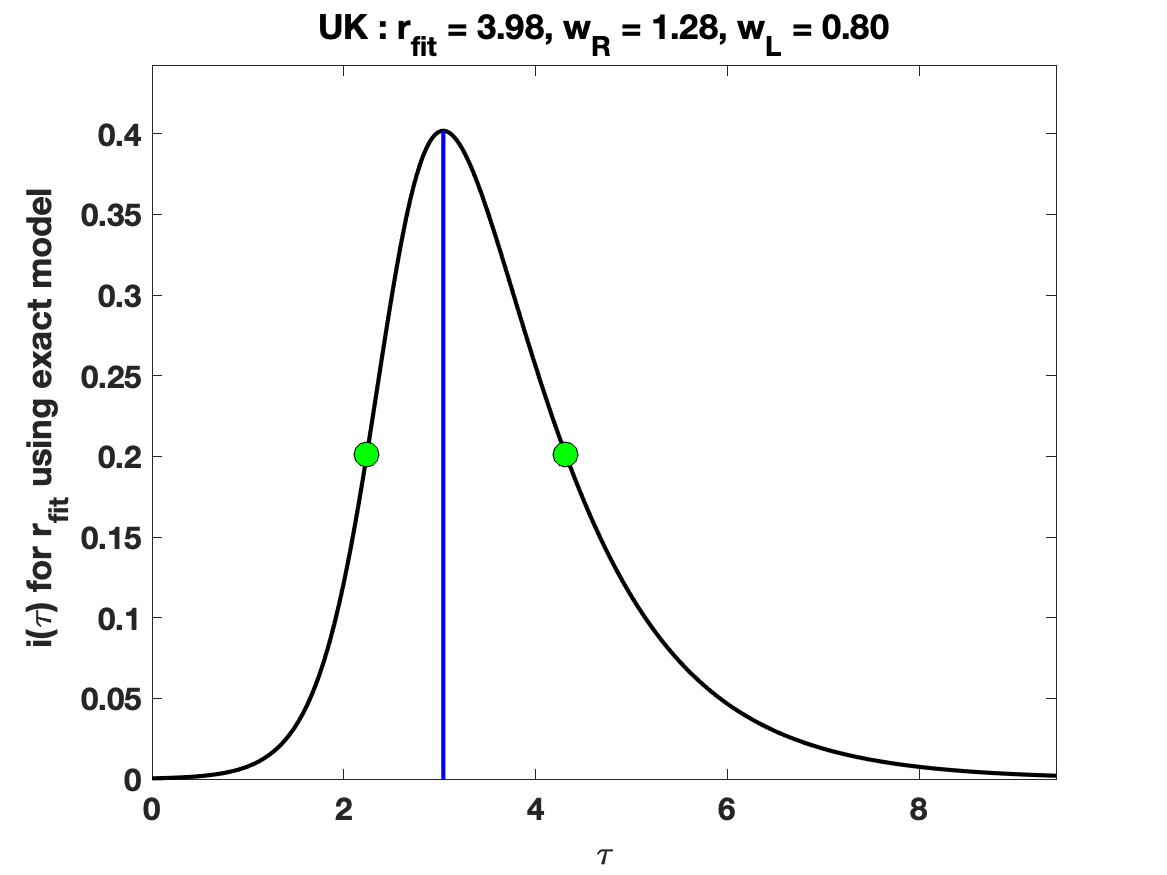

### UK_raw_data.png

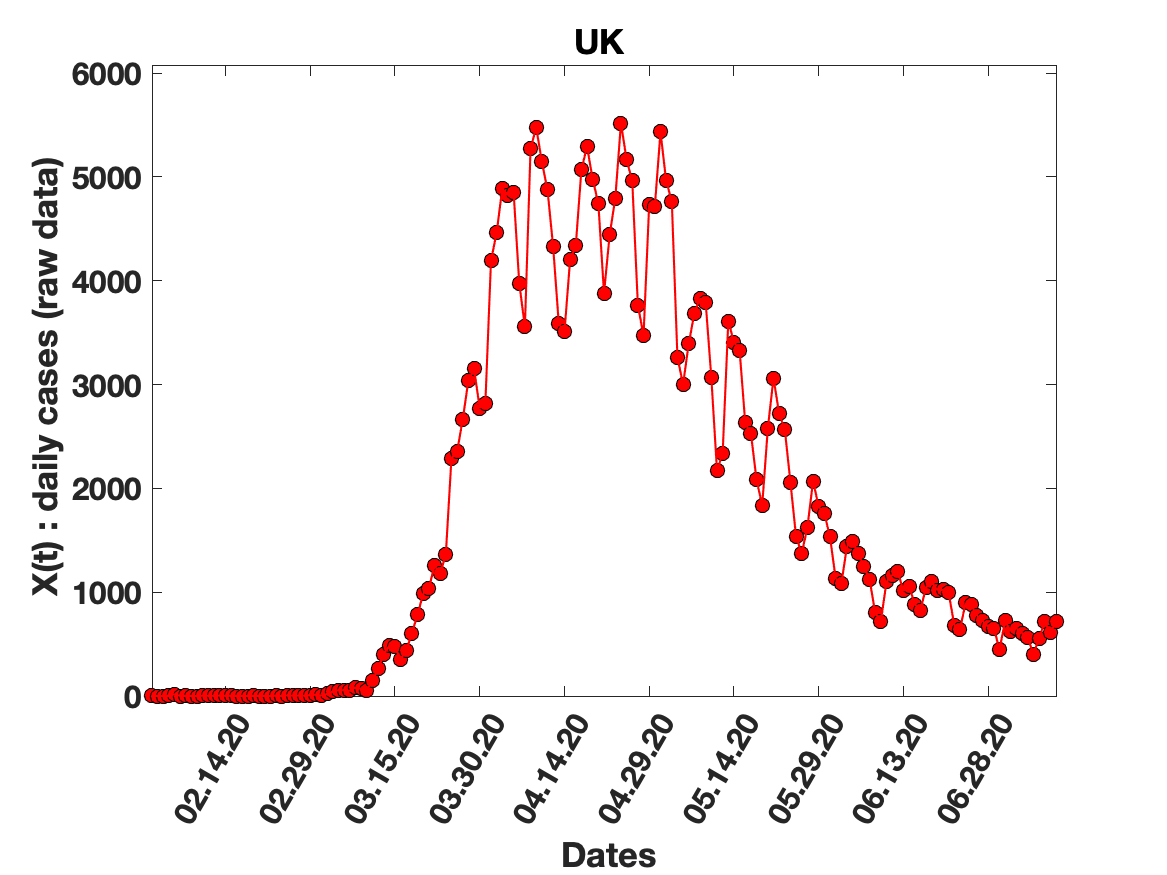
